# The effects of food-based versus supplement-based very low-energy diets on gut microbiome composition and health outcomes in women with high body mass index (The MicroFit Study): a randomised controlled trial

**DOI:** 10.1101/2024.08.11.24311823

**Authors:** Melissa M Lane, Amelia J McGuinness, Mohammadreza Mohebbi, Mojtaba Lotfaliany, Amy Loughman, Martin O’Hely, Adrienne O’Neil, Jessica Batti, Mark A Kotowicz, Michael Berk, Lucy Saunders, Richard Page, Sally Beatti, Wolfgang Marx, Felice N Jacka

**Affiliations:** Deakin University, Geelong, Australia, the Institute for Mental and Physical Health and Clinical Translation (IMPACT), Food & Mood Centre, School of Medicine and Barwon Health, Geelong, Australia; Deakin University, Faculty of Health, Biostatistics Unit, Geelong, Australia; Deakin University, Geelong, Australia, the Institute for Mental and Physical Health and Clinical Translation (IMPACT), School of Medicine and Barwon Health, Geelong, Australia; Murdoch Children’s Research Institute, Parkville, VIC, Australia; Department of Medicine-Western Health, The University of Melbourne, St Albans, Victoria, Australia; Barwon Health, University Hospital, Geelong, VIC, Australia; Orygen, The National Centre of Excellence in Youth Mental Health, Centre for Youth Mental Health, Florey Institute for Neuroscience and Mental Health and the Department of Psychiatry, The University of Melbourne, Melbourne, Australia; Barwon Centre of Orthopaedic Research and Education, Deakin University, Geelong, VIC, Australia; Australian Orthopaedic Association National Joint Replacement Registry (AOANJRR), Adelaide, SA, Australia; The University of Sydney, Sydney Medical School, Sydney, NSW, Australia; Centre for Adolescent Health, Murdoch Children’s Research Institute, Melbourne, VIC, Australia; College of Public Health, Medical & Veterinary Sciences, James Cook University, Townsville, QLD, Australia

**Keywords:** gut microbiome, very low-energy diet, dietary intervention, randomised controlled trial, obesity, women

## Abstract

**Objective:** To compare the effects of consuming food-based versus supplement-based very low-energy diet (VLED) programs on gut microbiome composition in women with a high body mass index (BMI).

**Design:** An investigator-initiated, single-blind, two-arm, parallel-group randomised controlled-feeding trial with computer-generated 1:1 randomisation. From May 2021 to February 2022, women aged 30– 65 years with BMI 30–45 kg/m^2^ were recruited from southwest Victoria, Australia, and randomised to a three-week food-based or supplement-based VLED program. The primary outcome was between-group differential change in faecal microbiome alpha diversity (Shannon index) from baseline to week three, assessed using shotgun metagenomics. Outcome assessors, study investigators, and analysing statisticians were blinded to group allocation until analysis completion. Allocation concealment was managed by an independent researcher using a computer software system. Modified intention-to-treat (mITT) analyses using linear mixed-effects regression models estimated mean between-group differential changes, reported as beta-coefficient point estimates (β) and 95% confidence intervals (95%CI), adjusted for multiple comparisons.

**Results:** Forty-seven participants were randomised (food-based: n=23, supplement-based: n=24). Of the 45 participants analysed, there was a between-group differential change in the Shannon index (mITT β: 0.37, 95%CI: 0.15 to 0.60) from baseline to week three, with a greater increase in the food-based group (mean change: 0.26, 95%CI: 0.09 to 0.44; n=23) versus supplement-based group (mean change: −0.10, 95%CI: −0.25 to 0.05; n=22). There were 27 non-serious adverse events (food-based: 8, supplement-based: 19), all non-serious.

**Conclusion:** A food-based VLED, with more whole food components and fewer highly processed industrial ingredients, increases gut microbiome diversity more than a supplement-based VLED.

**Summary Box:** **What is already known on this topic**

Dietary interventions can alter gut microbiome composition, but the impact of food processing, including in nutritionally balanced very low energy diets (VLEDs), is less understood.

**What this study adds**

This study shows that a food-based VLED, with more whole food components and fewer highly processed industrial ingredients, increases gut microbiome diversity more than a supplement-based VLED.

**How this study might affect research, practice, or policy. Summarise the implications of this study**

Our findings underscore the need for further research into how specific components and attributes of diets, both including and beyond nutritional composition, influence the gut microbiome.

## Introduction

The gut microbiome is intricately connected to human health and disease ^1^. Understanding the influence of diet on its composition and function may inform gut-focussed treatment strategies ^1^. The gut microbiome is shaped by both short- and long-term dietary exposures ^2–5^. Dietary interventions, including high-fibre and Mediterranean-style diets, have been shown to beneficially alter the gut microbiome ^6^ ^7^. This includes increasing bacterial diversity and the abundances of bacterial species considered beneficial for health, enhancing carbohydrate breakdown by microbiome enzymes, and reducing inflammation ^4^ ^7^. Conversely, more “Westernised” diets, characterised by higher intakes of sugar, fat, and protein and lower intakes of fibre, are linked to reduced gut microbiome diversity and functional capacity, higher body mass index (BMI), increased inflammatory markers, elevated risk of diseases such as cancer ^8^ ^9^, and decreased hippocampal function within as little as four days ^10^.

Although dietary interventions have been linked to alterations in gut microbiome composition and potential function ^4^ ^6^ ^7^ ^11^, the effects of food processing on the gut microbiome have yet to be directly evaluated. Examples of heavily processed food items include the supplement-based shakes, bars, and soups consumed as meal replacements in very low-energy diet (VLED) programs, which are designed to provide an adequate ratio of macronutrients (proteins, fats, carbohydrates) and sufficient levels of essential vitamins and minerals, whilst limiting energy. These VLEDs, designed for individuals with a BMI of 25–30 kg/m^2^ or higher, work by restricting energy intake to approximately 800–900 kcal per day. While VLEDs have shown effectiveness in reducing weight and improving markers of type 2 diabetes and cardiovascular disease ^12^ ^13^, the impact of highly processed supplement-based VLEDs on the gut microbiome is not well understood, especially compared to food-based VLEDs ^14^. Given the crucial role of the gut microbiome in health ^1^, understanding whether differential effects of these VLEDs on the gut microbiome exist will help assess the full risk-benefit profile of these weight loss and disease prevention approaches.

This study aimed to explore the effects of a food-based versus supplement-based VLED program on gut microbiome composition and function, and physical and mental health outcomes, in women with high BMI. As preclinical models have demonstrated that diet-microbiome associations are sex-dependent ^15^, the present study was conducted entirely in women, with the next step being a larger study to determine if findings extend to both sexes.

## Materials and Methods

### Trial design

The MicroFit Study was a three-week, investigator-initiated, single-blind, two-arm, parallel-group randomised controlled-feeding trial with computer-generated 1:1 randomisation. Women with BMI 30–45 kg/m^2^ were recruited from the south-west region of Victoria, Australia. Participants were screened against eligibility criteria online via the REDCap platform ^16^ ^17^, and research assistants obtained informed consent via telephone once eligibility was confirmed. At the baseline assessment, research assistants collected anthropometric measurements to verify participants’ eligibility based on BMI. Eligible participants were randomised to consume a meal replacement program comprising either primarily food-based or supplement-based VLED options. Participants attended in-person study visits at baseline and week three at Australian Clinical Labs (ACL, Geelong, Australia) where a phlebotomist collected serum samples after an overnight fast. Participants were provided kits to collect faecal samples at home at baseline and week three, which they sent via prepaid mail to Microba Pty Ltd (Brisbane, Australia) for analysis. Questionnaire data, including sociodemographic, dietary, and health-related factors, were self-reported by participants at home using REDCap online at both study timepoints ^18^ ^19^. Participants logged their food consumption using the Easy Diet Diary application (Xyris Software Pty Ltd, Australia) every day for the entire study period. An *a priori* power calculation was conducted, based on 40 participants (20 in each arm). However, by study completion, we randomised 47 participants to account for missing baseline data and higher than anticipated dropout, and the power calculation was adjusted accordingly.

This trial received ethical approval from the Barwon Health (19/112) and Deakin University (2018/211) Human Research Ethics Committees and was registered on the Australian New Zealand Clinical Trials Registry (ACTRN12620000301965). This manuscript is presented as per the Consolidated Standards of Reporting Trials (CONSORT) statement and checklist ^20^ and gut microbiome data are reported as per the Strengthening the Organising and Reporting of Microbiome Studies (STORMS) checklist ^21^. The completed checklists and additional details are provided in the Supplementary Methods. The trial and manuscript development did not involve patients or the public owing to the absence of funding to support consumer engagement for this research.

### Recruitment and participants

Community-based recruitment was conducted from May 2021 to February 2022 using online platforms hosted by Deakin University and Barwon Health (e.g. Facebook, Instagram, Twitter, Barwon Health online newsletter), distributing flyers to local general practitioner offices, and through a variety of both paid and free online advertising services.

Inclusion criteria were: of female sex (as a means of reducing interindividual heterogeneity); aged 30– 65 years; with a BMI of 30–45 kg/m^2^; able to commit to all study procedures, including attending in-person appointments and consuming only the investigational products and recommended extras for the study duration; able to understand study materials and directions presented in English; with access to the internet and a computer, smartphone, or tablet; and able to agree to not to enrol in another clinical trial while taking part in the study.

Exclusion criteria were: currently consuming VLED products; having a diagnosed food allergy or food intolerance; receiving treatment with medications related to obesity; confirmed/suspected/planned pregnancy, or lactating; diagnosed with or having commenced a new treatment for, anxiety and/or depression within one month before baseline; having gastrointestinal disease or history of major gastrointestinal surgery; having a pre-existing cardiometabolic conditions; having had a heart attack within the past six months; having a diagnosed eating disorder; having other major medical conditions likely to have systemic effects or deemed unfit for study participation by the research team (e.g., type 2 diabetes, prediabetic, insulin resistance); regularly using opioid-based medications; regularly using recreational or illicit drugs; regularly using sodium-glucose co-transporter-2 inhibitors (i.e., gliflozins); having used antibiotics, prebiotics, and/or probiotics in the month before baseline; and having been enrolled in another clinical trial within the past three months.

### Randomisation, allocation, and blinding

Eligible participants were randomly assigned in a 1:1 ratio to either the food-based or supplement-based VLED using a computer-generated randomisation sequence with randomly ordered blocks of sizes 2 and 4. This sequence was created by a study statistician and input into REDCap online by an independent researcher to ensure allocation concealment from the study investigators. An unblinded trial coordinator enrolled and informed participants of the VLED program to which they had been assigned. The outcome assessors (research assistants) and all other study investigators, including the analysing statisticians, remained blinded to the group allocations until data analysis completion. As the study was single-blind, participants were aware of their group allocations and instructed not to discuss their allocation with the outcome assessors to preserve blinding.

### Interventions

The VLEDs were intended to be matched in overall energy (800–900 kcal per day), macronutrient profiles, sugar, sodium, and fibre. The supplement-based VLED comprised three daily total meal replacement options. Participants chose from a selection of powdered shakes and soups, bars, and desserts (16 items). On average, approximately 70% of the composition of these options was made of extracted, refined, fractionated, modified, and/or isolated proteins (e.g., calcium caseinate), carbohydrates (e.g., maltodextrin), fats (e.g., medium chain triglycerides), and fibres (e.g., fructo-oligosaccharide), as well as added vitamins (e.g., B1) and minerals (e.g., potassium citrate), and additives like emulsifiers (e.g., 472c), non-sugar sweeteners (e.g., aspartame), flavours (e.g., unspecified “flavour”), colours (e.g., curcumin), thickeners and stabilisers (e.g., vegetable gum 414). The remaining 30% consisted primarily of whole powdered milk. The food-based VLED comprised three daily total meal replacement options (55 items) and a discretionary snack (11 items). Participants chose from a selection of pre-prepared meals. On average, approximately 93% of the composition of these options was made of vegetables (e.g., green cabbage), fruits (e.g., banana), whole grains (e.g., oats), beans (e.g., cannellini beans), legumes (e.g., chickpeas), lean meats (e.g., chicken), dairy cheeses (e.g., ricotta), nuts (e.g., almond meal), seeds (e.g., flaxseed), herbs (e.g., parsley), and spices (e.g., cinnamon). The remaining 7% of the composition consisted primarily of protein isolates (e.g., whey protein isolate), as well as additives such as emulsifiers (e.g., soy lecithin), non-sugar sweeteners (e.g., stevia), flavours (e.g., vanilla extract), thickeners and stabilisers (e.g., guar gum), with approximately less than 1% of the composition of discretionary snacks including an added and isolated fibre (e.g., oligofructose) and the probiotic *Lactobacillus plantarum* (now *Lactiplantibacillus plantarum*). The full list of the options in each group and their ingredients are detailed in the Supplementary Methods.

Participants received all meal replacements without cost, delivered directly to their homes. They could choose any three meal replacements to consume at any time throughout the day. Participants were also permitted to include additional ‘recommended extras’ foods from a predetermined list (see Supplementary Methods). The supplement-based VLED group was recommended to consume at least two cups of low starch vegetables. The food-based VLED group was recommended to include one additional fruit or protein snack and three serves of side salads or vegetables. Non-sugar sweeteners, diet jelly desserts, and sugar-free lollies and gum were not recommended for daily use in the food-based VLED group, with diet cordial and diet soft drinks recommended as occasional options. The supplement-based VLED group were recommended to consume these items *ad libitum*, with no restrictions. Participants logged their daily food consumption using the Easy Diet Diary application throughout the entire study period. These data were used to monitor adherence to the approximately 900 kcal per day target and to assess macronutrient intake, which was analysed using Australian food composition databases via the FoodWorks Professional nutrient analysis software (Xyris Software Pty Ltd, Brisbane, Australia ^22^ ^23^.

### Sample size

An *a priori* sample size calculation was conducted based on 40 participants (20 in each arm) and a similar study’s sample size at the time of protocol development ^24^. As per benchmarks proposed by Cohen (1998), the *a priori* power calculation showed that the study had above 80% power with an alpha of 5% to detect moderate between-group difference effect sizes (*d*=0.91) in gut microbiome alpha diversity (Shannon index). An *a posteriori* power calculation based on 45 participants had 80% power to detect effect sizes of *d*=0.85.

### Clinical outcomes measures

Anthropometric measurements included height (stadiometer), weight (electric scales), and hip and waist circumferences (measuring tape). BMI was calculated as: BMI = (weight, kg) / (height, m)^2^. Serum inflammation markers (homocysteine, interleukin (IL)-β, IL-6, and tumor necrosis factor (TNF)-α) were assayed using the BDtm Cytometric Bead Array platform (SA Pathology, Adelaide Women’s and Children’s Hospital). Serum leptin was measured using Merck Millipore radioimmunoassay kits (Royal Prince Alfred’s Central Sydney Pathology Services). Other serum biomarkers (glucose, insulin, liver function markers (ALT, GGT, ALP, AST, total bilirubin, albumin, protein, and globulin), and lipid markers (total cholesterol, HDL, LDL, non-HDL, LDL/HDL ratio, cholesterol/HDL ratio, triglycerides) were analysed using Siemens’ ADVIA® Chemistry kits (Australian Clinical Labs, Victoria). Self-reported measures included: mental health symptoms using the Depression Anxiety Stress Scale-21 (DASS-21) ^25^, with higher scores indicating more severe symptoms; perceived well-being using the World Health Organization Wellbeing Scale (WHO-5) ^26^, with higher scores indicating better well-being; sleep-related difficulties using the Athens Insomnia Scale (AIS) ^27^, with higher scores indicating more severe issues; gastrointestinal symptoms using the Visual Analogue Scale for Irritable Bowel Syndrome (VAS-IBS) ^28^ ^29^, with higher scores indicating better outcomes; stool consistency using the Bristol Stool Form Scale (BSFS) ^30^, a 7-point scale spanning from firmest to softest stool, with mean scores estimated for each participant across one week; physical activity using the International Physical Activity Questionnaire-Short Form (IPAQ-SF) ^31^, with categorical scores (i.e., low, moderate, high) estimated as per guidelines ^32^; and habitual dietary intake at baseline using the Dietary Questionnaire for Epidemiological Studies v3.2 (DQES v3.2) ^33^. Further details are provided in the Supplementary Methods.

### Sample collection, transport, and storage

Faecal samples (∼15g) were collected by participants at baseline and week three using a Copan Italia SPA FLOQSwab in an active drying tube, including an internal desiccant to preserve samples at room temperature for up to four weeks, within 24–48 hours of their in-person appointments. Faecal samples were sent directly to Microba via post at room temperature where they were stored at –80°C until further processing. Fasted blood samples (40mL) were collected and stored at ACL as per standard procedures.

### DNA extraction

Faecal samples were extracted using the DNeasy 96 PowerSoil Pro QIAcube HT Kit (Qiagen 47021) in a 2mL deep well plate format with a modified initial processing step on the QIAcube HT DNA extraction system (Qiagen 9001793). Mechanical lysis was performed with PowerBead Pro beads (Qiagen 19311). DNA was quantified using a high-sensitivity dsDNA fluorometric assay (QuantIT, ThermoFisher, Q33120), with samples needing to reach a minimum of 0.2 ng/µL for quality control.

### Library preparation

Libraries were constructed using the Illumina DNA Prep (M) Tagmentation Kit (Illumina, 20018705) with IDT for Illumina DNA/RNA UD Index Sets A-D (Illumina 20027213-16), modified to accommodate processing in a 384-plate format. Individual libraries were pooled in equimolar amounts and assessed using a high-sensitivity dsDNA fluorometric assay (QuantIT, ThermoFisher, Q33120) and visualised with capillary gel electrophoresis using the QIAxcel DNA High Resolution Kit (Qiagen, 929002).

### Shotgun metagenomic sequencing

Samples were sequenced on the NovaSeq6000 (Illumina) using v1.5 300bp paired-end sequencing reagents. Sequence data were reviewed for yield and quality, with known control samples included in each run to monitor for background contamination. Paired-end DNA sequencing data were demultiplexed and adaptor trimmed using Illumina BaseSpace Bcl2fastq2 (v2.20) with one mismatch allowed in index sequences. Reads were quality trimmed and residual adaptors removed using Trimmomatic v0.39 with parameters: -phred33 LEADING:3 TRAILING:3 SLIDINGWINDOW:4:15 CROP:100000 HEADCROP:0 MINLEN:100. Human DNA was removed by aligning reads to the human genome reference assembly 38 (GRCh38.p12) using bwa-mem v0.7.17 with a minimum seed length of 31 (-k 31). Alignments were filtered using SAMtools v1.7, and reads mapping to the human genome with greater than 95% identity over more than 90% of the read length were flagged as human DNA and removed.

Species profiles were obtained using the Microba Community Profiler (MCP) v1.0 and the Microba Genome Database (MGDB) v1.0.3, with reads assigned to genomes within MGDB to estimate and report the relative cellular abundance of species clusters. Quantification of gene and pathway abundance was performed using the Microba Gene and Pathway Profiler (MGPP) v1.0 against the Microba Genes (MGENES) database v1.0.3. Open reading frames from genomes in MGDB were clustered against UniRef90 (release 2019/04) using MMSeqs2 with 90% identity over 80% of read length. Gene clusters were annotated with UniRef90 identifiers and linked to Enzyme Commission and Transporter Classification Database annotations via the UniProt ID mapping service. Enzyme Commission annotations were then used to determine MetaCyc pathway encoding using enrichM, with pathways classified as encoded if completeness exceeded 80%. DNA sequencing read pairs aligning with gene sequences from any protein within an MGENES protein cluster were summed. The abundances of encoded pathways for species detected by MCP were calculated by averaging the read counts of all genes for each enzyme in the pathway.

### Data preparation

Data normalisation was performed by down-sampling to a standardised number of reads before profiling within the MCP. One sample (food-based, week 3) had low read count (2,779,002) and was removed in sensitivity analyses. Alpha diversity was calculated using raw count data, with rarefaction applied to match the smallest total number of prokaryotic reads across all samples. Centred log-ratio (CLR) transformations were utilised before conducting beta-diversity and differential abundance statistical tests (species, genus, family, and phylum) given the compositional and non-normal nature of microbiome relative abundance data.

Alpha diversity, which summarises community structure within a sample, was evaluated using the Shannon index and Richness metrics. Richness quantifies the number of different species present in each sample. The Shannon index considers both the number of detected species (richness) and how evenly distributed the species are (evenness); communities with higher numbers of detected species and more even distributions of these species will result in higher Shannon index. Filtering was conducted to remove low prevalence taxa present in less than 5% of samples. Beta diversity, which summarises between-sample differences in community structure, was calculated using Aitchison distances, defined as the Euclidean distance between CLR-transformed samples, via the *stats* ^34^ package. Principal Component Analysis (PCA) was used to reduce data dimensionality for visualisation in two dimensions to identify patterns or clusters of samples within the dataset using the *Tjazi* ^35^ package.

### Statistical methods

Participant baseline characteristics were summarised with mean and standard deviation for continuous variables (or median and interquartile range where appropriate) and frequency and percentage for categorical variables. We conducted modified intention-to-treat (mITT) analyses, which included all participants who provided a baseline faecal sample. We also conducted complete case analyses, which included only participants who provided both a baseline and week three faecal sample as secondary analyses. We used linear mixed-effects regression (LMER) models to estimate between-group (food-based vs. supplement-based) differential changes (week three vs. baseline) in gut microbiome Shannon index (primary outcome) and secondary outcomes using the *lme4* ^36^ package. The models included participant as a random effect, and group allocation, nominal time point, and the interaction between diet group and time point (i.e., diet group x time point) as fixed effects. The interaction estimated the between-group differential changes from baseline to week three using beta-coefficient point estimates (β) with 95% confidence intervals (95%CI) and p-values (two-tailed; p<0.05 for significance) ^37^. The supplement-based VLED was set as the reference group. Missing covariate and secondary outcome data were imputed using predictive mean matching (five imputations) using the *mice* ^38^ package, with baseline auxiliary variables included to increase accuracy. Interaction plots of the estimated marginal means were created using the *emmeans* ^39^ package. Additionally, mean (95%CI) within-group changes are provided for descriptive purposes only and not as formal statistical tests, due to the limited sample size and low statistical power.

For beta diversity, we used complete cases to create individual CLR component-wise change scores (baseline minus week three) and then used permutational analysis of variance using adonis2 via the *vegan* ^40^ package with 999 permutations to calculate the between-group differential change (i.e., beta diversity change ∼ group). We report the R-squared (r^2^) statistic, providing a measure of the proportion of variance explained by the grouping factor (i.e., diet group) in the model.

We applied the Benjamini-Hochberg procedure ^41^ to adjust for multiple comparisons. Outcomes were grouped into related categories (e.g., gastrointestinal outcomes) and multiple comparisons testing was conducted within each category (q<0.1 for significance given the small sample size). Finally, in sensitivity analyses for the primary outcome, we adjusted for BMI due to baseline group imbalances, and removed one sample with low read count. Statistical analyses were conducted using R in the RStudio environment ^42^.

## Results

### Recruitment and trial retention

We screened 102 participants for eligibility, of whom 40 were initially randomised (**Figure 1**). Due to participant withdrawal/loss to follow-up (n=4) and missing faecal samples (n=4), we aimed to recruit an additional eight participants to reach a sample size of 40 with complete data. Overall, 47 participants were randomised, including 23 in the food-based VLED and 24 in the supplement-based VLED. Of these, 45 were included in mITT analyses (food-based: n=23, supplement-based: n=22), and 39 in complete case analysis (food-based: n=22, supplement-based: n=17) of the primary outcome.

**Figure 1.**
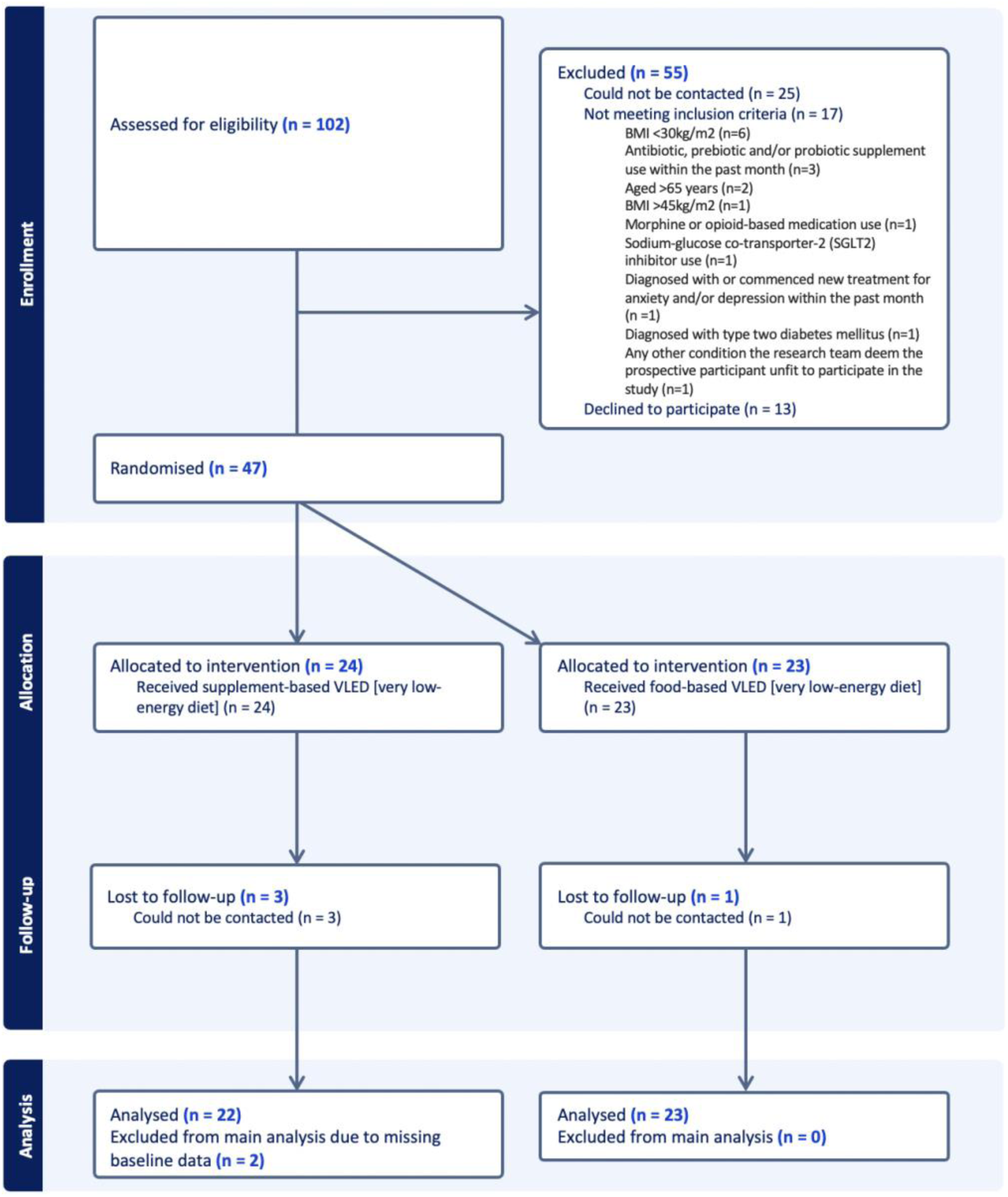
Consolidated Standards of Reporting Trials (CONSORT) flow diagram.

### Baseline characteristics

On average, participants in the food-based VLED were, less commonly married or employed compared to the supplement-based VLED group (**Table 1**). More participants in the food-based VLED were taking medication, and they also had higher average BMI, body weight, physical activity levels, and waist and hip circumferences, compared to the supplement-based VLED.

**Table 1.** Baseline characteristics of participants randomised to food-based versus supplement-based very low-energy diets.

|  | Food-based<br>VLED (N=23) | Supplement-based<br>VLED (N=24) | Total (N=47) |
| --- | --- | --- | --- |
| Sociodemographic factors |  |  |  |
| Age, years – *M* (SD) | 46.5 (10.3) | 48.3 (9.91) | 47.4 (10.0) |
| Born in Australia, *n* (%) | 22 (96%) | 19 (79%) | 41 (87%) |
| Married, *n* (%) | 15 (65%) | 19 (79%) | 34 (72%) |
| Post-secondary school education, *n* (%) | 21 (91%) | 20 (83%) | 41 (87%) |
| Employed, *n* (%) | 18 (78%) | 23 (96%) | 41 (87%) |
| Household income above \$74,999, *n* (%) | 13 (57%) | 13 (54%) | 26 (55%) |
| Health factors |  |  |  |
| Current smoker, *n* (%) | 1 (4%) | 2 (8%) | 3 (6%) |
| Any medication use, *n* (%) | 18 (78%) | 13 (59%) | 31 (69%) |
| Stool consistency – *M* (SD) | 3.8 (1.1) | 4.3 (1.2) | 4.0 (1.2) |
| Body mass index – *M* (SD) | 36.8 (3.6) | 34.8 (3.2) | 35.8 (3.5) |
| Weight, kilograms – *M* (SD) | 101 (10.1) | 94.1 (10.3) | 97.3 (10.6) |
| Waist circumference, centimeters – *M* (SD) | 107 (8.7) | 103 (9.1) | 105 (9.1) |
| Hip circumference, centimeters – *M* (SD) | 124 (9.1) | 119 (7.1) | 122 (8.5) |
| Low physical activity, *n* (%) | 13 (57%) | 18 (75%) | 31 (66%) |

Notes: Stool consistency measured using the Bristol Stool Form Scale; Low physical activity measured using the International Physical Activity Questionnaire. Abbreviations: BSFS, Bristol Stool Form Scale; *M*, Mean; *n* (%), Number and proportion of participants; VLED, very low-energy diet; SD, Standard deviation.

## ADHERENCE & SAFETY

### Diet adherence

Of the complete cases (n=39), on average, dietary intake was recorded on 20 out of the requested 21 days. The average daily energy intake of 825 kcal was within the 800–900 kcal VLED target range (**Table S1**). Both groups had similar average daily intakes of carbohydrate, protein, fibre, sugar, and sodium. The food-based VLED had higher average total energy intake (903 vs. 748 kcal/d) and total fat intake (40.1 vs. 22.5 g/d, equivalent to ∼158kcal) compared to the supplement-based VLED, respectively. The difference in average energy intake between the two groups closely matches the energy difference attributable to the variation in fat intake.

### Adverse Events

Participants in the supplement-based VLED reported more adverse events compared to the food-based VLED (19 vs. 8, respectively) (**Table S2**). The most common adverse event reported for both groups was headaches (5 vs. 3 events, respectively). No serious adverse events were reported in either group.

## PRIMARY OUTCOME

We observed a statistically significant between-group differential change in species-level Shannon index (mITT β: 0.37 95%CI: 0.15 to 0.60) (**Table S3**), with the food-based VLED group experiencing a greater increase in Shannon index (mean change: 0.27, 95%CI: 0.09 to 0.44) compared to the statistically non-significant decrease observed in the supplement-based VLED group (mean change: −0.11, −0.27 to 0.05). (**Figure 2 - A**). Results of complete case analyses (**Table S3**) and sensitivity analyses adjusting for BMI and removing a sample with low read count (**Table S11**) were similar.

**Figure 2.**
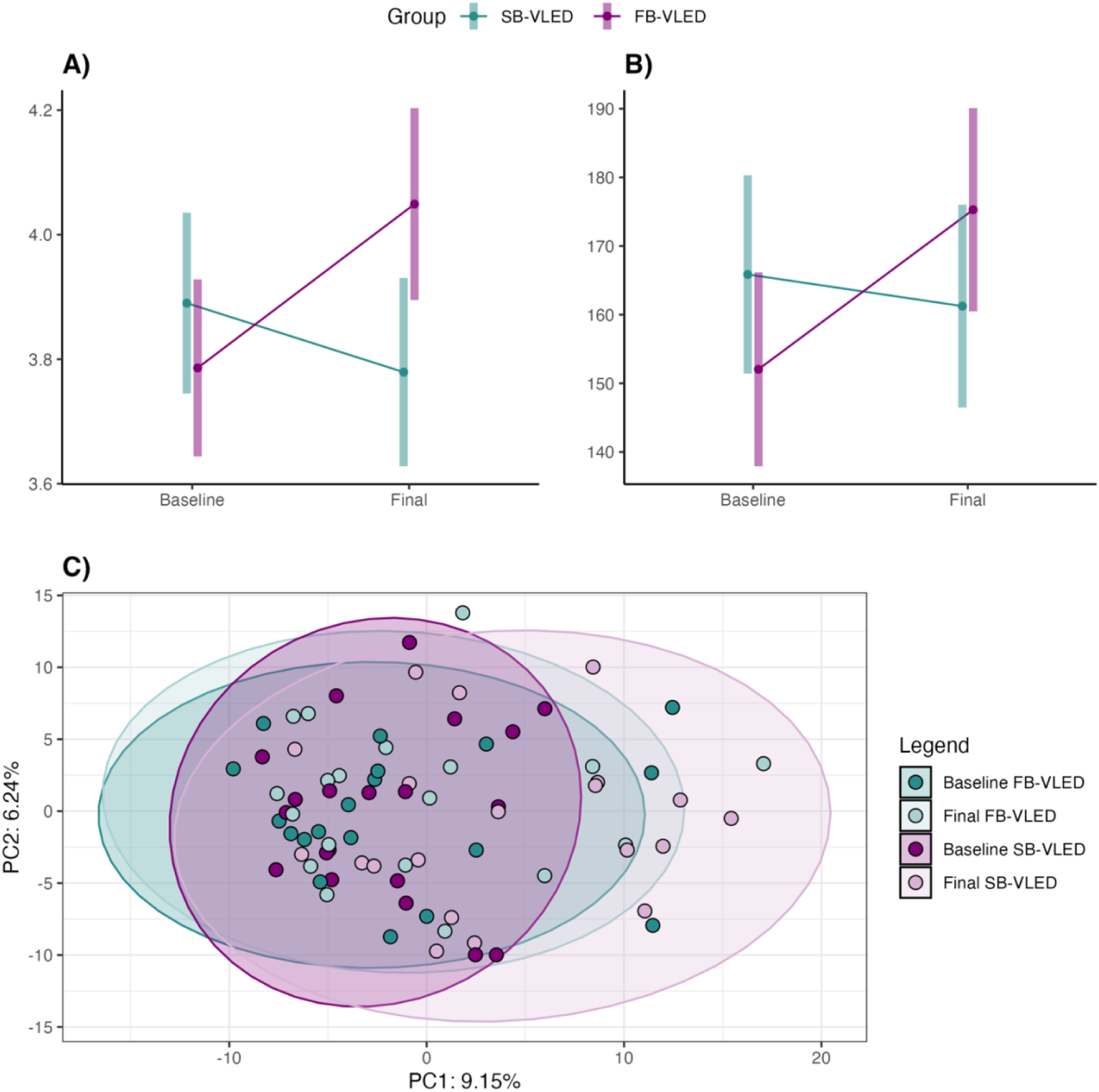
Interaction plots for the estimated marginal means of **A)** species-level Shannon index (primary outcome) and **B)** species Richness, in participants that received a food-based very low-energy diet (FB-VLED; purple) versus supplement-based very low-energy diet (SB-VLED; blue) from baseline to week three (final). Vertical bars represent 95% confidence intervals. **C)** Principal component analysis of Aitchison distances in those that received a food-based very low-energy diet (FB-VLED; purple) versus supplement-based very low-energy diet (SB-VLED; blue) at baseline and week three (final). The scatter plot shows the first two principal components (PC1 and PC2), explaining 9.15% and 6.24% of the variance, respectively. Each point represents an individual sample. Ellipses around each group represent the 95% confidence intervals, illustrating the spread and central tendency of the samples within each group. The ellipses provide a visual indication of the multivariate normal distribution of the data points, helping to identify the degree of overlap or separation between different groups.

## SECONDARY OUTCOMES

From baseline to week three, we observed a between-group differential change in species richness (mITTβ: 27.9, 12.1 to 43.7), with the food-based VLED group experiencing a greater increase in richness (mean change: 23.2, 12.7 to 33.7) compared to the statistically non-significant decrease exhibited in the supplement-based VLED group (mean change: −4.59, −17.0 to 7.78) (**Table S3; Figure 2 – B**).

We also observed a between-group differential change in beta diversity (complete case r^2^=0.051, p=0.001) (**Table S4**), suggesting that 5.1% of the difference in the shift in beta diversity observed between groups could be explained by the VLEDs. Visual inspection of the data using PCA suggested a greater shift in beta diversity in the supplement-based VLED compared to the food-based VLED (**Figure 2 – C**). The variance in Aitchison distances explained by time point was 1.8% in the food-based VLED group and 4.5% in the supplement-based VLED group (**Table S4**).

There were 619 species for analysis after prevalence filtering. Of these, we observed between-group differential changes in 72 bacterial species (**Table S5**); however, these results were not upheld after adjustment for multiple comparisons. Results of complete case analyses were similar (**Table S6**). There were between-group differential changes in 56 genera (**Table S7**), eight families (**Table S8**), three MetaCyc groups (**Table S10**), and one phylum (**Table S9**); of these, 15 genera, 2 families, 6 MetaCyc groups, and no phyla survived adjustment for multiple comparisons. For other secondary outcomes, there was a between-group differential change in constipation symptoms; however, this result was not upheld after adjustment for multiple comparisons (**Table S3**). No other between-group differential changes, including in body weight, were observed across other secondary outcomes (**Table S3**).

## Discussion

We present novel evidence of the differential effects of food-based versus supplement-based VLEDs on gut microbiome alpha diversity in women with high BMI. Additionally, there were differences in beta diversity as a secondary outcome. Compared to the supplement-based VLED group, participants in the food-based VLED exhibited greater increases in alpha diversity, a marker of better gut health in adults ^43^, and smaller shifts in beta diversity, suggesting less variability or dissimilarity in overall gut microbiome community structure between baseline and week three. We did not observe differences between the two groups in terms of changes to other secondary outcomes, including gut microbiome functional potential, body weight measures, serum metabolic and inflammatory markers, mental health, or gastrointestinal parameters.

### Limitations

Several limitations must be considered when interpreting our findings. While the single-blind nature of the trial likely had little impact on the objective gut microbiome measures, knowledge of intervention allocation may have influenced self-reported gastrointestinal and mental health symptoms. Our study was conducted in women with a BMI at or above 30 kg/m^2^, therefore findings may not be directly applicable to other populations. Our study was of short duration and VLEDs are typically followed for 12 weeks or more. Future research should involve more diverse populations and extended follow-up periods to enhance generalisability. We randomised 47 participants instead of the planned 40 to address missing data, withdrawals, and loss to follow up; however, mITT principles were maintained, and LMER models, which partially mitigate bias from missing data, showed consistent results aligned with complete case analyses. The large number of outcomes assessed increases the potential for type I errors; however, outcomes were grouped into related categories and adjustments for multiple comparisons were made for each analysis. There were differences between the groups in BMI at baseline, likely due to the small sample size, which was not large enough to balance off all confounders. Whilst results of sensitivity analyses adjusted for BMI at baseline were similar to primary analyses, larger trials are required to confirm findings. Lastly, while the two VLED programs were intended to be isocaloric and nutritionally equivalent in terms of macronutrients, differences in fat and energy intake, as well as other program aspects such as restrictions on non-sugar sweeteners and the inclusion of the probiotic *L. plantarum* in the discretionary snacks of the food-based VLED, introduce additional variability. Although a small number of food-based VLED snacks contained less than 1% of *L. plantarum*, this species was not detected in any of the faecal samples. Therefore, the possible implications of these dietary differences on our findings remains unclear.

### Implications and future directions

Our study is of potential relevance to the ongoing research and policy discussions regarding the role of ultra-processed foods in health and disease. Although our study was not specifically designed to investigate the impact of ultra-processed foods as defined by the widely used Nova food classification system ^44^, existing evidence links diets high in these foods to poorer health outcomes, including greater risks of cardiovascular disease, type 2 diabetes, common mental disorders, and mortality ^45^. However, an oft-cited limitation to the current ultra-processed food literature is a lack of causal and mechanistic understanding of these associations ^46^ ^47^. Although considered nutritionally complete, the supplement-based VLED meal replacements were comprised almost entirely of highly processed industrial ingredients, including substances extracted from foods (such as combinations of macronutrient isolates like calcium caseinate, sodium caseinate, and medium chain triglycerides), as well as additives (such as emulsifiers and non-sugar sweeteners like 472c and aspartame, respectively). Conversely, the food-based VLED meal replacements primarily consisted of vegetables, whole grains, and legumes, with far fewer heavily processed industrial ingredients. Emerging evidence suggests that additives such as emulsifiers ^48^ and non-nutritive sweeteners ^49^ may adversely impact the gut microbiome and related health. In contrast, intact food structures and matrices, along with polyphenols and phytonutrients and diverse sources of fibre found in whole food components may positively modulate the gut microbiome ^50^ ^51^. Acknowledging the limitations of our study, the findings support future research specifically designed to investigate the gut microbiome as a potential mechanism through which diets high in ultra-processed foods impact health outcomes.

## Conclusion

Our study provides novel evidence of the differential impacts of food-based versus supplement-based VLEDs on the diversity of the gut microbiome of women with high BMI. Our findings show that a food-based VLED, with greater whole food components and fewer highly processed industrial ingredients, increases gut microbiome diversity more than a supplement-based VLED. Further research is needed to better understand specific components or attributes of these diets responsible for the observed differential effects.

## Data Availability

To access additional data from this study, the code and raw data can be found on GitHub and NCBI Sequence Read Archive, respectively. For further assistance or inquiries, please contact the corresponding author.

## Contributors

## Acknowledgements

We would like to thank Dr Jerry Lai for their assistance with REDCap, Dr Jessica Davis and Samantha Kernaghan for their assistance with data collection, and Chantelle Erwin for their assistance with data cleaning of dietary data.

## Ethical approval

The study was conducted following the principles of the Declaration of Helsinki and had ethical approval from the Barwon Health (19/112) and Deakin University (2018/211) Human Research Ethics Committees.

## Data sharing

To access additional data from this study, the code and raw data can be found on GitHub and NCBI Sequence Read Archive, respectively. For further assistance or inquiries, please contact the corresponding author at

## Transparency

The corresponding (the manuscript’s guarantor) affirms that the manuscript is an honest, accurate, and transparent account of the study being reported; that no important aspects of the study have been omitted; and that any discrepancies from the study as planned (and, if relevant, registered) have been explained.

## Dissemination to participants and related patient and public communities

We will share our review findings with academic, clinical, policy, and public audiences post-publication through various channels, including conferences as well as personal and institutional media hosted primarily by Deakin University and the Food & Mood Centre.

## Provenance and peer review

Provenance and peer review: Not commissioned; externally peer reviewed.

The default licence, a CC BY NC licence, is needed.

