## Supplementary material for "The effects of food-based versus supplement-based very low-energy diets on gut microbiome composition and health outcomes in women with high body mass index (The MicroFit Study): a randomised controlled trial": Table S1

**Table S1.** Average daily energy and nutrient intakes in the food-based and supplement-based very low-energy diets across the three-week intervention period using complete cases (n=39).

| | **FB-VLED (N=20)**<br>**Mean (SD)** | **SB-VLED (N=19)**<br>**Mean (SD)** | **Overall (N=39)**<br>**Mean (SD)** |
|---|---|---|---|
| Total reported days | 20 (3.0) | 20.3 (2.7) | 20.2 (2.8) |
| Energy [a] , kilocalories/day | 903 (249) | 748 (137) | 825 (211) |
| kilojoules/day | 3780 (1040) | 3130 (575) | 3450 (883) |
| Carbohydrate, grams/day | 66.9 (17.2) | 64.0 (10.9) | 66.6 (18.2) |
| Total fat, grams/day | 40.1 (12.1) | 22.5 (6.95) | 31.0 (13.1) |
| Protein, grams/day | 63.9 (23.3) | 69.1 (11.5) | 65.4 (14.2) |
| Fibre, grams/day | 22.6 (8.49) | 20.0 (3.11) | 21.3 (6.37) |
| Total sugar, grams/day | 38.1 (14.9) | 37.3 (9.35) | 37.7 (12.2) |
| Sodium, milligrams/day | 1260 (343) | 1390 (306) | 1330 (328) |

[a] Energy intake including dietary fibre.

Abbreviations: FB-VLED, food-based very low-energy diet; SB-VLED, supplement-based very low-energy diet; SD, standard deviation.
