## Supplementary material for "The effects of food-based versus supplement-based very low-energy diets on gut microbiome composition and health outcomes in women with high body mass index (The MicroFit Study): a randomised controlled trial": Table S2

**Table S2.** Non-serious adverse events reported in the food-based and supplement-based very low-energy diet groups over three weeks.

| | **FB-VLED (n=23)** | **SB-VLED (n=24)** | **Overall (n=47)** |
|---|---|---|---|
| Headache | 3 | 5 | 8 |
| Gastrointestinal aberrations | 2 | 2 | 4 |
| Fatigue | 1 | 1 | 2 |
| Dizziness | 0 | 1 | 1 |
| Delayed menstrual cycle | 0 | 1 | 1 |
| Migraine | 1 | 0 | 1 |
| Acne | 1 | 0 | 1 |
| Hormonal aberrations | 0 | 1 | 1 |
| Emotional dysregulation | 0 | 1 | 1 |
| Water retention | 0 | 1 | 1 |
| Urinary tract infection | 0 | 1 | 1 |
| Hypothyroidism | 0 | 1 | 1 |
| Acute viral rhinosinusitis | 1 | 0 | 1 |
| Herpes labialis | 0 | 1 | 1 |
| Paraesthesia | 0 | 1 | 1 |
| Xerosis | 0 | 1 | 1 |
| **Total adverse events** | 9 | 18 | 27 |
| *Abbreviations: FB-VLED, food-based very low-energy diet; SB-VLED, supplement-based very low-energy diet* | | | |
