## Supplementary material for "The effects of food-based versus supplement-based very low-energy diets on gut microbiome composition and health outcomes in women with high body mass index (The MicroFit Study): a randomised controlled trial": Table S3

**Table S3.** Unadjusted between-group differential changes in alpha diversity (primary outcome) and secondary outcomes in those that consumed a food-based versus supplement-based very low-energy diet for three weeks using modified intention-to-treat and complete case analysis.

| | Within-group change from baseline to week three Mean (95%CI) | | Between-group differential changes from baseline to week three | | |
|---|---|---|---|---|---|
| | Food-based VLED | Supplement-based VLED | Unadjusted β (95%CI) | p-value | q-value |
| **ALPHA DIVERSITY** | | | | | |
| Shannon index | | | | | |
| ITT (n=45) | 0.27 (0.09 to 0.44) | −0.11 (−0.27 to 0.04) | **0.37 (0.15 to 0.60)** | **0.002** | **0.002** |
| CC (n=39) | 0.25 (0.07 to 0.43) | −0.11 (−0.27 to 0.05) | **0.35 (0.12 to 0.59)** | **0.004** | **0.004** |
| Richness | | | | | |
| ITT (n=45) | 23.2 (12.7 to 33.7) | −4.59 (−17.0 to 7.78) | **27.9 (12.1 to 43.7)** | **0.001** | **0.002** |
| CC (n=39) | 22.4 (11.9 to 32.9) | −5.10 (−17.6 to 7.44) | **27.5 (11.6 to 43.4)** | **0.001** | **0.002** |
| **ANTHROPOMETRICS** | | | | | |
| Weight, kg | | | | | |
| ITT (n=45) | −4.58 (−5.58 to −3.58) | −4.85 (−5.67 to −4.03) | 0.27 (−1.01 to 1.56) | 0.669 | 0.669 |
| CC (n=41) | −4.57 (−5.58 to −3.57) | −4.85 (−5.68 to −4.03) | 0.28 (−1.01 to 1.57) | 0.662 | 0.662 |
| Body mass index, $kg/m^2$ | | | | | |
| ITT (n=45) | −1.66 (−2.00 to −1.31) | −1.79 (−2.09 to −1.49) | 0.13 (−0.32 to 0.58) | 0.555 | 0.669 |
| CC (n=41) | −1.65 (−2.00 to −1.31) | −1.79 (−2.09 to −1.49) | 0.14 (−0.31 to 0.59) | 0.541 | 0.662 |
| Hip circumference, cm | | | | | |
| ITT (n=45) | −1.18 (−3.40 to 1.03) | −4.53 (−6.87 to −2.20) | **3.29 (0.17 to 6.42)** | **0.039** | 0.157 |
| CC (n=40) | −1.13 (−3.35 to 1.10) | −4.43 (−6.80 to −2.05) | **3.30 (0.15 to 6.45)** | **0.041** | 0.162 |
| Waist circumference, cm | | | | | |
| ITT (n=45) | −4.23 (−7.11 to −1.35) | −5.50 (−8.36 to −2.64) | 1.27 (−2.68 to 5.22) | 0.520 | 0.669 |
| CC (n=40) | −4.28 (−7.20 to −1.36) | −5.48 (−8.40 to −2.57) | 1.20 (−2.81 to 5.21) | 0.548 | 0.662 |
| **BLOOD BIOMARKERS: LIVER FUNCTION** | | | | | |
| Alanine transaminase, g/L | | | | | |
| ITT (n=45) | 0.20 (−2.66 to 3.07) | 1.96 (−6.30 to 10.2) | −1.92 (−9.59 to 5.74) | 0.613 | 0.833 |
| CC (n=40) | 0.05 (−2.84 to 2.93) | 3.16 (−4.08 to 10.4) | −3.11 (−10.4 to 4.16) | 0.392 | 0.814 |
| Aspartate transaminase, g/L | | | | | |
| ITT (n=45) | 0.69 (−1.40 to 2.79) | 2.79 (−0.59 to 6.16) | −2.10 (−5.92 to 1.72) | 0.271 | 0.720 |
| CC (n=40) | 0.52 (−1.59 to 2.64) | 3.05 (−0.23 to 6.34) | −2.53 (−6.24 to 1.18) | 0.176 | 0.814 |
| Alkaline phosphatase, g/L | | | | | |
| ITT (n=45) | −6.70 (−12.3 to −1.06) | −6.96 (−11.1 to −2.85) | 0.36 (−6.54 to 7.26) | 0.917 | 0.917 |
| CC (n=40) | −6.81 (−12.5 to −1.09) | −6.79 (−10.9 to −2.65) | −0.02 (−6.99 to 6.95) | 0.995 | 0.995 |
| Gamma-glutamyltransferase, g/L | | | | | |
| ITT (n=45) | −4.22 (−10.6 to 2.18) | −6.89 (−11.2 to −2.59) | 2.69 (−4.96 to 10.3) | 0.481 | 0.833 |
| CC (n=40) | −4.48 (−10.9 to 1.99) | −6.74 (−11.0 to −2.49) | 2.26 (−5.41 to 9.93) | 0.555 | 0.814 |
| Albumin, g/L | | | | | |
| ITT (n=45) | 1.32 (0.48 to 2.15) | 2.91 (1.90 to 3.92) | **−1.60 (−2.86 to −0.33)** | **0.015** | 0.323 |
| CC (n=40) | 1.33 (0.49 to 2.18) | 2.89 (1.85 to 3.94) | **−1.56 (−2.85 to −0.27)** | **0.019** | 0.416 |

| | Within-group change from baseline to week three Mean (95%CI) | | Between-group differential changes from baseline to week three | | |
|---|---|---|---|---|---|
| | Food-based VLED | Supplement-based VLED | Unadjusted β (95%CI) | p-value | q-value |
| Globulin, g/L | | | | | |
| ITT (n=45) | −1.51 (−2.37 to −0.64) | −0.88 (−2.26 to 0.51) | −0.60 (−2.16 to 0.97) | 0.446 | 0.833 |
| CC (n=40) | −1.57 (−2.44 to −0.70) | −0.84 (−2.27 to 0.59) | −0.73 (−2.32 to 0.86) | 0.358 | 0.814 |
| Bilirubin, µmol/L | | | | | |
| ITT (n=45) | 0.60 (−0.30 to 1.51) | 0.97 (0.04 to 1.90) | −0.36 (−1.62 to 0.90) | 0.570 | 0.833 |
| CC (n=40) | 0.57 (−0.34 to 1.49) | 0.95 (0.01 to 1.89) | −0.38 (−1.65 to 0.90) | 0.554 | 0.814 |
| Total protein, g/L | | | | | |
| ITT (n=45) | −0.13 (−1.53 to 1.27) | 1.51 (−0.01 to 3.02) | −1.62 (−3.62 to 0.38) | 0.109 | 0.720 |
| CC (n=40) | −0.24 (−1.66 to 1.18) | 1.53 (−0.04 to 3.09) | −1.76 (−3.81 to 0.28) | 0.089 | 0.814 |
| **BLOOD BIOMARKERS: LIPIDS** | | | | | |
| Total cholesterol, mmol/L | | | | | |
| ITT (n=45) | −0.89 (−1.22 to −0.56) | −1.09 (−1.33 to −0.85) | 0.20 (−0.20 to 0.61) | 0.319 | 0.720 |
| CC (n=40) | −0.91 (−1.25 to −0.57) | −1.08 (−1.32 to −0.84) | 0.17 (−0.24 to 0.58) | 0.408 | 0.814 |
| Triglycerides, mmol/L | | | | | |
| ITT (n=45) | −0.31 (−0.49 to −0.12) | −0.47 (−0.77 to −0.17) | 0.16 (−0.17 to 0.50) | 0.327 | 0.720 |
| CC (n=40) | −0.32 (−0.51 to −0.14) | −0.44 (−0.73 to −0.14) | 0.11 (−0.22 to 0.44) | 0.494 | 0.814 |
| HDL cholesterol, mmol/L | | | | | |
| ITT (n=45) | −0.22 (−0.31 to −0.13) | −0.30 (−0.40 to −0.21) | 0.08 (−0.05 to 0.21) | 0.210 | 0.720 |
| CC (n=40) | −0.23 (−0.32 to −0.14) | −0.30 (−0.39 to −0.21) | 0.07 (−0.06 to 0.20) | 0.264 | 0.814 |
| LDL cholesterol, mmol/L | | | | | |
| ITT (n=45) | −0.52 (−0.82 to −0.22) | −0.56 (−0.77 to −0.36) | 0.04 (−0.32 to 0.40) | 0.833 | 0.881 |
| CC (n=40) | −0.53 (−0.83 to −0.22) | −0.57 (−0.78 to −0.36) | 0.04 (−0.33 to 0.40) | 0.827 | 0.946 |
| Non-HDL cholesterol, mmol/L | | | | | |
| ITT (n=45) | −0.67 (−1.00 to −0.33) | −0.79 (−1.03 to −0.55) | 0.12 (−0.28 to 0.53) | 0.541 | 0.833 |
| CC (n=40) | −0.68 (−1.02 to −0.34) | −0.78 (−1.02 to −0.54) | 0.10 (−0.32 to 0.51) | 0.635 | 0.874 |
| LDL:HDL ratio, mmol/L | | | | | |
| ITT (n=45) | −0.07 (−0.39 to 0.25) | −0.03 (−0.29 to 0.22) | −0.04 (−0.44 to 0.36) | 0.841 | 0.881 |
| CC (n=40) | −0.06 (−0.39 to 0.27) | −0.04 (−0.30 to 0.23) | −0.03 (−0.44 to 0.39) | 0.903 | 0.946 |
| Cholesterol:ratio, mmol/L | | | | | |
| ITT (n=45) | −0.11 (−0.47 to 0.25) | −0.04 (−0.33 to 0.24) | −0.07 (−0.52 to 0.38) | 0.768 | 0.881 |
| CC (n=40) | −0.10 (−0.47 to 0.26) | −0.05 (−0.33 to 0.24) | −0.06 (−0.52 to 0.40) | 0.802 | 0.946 |
| **BLOOD BIOMARKERS: INFLAMMATORY MARKERS** | | | | | |
| Interleukin-1b, pg/mL | | | | | |
| ITT (n=45) | 0.21 (−0.37 to 0.79) | −0.03 (−0.81 to 0.76) | 0.23 (−0.73 to 1.20) | 0.634 | 0.833 |
| CC (n=39) | 0.25 (−0.36 to 0.85) | −0.08 (−0.92 to 0.76) | 0.33 (−0.69 to 1.36) | 0.521 | 0.814 |
| Interleukin-6, pg/mL | | | | | |
| ITT (n=45) | 0.94 (−5.24 to 7.12) | 5.16 (−0.66 to 11.0) | −4.24 (−12.7 to 4.20) | 0.317 | 0.720 |
| CC (n=39) | 0.89 (−5.74 to 7.52) | 5.11 (−1.02 to 11.3) | −4.22 (−13.2 to 4.74) | 0.346 | 0.814 |
| Tumor Necrosis Factor-alpha, pg/mL | | | | | |
| ITT (n=45) | 0.03 (−0.17 to 0.22) | 0.17 (0.07 to 0.28) | −0.14 (−0.36 to 0.08) | 0.195 | 0.720 |
| CC (n=39) | 0.03 (−0.17 to 0.23) | 0.16 (0.06 to 0.27) | −0.13 (−0.36 to 0.09) | 0.230 | 0.814 |

| | Within-group change from baseline to week three Mean (95%CI) | | Between-group differential changes from baseline to week three | | |
|---|---|---|---|---|---|
| | Food-based VLED | Supplement-based VLED | Unadjusted β (95%CI) | p-value | q-value |
| Homocysteine, (μmol/L) | | | | | |
| ITT (n=45) | 0.64 (−0.37 to 1.66) | 1.40 (0.49 to 2.31) | −0.71 (−2.04 to 0.61) | 0.284 | 0.720 |
| CC (n=39) | 0.57 (−0.47 to 1.60) | 1.29 (0.36 to 2.22) | −0.73 (−2.07 to 0.62) | 0.283 | 0.814 |
| **BLOOD BIOMARKERS: METABOLIC MARKERS** | | | | | |
| Leptin, ng/mL | | | | | |
| ITT (n=45) | −41.6 (−57.2 to −26.1) | −47.4 (−67.5 to −27.3) | 5.46 (−18.3 to 29.2) | 0.644 | 0.833 |
| CC (n=40) | −42.6 (−58.2 to −26.9) | −44.9 (−63.2 to −26.7) | 2.37 (−20.8 to 25.5) | 0.837 | 0.946 |
| Glucose, mmol/L | | | | | |
| ITT (n=45) | −0.32 (−0.57 to −0.08) | −0.26 (−0.47 to −0.05) | −0.06 (−0.37 to 0.26) | 0.728 | 0.881 |
| CC (n=40) | −0.29 (−0.54 to −0.04) | −0.26 (−0.48 to −0.05) | −0.03 (−0.35 to 0.30) | 0.865 | 0.946 |
| Insulin, mU/L | | | | | |
| ITT (n=45) | −2.55 (−4.97 to −0.13) | −4.82 (−6.98 to −2.67) | 2.19 (−0.98 to 5.36) | 0.171 | 0.720 |
| CC (n=40) | −2.71 (−5.17 to −0.26) | −5.05 (−7.23 to −2.87) | 2.34 (−0.87 to 5.55) | 0.149 | 0.814 |
| **MENTAL HEALTH** | | | | | |
| Total DASS | | | | | |
| ITT (n=45) | −5.21 (−8.76 to −1.67) | −2.26 (−5.48 to 0.96) | −2.94 (−7.61 to 1.73) | 0.210 | 0.397 |
| CC (n=40) | −4.95 (−8.47 to −1.44) | −2.26 (−5.53 to 1.00) | −2.69 (−7.37 to 1.99) | 0.253 | 0.492 |
| DASS Depression subscale | | | | | |
| ITT (n=45) | −1.52 (−3.07 to 0.02) | −0.84 (−2.32 to 0.65) | −0.69 (−2.78 to 1.40) | 0.508 | 0.610 |
| CC (n=40) | −1.43 (−2.97 to 0.11) | −0.84 (−2.37 to 0.69) | −0.59 (−2.69 to 1.52) | 0.577 | 0.692 |
| DASS Anxiety subscale | | | | | |
| ITT (n=45) | −1.93 (−2.88 to −0.98) | −0.99 (−2.15 to 0.16) | −0.95 (−2.39 to 0.49) | 0.189 | 0.397 |
| CC (n=40) | −1.86 (−2.80 to −0.91) | −1.00 (−2.17 to 0.17) | −0.86 (−2.30 to 0.58) | 0.236 | 0.492 |
| DASS Stress subscale | | | | | |
| ITT (n=45) | −1.73 (−3.11 to −0.35) | −0.43 (−1.25 to 0.39) | −1.27 (−2.85 to 0.30) | 0.111 | 0.397 |
| CC (n=40) | −1.67 (−3.03 to −0.31) | −0.42 (−1.25 to 0.41) | −1.25 (−2.83 to 0.34) | 0.120 | 0.492 |
| WHO-5 Well-being Index | | | | | |
| ITT (n=45) | 14.7 (7.07 to 22.4) | 8.95 (1.80 to 16.1) | 5.68 (−4.46 to 15.8) | 0.265 | 0.397 |
| CC (n=40) | 14.1 (6.52 to 21.7) | 9.05 (1.66 to 16.5) | 5.04 (−5.25 to 15.3) | 0.328 | 0.492 |
| Athens Insomnia Scale | | | | | |
| ITT (n=45) | −2.44 (−4.09 to −0.80) | −2.38 (−4.04 to −0.72) | −0.07 (−2.34 to 2.20) | 0.952 | 0.952 |
| CC (n=40) | −2.43 (−4.11 to −0.75) | −2.47 (−4.20 to −0.75) | 0.05 (−2.29 to 2.38) | 0.969 | 0.969 |
| **GASTROINTESTINAL HEALTH** | | | | | |
| Bristol stool total score | | | | | |
| ITT (n=45) | −0.31 (−0.93 to 0.32) | −0.60 (−1.35 to 0.14) | 0.28 (−0.66 to 1.22) | 0.549 | 0.713 |
| CC (n=40) | −0.29 (−0.93 to 0.36) | −0.74 (−1.50 to 0.02) | 0.45 (−0.51 to 1.41) | 0.348 | 0.723 |
| Abdominal pain | | | | | |
| ITT (n=45) | 11.6 (3.01 to 20.1) | −1.08 (−13.6 to 11.4) | 12.6 (−1.86 to 27.1) | 0.086 | 0.429 |
| CC (n=40) | 10.5 (1.95 to 19.0) | 0.95 (−11.9 to 13.8) | 9.53 (−5.11 to 24.2) | 0.196 | 0.723 |
| Diarrhoea | | | | | |

| | Within-group change from baseline to week three Mean (95%CI) | | Between-group differential changes from baseline to week three | | |
|---|---|---|---|---|---|
| | Food-based VLED | Supplement-based VLED | Unadjusted β (95%CI) | p-value | q-value |
| ITT (n=45) | 2.58 (−8.54 to 13.7) | −6.75 (−22.5 to 9.04) | 9.27 (−9.29 to 27.8) | 0.320 | 0.533 |
| CC (n=40) | 2.52 (−8.96 to 14.0) | −4.21 (−20.9 to 12.5) | 6.73 (−12.6 to 26.0) | 0.485 | 0.723 |
| Constipation | | | | | |
| ITT (n=45) | 1.95 (−6.20 to 10.1) | −18.5 (−33.1 to −3.98) | **19.5 (3.59 to 35.5)** | **0.017** | 0.175 |
| CC (n=40) | 2.38 (−5.88 to 10.6) | −16.3 (−31.6 to −1.08) | **18.7 (2.35 to 35.0)** | **0.026** | 0.260 |
| Bloating/flatulence | | | | | |
| ITT (n=45) | 20.3 (9.33 to 31.2) | 7.23 (−9.61 to 24.1) | 12.8 (−6.40 to 32.0) | 0.185 | 0.446 |
| CC (n=40) | 19.1 (8.14 to 30.0) | 10.7 (−6.47 to 27.8) | 8.36 (−10.9 to 27.7) | 0.386 | 0.723 |
| Vomiting/nausea | | | | | |
| ITT (n=45) | 8.63 (−0.48 to 17.7) | 0.14 (−6.32 to 6.61) | 8.46 (−2.74 to 19.7) | 0.134 | 0.446 |
| CC (n=40) | 7.29 (−1.61 to 16.2) | 1.00 (−5.74 to 7.74) | 6.29 (−4.71 to 17.3) | 0.255 | 0.723 |
| Psychological wellbeing | | | | | |
| ITT (n=45) | 12.1 (3.69 to 20.5) | 7.90 (−6.55 to 22.4) | 4.54 (−11.5 to 20.6) | 0.571 | 0.713 |
| CC (n=40) | 11.3 (2.92 to 19.8) | 8.68 (−6.30 to 23.67) | 2.65 (−13.6 to 18.9) | 0.743 | 0.826 |
| Influence on daily life | | | | | |
| ITT (n=45) | 20.0 (7.39 to 32.6) | 7.39 (−9.68 to 24.5) | 13.1 (−8.26 to 34.4) | 0.223 | 0.446 |
| CC (n=40) | 17.8 (5.56 to 30.1) | 10.7 (−7.15 to 28.6) | 7.07 (−14.2 to 28.4) | 0.506 | 0.723 |
| Urgency to defecation | | | | | |
| ITT (n=45) | −0.15 (−0.40 to 0.10) | −0.19 (−0.48 to 0.09) | 0.25 (−1.76 to 2.26) | 0.810 | 0.877 |
| CC (n=40) | −0.14 (−0.40 to 0.11) | −0.21 (−0.51 to 0.08) | 0.36 (−1.71 to 2.42) | 0.735 | 0.826 |
| Incomplete defecation | | | | | |
| ITT (n=45) | 0.04 (−0.22 to 0.29) | 0.06 (−0.18 to 0.30) | −0.17 (−2.28 to 1.95) | 0.877 | 0.877 |
| CC (n=40) | 0.05 (−0.21 to 0.31) | 0.05 (−0.19 to 0.30) | −0.02 (−2.20 to 2.16) | 0.987 | 0.987 |
| **METACYC GROUP ALPHA DIVERSITY** | | | | | |
| MetaCyc Shannon index | | | | | |
| ITT (n=45) | 0.01 (−0.01 to 0.03) | 0.01 (−0.01 to 0.03) | 0.00 (−0.03 to 0.02) | 0.764 | 0.764 |
| CC (n=39) | 0.01 (−0.01 to 0.03) | 0.01 (−0.01 to 0.03) | 0.01 (−0.01 to 0.03) | 0.187 | 0.375 |
| MetaCyc richness | | | | | |
| ITT (n=45) | −0.17 (−1.24 to 0.89) | 1.05 (−0.46 to 2.55) | −1.21 (−3.02 to 0.61) | 0.187 | 0.375 |
| CC (n=39) | −0.13 (−1.21 to 0.95) | 0.91 (−0.65 to 2.46) | 0.40 (−0.54 to 1.34) | 0.393 | 0.393 |
