## Supplementary material for "The effects of food-based versus supplement-based very low-energy diets on gut microbiome composition and health outcomes in women with high body mass index (The MicroFit Study): a randomised controlled trial": Table S4

**Table S4.** Between-group differential change in beta diversity (complete cases n=39) in those that consumed a food-based versus supplement-based very low energy diet for three weeks.

| | Aitchison distance | | | | |
|---|---|---|---|---|---|
| | Df | Sum of squares | $R^2$ | F | p-value |
| **Between-group differential change** | | | | | |
| Group | 1 | 783.8 | 0.051 | 1.977 | **0.001** |
| Residual | 37 | 14648 | 0.949 | NA | NA |
| Total | 38 | 15431 | 1.000 | NA | NA |
| **Within-group: food-based VLED group** | | | | | |
| Time point | 1 | 305.2 | 0.018 | | |
| Residual | 36 | 16636 | 0.982 | | |
| Total | 37 | 16941 | 1 | | |
| **Within-group: supplement-based VLED group** | | | | | |
| Time point | 1 | 785.8 | 0.045 | | |
| Residual | 38 | 16828 | 0.955 | | |
| Total | 39 | 17614 | 1 | | |

Between-group model: beta_change_score ~ group

Within-group model: beta_score ~ time_point

Abbreviations: Df, degrees of freedom.
