## Supplementary material for "The effects of food-based versus supplement-based very low-energy diets on gut microbiome composition and health outcomes in women with high body mass index (The MicroFit Study): a randomised controlled trial": Table S5

**Table S5.** Unadjusted modified intention-to-treat analysis (n=45) of the differential changes in bacterial species between those that consumed a food-based versus supplement-based very low-energy diet for three weeks.

| OUTCOMES - SPECIES | EFFECT | GROUP | TERM | ESTIMATE | STD ERROR | STATISTIC | DF | CONF LOW | CONF HIGH | P VALUE | Q VALUE |
|---|---|---|---|---|---|---|---|---|---|---|---|
| Oscillibacter sp001916835 | fixed | NA | groupFBVLED:time_pointFinal | 0.72 | 0.19 | 3.77 | 41.06 | 0.34 | 1.11 | **0.001** | 0.204 |
| Faecalibacterium prausnitzii_G | fixed | NA | groupFBVLED:time_pointFinal | 1.25 | 0.37 | 3.40 | 41.82 | 0.51 | 2.00 | **0.001** | 0.204 |
| Gemmiger sp003476825 | fixed | NA | groupFBVLED:time_pointFinal | 1.26 | 0.38 | 3.35 | 40.76 | 0.50 | 2.02 | **0.002** | 0.204 |
| UBA11524 sp000437595 | fixed | NA | groupFBVLED:time_pointFinal | 1.22 | 0.38 | 3.22 | 42.19 | 0.45 | 1.98 | **0.002** | 0.204 |
| GCA 900066135 MIC6659 | fixed | NA | groupFBVLED:time_pointFinal | 0.76 | 0.24 | 3.21 | 41.20 | 0.28 | 1.24 | **0.003** | 0.204 |
| Anaerostipes hadrus | fixed | NA | groupFBVLED:time_pointFinal | -1.20 | 0.38 | -3.17 | 40.93 | -1.96 | -0.43 | **0.003** | 0.204 |
| UBA11774 sp003507655 | fixed | NA | groupFBVLED:time_pointFinal | 1.33 | 0.43 | 3.12 | 43.73 | 0.47 | 2.19 | **0.003** | 0.204 |
| GCA 900066905 sp900066905 | fixed | NA | groupFBVLED:time_pointFinal | -0.36 | 0.12 | -3.00 | 84.00 | -0.60 | -0.12 | **0.004** | 0.204 |
| Lawsonibacter sp900066645 | fixed | NA | groupFBVLED:time_pointFinal | -0.36 | 0.12 | -3.06 | 44.47 | -0.59 | -0.12 | **0.004** | 0.204 |
| Agathobacter faecis | fixed | NA | groupFBVLED:time_pointFinal | 1.58 | 0.51 | 3.09 | 37.86 | 0.55 | 2.62 | **0.004** | 0.204 |
| Lachnospiraceae MIC8879 | fixed | NA | groupFBVLED:time_pointFinal | 0.56 | 0.18 | 3.06 | 42.23 | 0.19 | 0.93 | **0.004** | 0.204 |
| Ruminococcus_A sp003011855 | fixed | NA | groupFBVLED:time_pointFinal | -0.91 | 0.30 | -3.04 | 41.41 | -1.52 | -0.31 | **0.004** | 0.204 |
| Streptococcus thermophilus | fixed | NA | groupFBVLED:time_pointFinal | 1.65 | 0.55 | 2.99 | 44.02 | 0.54 | 2.75 | **0.005** | 0.204 |
| Alistipes MIC8513 | fixed | NA | groupFBVLED:time_pointFinal | 0.37 | 0.13 | 2.96 | 40.44 | 0.12 | 0.62 | **0.005** | 0.204 |
| Lachnospiraceae MIC9331 | fixed | NA | groupFBVLED:time_pointFinal | -0.65 | 0.22 | -2.93 | 46.37 | -1.09 | -0.20 | **0.005** | 0.204 |
| ER4 sp000765235 | fixed | NA | groupFBVLED:time_pointFinal | 0.86 | 0.30 | 2.93 | 41.48 | 0.27 | 1.46 | **0.006** | 0.204 |
| Eisenbergiella massiliensis | fixed | NA | groupFBVLED:time_pointFinal | -0.59 | 0.21 | -2.84 | 84.00 | -1.00 | -0.18 | **0.006** | 0.204 |
| Faecalibacterium prausnitzii_D | fixed | NA | groupFBVLED:time_pointFinal | 1.09 | 0.38 | 2.89 | 39.88 | 0.33 | 1.85 | **0.006** | 0.204 |
| Faecalicatena lactaris | fixed | NA | groupFBVLED:time_pointFinal | 0.99 | 0.34 | 2.89 | 39.32 | 0.30 | 1.68 | **0.006** | 0.204 |
| Oscillibacter MIC9597 | fixed | NA | groupFBVLED:time_pointFinal | 0.53 | 0.19 | 2.82 | 44.69 | 0.15 | 0.90 | **0.007** | 0.222 |
| Clostridium_A leptum | fixed | NA | groupFBVLED:time_pointFinal | -0.81 | 0.29 | -2.80 | 44.78 | -1.39 | -0.23 | **0.008** | 0.223 |
| Dorea sp000433215 | fixed | NA | groupFBVLED:time_pointFinal | 0.53 | 0.19 | 2.78 | 40.44 | 0.15 | 0.92 | **0.008** | 0.226 |
| Eubacterium_F sp000433735 | fixed | NA | groupFBVLED:time_pointFinal | 0.46 | 0.17 | 2.77 | 39.68 | 0.13 | 0.80 | **0.008** | 0.226 |
| Ruminococcus_D bicirculans | fixed | NA | groupFBVLED:time_pointFinal | 1.43 | 0.52 | 2.74 | 41.36 | 0.38 | 2.48 | **0.009** | 0.233 |
| Bacteroides finegoldii | fixed | NA | groupFBVLED:time_pointFinal | 0.58 | 0.22 | 2.64 | 39.85 | 0.14 | 1.03 | **0.012** | 0.276 |
| Bifidobacterium longum | fixed | NA | groupFBVLED:time_pointFinal | -1.21 | 0.46 | -2.63 | 40.76 | -2.13 | -0.28 | **0.012** | 0.276 |
| Lachnospira eligens_B | fixed | NA | groupFBVLED:time_pointFinal | 1.32 | 0.51 | 2.60 | 43.15 | 0.30 | 2.35 | **0.013** | 0.276 |

| OUTCOMES - SPECIES | EFFECT | GROUP | TERM | ESTIMATE | STD ERROR | STATISTIC | DF | CONF LOW | CONF HIGH | P VALUE | Q VALUE |
|---|---|---|---|---|---|---|---|---|---|---|---|
| CAG 74 MIC8062 | fixed | NA | groupFBVLED:time_pointFinal | -1.12 | 0.43 | -2.58 | 44.79 | -2.00 | -0.25 | **0.013** | 0.276 |
| Bacteroides faecis | fixed | NA | groupFBVLED:time_pointFinal | -0.52 | 0.20 | -2.58 | 40.42 | -0.93 | -0.11 | **0.014** | 0.276 |
| Lachnospira rogosae | fixed | NA | groupFBVLED:time_pointFinal | 1.08 | 0.42 | 2.57 | 38.89 | 0.23 | 1.93 | **0.014** | 0.276 |
| Faecalibacterium MIC8666 | fixed | NA | groupFBVLED:time_pointFinal | 1.00 | 0.39 | 2.55 | 44.05 | 0.21 | 1.79 | **0.014** | 0.276 |
| Hungatella effluvii | fixed | NA | groupFBVLED:time_pointFinal | -0.42 | 0.17 | -2.49 | 84.00 | -0.75 | -0.09 | **0.015** | 0.276 |
| Eisenbergiella sp900066775 | fixed | NA | groupFBVLED:time_pointFinal | 0.67 | 0.26 | 2.54 | 42.57 | 0.14 | 1.20 | **0.015** | 0.276 |
| Agathobaculum butyriciproducens | fixed | NA | groupFBVLED:time_pointFinal | 0.79 | 0.31 | 2.52 | 43.94 | 0.16 | 1.42 | **0.016** | 0.284 |
| Ruthenibacterium lactatiformans | fixed | NA | groupFBVLED:time_pointFinal | -1.14 | 0.46 | -2.49 | 44.38 | -2.07 | -0.22 | **0.017** | 0.288 |
| Faecalibacterium MIC7145 | fixed | NA | groupFBVLED:time_pointFinal | 0.90 | 0.36 | 2.48 | 40.22 | 0.17 | 1.63 | **0.017** | 0.288 |
| Gemmiger MIC9530 | fixed | NA | groupFBVLED:time_pointFinal | 0.66 | 0.27 | 2.47 | 42.17 | 0.12 | 1.21 | **0.018** | 0.288 |
| UBA1820 sp002314265 | fixed | NA | groupFBVLED:time_pointFinal | 0.46 | 0.19 | 2.46 | 43.92 | 0.08 | 0.84 | **0.018** | 0.288 |
| Lachnospiraceae MIC6495 | fixed | NA | groupFBVLED:time_pointFinal | 0.43 | 0.17 | 2.46 | 43.09 | 0.08 | 0.78 | **0.018** | 0.288 |
| Eubacterium_R sp000434995 | fixed | NA | groupFBVLED:time_pointFinal | 0.51 | 0.21 | 2.44 | 41.00 | 0.09 | 0.94 | **0.019** | 0.296 |
| Intestinimonas butyriciproducens | fixed | NA | groupFBVLED:time_pointFinal | -0.54 | 0.23 | -2.40 | 45.75 | -1.00 | -0.09 | **0.021** | 0.312 |
| COE1 sp001916965 | fixed | NA | groupFBVLED:time_pointFinal | 0.51 | 0.21 | 2.40 | 39.96 | 0.08 | 0.93 | **0.021** | 0.314 |
| Eggerthella lenta | fixed | NA | groupFBVLED:time_pointFinal | -0.71 | 0.30 | -2.38 | 41.84 | -1.32 | -0.11 | **0.022** | 0.314 |
| UBA9502 MIC6887 | fixed | NA | groupFBVLED:time_pointFinal | 0.31 | 0.13 | 2.36 | 43.12 | 0.04 | 0.58 | **0.023** | 0.324 |
| GCA 900066995 sp900291955 | fixed | NA | groupFBVLED:time_pointFinal | 0.62 | 0.26 | 2.34 | 45.81 | 0.09 | 1.15 | **0.024** | 0.328 |
| Blautia_A MIC7250 | fixed | NA | groupFBVLED:time_pointFinal | 0.21 | 0.09 | 2.29 | 84.00 | 0.03 | 0.39 | **0.025** | 0.333 |
| Faecalibacterium prausnitzii_I | fixed | NA | groupFBVLED:time_pointFinal | 0.70 | 0.30 | 2.30 | 41.20 | 0.09 | 1.31 | **0.026** | 0.343 |
| UBA1417 sp003531055 | fixed | NA | groupFBVLED:time_pointFinal | -0.95 | 0.42 | -2.29 | 41.51 | -1.79 | -0.11 | **0.027** | 0.343 |
| Coprococcus sp000154245 | fixed | NA | groupFBVLED:time_pointFinal | 0.74 | 0.32 | 2.28 | 45.47 | 0.09 | 1.39 | **0.027** | 0.343 |
| Clostridium sp001916075 | fixed | NA | groupFBVLED:time_pointFinal | 0.86 | 0.38 | 2.27 | 44.77 | 0.10 | 1.61 | **0.028** | 0.343 |
| Escherichia flexneri | fixed | NA | groupFBVLED:time_pointFinal | -0.90 | 0.41 | -2.22 | 84.00 | -1.71 | -0.09 | **0.029** | 0.343 |
| UBA1191 MIC6579 | fixed | NA | groupFBVLED:time_pointFinal | -0.38 | 0.17 | -2.25 | 43.82 | -0.71 | -0.04 | **0.029** | 0.343 |
| Lachnospira sp900316325 | fixed | NA | groupFBVLED:time_pointFinal | 0.82 | 0.37 | 2.25 | 42.28 | 0.08 | 1.56 | **0.03** | 0.343 |
| Parabacteroides goldsteinii | fixed | NA | groupFBVLED:time_pointFinal | -0.74 | 0.33 | -2.24 | 43.62 | -1.40 | -0.08 | **0.03** | 0.343 |
| Lactobacillus_C rhamnosus | fixed | NA | groupFBVLED:time_pointFinal | 0.39 | 0.18 | 2.18 | 84.00 | 0.03 | 0.74 | **0.032** | 0.357 |
| Blautia_A MIC8343 | fixed | NA | groupFBVLED:time_pointFinal | 0.33 | 0.15 | 2.22 | 39.24 | 0.03 | 0.64 | **0.032** | 0.357 |

| OUTCOMES - SPECIES | EFFECT | GROUP | TERM | ESTIMATE | STD ERROR | STATISTIC | DF | CONF LOW | CONF HIGH | P VALUE | Q VALUE |
|---|---|---|---|---|---|---|---|---|---|---|---|
| Terrisporobacter MIC9205 | fixed | NA | groupFBVLED:time_pointFinal | 0.67 | 0.31 | 2.19 | 41.91 | 0.05 | 1.28 | **0.034** | 0.364 |
| Coprococcus_A catus | fixed | NA | groupFBVLED:time_pointFinal | 0.58 | 0.26 | 2.19 | 40.53 | 0.04 | 1.11 | **0.034** | 0.364 |
| Oxalobacter MIC6654 | fixed | NA | groupFBVLED:time_pointFinal | 0.30 | 0.14 | 2.18 | 43.82 | 0.02 | 0.58 | **0.035** | 0.364 |
| Absiella sp000163515 | fixed | NA | groupFBVLED:time_pointFinal | -0.31 | 0.14 | -2.16 | 42.28 | -0.60 | -0.02 | **0.036** | 0.364 |
| CAG 81 sp900066535 | fixed | NA | groupFBVLED:time_pointFinal | 0.42 | 0.20 | 2.16 | 42.41 | 0.03 | 0.81 | **0.037** | 0.364 |
| Blautia_A sp900066145 | fixed | NA | groupFBVLED:time_pointFinal | 0.70 | 0.33 | 2.16 | 41.32 | 0.04 | 1.36 | **0.037** | 0.364 |
| 4C28d 15 MIC7065 | fixed | NA | groupFBVLED:time_pointFinal | 0.18 | 0.08 | 2.15 | 43.84 | 0.01 | 0.35 | **0.037** | 0.364 |
| Lachnospiraceae MIC7543 | fixed | NA | groupFBVLED:time_pointFinal | -0.21 | 0.10 | -2.14 | 41.94 | -0.41 | -0.01 | **0.038** | 0.364 |
| Massilimaliae timonensis | fixed | NA | groupFBVLED:time_pointFinal | -0.64 | 0.30 | -2.11 | 83.99 | -1.24 | -0.04 | **0.038** | 0.364 |
| CAG 269 sp003525075 | fixed | NA | groupFBVLED:time_pointFinal | -0.45 | 0.21 | -2.13 | 40.94 | -0.87 | -0.02 | **0.039** | 0.364 |
| Eisenbergiella tayi | fixed | NA | groupFBVLED:time_pointFinal | -0.79 | 0.37 | -2.11 | 46.18 | -1.55 | -0.04 | **0.04** | 0.364 |
| CAG 103 sp000432375 | fixed | NA | groupFBVLED:time_pointFinal | 0.68 | 0.32 | 2.12 | 41.54 | 0.03 | 1.33 | **0.04** | 0.364 |
| Erysipelatoclostridium ramosum | fixed | NA | groupFBVLED:time_pointFinal | -0.74 | 0.35 | -2.11 | 42.82 | -1.44 | -0.03 | **0.041** | 0.365 |
| QAND01 sp003150225 | fixed | NA | groupFBVLED:time_pointFinal | -0.22 | 0.11 | -2.07 | 84.00 | -0.43 | -0.01 | **0.042** | 0.365 |
| Faecalitalea cylindroides | fixed | NA | groupFBVLED:time_pointFinal | -0.52 | 0.25 | -2.10 | 38.50 | -1.03 | -0.02 | **0.042** | 0.365 |
| CAG 81 sp900066055 | fixed | NA | groupFBVLED:time_pointFinal | 0.26 | 0.12 | 2.09 | 44.42 | 0.01 | 0.51 | **0.043** | 0.365 |
| CAG 313 sp003539625 | fixed | NA | groupFBVLED:time_pointFinal | 0.76 | 0.37 | 2.06 | 46.30 | 0.02 | 1.50 | 0.045 | 0.374 |
| CAG 74 MIC7649 | fixed | NA | groupFBVLED:time_pointFinal | -0.52 | 0.25 | -2.07 | 41.71 | -1.03 | -0.01 | 0.045 | 0.374 |
| Lachnospiraceae MIC8643 | fixed | NA | groupFBVLED:time_pointFinal | -0.24 | 0.12 | -2.06 | 45.40 | -0.48 | 0.00 | 0.046 | 0.376 |
| Cloacibacillus evryensis | fixed | NA | groupFBVLED:time_pointFinal | -0.27 | 0.13 | -2.03 | 43.49 | -0.54 | 0.00 | 0.048 | 0.393 |
| Oscillibacter MIC7603 | fixed | NA | groupFBVLED:time_pointFinal | -0.18 | 0.09 | -1.98 | 84.00 | -0.36 | 0.00 | 0.051 | 0.41 |
| Coprococcus_B comes | fixed | NA | groupFBVLED:time_pointFinal | 0.51 | 0.25 | 2.00 | 41.27 | 0.00 | 1.01 | 0.052 | 0.41 |
| Oscillospiraceae MIC8045 | fixed | NA | groupFBVLED:time_pointFinal | 0.21 | 0.10 | 2.00 | 32.96 | 0.00 | 0.42 | 0.054 | 0.418 |
| UBA9475 MIC7490 | fixed | NA | groupFBVLED:time_pointFinal | 0.18 | 0.09 | 1.98 | 44.45 | 0.00 | 0.36 | 0.054 | 0.418 |
| Dialister invisus | fixed | NA | groupFBVLED:time_pointFinal | 0.39 | 0.20 | 1.98 | 39.30 | -0.01 | 0.78 | 0.055 | 0.422 |
| An200 sp003268275 | fixed | NA | groupFBVLED:time_pointFinal | -0.20 | 0.10 | -1.96 | 44.58 | -0.41 | 0.01 | 0.056 | 0.422 |
| Odoribacter splanchnicus | fixed | NA | groupFBVLED:time_pointFinal | 0.52 | 0.27 | 1.96 | 40.70 | -0.02 | 1.06 | 0.057 | 0.422 |
| Bifidobacterium animalis | fixed | NA | groupFBVLED:time_pointFinal | 0.45 | 0.23 | 1.95 | 44.78 | -0.02 | 0.92 | 0.057 | 0.422 |
| UBA7102 MIC9705 | fixed | NA | groupFBVLED:time_pointFinal | -0.38 | 0.20 | -1.94 | 45.19 | -0.78 | 0.01 | 0.058 | 0.422 |

| OUTCOMES - SPECIES | EFFECT | GROUP | TERM | ESTIMATE | STD ERROR | STATISTIC | DF | CONF LOW | CONF HIGH | P VALUE | Q VALUE |
|---|---|---|---|---|---|---|---|---|---|---|---|
| CAG 95 sp900066375 | fixed | NA | groupFBVLED:time_pointFinal | 0.57 | 0.30 | 1.90 | 84.00 | -0.03 | 1.17 | 0.061 | 0.422 |
| Eubacterium_F sp003491505 | fixed | NA | groupFBVLED:time_pointFinal | 0.49 | 0.26 | 1.92 | 41.99 | -0.02 | 1.01 | 0.061 | 0.422 |
| Turicibacter sp001543345 | fixed | NA | groupFBVLED:time_pointFinal | 0.35 | 0.18 | 1.92 | 42.71 | -0.02 | 0.72 | 0.061 | 0.422 |
| Lachnospiraceae MIC9183 | fixed | NA | groupFBVLED:time_pointFinal | -0.30 | 0.15 | -1.92 | 45.46 | -0.61 | 0.02 | 0.062 | 0.422 |
| Clostridium MIC8163 | fixed | NA | groupFBVLED:time_pointFinal | 0.81 | 0.42 | 1.93 | 36.50 | -0.04 | 1.67 | 0.062 | 0.422 |
| Butyricimonas sp002161485 | fixed | NA | groupFBVLED:time_pointFinal | 0.57 | 0.30 | 1.91 | 42.48 | -0.03 | 1.17 | 0.062 | 0.422 |
| Alistipes finegoldii | fixed | NA | groupFBVLED:time_pointFinal | 0.49 | 0.26 | 1.91 | 40.61 | -0.03 | 1.01 | 0.063 | 0.422 |
| Victivallis sp002998355 | fixed | NA | groupFBVLED:time_pointFinal | 0.35 | 0.18 | 1.90 | 43.93 | -0.02 | 0.71 | 0.064 | 0.423 |
| Desulfovibrio fairfieldensis | fixed | NA | groupFBVLED:time_pointFinal | -0.32 | 0.17 | -1.90 | 44.07 | -0.66 | 0.02 | 0.064 | 0.423 |
| Gemmiger MIC8010 | fixed | NA | groupFBVLED:time_pointFinal | -0.50 | 0.27 | -1.88 | 41.43 | -1.04 | 0.04 | 0.068 | 0.433 |
| Flavonifractor plautii | fixed | NA | groupFBVLED:time_pointFinal | -0.53 | 0.28 | -1.87 | 44.58 | -1.09 | 0.04 | 0.068 | 0.433 |
| Negativibacillus sp000435195 | fixed | NA | groupFBVLED:time_pointFinal | 0.41 | 0.22 | 1.87 | 39.58 | -0.03 | 0.86 | 0.068 | 0.433 |
| CAG 81 sp000435795 | fixed | NA | groupFBVLED:time_pointFinal | 0.40 | 0.21 | 1.87 | 41.24 | -0.03 | 0.83 | 0.069 | 0.433 |
| CAG 74 MIC9837 | fixed | NA | groupFBVLED:time_pointFinal | -0.51 | 0.27 | -1.86 | 42.94 | -1.06 | 0.04 | 0.069 | 0.433 |
| CAG 475 sp000434435 | fixed | NA | groupFBVLED:time_pointFinal | 0.47 | 0.25 | 1.86 | 41.93 | -0.04 | 0.99 | 0.07 | 0.433 |
| Alistipes obesi | fixed | NA | groupFBVLED:time_pointFinal | 0.63 | 0.34 | 1.85 | 42.35 | -0.06 | 1.32 | 0.071 | 0.433 |
| Faecalibacterium MIC8693 | fixed | NA | groupFBVLED:time_pointFinal | 0.20 | 0.11 | 1.82 | 84.00 | -0.02 | 0.42 | 0.073 | 0.433 |
| CAG 1427 sp000435475 | fixed | NA | groupFBVLED:time_pointFinal | -0.32 | 0.17 | -1.84 | 40.70 | -0.67 | 0.03 | 0.074 | 0.433 |
| Ruminococcus MIC6917 | fixed | NA | groupFBVLED:time_pointFinal | 0.39 | 0.21 | 1.83 | 43.88 | -0.04 | 0.82 | 0.074 | 0.433 |
| Streptococcus parasanguinis | fixed | NA | groupFBVLED:time_pointFinal | 0.44 | 0.24 | 1.83 | 40.77 | -0.05 | 0.92 | 0.075 | 0.433 |
| Ruthenibacterium MIC9855 | fixed | NA | groupFBVLED:time_pointFinal | -0.27 | 0.15 | -1.82 | 44.48 | -0.57 | 0.03 | 0.075 | 0.433 |
| Alistipes putredinis | fixed | NA | groupFBVLED:time_pointFinal | 0.42 | 0.23 | 1.83 | 39.99 | -0.04 | 0.89 | 0.075 | 0.433 |
| TF01 11 sp003529475 | fixed | NA | groupFBVLED:time_pointFinal | 0.61 | 0.34 | 1.82 | 43.07 | -0.07 | 1.29 | 0.076 | 0.436 |
| Acutalibacter timonensis | fixed | NA | groupFBVLED:time_pointFinal | -0.62 | 0.34 | -1.81 | 45.37 | -1.31 | 0.07 | 0.077 | 0.436 |
| Faecalicatena glycyrrhizinilyticum | fixed | NA | groupFBVLED:time_pointFinal | 0.25 | 0.14 | 1.80 | 42.84 | -0.03 | 0.53 | 0.08 | 0.445 |
| QANA01 MIC6812 | fixed | NA | groupFBVLED:time_pointFinal | 0.17 | 0.10 | 1.79 | 44.80 | -0.02 | 0.37 | 0.08 | 0.445 |
| Clostridium_Q sp003024715 | fixed | NA | groupFBVLED:time_pointFinal | 0.42 | 0.24 | 1.79 | 41.29 | -0.05 | 0.90 | 0.08 | 0.445 |
| Roseburia inulinivorans | fixed | NA | groupFBVLED:time_pointFinal | 0.59 | 0.33 | 1.77 | 38.40 | -0.08 | 1.25 | 0.084 | 0.445 |
| Lachnospirales MIC7715 | fixed | NA | groupFBVLED:time_pointFinal | -0.22 | 0.12 | -1.77 | 44.51 | -0.47 | 0.03 | 0.084 | 0.445 |

| OUTCOMES - SPECIES | EFFECT | GROUP | TERM | ESTIMATE | STD ERROR | STATISTIC | DF | CONF LOW | CONF HIGH | P VALUE | Q VALUE |
|---|---|---|---|---|---|---|---|---|---|---|---|
| Bifidobacterium adolescentis | fixed | NA | groupFBVLED:time_pointFinal | -0.75 | 0.43 | -1.77 | 39.10 | -1.61 | 0.11 | 0.084 | 0.445 |
| F23 B02 sp001916715 | fixed | NA | groupFBVLED:time_pointFinal | 0.58 | 0.33 | 1.76 | 43.43 | -0.08 | 1.25 | 0.085 | 0.445 |
| UBA1820 sp003150615 | fixed | NA | groupFBVLED:time_pointFinal | -0.25 | 0.14 | -1.76 | 43.80 | -0.55 | 0.04 | 0.086 | 0.445 |
| UBA1191 MIC7533 | fixed | NA | groupFBVLED:time_pointFinal | 0.10 | 0.05 | 1.76 | 39.60 | -0.01 | 0.20 | 0.086 | 0.445 |
| CAG 452 sp000434035 | fixed | NA | groupFBVLED:time_pointFinal | -0.20 | 0.11 | -1.76 | 40.11 | -0.44 | 0.03 | 0.086 | 0.445 |
| Ruminococcus_E bromii_B | fixed | NA | groupFBVLED:time_pointFinal | 1.08 | 0.61 | 1.75 | 40.45 | -0.16 | 2.32 | 0.087 | 0.448 |
| CAG 45 sp900066395 | fixed | NA | groupFBVLED:time_pointFinal | 0.59 | 0.34 | 1.74 | 43.91 | -0.09 | 1.27 | 0.089 | 0.448 |
| Lachnospira sp000437735 | fixed | NA | groupFBVLED:time_pointFinal | 0.65 | 0.37 | 1.74 | 41.84 | -0.10 | 1.40 | 0.089 | 0.448 |
| Dorea sp900066555 | fixed | NA | groupFBVLED:time_pointFinal | 0.38 | 0.22 | 1.73 | 41.87 | -0.06 | 0.82 | 0.091 | 0.448 |
| Ruthenibacterium MIC9423 | fixed | NA | groupFBVLED:time_pointFinal | -0.24 | 0.14 | -1.73 | 43.95 | -0.51 | 0.04 | 0.091 | 0.448 |
| CAG 103 MIC7540 | fixed | NA | groupFBVLED:time_pointFinal | 0.30 | 0.17 | 1.73 | 40.75 | -0.05 | 0.65 | 0.091 | 0.448 |
| Oscillibacter MIC9243 | fixed | NA | groupFBVLED:time_pointFinal | -0.12 | 0.07 | -1.72 | 39.57 | -0.26 | 0.02 | 0.093 | 0.448 |
| Parasutterella excrementihominis | fixed | NA | groupFBVLED:time_pointFinal | 0.38 | 0.22 | 1.72 | 39.17 | -0.07 | 0.83 | 0.093 | 0.448 |
| Marvinbryantia sp900066075 | fixed | NA | groupFBVLED:time_pointFinal | 0.19 | 0.11 | 1.72 | 42.35 | -0.03 | 0.40 | 0.093 | 0.448 |
| Eubacterium_G ventriosum | fixed | NA | groupFBVLED:time_pointFinal | 0.39 | 0.23 | 1.71 | 41.48 | -0.07 | 0.84 | 0.094 | 0.448 |
| Blautia sp000436935 | fixed | NA | groupFBVLED:time_pointFinal | 0.53 | 0.31 | 1.71 | 38.06 | -0.10 | 1.15 | 0.095 | 0.448 |
| Ruthenibacterium sp003149955 | fixed | NA | groupFBVLED:time_pointFinal | -0.28 | 0.16 | -1.69 | 84.00 | -0.60 | 0.05 | 0.095 | 0.448 |
| Lachnospiraceae MIC7886 | fixed | NA | groupFBVLED:time_pointFinal | 0.17 | 0.10 | 1.70 | 42.29 | -0.03 | 0.37 | 0.096 | 0.449 |
| CAG 492 sp000434335 | fixed | NA | groupFBVLED:time_pointFinal | 0.37 | 0.22 | 1.69 | 44.81 | -0.07 | 0.81 | 0.098 | 0.452 |
| Ruminococcus_E sp003526955 | fixed | NA | groupFBVLED:time_pointFinal | 0.69 | 0.41 | 1.69 | 41.88 | -0.13 | 1.50 | 0.098 | 0.452 |
| Alistipes_A sp900240235 | fixed | NA | groupFBVLED:time_pointFinal | 0.33 | 0.20 | 1.68 | 40.83 | -0.07 | 0.73 | 0.1 | 0.453 |
| CAG 217 sp000436335 | fixed | NA | groupFBVLED:time_pointFinal | 0.31 | 0.19 | 1.68 | 39.65 | -0.06 | 0.69 | 0.1 | 0.453 |
| ER4 MIC9395 | fixed | NA | groupFBVLED:time_pointFinal | 0.21 | 0.13 | 1.68 | 43.13 | -0.04 | 0.47 | 0.1 | 0.453 |
| Clostridium_M asparagiforme | fixed | NA | groupFBVLED:time_pointFinal | -0.31 | 0.19 | -1.65 | 84.00 | -0.68 | 0.06 | 0.103 | 0.453 |
| Blautia_A MIC9663 | fixed | NA | groupFBVLED:time_pointFinal | -0.17 | 0.10 | -1.67 | 40.06 | -0.38 | 0.04 | 0.104 | 0.453 |
| CAG 272 MIC6999 | fixed | NA | groupFBVLED:time_pointFinal | -0.43 | 0.26 | -1.66 | 44.27 | -0.94 | 0.09 | 0.104 | 0.453 |
| Lawsonibacter sp000177015 | fixed | NA | groupFBVLED:time_pointFinal | 0.10 | 0.06 | 1.66 | 40.11 | -0.02 | 0.22 | 0.104 | 0.453 |
| CAG 314 sp000437915 | fixed | NA | groupFBVLED:time_pointFinal | 0.52 | 0.32 | 1.65 | 43.97 | -0.11 | 1.16 | 0.106 | 0.453 |
| Peptoniphilus_C sp900088125 | fixed | NA | groupFBVLED:time_pointFinal | 0.28 | 0.17 | 1.63 | 84.00 | -0.06 | 0.61 | 0.106 | 0.453 |

| OUTCOMES - SPECIES | EFFECT | GROUP | TERM | ESTIMATE | STD ERROR | STATISTIC | DF | CONF LOW | CONF HIGH | P VALUE | Q VALUE |
|---|---|---|---|---|---|---|---|---|---|---|---|
| Oscillibacter sp900066435 | fixed | NA | groupFBVLED:time_pointFinal | -0.62 | 0.38 | -1.64 | 44.73 | -1.39 | 0.14 | 0.108 | 0.453 |
| CAG 41 sp900066215 | fixed | NA | groupFBVLED:time_pointFinal | 0.78 | 0.48 | 1.64 | 43.06 | -0.18 | 1.75 | 0.108 | 0.453 |
| Clostridium_M MIC6986 | fixed | NA | groupFBVLED:time_pointFinal | 0.25 | 0.15 | 1.64 | 39.27 | -0.06 | 0.55 | 0.109 | 0.453 |
| CAG 1427 sp000435675 | fixed | NA | groupFBVLED:time_pointFinal | 0.44 | 0.27 | 1.63 | 43.27 | -0.10 | 0.99 | 0.11 | 0.453 |
| CAG 313 MIC9072 | fixed | NA | groupFBVLED:time_pointFinal | 0.33 | 0.20 | 1.63 | 45.32 | -0.08 | 0.74 | 0.11 | 0.453 |
| CAG 145 MIC8493 | fixed | NA | groupFBVLED:time_pointFinal | 0.13 | 0.08 | 1.63 | 41.04 | -0.03 | 0.28 | 0.111 | 0.453 |
| Bilophila MIC7011 | fixed | NA | groupFBVLED:time_pointFinal | 0.08 | 0.05 | 1.63 | 37.34 | -0.02 | 0.18 | 0.111 | 0.453 |
| Barnesiella intestinihominis | fixed | NA | groupFBVLED:time_pointFinal | 0.54 | 0.34 | 1.62 | 40.51 | -0.13 | 1.22 | 0.112 | 0.453 |
| Peptoniphilus_A harei_A | fixed | NA | groupFBVLED:time_pointFinal | 0.26 | 0.16 | 1.63 | 31.63 | -0.07 | 0.59 | 0.112 | 0.453 |
| Bacteroides fragilis | fixed | NA | groupFBVLED:time_pointFinal | 0.40 | 0.25 | 1.62 | 41.29 | -0.10 | 0.90 | 0.113 | 0.453 |
| Dorea longicatena | fixed | NA | groupFBVLED:time_pointFinal | 0.50 | 0.31 | 1.62 | 39.57 | -0.12 | 1.12 | 0.114 | 0.453 |
| Pseudoflavonifractor MIC6616 | fixed | NA | groupFBVLED:time_pointFinal | -0.35 | 0.22 | -1.60 | 84.00 | -0.80 | 0.09 | 0.114 | 0.453 |
| QANA01 sp003149735 | fixed | NA | groupFBVLED:time_pointFinal | -0.14 | 0.09 | -1.61 | 43.84 | -0.32 | 0.04 | 0.115 | 0.453 |
| CAG 170 MIC9129 | fixed | NA | groupFBVLED:time_pointFinal | 0.20 | 0.13 | 1.61 | 41.35 | -0.05 | 0.46 | 0.115 | 0.453 |
| CAG 74 MIC7044 | fixed | NA | groupFBVLED:time_pointFinal | 0.42 | 0.26 | 1.59 | 42.13 | -0.11 | 0.96 | 0.119 | 0.46 |
| CAG 269 sp000437215 | fixed | NA | groupFBVLED:time_pointFinal | 0.24 | 0.15 | 1.59 | 42.44 | -0.06 | 0.54 | 0.119 | 0.46 |
| Bifidobacterium MIC6680 | fixed | NA | groupFBVLED:time_pointFinal | -0.27 | 0.17 | -1.59 | 40.54 | -0.61 | 0.07 | 0.12 | 0.46 |
| Akkermansia muciniphila_B | fixed | NA | groupFBVLED:time_pointFinal | 0.65 | 0.41 | 1.58 | 44.42 | -0.18 | 1.48 | 0.121 | 0.46 |
| Ruminiclostridium_C MIC9168 | fixed | NA | groupFBVLED:time_pointFinal | -0.06 | 0.04 | -1.58 | 39.44 | -0.14 | 0.02 | 0.121 | 0.46 |
| Akkermansia muciniphila | fixed | NA | groupFBVLED:time_pointFinal | -0.76 | 0.48 | -1.58 | 41.34 | -1.74 | 0.21 | 0.121 | 0.46 |
| CAG 313 sp000433035 | fixed | NA | groupFBVLED:time_pointFinal | -0.32 | 0.21 | -1.58 | 41.12 | -0.74 | 0.09 | 0.122 | 0.461 |
| CAG 727 MIC8506 | fixed | NA | groupFBVLED:time_pointFinal | 0.32 | 0.20 | 1.57 | 42.14 | -0.09 | 0.73 | 0.123 | 0.461 |
| QAND01 MIC9470 | fixed | NA | groupFBVLED:time_pointFinal | -0.56 | 0.36 | -1.56 | 46.27 | -1.28 | 0.16 | 0.125 | 0.465 |
| Alistipes shahii | fixed | NA | groupFBVLED:time_pointFinal | -0.54 | 0.35 | -1.56 | 42.00 | -1.24 | 0.16 | 0.127 | 0.47 |
| Anaerostipes caccae | fixed | NA | groupFBVLED:time_pointFinal | -0.31 | 0.20 | -1.55 | 44.52 | -0.71 | 0.09 | 0.128 | 0.47 |
| CAG 110 sp000435995 | fixed | NA | groupFBVLED:time_pointFinal | -0.48 | 0.31 | -1.55 | 41.57 | -1.11 | 0.15 | 0.129 | 0.47 |
| Blautia_A sp900066165 | fixed | NA | groupFBVLED:time_pointFinal | 0.44 | 0.29 | 1.55 | 42.24 | -0.13 | 1.02 | 0.129 | 0.47 |
| Acutalibacter MIC8008 | fixed | NA | groupFBVLED:time_pointFinal | -0.44 | 0.29 | -1.54 | 45.84 | -1.03 | 0.14 | 0.131 | 0.473 |
| CAG 74 MIC6989 | fixed | NA | groupFBVLED:time_pointFinal | -0.45 | 0.30 | -1.53 | 45.37 | -1.05 | 0.14 | 0.134 | 0.473 |

| OUTCOMES - SPECIES | EFFECT | GROUP | TERM | ESTIMATE | STD ERROR | STATISTIC | DF | CONF LOW | CONF HIGH | P VALUE | Q VALUE |
|---|---|---|---|---|---|---|---|---|---|---|---|
| Slackia_A piriformis | fixed | NA | groupFBVLED:time_pointFinal | 0.07 | 0.05 | 1.53 | 38.97 | -0.02 | 0.16 | 0.134 | 0.473 |
| QAND01 MIC9113 | fixed | NA | groupFBVLED:time_pointFinal | -0.39 | 0.26 | -1.53 | 44.19 | -0.91 | 0.13 | 0.134 | 0.473 |
| UBA6398 MIC8483 | fixed | NA | groupFBVLED:time_pointFinal | 0.06 | 0.04 | 1.52 | 39.70 | -0.02 | 0.13 | 0.137 | 0.473 |
| Bacteroides salyersiae | fixed | NA | groupFBVLED:time_pointFinal | 0.28 | 0.19 | 1.51 | 39.33 | -0.10 | 0.66 | 0.138 | 0.473 |
| CAG 83 MIC7389 | fixed | NA | groupFBVLED:time_pointFinal | -0.31 | 0.20 | -1.51 | 43.80 | -0.72 | 0.10 | 0.138 | 0.473 |
| Blautia_A sp900066505 | fixed | NA | groupFBVLED:time_pointFinal | 0.35 | 0.23 | 1.51 | 43.74 | -0.12 | 0.82 | 0.14 | 0.473 |
| Adlercreutzia MIC8014 | fixed | NA | groupFBVLED:time_pointFinal | 0.43 | 0.29 | 1.50 | 41.16 | -0.15 | 1.01 | 0.141 | 0.473 |
| UBA1777 MIC6732 | fixed | NA | groupFBVLED:time_pointFinal | 0.25 | 0.17 | 1.50 | 41.85 | -0.09 | 0.59 | 0.141 | 0.473 |
| CAG 272 MIC8971 | fixed | NA | groupFBVLED:time_pointFinal | 0.17 | 0.11 | 1.50 | 44.96 | -0.06 | 0.40 | 0.142 | 0.473 |
| Acutalibacteraceae MIC7526 | fixed | NA | groupFBVLED:time_pointFinal | -0.25 | 0.17 | -1.50 | 42.61 | -0.60 | 0.09 | 0.142 | 0.473 |
| CAG 83 MIC7830 | fixed | NA | groupFBVLED:time_pointFinal | 0.17 | 0.12 | 1.50 | 41.23 | -0.06 | 0.41 | 0.142 | 0.473 |
| Flavonifractor sp000508885 | fixed | NA | groupFBVLED:time_pointFinal | -0.45 | 0.30 | -1.49 | 45.87 | -1.06 | 0.16 | 0.143 | 0.473 |
| CAG 460 sp000437355 | fixed | NA | groupFBVLED:time_pointFinal | 0.12 | 0.08 | 1.49 | 40.79 | -0.04 | 0.28 | 0.143 | 0.473 |
| Oscillospiraceae MIC9607 | fixed | NA | groupFBVLED:time_pointFinal | -0.17 | 0.12 | -1.49 | 46.21 | -0.41 | 0.06 | 0.144 | 0.473 |
| TF01 11 sp001916135 | fixed | NA | groupFBVLED:time_pointFinal | 0.19 | 0.13 | 1.50 | 34.00 | -0.07 | 0.45 | 0.144 | 0.473 |
| Oscillibacter MIC6445 | fixed | NA | groupFBVLED:time_pointFinal | 0.17 | 0.12 | 1.49 | 39.80 | -0.06 | 0.41 | 0.144 | 0.473 |
| UBA5394 sp003150565 | fixed | NA | groupFBVLED:time_pointFinal | -0.36 | 0.25 | -1.47 | 84.00 | -0.86 | 0.13 | 0.144 | 0.473 |
| UBA5416 MIC7893 | fixed | NA | groupFBVLED:time_pointFinal | -0.29 | 0.20 | -1.48 | 45.39 | -0.69 | 0.11 | 0.145 | 0.473 |
| NK3B98 MIC8354 | fixed | NA | groupFBVLED:time_pointFinal | 0.21 | 0.14 | 1.48 | 40.55 | -0.08 | 0.50 | 0.147 | 0.476 |
| CAG 495 sp001917125 | fixed | NA | groupFBVLED:time_pointFinal | 0.18 | 0.12 | 1.48 | 33.74 | -0.07 | 0.44 | 0.148 | 0.476 |
| Bacteroides_B MIC8119 | fixed | NA | groupFBVLED:time_pointFinal | -0.20 | 0.13 | -1.47 | 39.83 | -0.47 | 0.07 | 0.149 | 0.476 |
| Gordonibacter pamelaeae | fixed | NA | groupFBVLED:time_pointFinal | -0.28 | 0.19 | -1.47 | 43.37 | -0.67 | 0.11 | 0.149 | 0.476 |
| Blautia_A MIC7329 | fixed | NA | groupFBVLED:time_pointFinal | 0.14 | 0.09 | 1.47 | 42.04 | -0.05 | 0.33 | 0.15 | 0.476 |
| Parabacteroides johnsonii | fixed | NA | groupFBVLED:time_pointFinal | -0.21 | 0.14 | -1.46 | 42.61 | -0.50 | 0.08 | 0.151 | 0.476 |
| Massilioclostridium coli | fixed | NA | groupFBVLED:time_pointFinal | -0.14 | 0.10 | -1.46 | 45.19 | -0.34 | 0.05 | 0.151 | 0.476 |
| Bifidobacterium bifidum | fixed | NA | groupFBVLED:time_pointFinal | -0.24 | 0.17 | -1.45 | 40.46 | -0.58 | 0.09 | 0.154 | 0.479 |
| Bacteroides nordii | fixed | NA | groupFBVLED:time_pointFinal | -0.38 | 0.26 | -1.45 | 44.83 | -0.91 | 0.15 | 0.155 | 0.479 |
| Erysipelatoclostridium sp000752095 | fixed | NA | groupFBVLED:time_pointFinal | -0.44 | 0.30 | -1.45 | 39.64 | -1.04 | 0.17 | 0.155 | 0.479 |
| CAG 312 MIC7338 | fixed | NA | groupFBVLED:time_pointFinal | 0.25 | 0.17 | 1.44 | 45.00 | -0.10 | 0.59 | 0.156 | 0.479 |

| OUTCOMES - SPECIES | EFFECT | GROUP | TERM | ESTIMATE | STD ERROR | STATISTIC | DF | CONF LOW | CONF HIGH | P VALUE | Q VALUE |
|---|---|---|---|---|---|---|---|---|---|---|---|
| Intestinibacter bartlettii | fixed | NA | groupFBVLED:time_pointFinal | 0.49 | 0.34 | 1.43 | 40.60 | -0.20 | 1.17 | 0.159 | 0.483 |
| CAG 354 sp001915925 | fixed | NA | groupFBVLED:time_pointFinal | 0.11 | 0.08 | 1.43 | 38.58 | -0.05 | 0.27 | 0.16 | 0.483 |
| Romboutsia timonensis | fixed | NA | groupFBVLED:time_pointFinal | 0.49 | 0.34 | 1.43 | 41.93 | -0.20 | 1.18 | 0.16 | 0.483 |
| Blautia_A MIC7810 | fixed | NA | groupFBVLED:time_pointFinal | 0.15 | 0.10 | 1.43 | 42.08 | -0.06 | 0.36 | 0.16 | 0.483 |
| Desulfovibrio piger | fixed | NA | groupFBVLED:time_pointFinal | -0.12 | 0.08 | -1.43 | 39.64 | -0.29 | 0.05 | 0.161 | 0.483 |
| Peptoniphilus_B duerdenii | fixed | NA | groupFBVLED:time_pointFinal | 0.24 | 0.17 | 1.41 | 84.00 | -0.10 | 0.58 | 0.162 | 0.485 |
| UBA1829 sp002338895 | fixed | NA | groupFBVLED:time_pointFinal | 0.16 | 0.11 | 1.42 | 43.24 | -0.07 | 0.39 | 0.164 | 0.487 |
| Prevotella sp001275135 | fixed | NA | groupFBVLED:time_pointFinal | 0.08 | 0.06 | 1.42 | 39.84 | -0.04 | 0.20 | 0.165 | 0.487 |
| Faecalibacterium prausnitzii_J | fixed | NA | groupFBVLED:time_pointFinal | 0.25 | 0.18 | 1.41 | 39.90 | -0.11 | 0.62 | 0.165 | 0.487 |
| Parasutterella sp000980495 | fixed | NA | groupFBVLED:time_pointFinal | 0.18 | 0.13 | 1.40 | 42.83 | -0.08 | 0.45 | 0.168 | 0.491 |
| Murdochiella MIC8251 | fixed | NA | groupFBVLED:time_pointFinal | 0.28 | 0.20 | 1.40 | 46.09 | -0.12 | 0.68 | 0.168 | 0.491 |
| UBA738 sp003522945 | fixed | NA | groupFBVLED:time_pointFinal | -0.30 | 0.22 | -1.36 | 42.92 | -0.75 | 0.15 | 0.182 | 0.527 |
| Faecalibacterium prausnitzii_C | fixed | NA | groupFBVLED:time_pointFinal | 0.43 | 0.31 | 1.35 | 42.64 | -0.21 | 1.06 | 0.183 | 0.527 |
| Lachnospiraceae MIC6885 | fixed | NA | groupFBVLED:time_pointFinal | 0.01 | 0.01 | 1.36 | 39.04 | 0.00 | 0.02 | 0.183 | 0.527 |
| Bacteroides_B vulgatus | fixed | NA | groupFBVLED:time_pointFinal | 0.52 | 0.39 | 1.35 | 40.04 | -0.26 | 1.30 | 0.185 | 0.527 |
| Coprobacter fastidiosus | fixed | NA | groupFBVLED:time_pointFinal | 0.36 | 0.27 | 1.35 | 41.13 | -0.18 | 0.89 | 0.186 | 0.527 |
| Peptoniphilus_A grossensis_B | fixed | NA | groupFBVLED:time_pointFinal | 0.57 | 0.42 | 1.34 | 42.06 | -0.28 | 1.42 | 0.186 | 0.527 |
| Acutalibacteraceae MIC6990 | fixed | NA | groupFBVLED:time_pointFinal | 0.34 | 0.26 | 1.34 | 43.42 | -0.17 | 0.86 | 0.186 | 0.527 |
| CAG 145 MIC7639 | fixed | NA | groupFBVLED:time_pointFinal | -0.13 | 0.10 | -1.33 | 43.49 | -0.32 | 0.07 | 0.191 | 0.535 |
| Bacteroides intestinalis_A | fixed | NA | groupFBVLED:time_pointFinal | -0.10 | 0.08 | -1.32 | 43.98 | -0.26 | 0.05 | 0.194 | 0.535 |
| Anaerotruncus sp900199635 | fixed | NA | groupFBVLED:time_pointFinal | 0.14 | 0.11 | 1.32 | 42.58 | -0.07 | 0.35 | 0.195 | 0.535 |
| Fusicatenibacter MIC7088 | fixed | NA | groupFBVLED:time_pointFinal | 0.10 | 0.08 | 1.30 | 84.00 | -0.05 | 0.25 | 0.195 | 0.535 |
| CAG 95 sp000438155 | fixed | NA | groupFBVLED:time_pointFinal | 0.25 | 0.19 | 1.31 | 46.24 | -0.13 | 0.64 | 0.197 | 0.535 |
| Lachnospira sp000436535 | fixed | NA | groupFBVLED:time_pointFinal | 0.23 | 0.18 | 1.31 | 37.33 | -0.13 | 0.59 | 0.198 | 0.535 |
| Lachnospirales MIC6978 | fixed | NA | groupFBVLED:time_pointFinal | -0.11 | 0.08 | -1.31 | 44.33 | -0.27 | 0.06 | 0.199 | 0.535 |
| Lachnospiraceae MIC9747 | fixed | NA | groupFBVLED:time_pointFinal | -0.19 | 0.14 | -1.30 | 44.38 | -0.48 | 0.10 | 0.199 | 0.535 |
| Ezakiella MIC8494 | fixed | NA | groupFBVLED:time_pointFinal | 0.32 | 0.25 | 1.29 | 84.00 | -0.17 | 0.81 | 0.2 | 0.535 |
| Gabonibacter massiliensis | fixed | NA | groupFBVLED:time_pointFinal | -0.09 | 0.07 | -1.30 | 42.16 | -0.23 | 0.05 | 0.201 | 0.535 |
| Christensenellales MIC6424 | fixed | NA | groupFBVLED:time_pointFinal | -0.05 | 0.04 | -1.30 | 39.88 | -0.14 | 0.03 | 0.202 | 0.535 |

| OUTCOMES - SPECIES | EFFECT | GROUP | TERM | ESTIMATE | STD ERROR | STATISTIC | DF | CONF LOW | CONF HIGH | P VALUE | Q VALUE |
|---|---|---|---|---|---|---|---|---|---|---|---|
| GCA 900066575 MIC7948 | fixed | NA | groupFBVLED:time_pointFinal | 0.24 | 0.19 | 1.30 | 43.26 | -0.14 | 0.62 | 0.202 | 0.535 |
| CAG 74 MIC8660 | fixed | NA | groupFBVLED:time_pointFinal | -0.13 | 0.10 | -1.29 | 43.37 | -0.33 | 0.07 | 0.202 | 0.535 |
| UBA11452 sp003526375 | fixed | NA | groupFBVLED:time_pointFinal | -0.17 | 0.13 | -1.29 | 42.11 | -0.44 | 0.10 | 0.203 | 0.535 |
| Clostridium_M citroniae | fixed | NA | groupFBVLED:time_pointFinal | -0.23 | 0.18 | -1.28 | 84.00 | -0.59 | 0.13 | 0.203 | 0.535 |
| Blautia_A sp900066355 | fixed | NA | groupFBVLED:time_pointFinal | 0.36 | 0.28 | 1.29 | 41.05 | -0.20 | 0.92 | 0.203 | 0.535 |
| Prevotella sp003447235 | fixed | NA | groupFBVLED:time_pointFinal | 0.38 | 0.29 | 1.29 | 39.05 | -0.21 | 0.97 | 0.205 | 0.535 |
| Acidaminococcus intestini | fixed | NA | groupFBVLED:time_pointFinal | 0.06 | 0.04 | 1.29 | 39.10 | -0.03 | 0.14 | 0.205 | 0.535 |
| Bacteroides xylanisolvens | fixed | NA | groupFBVLED:time_pointFinal | 0.48 | 0.38 | 1.29 | 41.34 | -0.28 | 1.24 | 0.206 | 0.535 |
| Alistipes sp000434235 | fixed | NA | groupFBVLED:time_pointFinal | 0.22 | 0.17 | 1.28 | 39.41 | -0.13 | 0.56 | 0.207 | 0.535 |
| CAG 83 sp001916855 | fixed | NA | groupFBVLED:time_pointFinal | 0.38 | 0.30 | 1.28 | 41.11 | -0.22 | 0.99 | 0.208 | 0.535 |
| CAG 170 MIC6856 | fixed | NA | groupFBVLED:time_pointFinal | 0.30 | 0.24 | 1.28 | 45.21 | -0.18 | 0.78 | 0.208 | 0.535 |
| Parvimonas sp001553085 | fixed | NA | groupFBVLED:time_pointFinal | -0.07 | 0.05 | -1.28 | 39.83 | -0.17 | 0.04 | 0.209 | 0.536 |
| UBA5394 MIC7155 | fixed | NA | groupFBVLED:time_pointFinal | -0.21 | 0.16 | -1.27 | 45.68 | -0.54 | 0.12 | 0.211 | 0.537 |
| Alistipes_A indistinctus | fixed | NA | groupFBVLED:time_pointFinal | -0.37 | 0.29 | -1.26 | 43.30 | -0.95 | 0.22 | 0.215 | 0.544 |
| UBA1394 MIC7236 | fixed | NA | groupFBVLED:time_pointFinal | -0.08 | 0.06 | -1.26 | 39.06 | -0.21 | 0.05 | 0.216 | 0.544 |
| Lachnospiraceae MIC6593 | fixed | NA | groupFBVLED:time_pointFinal | 0.18 | 0.14 | 1.26 | 31.45 | -0.11 | 0.48 | 0.216 | 0.544 |
| Faecalicatena torques | fixed | NA | groupFBVLED:time_pointFinal | -0.75 | 0.60 | -1.25 | 42.47 | -1.95 | 0.46 | 0.218 | 0.545 |
| Dorea sp000509125 | fixed | NA | groupFBVLED:time_pointFinal | -0.20 | 0.16 | -1.25 | 43.10 | -0.53 | 0.12 | 0.218 | 0.545 |
| Anaerovoracaceae MIC7161 | fixed | NA | groupFBVLED:time_pointFinal | 0.10 | 0.08 | 1.24 | 45.69 | -0.06 | 0.27 | 0.221 | 0.55 |
| Anaerococcus prevotii_A | fixed | NA | groupFBVLED:time_pointFinal | 0.22 | 0.18 | 1.23 | 84.00 | -0.14 | 0.57 | 0.222 | 0.551 |
| Levyella massiliensis | fixed | NA | groupFBVLED:time_pointFinal | -0.37 | 0.30 | -1.23 | 43.46 | -0.99 | 0.24 | 0.224 | 0.553 |
| CAG 74 MIC9156 | fixed | NA | groupFBVLED:time_pointFinal | -0.41 | 0.33 | -1.23 | 45.99 | -1.07 | 0.26 | 0.226 | 0.555 |
| PeH17 sp000435055 | fixed | NA | groupFBVLED:time_pointFinal | 0.58 | 0.48 | 1.20 | 42.13 | -0.39 | 1.56 | 0.236 | 0.577 |
| Lactobacillus_C paracasei | fixed | NA | groupFBVLED:time_pointFinal | 0.14 | 0.12 | 1.19 | 84.00 | -0.10 | 0.38 | 0.237 | 0.577 |
| Holdemania massiliensis | fixed | NA | groupFBVLED:time_pointFinal | 0.12 | 0.10 | 1.19 | 84.00 | -0.08 | 0.32 | 0.238 | 0.579 |
| Peptoniphilus_A MIC9267 | fixed | NA | groupFBVLED:time_pointFinal | 0.35 | 0.29 | 1.19 | 45.82 | -0.24 | 0.93 | 0.242 | 0.581 |
| UBA7182 MIC8257 | fixed | NA | groupFBVLED:time_pointFinal | 0.13 | 0.11 | 1.19 | 44.36 | -0.09 | 0.35 | 0.242 | 0.581 |
| UCG 010 sp003150215 | fixed | NA | groupFBVLED:time_pointFinal | -0.20 | 0.17 | -1.18 | 84.00 | -0.54 | 0.14 | 0.243 | 0.581 |
| Anaerococcus MIC7502 | fixed | NA | groupFBVLED:time_pointFinal | 0.22 | 0.19 | 1.18 | 40.66 | -0.16 | 0.59 | 0.243 | 0.581 |

| OUTCOMES - SPECIES | EFFECT | GROUP | TERM | ESTIMATE | STD ERROR | STATISTIC | DF | CONF LOW | CONF HIGH | P VALUE | Q VALUE |
|---|---|---|---|---|---|---|---|---|---|---|---|
| Tyzzerella MIC9817 | fixed | NA | groupFBVLED:time_pointFinal | -0.19 | 0.16 | -1.18 | 42.46 | -0.53 | 0.14 | 0.245 | 0.584 |
| Bacteroides_A plebeius_A | fixed | NA | groupFBVLED:time_pointFinal | 0.27 | 0.23 | 1.17 | 41.94 | -0.20 | 0.74 | 0.25 | 0.594 |
| CAG 74 MIC8853 | fixed | NA | groupFBVLED:time_pointFinal | -0.07 | 0.06 | -1.16 | 40.75 | -0.20 | 0.05 | 0.252 | 0.594 |
| UBA5446 MIC7746 | fixed | NA | groupFBVLED:time_pointFinal | -0.32 | 0.28 | -1.15 | 44.89 | -0.89 | 0.24 | 0.254 | 0.594 |
| Anaerostipes hadrus_A | fixed | NA | groupFBVLED:time_pointFinal | -0.36 | 0.31 | -1.16 | 41.15 | -0.99 | 0.27 | 0.254 | 0.594 |
| CAG 267 sp001917135 | fixed | NA | groupFBVLED:time_pointFinal | -0.25 | 0.21 | -1.16 | 39.66 | -0.68 | 0.19 | 0.254 | 0.594 |
| Negativibacillus MIC7916 | fixed | NA | groupFBVLED:time_pointFinal | 0.15 | 0.13 | 1.15 | 43.69 | -0.11 | 0.40 | 0.258 | 0.6 |
| S5 A14a MIC8631 | fixed | NA | groupFBVLED:time_pointFinal | 0.18 | 0.16 | 1.13 | 84.00 | -0.13 | 0.49 | 0.26 | 0.603 |
| CAG 312 sp000438015 | fixed | NA | groupFBVLED:time_pointFinal | 0.12 | 0.10 | 1.14 | 43.00 | -0.09 | 0.32 | 0.261 | 0.604 |
| Mogibacterium massiliense | fixed | NA | groupFBVLED:time_pointFinal | 0.31 | 0.27 | 1.13 | 84.00 | -0.24 | 0.86 | 0.263 | 0.604 |
| CAG 269 sp001915995 | fixed | NA | groupFBVLED:time_pointFinal | -0.05 | 0.04 | -1.13 | 42.13 | -0.13 | 0.04 | 0.266 | 0.604 |
| UBA11471 sp000434215 | fixed | NA | groupFBVLED:time_pointFinal | 0.27 | 0.24 | 1.13 | 40.39 | -0.21 | 0.74 | 0.266 | 0.604 |
| Mailhella sp003150275 | fixed | NA | groupFBVLED:time_pointFinal | -0.07 | 0.07 | -1.13 | 42.27 | -0.21 | 0.06 | 0.267 | 0.604 |
| UBA866 MIC8205 | fixed | NA | groupFBVLED:time_pointFinal | -0.18 | 0.16 | -1.13 | 43.70 | -0.50 | 0.14 | 0.267 | 0.604 |
| Acutalibacter MIC8974 | fixed | NA | groupFBVLED:time_pointFinal | -0.18 | 0.16 | -1.12 | 84.00 | -0.51 | 0.14 | 0.268 | 0.605 |
| Acetatifactor sp900066565 | fixed | NA | groupFBVLED:time_pointFinal | 0.54 | 0.49 | 1.12 | 42.31 | -0.44 | 1.53 | 0.271 | 0.607 |
| CAG 81 sp900066785 | fixed | NA | groupFBVLED:time_pointFinal | 0.19 | 0.17 | 1.11 | 41.43 | -0.15 | 0.53 | 0.272 | 0.607 |
| Clostridium_Q sp000435655 | fixed | NA | groupFBVLED:time_pointFinal | 0.14 | 0.12 | 1.12 | 29.06 | -0.11 | 0.39 | 0.272 | 0.607 |
| CAG 74 MIC8780 | fixed | NA | groupFBVLED:time_pointFinal | -0.08 | 0.07 | -1.11 | 42.51 | -0.22 | 0.06 | 0.274 | 0.607 |
| UBA1394 sp900066845 | fixed | NA | groupFBVLED:time_pointFinal | 0.27 | 0.24 | 1.11 | 41.90 | -0.22 | 0.75 | 0.274 | 0.607 |
| Holdemania sp900120005 | fixed | NA | groupFBVLED:time_pointFinal | 0.10 | 0.09 | 1.10 | 84.00 | -0.08 | 0.29 | 0.276 | 0.607 |
| CAG 83 sp000435975 | fixed | NA | groupFBVLED:time_pointFinal | -0.38 | 0.35 | -1.10 | 40.81 | -1.08 | 0.32 | 0.276 | 0.607 |
| CAG 45 sp000438375 | fixed | NA | groupFBVLED:time_pointFinal | 0.33 | 0.30 | 1.10 | 45.30 | -0.27 | 0.92 | 0.276 | 0.607 |
| CAG 417 sp000432835 | fixed | NA | groupFBVLED:time_pointFinal | 0.20 | 0.19 | 1.10 | 43.47 | -0.17 | 0.58 | 0.279 | 0.607 |
| CAG 110 MIC9276 | fixed | NA | groupFBVLED:time_pointFinal | -0.29 | 0.26 | -1.09 | 45.40 | -0.81 | 0.24 | 0.279 | 0.607 |
| Clostridium_M sp000431375 | fixed | NA | groupFBVLED:time_pointFinal | 0.43 | 0.39 | 1.10 | 40.31 | -0.36 | 1.22 | 0.279 | 0.607 |
| Collinsella MIC8209 | fixed | NA | groupFBVLED:time_pointFinal | -0.14 | 0.13 | -1.07 | 42.34 | -0.39 | 0.12 | 0.289 | 0.625 |
| Lactococcus lactis | fixed | NA | groupFBVLED:time_pointFinal | -0.23 | 0.21 | -1.07 | 46.19 | -0.66 | 0.20 | 0.291 | 0.626 |
| CAG 382 MIC9861 | fixed | NA | groupFBVLED:time_pointFinal | 0.07 | 0.07 | 1.07 | 45.64 | -0.06 | 0.21 | 0.291 | 0.626 |

| OUTCOMES - SPECIES | EFFECT | GROUP | TERM | ESTIMATE | STD ERROR | STATISTIC | DF | CONF LOW | CONF HIGH | P VALUE | Q VALUE |
|---|---|---|---|---|---|---|---|---|---|---|---|
| Acutalibacteraceae MIC7795 | fixed | NA | groupFBVLED:time_pointFinal | -0.23 | 0.22 | -1.06 | 41.33 | -0.66 | 0.21 | 0.293 | 0.628 |
| Desulfovibrio piger_A | fixed | NA | groupFBVLED:time_pointFinal | -0.06 | 0.05 | -1.06 | 41.37 | -0.16 | 0.05 | 0.295 | 0.629 |
| CAG 83 sp000435555 | fixed | NA | groupFBVLED:time_pointFinal | -0.42 | 0.39 | -1.06 | 43.76 | -1.21 | 0.38 | 0.297 | 0.629 |
| Bacteroides clarus | fixed | NA | groupFBVLED:time_pointFinal | 0.20 | 0.19 | 1.06 | 39.53 | -0.19 | 0.59 | 0.297 | 0.629 |
| Blautia producta | fixed | NA | groupFBVLED:time_pointFinal | -0.19 | 0.18 | -1.05 | 45.49 | -0.56 | 0.18 | 0.298 | 0.629 |
| UBA644 MIC9596 | fixed | NA | groupFBVLED:time_pointFinal | -0.11 | 0.10 | -1.04 | 43.99 | -0.31 | 0.10 | 0.302 | 0.633 |
| Lachnospiraceae MIC9280 | fixed | NA | groupFBVLED:time_pointFinal | -0.17 | 0.16 | -1.04 | 41.02 | -0.48 | 0.15 | 0.302 | 0.633 |
| CAG 170 sp003516765 | fixed | NA | groupFBVLED:time_pointFinal | 0.18 | 0.17 | 1.04 | 43.56 | -0.17 | 0.53 | 0.303 | 0.633 |
| Agathobacter MIC9834 | fixed | NA | groupFBVLED:time_pointFinal | 0.19 | 0.19 | 1.03 | 84.00 | -0.18 | 0.56 | 0.304 | 0.634 |
| Streptococcus parasanguinis_B | fixed | NA | groupFBVLED:time_pointFinal | 0.21 | 0.20 | 1.03 | 84.00 | -0.19 | 0.60 | 0.306 | 0.634 |
| CAG 110 MIC9052 | fixed | NA | groupFBVLED:time_pointFinal | -0.18 | 0.17 | -1.04 | 40.91 | -0.52 | 0.17 | 0.306 | 0.634 |
| Urmitella timonensis | fixed | NA | groupFBVLED:time_pointFinal | -0.32 | 0.31 | -1.03 | 43.85 | -0.94 | 0.30 | 0.309 | 0.637 |
| CAG 145 sp000435715 | fixed | NA | groupFBVLED:time_pointFinal | -0.30 | 0.30 | -1.03 | 43.64 | -0.90 | 0.29 | 0.31 | 0.637 |
| Anaerovoracaceae MIC7478 | fixed | NA | groupFBVLED:time_pointFinal | 0.16 | 0.16 | 1.02 | 39.84 | -0.16 | 0.48 | 0.312 | 0.637 |
| Blautia_A MIC7077 | fixed | NA | groupFBVLED:time_pointFinal | 0.31 | 0.31 | 1.02 | 40.94 | -0.31 | 0.93 | 0.313 | 0.637 |
| Prevotella bivia | fixed | NA | groupFBVLED:time_pointFinal | -0.17 | 0.16 | -1.01 | 84.00 | -0.49 | 0.16 | 0.313 | 0.637 |
| CAG 74 MIC8717 | fixed | NA | groupFBVLED:time_pointFinal | 0.13 | 0.13 | 1.01 | 84.00 | -0.13 | 0.39 | 0.315 | 0.637 |
| Acetatifactor sp003447295 | fixed | NA | groupFBVLED:time_pointFinal | 0.22 | 0.22 | 1.02 | 45.16 | -0.22 | 0.66 | 0.315 | 0.637 |
| Oscillospiraceae MIC9346 | fixed | NA | groupFBVLED:time_pointFinal | 0.12 | 0.12 | 1.01 | 41.52 | -0.12 | 0.35 | 0.319 | 0.642 |
| UBA9502 MIC7149 | fixed | NA | groupFBVLED:time_pointFinal | 0.17 | 0.16 | 1.01 | 40.86 | -0.17 | 0.50 | 0.32 | 0.642 |
| UBA5446 MIC7134 | fixed | NA | groupFBVLED:time_pointFinal | -0.33 | 0.33 | -1.00 | 45.88 | -0.99 | 0.33 | 0.321 | 0.642 |
| CAG 312 MIC7045 | fixed | NA | groupFBVLED:time_pointFinal | -0.09 | 0.10 | -0.99 | 42.66 | -0.29 | 0.10 | 0.33 | 0.657 |
| UBA7182 MIC8422 | fixed | NA | groupFBVLED:time_pointFinal | 0.17 | 0.17 | 0.99 | 41.05 | -0.18 | 0.52 | 0.33 | 0.657 |
| Oscillospiraceae MIC9482 | fixed | NA | groupFBVLED:time_pointFinal | 0.13 | 0.13 | 0.98 | 44.26 | -0.14 | 0.40 | 0.333 | 0.66 |
| Negativibacillus massiliensis | fixed | NA | groupFBVLED:time_pointFinal | 0.04 | 0.04 | 0.98 | 39.68 | -0.04 | 0.11 | 0.335 | 0.662 |
| CAG 288 sp000437395 | fixed | NA | groupFBVLED:time_pointFinal | 0.23 | 0.24 | 0.97 | 42.95 | -0.25 | 0.71 | 0.336 | 0.662 |
| Streptococcus anginosus_C | fixed | NA | groupFBVLED:time_pointFinal | 0.20 | 0.21 | 0.96 | 84.00 | -0.21 | 0.61 | 0.34 | 0.665 |
| 51 20 sp001917175 | fixed | NA | groupFBVLED:time_pointFinal | 0.29 | 0.31 | 0.96 | 41.27 | -0.32 | 0.91 | 0.341 | 0.665 |
| CAG 83 sp000431575 | fixed | NA | groupFBVLED:time_pointFinal | 0.23 | 0.24 | 0.96 | 40.43 | -0.26 | 0.72 | 0.343 | 0.665 |

| OUTCOMES - SPECIES | EFFECT | GROUP | TERM | ESTIMATE | STD ERROR | STATISTIC | DF | CONF LOW | CONF HIGH | P VALUE | Q VALUE |
|---|---|---|---|---|---|---|---|---|---|---|---|
| Ruminococcus_C sp000980705 | fixed | NA | groupFBVLED:time_pointFinal | 0.34 | 0.35 | 0.96 | 42.84 | -0.37 | 1.04 | 0.344 | 0.665 |
| CAG 272 MIC9176 | fixed | NA | groupFBVLED:time_pointFinal | -0.02 | 0.02 | -0.96 | 38.31 | -0.06 | 0.02 | 0.345 | 0.665 |
| Bacteroides caccae | fixed | NA | groupFBVLED:time_pointFinal | 0.29 | 0.30 | 0.96 | 40.60 | -0.32 | 0.89 | 0.345 | 0.665 |
| Blautia_A hydrogenotrophica | fixed | NA | groupFBVLED:time_pointFinal | -0.17 | 0.18 | -0.96 | 40.38 | -0.53 | 0.19 | 0.345 | 0.665 |
| Phil1 sp001940855 | fixed | NA | groupFBVLED:time_pointFinal | 0.38 | 0.40 | 0.95 | 43.29 | -0.43 | 1.19 | 0.346 | 0.666 |
| Bifidobacterium dentium | fixed | NA | groupFBVLED:time_pointFinal | -0.17 | 0.18 | -0.95 | 31.28 | -0.52 | 0.19 | 0.347 | 0.666 |
| Clostridium MIC8573 | fixed | NA | groupFBVLED:time_pointFinal | 0.30 | 0.31 | 0.95 | 45.96 | -0.34 | 0.93 | 0.349 | 0.666 |
| Acutalibacter MIC8741 | fixed | NA | groupFBVLED:time_pointFinal | 0.11 | 0.11 | 0.94 | 45.42 | -0.12 | 0.33 | 0.352 | 0.666 |
| Adlercreutzia equolifaciens | fixed | NA | groupFBVLED:time_pointFinal | 0.15 | 0.16 | 0.94 | 39.48 | -0.17 | 0.46 | 0.352 | 0.666 |
| Senegalimassilia anaerobia | fixed | NA | groupFBVLED:time_pointFinal | -0.13 | 0.14 | -0.94 | 40.56 | -0.41 | 0.15 | 0.354 | 0.666 |
| Oscillibacter MIC7169 | fixed | NA | groupFBVLED:time_pointFinal | 0.13 | 0.14 | 0.93 | 44.81 | -0.15 | 0.41 | 0.355 | 0.666 |
| Facklamia hominis | fixed | NA | groupFBVLED:time_pointFinal | 0.20 | 0.22 | 0.93 | 84.00 | -0.23 | 0.63 | 0.356 | 0.666 |
| Dorea formicigenerans | fixed | NA | groupFBVLED:time_pointFinal | 0.19 | 0.21 | 0.93 | 41.71 | -0.22 | 0.61 | 0.357 | 0.666 |
| Butyricimonas virosa | fixed | NA | groupFBVLED:time_pointFinal | -0.19 | 0.21 | -0.93 | 40.58 | -0.61 | 0.23 | 0.357 | 0.666 |
| Christensenellaceae MIC9015 | fixed | NA | groupFBVLED:time_pointFinal | -0.13 | 0.14 | -0.92 | 83.95 | -0.40 | 0.15 | 0.359 | 0.666 |
| Haemophilus_D parainfluenzae | fixed | NA | groupFBVLED:time_pointFinal | -0.17 | 0.18 | -0.93 | 45.78 | -0.53 | 0.20 | 0.359 | 0.666 |
| Peptoniphilus_C coxii | fixed | NA | groupFBVLED:time_pointFinal | -0.14 | 0.15 | -0.93 | 36.29 | -0.44 | 0.16 | 0.359 | 0.666 |
| Coprobacter secundus | fixed | NA | groupFBVLED:time_pointFinal | -0.16 | 0.17 | -0.91 | 43.45 | -0.51 | 0.19 | 0.367 | 0.676 |
| UBA1390 sp002305315 | fixed | NA | groupFBVLED:time_pointFinal | -0.14 | 0.15 | -0.91 | 44.15 | -0.45 | 0.17 | 0.367 | 0.676 |
| Erysipelatoclostridium MIC9185 | fixed | NA | groupFBVLED:time_pointFinal | -0.11 | 0.12 | -0.91 | 38.24 | -0.34 | 0.13 | 0.368 | 0.676 |
| Sutterella wadsworthensis_A | fixed | NA | groupFBVLED:time_pointFinal | -0.24 | 0.27 | -0.90 | 38.82 | -0.78 | 0.30 | 0.372 | 0.681 |
| Porphyromonas somerae | fixed | NA | groupFBVLED:time_pointFinal | 0.15 | 0.17 | 0.89 | 84.00 | -0.18 | 0.48 | 0.374 | 0.681 |
| QALS01 MIC9566 | fixed | NA | groupFBVLED:time_pointFinal | 0.16 | 0.18 | 0.90 | 44.98 | -0.20 | 0.51 | 0.374 | 0.681 |
| UBA1255 MIC8514 | fixed | NA | groupFBVLED:time_pointFinal | -0.23 | 0.26 | -0.89 | 46.08 | -0.74 | 0.29 | 0.376 | 0.682 |
| Eubacterium_R sp000436835 | fixed | NA | groupFBVLED:time_pointFinal | 0.14 | 0.16 | 0.88 | 39.81 | -0.18 | 0.47 | 0.383 | 0.687 |
| CAG 194 sp000432915 | fixed | NA | groupFBVLED:time_pointFinal | -0.16 | 0.19 | -0.88 | 42.45 | -0.54 | 0.21 | 0.383 | 0.687 |
| UBA7160 MIC6745 | fixed | NA | groupFBVLED:time_pointFinal | 0.22 | 0.25 | 0.88 | 40.90 | -0.28 | 0.72 | 0.385 | 0.687 |
| Sutterella wadsworthensis_B | fixed | NA | groupFBVLED:time_pointFinal | -0.17 | 0.19 | -0.88 | 39.76 | -0.56 | 0.22 | 0.386 | 0.687 |
| Bacteroides cellulosilyticus | fixed | NA | groupFBVLED:time_pointFinal | 0.33 | 0.38 | 0.87 | 42.09 | -0.43 | 1.09 | 0.387 | 0.687 |

| OUTCOMES - SPECIES | EFFECT | GROUP | TERM | ESTIMATE | STD ERROR | STATISTIC | DF | CONF LOW | CONF HIGH | P VALUE | Q VALUE |
|---|---|---|---|---|---|---|---|---|---|---|---|
| Butyricimonas synergistica_A | fixed | NA | groupFBVLED:time_pointFinal | -0.18 | 0.20 | -0.87 | 40.73 | -0.59 | 0.23 | 0.389 | 0.687 |
| Bifidobacterium MIC7686 | fixed | NA | groupFBVLED:time_pointFinal | -0.11 | 0.12 | -0.87 | 42.21 | -0.35 | 0.14 | 0.389 | 0.687 |
| Blautia_A wexlerae | fixed | NA | groupFBVLED:time_pointFinal | 0.31 | 0.36 | 0.87 | 41.11 | -0.41 | 1.03 | 0.391 | 0.687 |
| Merdibacter MIC8457 | fixed | NA | groupFBVLED:time_pointFinal | -0.04 | 0.04 | -0.87 | 39.35 | -0.13 | 0.05 | 0.391 | 0.687 |
| Bifidobacterium pseudocatenulatum | fixed | NA | groupFBVLED:time_pointFinal | -0.22 | 0.25 | -0.87 | 37.71 | -0.73 | 0.29 | 0.391 | 0.687 |
| Campylobacter_B hominis | fixed | NA | groupFBVLED:time_pointFinal | 0.21 | 0.24 | 0.86 | 44.89 | -0.28 | 0.70 | 0.392 | 0.687 |
| Agathobacter MIC6414 | fixed | NA | groupFBVLED:time_pointFinal | 0.12 | 0.14 | 0.87 | 28.83 | -0.17 | 0.42 | 0.392 | 0.687 |
| Intestinimonas massiliensis | fixed | NA | groupFBVLED:time_pointFinal | -0.32 | 0.37 | -0.86 | 45.35 | -1.06 | 0.42 | 0.394 | 0.687 |
| CAG 138 MIC9630 | fixed | NA | groupFBVLED:time_pointFinal | -0.43 | 0.50 | -0.86 | 43.36 | -1.43 | 0.58 | 0.394 | 0.687 |
| Lachnospirales MIC9617 | fixed | NA | groupFBVLED:time_pointFinal | 0.13 | 0.15 | 0.86 | 42.95 | -0.17 | 0.43 | 0.395 | 0.687 |
| Massiliomicrobiota sp002160815 | fixed | NA | groupFBVLED:time_pointFinal | -0.08 | 0.09 | -0.86 | 41.11 | -0.26 | 0.10 | 0.397 | 0.687 |
| CAG 273 sp003507395 | fixed | NA | groupFBVLED:time_pointFinal | -0.21 | 0.24 | -0.86 | 42.44 | -0.69 | 0.28 | 0.397 | 0.687 |
| Hungatella hathewayi | fixed | NA | groupFBVLED:time_pointFinal | -0.33 | 0.39 | -0.84 | 45.53 | -1.12 | 0.46 | 0.403 | 0.695 |
| CAG 145 MIC9666 | fixed | NA | groupFBVLED:time_pointFinal | -0.09 | 0.10 | -0.83 | 46.11 | -0.30 | 0.12 | 0.409 | 0.703 |
| Blautia_A obeum | fixed | NA | groupFBVLED:time_pointFinal | 0.31 | 0.37 | 0.83 | 43.24 | -0.44 | 1.06 | 0.412 | 0.706 |
| Clostridium_Q symbiosum | fixed | NA | groupFBVLED:time_pointFinal | -0.17 | 0.20 | -0.82 | 46.00 | -0.57 | 0.24 | 0.416 | 0.709 |
| Ruminiclostridium_C sp000435295 | fixed | NA | groupFBVLED:time_pointFinal | -0.13 | 0.16 | -0.82 | 39.63 | -0.47 | 0.20 | 0.417 | 0.709 |
| Bacteroides sp900066265 | fixed | NA | groupFBVLED:time_pointFinal | 0.07 | 0.08 | 0.82 | 36.39 | -0.10 | 0.23 | 0.418 | 0.709 |
| Haemophilus_D sp001815355 | fixed | NA | groupFBVLED:time_pointFinal | -0.12 | 0.14 | -0.81 | 84.00 | -0.40 | 0.17 | 0.419 | 0.709 |
| Coprococcus_B MIC8649 | fixed | NA | groupFBVLED:time_pointFinal | -0.02 | 0.03 | -0.82 | 38.32 | -0.09 | 0.04 | 0.419 | 0.709 |
| CAG 110 sp003525905 | fixed | NA | groupFBVLED:time_pointFinal | -0.30 | 0.37 | -0.81 | 42.29 | -1.05 | 0.45 | 0.422 | 0.712 |
| Eubacterium_I ramulus_A | fixed | NA | groupFBVLED:time_pointFinal | 0.14 | 0.18 | 0.81 | 43.11 | -0.21 | 0.50 | 0.424 | 0.713 |
| Peptococcaceae MIC7269 | fixed | NA | groupFBVLED:time_pointFinal | 0.14 | 0.18 | 0.80 | 41.93 | -0.22 | 0.51 | 0.427 | 0.716 |
| UC5 1 2E3 sp001304875 | fixed | NA | groupFBVLED:time_pointFinal | -0.06 | 0.07 | -0.80 | 41.64 | -0.20 | 0.09 | 0.428 | 0.716 |
| UBA7160 MIC9207 | fixed | NA | groupFBVLED:time_pointFinal | 0.20 | 0.25 | 0.80 | 41.91 | -0.30 | 0.70 | 0.431 | 0.718 |
| Anaerococcus senegalensis | fixed | NA | groupFBVLED:time_pointFinal | 0.12 | 0.15 | 0.79 | 84.00 | -0.18 | 0.42 | 0.432 | 0.718 |
| CAG 272 MIC7215 | fixed | NA | groupFBVLED:time_pointFinal | 0.18 | 0.23 | 0.79 | 45.56 | -0.28 | 0.63 | 0.433 | 0.718 |
| CAG 302 sp000431795 | fixed | NA | groupFBVLED:time_pointFinal | 0.10 | 0.13 | 0.78 | 40.41 | -0.16 | 0.37 | 0.437 | 0.723 |
| Coprococcus sp900066115 | fixed | NA | groupFBVLED:time_pointFinal | 0.16 | 0.20 | 0.78 | 40.85 | -0.25 | 0.56 | 0.438 | 0.723 |

| OUTCOMES - SPECIES | EFFECT | GROUP | TERM | ESTIMATE | STD ERROR | STATISTIC | DF | CONF LOW | CONF HIGH | P VALUE | Q VALUE |
|---|---|---|---|---|---|---|---|---|---|---|---|
| Dorea sp900312975 | fixed | NA | groupFBVLED:time_pointFinal | -0.19 | 0.25 | -0.78 | 44.82 | -0.70 | 0.31 | 0.439 | 0.723 |
| Agathobacter rectale | fixed | NA | groupFBVLED:time_pointFinal | 0.38 | 0.49 | 0.78 | 41.16 | -0.62 | 1.38 | 0.442 | 0.726 |
| UCG 010 MIC8371 | fixed | NA | groupFBVLED:time_pointFinal | 0.10 | 0.13 | 0.77 | 42.37 | -0.16 | 0.36 | 0.447 | 0.731 |
| CAG 274 sp000432155 | fixed | NA | groupFBVLED:time_pointFinal | 0.22 | 0.29 | 0.77 | 40.14 | -0.37 | 0.81 | 0.448 | 0.731 |
| Slackia_A MIC8451 | fixed | NA | groupFBVLED:time_pointFinal | 0.02 | 0.02 | 0.77 | 40.11 | -0.03 | 0.07 | 0.449 | 0.731 |
| Flavonifractor MIC8104 | fixed | NA | groupFBVLED:time_pointFinal | -0.18 | 0.23 | -0.76 | 84.00 | -0.64 | 0.29 | 0.451 | 0.733 |
| Fusicatenibacter saccharivorans | fixed | NA | groupFBVLED:time_pointFinal | 0.35 | 0.46 | 0.76 | 42.93 | -0.58 | 1.28 | 0.454 | 0.736 |
| QALW01 sp003150515 | fixed | NA | groupFBVLED:time_pointFinal | -0.10 | 0.13 | -0.75 | 41.98 | -0.35 | 0.16 | 0.457 | 0.736 |
| Anaerotruncus colihominis | fixed | NA | groupFBVLED:time_pointFinal | -0.12 | 0.15 | -0.75 | 43.54 | -0.43 | 0.20 | 0.459 | 0.736 |
| CAG 103 MIC6381 | fixed | NA | groupFBVLED:time_pointFinal | 0.29 | 0.39 | 0.74 | 45.94 | -0.49 | 1.07 | 0.46 | 0.736 |
| CAG 83 sp003539495 | fixed | NA | groupFBVLED:time_pointFinal | 0.05 | 0.07 | 0.74 | 40.86 | -0.09 | 0.20 | 0.461 | 0.736 |
| Roseburia intestinalis | fixed | NA | groupFBVLED:time_pointFinal | 0.41 | 0.55 | 0.74 | 36.47 | -0.70 | 1.52 | 0.461 | 0.736 |
| Peptoniphilus_A lacrimalis | fixed | NA | groupFBVLED:time_pointFinal | 0.31 | 0.41 | 0.74 | 42.70 | -0.53 | 1.14 | 0.461 | 0.736 |
| UBA737 MIC7964 | fixed | NA | groupFBVLED:time_pointFinal | -0.02 | 0.03 | -0.74 | 39.85 | -0.09 | 0.04 | 0.464 | 0.738 |
| Bacteroides uniformis | fixed | NA | groupFBVLED:time_pointFinal | -0.29 | 0.39 | -0.74 | 43.06 | -1.08 | 0.50 | 0.465 | 0.738 |
| TF01 11 sp000436755 | fixed | NA | groupFBVLED:time_pointFinal | 0.08 | 0.11 | 0.74 | 43.99 | -0.14 | 0.31 | 0.466 | 0.738 |
| Blautia_A MIC9206 | fixed | NA | groupFBVLED:time_pointFinal | -0.05 | 0.07 | -0.73 | 39.97 | -0.20 | 0.10 | 0.471 | 0.743 |
| CAG 56 sp900066615 | fixed | NA | groupFBVLED:time_pointFinal | 0.31 | 0.43 | 0.72 | 40.71 | -0.56 | 1.18 | 0.475 | 0.746 |
| QAMH01 MIC6543 | fixed | NA | groupFBVLED:time_pointFinal | 0.11 | 0.15 | 0.72 | 40.70 | -0.19 | 0.41 | 0.477 | 0.746 |
| S5 A14a sp000758905 | fixed | NA | groupFBVLED:time_pointFinal | -0.14 | 0.19 | -0.71 | 84.00 | -0.51 | 0.24 | 0.478 | 0.746 |
| Ezakiella coagulans | fixed | NA | groupFBVLED:time_pointFinal | 0.20 | 0.28 | 0.71 | 84.00 | -0.35 | 0.74 | 0.48 | 0.746 |
| Ruminococcus_C sp000437175 | fixed | NA | groupFBVLED:time_pointFinal | 0.09 | 0.13 | 0.71 | 38.28 | -0.17 | 0.35 | 0.48 | 0.746 |
| Angelakisella MIC6791 | fixed | NA | groupFBVLED:time_pointFinal | 0.16 | 0.23 | 0.71 | 41.90 | -0.30 | 0.63 | 0.48 | 0.746 |
| Clostridium sp000435835 | fixed | NA | groupFBVLED:time_pointFinal | 0.26 | 0.37 | 0.71 | 44.83 | -0.48 | 1.00 | 0.481 | 0.746 |
| Lawsonibacter asaccharolyticus | fixed | NA | groupFBVLED:time_pointFinal | -0.16 | 0.22 | -0.71 | 39.99 | -0.60 | 0.29 | 0.484 | 0.749 |
| Ruminococcus_C sp000433635 | fixed | NA | groupFBVLED:time_pointFinal | 0.28 | 0.40 | 0.70 | 40.95 | -0.52 | 1.08 | 0.487 | 0.752 |
| Oscillospiraceae MIC8511 | fixed | NA | groupFBVLED:time_pointFinal | 0.07 | 0.10 | 0.68 | 84.00 | -0.13 | 0.27 | 0.497 | 0.763 |
| CAG 272 MIC9439 | fixed | NA | groupFBVLED:time_pointFinal | -0.10 | 0.15 | -0.69 | 42.21 | -0.41 | 0.20 | 0.497 | 0.763 |
| Gemmiger MIC8198 | fixed | NA | groupFBVLED:time_pointFinal | -0.07 | 0.10 | -0.68 | 40.44 | -0.28 | 0.14 | 0.499 | 0.763 |

| OUTCOMES - SPECIES | EFFECT | GROUP | TERM | ESTIMATE | STD ERROR | STATISTIC | DF | CONF LOW | CONF HIGH | P VALUE | Q VALUE |
|---|---|---|---|---|---|---|---|---|---|---|---|
| Acutalibacter sp000435395 | fixed | NA | groupFBVLED:time_pointFinal | -0.06 | 0.09 | -0.68 | 41.67 | -0.25 | 0.13 | 0.499 | 0.763 |
| Bacteroides thetaiotaomicron | fixed | NA | groupFBVLED:time_pointFinal | -0.27 | 0.39 | -0.68 | 41.41 | -1.06 | 0.53 | 0.502 | 0.763 |
| Holdemanella MIC8202 | fixed | NA | groupFBVLED:time_pointFinal | 0.06 | 0.08 | 0.68 | 28.30 | -0.11 | 0.23 | 0.502 | 0.763 |
| Coprococcus eutactus_A | fixed | NA | groupFBVLED:time_pointFinal | 0.30 | 0.44 | 0.68 | 39.82 | -0.59 | 1.19 | 0.503 | 0.763 |
| Paraprevotella clara | fixed | NA | groupFBVLED:time_pointFinal | 0.10 | 0.15 | 0.67 | 42.25 | -0.21 | 0.41 | 0.505 | 0.764 |
| CAG 727 MIC7825 | fixed | NA | groupFBVLED:time_pointFinal | -0.16 | 0.23 | -0.67 | 42.91 | -0.62 | 0.31 | 0.507 | 0.765 |
| CAG 1427 sp000436075 | fixed | NA | groupFBVLED:time_pointFinal | 0.07 | 0.11 | 0.67 | 38.52 | -0.15 | 0.30 | 0.509 | 0.767 |
| Streptococcus vestibularis | fixed | NA | groupFBVLED:time_pointFinal | 0.13 | 0.19 | 0.66 | 84.00 | -0.25 | 0.51 | 0.511 | 0.768 |
| Tidjanibacter inops | fixed | NA | groupFBVLED:time_pointFinal | -0.06 | 0.09 | -0.66 | 39.66 | -0.25 | 0.13 | 0.513 | 0.768 |
| Bacteroides_A MIC8078 | fixed | NA | groupFBVLED:time_pointFinal | 0.04 | 0.06 | 0.66 | 41.42 | -0.09 | 0.17 | 0.515 | 0.77 |
| Ruminococcaceae MIC8156 | fixed | NA | groupFBVLED:time_pointFinal | 0.06 | 0.09 | 0.64 | 43.14 | -0.13 | 0.25 | 0.526 | 0.782 |
| Streptococcus sp001556435 | fixed | NA | groupFBVLED:time_pointFinal | 0.17 | 0.27 | 0.64 | 44.21 | -0.37 | 0.71 | 0.527 | 0.782 |
| Lawsonella MIC8032 | fixed | NA | groupFBVLED:time_pointFinal | 0.19 | 0.31 | 0.63 | 84.00 | -0.42 | 0.80 | 0.531 | 0.782 |
| ER4 sp003522105 | fixed | NA | groupFBVLED:time_pointFinal | -0.18 | 0.29 | -0.63 | 43.37 | -0.76 | 0.40 | 0.532 | 0.782 |
| Faecalibacterium prausnitzii_K | fixed | NA | groupFBVLED:time_pointFinal | 0.22 | 0.36 | 0.63 | 39.61 | -0.49 | 0.94 | 0.532 | 0.782 |
| Hungatella_A MIC8772 | fixed | NA | groupFBVLED:time_pointFinal | 0.11 | 0.18 | 0.63 | 43.14 | -0.25 | 0.48 | 0.535 | 0.782 |
| Coprococcus eutactus | fixed | NA | groupFBVLED:time_pointFinal | -0.13 | 0.21 | -0.62 | 42.06 | -0.54 | 0.29 | 0.536 | 0.782 |
| Peptoniphilus_C MIC7895 | fixed | NA | groupFBVLED:time_pointFinal | 0.14 | 0.23 | 0.62 | 84.00 | -0.31 | 0.59 | 0.536 | 0.782 |
| Bifidobacterium vaginale | fixed | NA | groupFBVLED:time_pointFinal | 0.14 | 0.22 | 0.62 | 84.00 | -0.30 | 0.58 | 0.536 | 0.782 |
| CAG 115 sp003531585 | fixed | NA | groupFBVLED:time_pointFinal | 0.26 | 0.41 | 0.62 | 40.72 | -0.57 | 1.09 | 0.537 | 0.782 |
| Porphyromonas MIC7597 | fixed | NA | groupFBVLED:time_pointFinal | -0.04 | 0.07 | -0.63 | 26.64 | -0.19 | 0.10 | 0.537 | 0.782 |
| Ruminococcaceae MIC7581 | fixed | NA | groupFBVLED:time_pointFinal | -0.10 | 0.16 | -0.62 | 46.10 | -0.42 | 0.22 | 0.539 | 0.783 |
| CAG 273 sp000438355 | fixed | NA | groupFBVLED:time_pointFinal | -0.16 | 0.26 | -0.62 | 41.25 | -0.67 | 0.36 | 0.54 | 0.783 |
| UBA1417 MIC7387 | fixed | NA | groupFBVLED:time_pointFinal | -0.10 | 0.16 | -0.61 | 45.56 | -0.41 | 0.22 | 0.545 | 0.787 |
| CAG 170 MIC6396 | fixed | NA | groupFBVLED:time_pointFinal | 0.13 | 0.22 | 0.61 | 45.31 | -0.31 | 0.58 | 0.547 | 0.787 |
| Ruminococcus_C callidus | fixed | NA | groupFBVLED:time_pointFinal | 0.22 | 0.37 | 0.60 | 42.09 | -0.52 | 0.97 | 0.549 | 0.787 |
| Clostridium_P perfringens | fixed | NA | groupFBVLED:time_pointFinal | 0.06 | 0.11 | 0.60 | 33.97 | -0.15 | 0.28 | 0.55 | 0.787 |
| Peptoniphilus_A harei | fixed | NA | groupFBVLED:time_pointFinal | 0.12 | 0.21 | 0.60 | 84.00 | -0.29 | 0.54 | 0.55 | 0.787 |
| Streptococcus salivarius | fixed | NA | groupFBVLED:time_pointFinal | 0.20 | 0.34 | 0.60 | 40.43 | -0.48 | 0.89 | 0.551 | 0.787 |

| OUTCOMES - SPECIES | EFFECT | GROUP | TERM | ESTIMATE | STD ERROR | STATISTIC | DF | CONF LOW | CONF HIGH | P VALUE | Q VALUE |
|---|---|---|---|---|---|---|---|---|---|---|---|
| Eubacterium_E hallii | fixed | NA | groupFBVLED:time_pointFinal | -0.18 | 0.30 | -0.60 | 41.56 | -0.77 | 0.42 | 0.554 | 0.788 |
| Prevotella timonensis | fixed | NA | groupFBVLED:time_pointFinal | -0.11 | 0.18 | -0.59 | 84.00 | -0.48 | 0.26 | 0.554 | 0.788 |
| Lawsonibacter sp900066825 | fixed | NA | groupFBVLED:time_pointFinal | -0.06 | 0.10 | -0.59 | 39.08 | -0.27 | 0.15 | 0.556 | 0.79 |
| Erysipelatoclostridium sp003024675 | fixed | NA | groupFBVLED:time_pointFinal | 0.04 | 0.07 | 0.59 | 39.33 | -0.11 | 0.19 | 0.559 | 0.79 |
| Streptococcus gordonii | fixed | NA | groupFBVLED:time_pointFinal | 0.07 | 0.12 | 0.59 | 84.00 | -0.17 | 0.31 | 0.559 | 0.79 |
| Holdemania filiformis | fixed | NA | groupFBVLED:time_pointFinal | -0.06 | 0.10 | -0.59 | 42.93 | -0.25 | 0.14 | 0.561 | 0.79 |
| UBA9502 MIC8595 | fixed | NA | groupFBVLED:time_pointFinal | -0.08 | 0.14 | -0.58 | 84.00 | -0.35 | 0.19 | 0.562 | 0.79 |
| CAG 83 MIC8701 | fixed | NA | groupFBVLED:time_pointFinal | -0.10 | 0.16 | -0.58 | 43.09 | -0.43 | 0.23 | 0.564 | 0.791 |
| UBA4285 MIC9245 | fixed | NA | groupFBVLED:time_pointFinal | -0.08 | 0.14 | -0.58 | 39.67 | -0.37 | 0.21 | 0.568 | 0.794 |
| Prevotella MIC8201 | fixed | NA | groupFBVLED:time_pointFinal | 0.11 | 0.19 | 0.57 | 46.05 | -0.28 | 0.49 | 0.57 | 0.794 |
| Agathobaculum MIC7900 | fixed | NA | groupFBVLED:time_pointFinal | 0.10 | 0.17 | 0.57 | 39.44 | -0.25 | 0.45 | 0.57 | 0.794 |
| QALW01 MIC7388 | fixed | NA | groupFBVLED:time_pointFinal | 0.04 | 0.08 | 0.57 | 84.00 | -0.11 | 0.19 | 0.571 | 0.794 |
| QAND01 MIC7514 | fixed | NA | groupFBVLED:time_pointFinal | -0.10 | 0.18 | -0.56 | 45.27 | -0.47 | 0.27 | 0.577 | 0.801 |
| GCA 900066495 MIC8689 | fixed | NA | groupFBVLED:time_pointFinal | -0.09 | 0.16 | -0.54 | 39.74 | -0.41 | 0.24 | 0.591 | 0.817 |
| Blautia_A MIC8910 | fixed | NA | groupFBVLED:time_pointFinal | -0.06 | 0.11 | -0.54 | 38.77 | -0.29 | 0.17 | 0.592 | 0.817 |
| Bacteroides_B dorei | fixed | NA | groupFBVLED:time_pointFinal | -0.18 | 0.33 | -0.54 | 39.65 | -0.85 | 0.49 | 0.592 | 0.817 |
| Marvinbryantia MIC9792 | fixed | NA | groupFBVLED:time_pointFinal | -0.05 | 0.09 | -0.53 | 43.61 | -0.22 | 0.13 | 0.596 | 0.82 |
| Fenollaria timonensis | fixed | NA | groupFBVLED:time_pointFinal | 0.13 | 0.24 | 0.53 | 45.78 | -0.36 | 0.61 | 0.6 | 0.82 |
| Collinsella sp003487125 | fixed | NA | groupFBVLED:time_pointFinal | -0.08 | 0.16 | -0.53 | 37.47 | -0.41 | 0.24 | 0.601 | 0.82 |
| CAG 269 MIC8518 | fixed | NA | groupFBVLED:time_pointFinal | 0.08 | 0.15 | 0.53 | 42.17 | -0.22 | 0.37 | 0.601 | 0.82 |
| Blautia_A obeum_B | fixed | NA | groupFBVLED:time_pointFinal | 0.04 | 0.08 | 0.52 | 44.40 | -0.12 | 0.21 | 0.603 | 0.82 |
| Blautia_A MIC8050 | fixed | NA | groupFBVLED:time_pointFinal | -0.19 | 0.36 | -0.52 | 42.93 | -0.90 | 0.53 | 0.606 | 0.82 |
| Collinsella MIC8447 | fixed | NA | groupFBVLED:time_pointFinal | -0.01 | 0.03 | -0.52 | 38.84 | -0.07 | 0.04 | 0.606 | 0.82 |
| Phascolarctobacterium faecium | fixed | NA | groupFBVLED:time_pointFinal | 0.10 | 0.20 | 0.52 | 39.71 | -0.30 | 0.50 | 0.607 | 0.82 |
| Lawsonella clevelandensis | fixed | NA | groupFBVLED:time_pointFinal | 0.09 | 0.17 | 0.52 | 84.00 | -0.26 | 0.43 | 0.608 | 0.82 |
| Oscillospirales MIC7398 | fixed | NA | groupFBVLED:time_pointFinal | -0.07 | 0.13 | -0.51 | 46.18 | -0.33 | 0.19 | 0.61 | 0.82 |
| Ruminiclostridium_C MIC7261 | fixed | NA | groupFBVLED:time_pointFinal | 0.05 | 0.10 | 0.51 | 40.81 | -0.16 | 0.26 | 0.61 | 0.82 |
| Acetatifactor sp900066365 | fixed | NA | groupFBVLED:time_pointFinal | -0.23 | 0.45 | -0.51 | 42.50 | -1.15 | 0.68 | 0.61 | 0.82 |
| CAG 273 sp003534295 | fixed | NA | groupFBVLED:time_pointFinal | 0.18 | 0.36 | 0.51 | 42.65 | -0.55 | 0.92 | 0.613 | 0.822 |

| OUTCOMES - SPECIES | EFFECT | GROUP | TERM | ESTIMATE | STD ERROR | STATISTIC | DF | CONF LOW | CONF HIGH | P VALUE | Q VALUE |
|---|---|---|---|---|---|---|---|---|---|---|---|
| Marseille P4683 sp900232885 | fixed | NA | groupFBVLED:time_pointFinal | -0.09 | 0.18 | -0.51 | 36.39 | -0.45 | 0.27 | 0.615 | 0.823 |
| Escherichia coli | fixed | NA | groupFBVLED:time_pointFinal | 0.21 | 0.41 | 0.50 | 41.61 | -0.63 | 1.04 | 0.618 | 0.824 |
| Faecalicatena sp000509105 | fixed | NA | groupFBVLED:time_pointFinal | -0.10 | 0.20 | -0.50 | 45.29 | -0.51 | 0.31 | 0.619 | 0.824 |
| Alistipes onderdonkii | fixed | NA | groupFBVLED:time_pointFinal | -0.23 | 0.46 | -0.50 | 43.49 | -1.15 | 0.70 | 0.622 | 0.826 |
| Eubacterium_G sp000432355 | fixed | NA | groupFBVLED:time_pointFinal | -0.09 | 0.18 | -0.49 | 44.84 | -0.45 | 0.27 | 0.624 | 0.828 |
| CAG 312 MIC7072 | fixed | NA | groupFBVLED:time_pointFinal | 0.08 | 0.17 | 0.49 | 41.88 | -0.26 | 0.43 | 0.628 | 0.83 |
| CAG 74 MIC8932 | fixed | NA | groupFBVLED:time_pointFinal | -0.07 | 0.15 | -0.48 | 42.78 | -0.37 | 0.23 | 0.635 | 0.838 |
| CAG 353 sp900066885 | fixed | NA | groupFBVLED:time_pointFinal | -0.13 | 0.27 | -0.47 | 43.06 | -0.66 | 0.41 | 0.64 | 0.843 |
| QANA01 MIC9070 | fixed | NA | groupFBVLED:time_pointFinal | 0.04 | 0.08 | 0.47 | 42.80 | -0.12 | 0.19 | 0.644 | 0.845 |
| CAG 354 MIC8844 | fixed | NA | groupFBVLED:time_pointFinal | 0.08 | 0.17 | 0.47 | 42.94 | -0.26 | 0.42 | 0.644 | 0.845 |
| UBA3818 MIC8425 | fixed | NA | groupFBVLED:time_pointFinal | -0.11 | 0.25 | -0.46 | 43.71 | -0.61 | 0.39 | 0.647 | 0.846 |
| Blautia_A sp000285855 | fixed | NA | groupFBVLED:time_pointFinal | 0.16 | 0.35 | 0.46 | 42.27 | -0.55 | 0.88 | 0.648 | 0.846 |
| CAG 83 MIC9166 | fixed | NA | groupFBVLED:time_pointFinal | -0.02 | 0.04 | -0.45 | 39.42 | -0.10 | 0.07 | 0.653 | 0.851 |
| Eubacterium_I ramulus | fixed | NA | groupFBVLED:time_pointFinal | -0.12 | 0.27 | -0.45 | 39.81 | -0.67 | 0.43 | 0.657 | 0.853 |
| Anaeromassilibacillus sp002159845 | fixed | NA | groupFBVLED:time_pointFinal | -0.05 | 0.12 | -0.44 | 84.00 | -0.29 | 0.18 | 0.658 | 0.853 |
| Finegoldia magna | fixed | NA | groupFBVLED:time_pointFinal | -0.19 | 0.44 | -0.44 | 39.88 | -1.09 | 0.70 | 0.661 | 0.856 |
| Butyricicoccus_A sp002395695 | fixed | NA | groupFBVLED:time_pointFinal | 0.08 | 0.17 | 0.44 | 44.45 | -0.27 | 0.43 | 0.663 | 0.856 |
| Faecalicatena faecis | fixed | NA | groupFBVLED:time_pointFinal | -0.13 | 0.30 | -0.43 | 40.39 | -0.74 | 0.48 | 0.667 | 0.859 |
| Enorma MIC9272 | fixed | NA | groupFBVLED:time_pointFinal | -0.02 | 0.04 | -0.43 | 40.54 | -0.11 | 0.07 | 0.668 | 0.859 |
| Peptostreptococcus anaerobius | fixed | NA | groupFBVLED:time_pointFinal | 0.05 | 0.12 | 0.43 | 43.11 | -0.19 | 0.29 | 0.669 | 0.859 |
| CAG 83 MIC8843 | fixed | NA | groupFBVLED:time_pointFinal | -0.04 | 0.09 | -0.43 | 39.97 | -0.21 | 0.14 | 0.672 | 0.861 |
| CAG 196 sp002102975 | fixed | NA | groupFBVLED:time_pointFinal | 0.13 | 0.30 | 0.42 | 38.80 | -0.48 | 0.74 | 0.676 | 0.865 |
| Bacteroides intestinalis | fixed | NA | groupFBVLED:time_pointFinal | 0.07 | 0.18 | 0.42 | 40.58 | -0.29 | 0.44 | 0.678 | 0.865 |
| Lactobacillus_B ruminis | fixed | NA | groupFBVLED:time_pointFinal | 0.03 | 0.08 | 0.41 | 39.49 | -0.12 | 0.18 | 0.686 | 0.874 |
| Eubacterium_G sp000434315 | fixed | NA | groupFBVLED:time_pointFinal | 0.10 | 0.24 | 0.40 | 44.50 | -0.39 | 0.59 | 0.69 | 0.876 |
| Escherichia coli_D | fixed | NA | groupFBVLED:time_pointFinal | 0.09 | 0.22 | 0.40 | 44.84 | -0.35 | 0.52 | 0.69 | 0.876 |
| Monoglobus pectinilyticus | fixed | NA | groupFBVLED:time_pointFinal | 0.10 | 0.25 | 0.40 | 45.24 | -0.41 | 0.61 | 0.693 | 0.878 |
| Turicibacter sanguinis | fixed | NA | groupFBVLED:time_pointFinal | 0.09 | 0.24 | 0.38 | 44.19 | -0.39 | 0.57 | 0.703 | 0.885 |
| Faecalibacterium prausnitzii_A | fixed | NA | groupFBVLED:time_pointFinal | -0.07 | 0.19 | -0.38 | 39.39 | -0.46 | 0.32 | 0.704 | 0.885 |

| OUTCOMES - SPECIES | EFFECT | GROUP | TERM | ESTIMATE | STD ERROR | STATISTIC | DF | CONF LOW | CONF HIGH | P VALUE | Q VALUE |
|---|---|---|---|---|---|---|---|---|---|---|---|
| UBA7102 MIC7325 | fixed | NA | groupFBVLED:time_pointFinal | 0.03 | 0.09 | 0.38 | 43.58 | -0.15 | 0.21 | 0.705 | 0.885 |
| CAG 180 sp000432435 | fixed | NA | groupFBVLED:time_pointFinal | 0.13 | 0.35 | 0.38 | 41.16 | -0.57 | 0.84 | 0.706 | 0.885 |
| Faecalibacterium MIC9210 | fixed | NA | groupFBVLED:time_pointFinal | -0.13 | 0.35 | -0.38 | 45.69 | -0.83 | 0.57 | 0.709 | 0.885 |
| KLE1615 sp900066985 | fixed | NA | groupFBVLED:time_pointFinal | 0.12 | 0.32 | 0.38 | 42.44 | -0.52 | 0.76 | 0.709 | 0.885 |
| Blautia_A sp900066335 | fixed | NA | groupFBVLED:time_pointFinal | -0.07 | 0.19 | -0.37 | 40.22 | -0.46 | 0.32 | 0.711 | 0.885 |
| Bacteroides ovatus | fixed | NA | groupFBVLED:time_pointFinal | -0.18 | 0.50 | -0.37 | 40.05 | -1.19 | 0.82 | 0.713 | 0.885 |
| Roseburia hominis | fixed | NA | groupFBVLED:time_pointFinal | 0.13 | 0.36 | 0.37 | 43.47 | -0.60 | 0.87 | 0.716 | 0.885 |
| Oscillibacter MIC6596 | fixed | NA | groupFBVLED:time_pointFinal | 0.02 | 0.05 | 0.36 | 43.52 | -0.09 | 0.13 | 0.718 | 0.885 |
| Prevotella copri | fixed | NA | groupFBVLED:time_pointFinal | 0.10 | 0.26 | 0.36 | 41.36 | -0.44 | 0.63 | 0.719 | 0.885 |
| Oscillibacter MIC7430 | fixed | NA | groupFBVLED:time_pointFinal | 0.03 | 0.08 | 0.36 | 84.00 | -0.13 | 0.19 | 0.72 | 0.885 |
| Phascolarctobacterium_A succinatutens | fixed | NA | groupFBVLED:time_pointFinal | 0.02 | 0.05 | 0.36 | 39.33 | -0.09 | 0.12 | 0.72 | 0.885 |
| Dorea sp000433535 | fixed | NA | groupFBVLED:time_pointFinal | 0.05 | 0.14 | 0.36 | 41.28 | -0.23 | 0.32 | 0.721 | 0.885 |
| Eubacterium_G sp000435815 | fixed | NA | groupFBVLED:time_pointFinal | 0.05 | 0.15 | 0.36 | 33.82 | -0.24 | 0.35 | 0.722 | 0.885 |
| Oscillibacter MIC7608 | fixed | NA | groupFBVLED:time_pointFinal | 0.06 | 0.15 | 0.36 | 44.45 | -0.26 | 0.37 | 0.723 | 0.885 |
| An200 MIC7965 | fixed | NA | groupFBVLED:time_pointFinal | -0.05 | 0.15 | -0.36 | 46.13 | -0.35 | 0.25 | 0.723 | 0.885 |
| UBA644 MIC9235 | fixed | NA | groupFBVLED:time_pointFinal | -0.08 | 0.22 | -0.34 | 43.91 | -0.52 | 0.37 | 0.734 | 0.894 |
| Fenollaria massiliensis | fixed | NA | groupFBVLED:time_pointFinal | 0.08 | 0.25 | 0.34 | 84.00 | -0.40 | 0.57 | 0.734 | 0.894 |
| UBA644_A MIC9239 | fixed | NA | groupFBVLED:time_pointFinal | 0.04 | 0.12 | 0.34 | 44.43 | -0.19 | 0.27 | 0.735 | 0.894 |
| Intestinibacter MIC8174 | fixed | NA | groupFBVLED:time_pointFinal | -0.10 | 0.30 | -0.34 | 37.41 | -0.70 | 0.50 | 0.738 | 0.896 |
| Bacteroides_B massiliensis | fixed | NA | groupFBVLED:time_pointFinal | -0.08 | 0.25 | -0.33 | 39.10 | -0.59 | 0.42 | 0.74 | 0.897 |
| CAG 245 sp000435175 | fixed | NA | groupFBVLED:time_pointFinal | 0.09 | 0.26 | 0.33 | 43.03 | -0.44 | 0.61 | 0.743 | 0.898 |
| Faecalibacterium prausnitzii_H | fixed | NA | groupFBVLED:time_pointFinal | 0.04 | 0.11 | 0.33 | 35.45 | -0.19 | 0.27 | 0.746 | 0.899 |
| Blautia_A massiliensis | fixed | NA | groupFBVLED:time_pointFinal | 0.13 | 0.41 | 0.33 | 40.29 | -0.70 | 0.96 | 0.747 | 0.899 |
| Gemmiger formicilis | fixed | NA | groupFBVLED:time_pointFinal | -0.08 | 0.26 | -0.32 | 40.85 | -0.60 | 0.44 | 0.754 | 0.906 |
| UBA5026 MIC9780 | fixed | NA | groupFBVLED:time_pointFinal | 0.03 | 0.09 | 0.31 | 41.03 | -0.15 | 0.21 | 0.755 | 0.906 |
| Anaerotignum sp000436415 | fixed | NA | groupFBVLED:time_pointFinal | -0.06 | 0.20 | -0.31 | 40.71 | -0.46 | 0.34 | 0.758 | 0.908 |
| Anaerococcus MIC7735 | fixed | NA | groupFBVLED:time_pointFinal | -0.06 | 0.20 | -0.30 | 84.00 | -0.46 | 0.34 | 0.762 | 0.909 |
| Dialister MIC7247 | fixed | NA | groupFBVLED:time_pointFinal | 0.05 | 0.17 | 0.30 | 84.00 | -0.29 | 0.39 | 0.764 | 0.909 |
| Porphyromonas MIC9772 | fixed | NA | groupFBVLED:time_pointFinal | -0.06 | 0.21 | -0.30 | 84.00 | -0.49 | 0.36 | 0.765 | 0.909 |

| OUTCOMES - SPECIES | EFFECT | GROUP | TERM | ESTIMATE | STD ERROR | STATISTIC | DF | CONF LOW | CONF HIGH | P VALUE | Q VALUE |
|---|---|---|---|---|---|---|---|---|---|---|---|
| Clostridium_M MIC9612 | fixed | NA | groupFBVLED:time_pointFinal | 0.06 | 0.21 | 0.30 | 45.54 | -0.36 | 0.48 | 0.766 | 0.909 |
| Blautia_A MIC6897 | fixed | NA | groupFBVLED:time_pointFinal | 0.05 | 0.17 | 0.30 | 39.90 | -0.29 | 0.39 | 0.767 | 0.909 |
| Collinsella sp002232035 | fixed | NA | groupFBVLED:time_pointFinal | 0.04 | 0.14 | 0.29 | 43.72 | -0.24 | 0.32 | 0.772 | 0.911 |
| CAG 302 sp001916775 | fixed | NA | groupFBVLED:time_pointFinal | 0.08 | 0.26 | 0.29 | 40.76 | -0.46 | 0.61 | 0.772 | 0.911 |
| CAG 74 MIC7629 | fixed | NA | groupFBVLED:time_pointFinal | -0.07 | 0.26 | -0.29 | 45.88 | -0.60 | 0.45 | 0.773 | 0.911 |
| Erysipelatoclostridium spiroforme | fixed | NA | groupFBVLED:time_pointFinal | -0.07 | 0.23 | -0.29 | 40.83 | -0.53 | 0.40 | 0.775 | 0.911 |
| Collinsella aerofaciens_F | fixed | NA | groupFBVLED:time_pointFinal | -0.06 | 0.20 | -0.29 | 38.29 | -0.47 | 0.35 | 0.775 | 0.911 |
| Finegoldia magna_H | fixed | NA | groupFBVLED:time_pointFinal | 0.10 | 0.33 | 0.28 | 40.07 | -0.58 | 0.77 | 0.777 | 0.911 |
| CAG 273 sp000435755 | fixed | NA | groupFBVLED:time_pointFinal | 0.04 | 0.14 | 0.28 | 40.88 | -0.25 | 0.33 | 0.782 | 0.915 |
| Bacteroides eggerthii | fixed | NA | groupFBVLED:time_pointFinal | 0.07 | 0.27 | 0.27 | 38.66 | -0.47 | 0.62 | 0.785 | 0.915 |
| Eubacterium_E MIC8705 | fixed | NA | groupFBVLED:time_pointFinal | -0.05 | 0.17 | -0.27 | 84.00 | -0.39 | 0.29 | 0.788 | 0.915 |
| Streptococcus anginosus | fixed | NA | groupFBVLED:time_pointFinal | 0.08 | 0.29 | 0.27 | 43.34 | -0.51 | 0.67 | 0.788 | 0.915 |
| Methanobrevibacter_A smithii | fixed | NA | groupFBVLED:time_pointFinal | 0.13 | 0.49 | 0.27 | 41.79 | -0.86 | 1.13 | 0.789 | 0.915 |
| Mailhella MIC8103 | fixed | NA | groupFBVLED:time_pointFinal | -0.04 | 0.14 | -0.27 | 43.85 | -0.33 | 0.25 | 0.79 | 0.915 |
| Catenibacterium sp000437715 | fixed | NA | groupFBVLED:time_pointFinal | -0.05 | 0.19 | -0.27 | 41.10 | -0.43 | 0.33 | 0.791 | 0.915 |
| Clostridium_M sp001517625 | fixed | NA | groupFBVLED:time_pointFinal | -0.06 | 0.22 | -0.26 | 35.83 | -0.50 | 0.39 | 0.796 | 0.919 |
| CAG 110 sp000434635 | fixed | NA | groupFBVLED:time_pointFinal | -0.07 | 0.27 | -0.26 | 43.38 | -0.61 | 0.47 | 0.797 | 0.919 |
| CAG 170 MIC8868 | fixed | NA | groupFBVLED:time_pointFinal | -0.05 | 0.20 | -0.25 | 84.00 | -0.44 | 0.34 | 0.805 | 0.925 |
| Parabacteroides distasonis | fixed | NA | groupFBVLED:time_pointFinal | 0.08 | 0.34 | 0.25 | 42.09 | -0.59 | 0.76 | 0.805 | 0.925 |
| Lachnospiraceae MIC7157 | fixed | NA | groupFBVLED:time_pointFinal | -0.03 | 0.13 | -0.24 | 45.47 | -0.29 | 0.23 | 0.81 | 0.927 |
| Lactobacillus crispatus | fixed | NA | groupFBVLED:time_pointFinal | 0.05 | 0.19 | 0.24 | 46.29 | -0.33 | 0.42 | 0.811 | 0.927 |
| Faecalicatena gnavus | fixed | NA | groupFBVLED:time_pointFinal | 0.06 | 0.26 | 0.24 | 37.36 | -0.47 | 0.60 | 0.811 | 0.927 |
| Ruminiclostridium_E siraeum | fixed | NA | groupFBVLED:time_pointFinal | 0.11 | 0.48 | 0.23 | 40.58 | -0.86 | 1.09 | 0.819 | 0.93 |
| Absiella innocuum | fixed | NA | groupFBVLED:time_pointFinal | 0.06 | 0.28 | 0.23 | 84.00 | -0.49 | 0.61 | 0.819 | 0.93 |
| Sellimonas intestinalis | fixed | NA | groupFBVLED:time_pointFinal | 0.04 | 0.18 | 0.23 | 43.95 | -0.33 | 0.41 | 0.82 | 0.93 |
| UBA1685 sp002320595 | fixed | NA | groupFBVLED:time_pointFinal | 0.05 | 0.24 | 0.23 | 42.47 | -0.42 | 0.53 | 0.822 | 0.93 |
| Clostridium_M bolteae | fixed | NA | groupFBVLED:time_pointFinal | 0.06 | 0.26 | 0.23 | 44.37 | -0.47 | 0.58 | 0.822 | 0.93 |
| UBA1191 MIC6632 | fixed | NA | groupFBVLED:time_pointFinal | 0.03 | 0.15 | 0.22 | 43.91 | -0.26 | 0.33 | 0.824 | 0.93 |
| Acutalibacteraceae MIC6974 | fixed | NA | groupFBVLED:time_pointFinal | 0.06 | 0.26 | 0.22 | 41.58 | -0.47 | 0.58 | 0.826 | 0.93 |

| OUTCOMES - SPECIES | EFFECT | GROUP | TERM | ESTIMATE | STD ERROR | STATISTIC | DF | CONF LOW | CONF HIGH | P VALUE | Q VALUE |
|---|---|---|---|---|---|---|---|---|---|---|---|
| CAG 83 MIC8731 | fixed | NA | groupFBVLED:time_pointFinal | 0.04 | 0.20 | 0.22 | 42.83 | -0.36 | 0.45 | 0.827 | 0.93 |
| Anaerococcus obesiensis | fixed | NA | groupFBVLED:time_pointFinal | 0.06 | 0.30 | 0.22 | 84.00 | -0.52 | 0.65 | 0.827 | 0.93 |
| S5 A14a MIC8656 | fixed | NA | groupFBVLED:time_pointFinal | 0.06 | 0.28 | 0.21 | 84.00 | -0.49 | 0.61 | 0.834 | 0.935 |
| UBA10677 MIC8686 | fixed | NA | groupFBVLED:time_pointFinal | 0.02 | 0.07 | 0.21 | 43.80 | -0.14 | 0.17 | 0.837 | 0.936 |
| ER4 sp900317525 | fixed | NA | groupFBVLED:time_pointFinal | 0.06 | 0.32 | 0.20 | 44.02 | -0.57 | 0.70 | 0.839 | 0.936 |
| Parabacteroides merdae | fixed | NA | groupFBVLED:time_pointFinal | 0.04 | 0.21 | 0.20 | 39.73 | -0.39 | 0.47 | 0.841 | 0.936 |
| QALS01 sp003150575 | fixed | NA | groupFBVLED:time_pointFinal | -0.06 | 0.28 | -0.20 | 44.03 | -0.62 | 0.50 | 0.841 | 0.936 |
| Pauljensenia sp000466265 | fixed | NA | groupFBVLED:time_pointFinal | -0.03 | 0.15 | -0.19 | 42.65 | -0.34 | 0.28 | 0.848 | 0.942 |
| Lachnospira sp003451515 | fixed | NA | groupFBVLED:time_pointFinal | -0.04 | 0.22 | -0.19 | 41.72 | -0.48 | 0.40 | 0.851 | 0.944 |
| TF01 11 sp001414325 | fixed | NA | groupFBVLED:time_pointFinal | -0.05 | 0.24 | -0.19 | 39.01 | -0.54 | 0.45 | 0.854 | 0.944 |
| Bacteroides stercoris | fixed | NA | groupFBVLED:time_pointFinal | 0.05 | 0.25 | 0.18 | 39.52 | -0.45 | 0.54 | 0.854 | 0.944 |
| Lawsonibacter MIC8046 | fixed | NA | groupFBVLED:time_pointFinal | -0.03 | 0.17 | -0.18 | 41.86 | -0.37 | 0.30 | 0.857 | 0.946 |
| Dorea longicatena_B | fixed | NA | groupFBVLED:time_pointFinal | 0.04 | 0.23 | 0.17 | 39.88 | -0.43 | 0.51 | 0.863 | 0.951 |
| Bilophila wadsworthia | fixed | NA | groupFBVLED:time_pointFinal | 0.05 | 0.32 | 0.17 | 41.65 | -0.59 | 0.70 | 0.869 | 0.956 |
| Blautia_A MIC8288 | fixed | NA | groupFBVLED:time_pointFinal | -0.02 | 0.12 | -0.16 | 84.00 | -0.25 | 0.21 | 0.873 | 0.958 |
| UBA1191 MIC6696 | fixed | NA | groupFBVLED:time_pointFinal | 0.03 | 0.22 | 0.16 | 42.78 | -0.40 | 0.47 | 0.876 | 0.959 |
| Intestinibacter MIC7152 | fixed | NA | groupFBVLED:time_pointFinal | 0.02 | 0.11 | 0.15 | 84.00 | -0.20 | 0.23 | 0.878 | 0.959 |
| Alistipes_A ihumii | fixed | NA | groupFBVLED:time_pointFinal | -0.05 | 0.31 | -0.15 | 41.91 | -0.67 | 0.57 | 0.879 | 0.959 |
| Phascolarctobacterium sp000436095 | fixed | NA | groupFBVLED:time_pointFinal | 0.02 | 0.12 | 0.15 | 40.75 | -0.22 | 0.26 | 0.88 | 0.959 |
| Eggerthellaceae MIC6427 | fixed | NA | groupFBVLED:time_pointFinal | -0.01 | 0.04 | -0.15 | 40.07 | -0.08 | 0.07 | 0.884 | 0.961 |
| Merdibacter MIC6626 | fixed | NA | groupFBVLED:time_pointFinal | 0.01 | 0.10 | 0.15 | 41.16 | -0.19 | 0.22 | 0.885 | 0.961 |
| CAG 177 sp003514385 | fixed | NA | groupFBVLED:time_pointFinal | 0.05 | 0.35 | 0.13 | 40.67 | -0.65 | 0.74 | 0.895 | 0.969 |
| Akkermansia sp001580195 | fixed | NA | groupFBVLED:time_pointFinal | -0.03 | 0.24 | -0.13 | 43.08 | -0.52 | 0.45 | 0.896 | 0.969 |
| UBA1191 sp900066305 | fixed | NA | groupFBVLED:time_pointFinal | 0.02 | 0.13 | 0.12 | 44.63 | -0.24 | 0.27 | 0.905 | 0.976 |
| Anaerovoracaceae MIC8502 | fixed | NA | groupFBVLED:time_pointFinal | 0.02 | 0.15 | 0.12 | 45.34 | -0.28 | 0.31 | 0.907 | 0.976 |
| Pauljensenia turicensis | fixed | NA | groupFBVLED:time_pointFinal | 0.03 | 0.24 | 0.12 | 45.30 | -0.46 | 0.52 | 0.908 | 0.976 |
| Prevotella disiens | fixed | NA | groupFBVLED:time_pointFinal | 0.02 | 0.17 | 0.12 | 43.43 | -0.31 | 0.35 | 0.909 | 0.976 |
| CAG 74 MIC7845 | fixed | NA | groupFBVLED:time_pointFinal | 0.01 | 0.10 | 0.11 | 84.00 | -0.19 | 0.21 | 0.91 | 0.976 |
| Butyricicoccaceae MIC8222 | fixed | NA | groupFBVLED:time_pointFinal | -0.01 | 0.08 | -0.11 | 43.36 | -0.18 | 0.16 | 0.912 | 0.976 |

| OUTCOMES - SPECIES | EFFECT | GROUP | TERM | ESTIMATE | STD ERROR | STATISTIC | DF | CONF LOW | CONF HIGH | P VALUE | Q VALUE |
|---|---|---|---|---|---|---|---|---|---|---|---|
| CAG 433 sp000433675 | fixed | NA | groupFBVLED:time_pointFinal | -0.03 | 0.27 | -0.11 | 32.54 | -0.58 | 0.52 | 0.914 | 0.976 |
| Lachnospirales MIC8074 | fixed | NA | groupFBVLED:time_pointFinal | -0.01 | 0.11 | -0.10 | 46.21 | -0.24 | 0.22 | 0.918 | 0.976 |
| CAG 74 MIC9091 | fixed | NA | groupFBVLED:time_pointFinal | -0.01 | 0.14 | -0.10 | 43.89 | -0.29 | 0.26 | 0.918 | 0.976 |
| Tyzzerella nexilis | fixed | NA | groupFBVLED:time_pointFinal | -0.04 | 0.39 | -0.10 | 43.06 | -0.83 | 0.75 | 0.919 | 0.976 |
| Bacteroides_A coprocola | fixed | NA | groupFBVLED:time_pointFinal | 0.02 | 0.19 | 0.10 | 40.60 | -0.37 | 0.41 | 0.919 | 0.976 |
| Prevotella corporis | fixed | NA | groupFBVLED:time_pointFinal | -0.04 | 0.37 | -0.10 | 35.03 | -0.79 | 0.72 | 0.922 | 0.976 |
| Bacteroides congonensis | fixed | NA | groupFBVLED:time_pointFinal | -0.02 | 0.17 | -0.10 | 45.23 | -0.36 | 0.33 | 0.922 | 0.976 |
| Alistipes MIC9770 | fixed | NA | groupFBVLED:time_pointFinal | 0.02 | 0.16 | 0.10 | 42.54 | -0.31 | 0.34 | 0.924 | 0.976 |
| Coprobacillus cateniformis | fixed | NA | groupFBVLED:time_pointFinal | -0.03 | 0.32 | -0.09 | 44.06 | -0.67 | 0.61 | 0.926 | 0.976 |
| Blastocystis sp subtype 3 | fixed | NA | groupFBVLED:time_pointFinal | 0.01 | 0.07 | 0.09 | 40.96 | -0.14 | 0.15 | 0.927 | 0.976 |
| CAG 170 sp002404795 | fixed | NA | groupFBVLED:time_pointFinal | 0.04 | 0.43 | 0.09 | 44.91 | -0.83 | 0.91 | 0.929 | 0.976 |
| UBA5446 MIC9241 | fixed | NA | groupFBVLED:time_pointFinal | 0.02 | 0.20 | 0.08 | 44.24 | -0.39 | 0.43 | 0.934 | 0.98 |
| Bacteroides MIC8726 | fixed | NA | groupFBVLED:time_pointFinal | 0.01 | 0.18 | 0.08 | 43.39 | -0.35 | 0.38 | 0.94 | 0.981 |
| Faecalicatena sp900066545 | fixed | NA | groupFBVLED:time_pointFinal | -0.01 | 0.15 | -0.07 | 42.70 | -0.32 | 0.30 | 0.941 | 0.981 |
| Absiella sp000165065 | fixed | NA | groupFBVLED:time_pointFinal | -0.02 | 0.20 | -0.07 | 44.47 | -0.43 | 0.40 | 0.941 | 0.981 |
| Dorea sp900066765 | fixed | NA | groupFBVLED:time_pointFinal | -0.01 | 0.12 | -0.07 | 40.89 | -0.26 | 0.24 | 0.942 | 0.981 |
| Corynebacterium sp001767255 | fixed | NA | groupFBVLED:time_pointFinal | 0.01 | 0.08 | 0.07 | 84.00 | -0.15 | 0.16 | 0.942 | 0.981 |
| Ruminococcaceae MIC8509 | fixed | NA | groupFBVLED:time_pointFinal | 0.01 | 0.08 | 0.07 | 45.17 | -0.16 | 0.17 | 0.948 | 0.982 |
| Coprococcus_A MIC9199 | fixed | NA | groupFBVLED:time_pointFinal | 0.01 | 0.21 | 0.06 | 40.48 | -0.42 | 0.44 | 0.95 | 0.982 |
| Blautia_A sp000433815 | fixed | NA | groupFBVLED:time_pointFinal | -0.02 | 0.29 | -0.06 | 42.78 | -0.60 | 0.57 | 0.95 | 0.982 |
| Blautia_A sp900066205 | fixed | NA | groupFBVLED:time_pointFinal | 0.02 | 0.30 | 0.06 | 41.48 | -0.59 | 0.62 | 0.953 | 0.982 |
| CAG 495 sp000436375 | fixed | NA | groupFBVLED:time_pointFinal | -0.02 | 0.41 | -0.06 | 40.86 | -0.86 | 0.81 | 0.955 | 0.982 |
| QALS01 MIC6548 | fixed | NA | groupFBVLED:time_pointFinal | -0.01 | 0.21 | -0.05 | 36.39 | -0.43 | 0.41 | 0.957 | 0.982 |
| Holdemanella sp002299315 | fixed | NA | groupFBVLED:time_pointFinal | -0.01 | 0.17 | -0.05 | 39.39 | -0.36 | 0.34 | 0.959 | 0.982 |
| Lachnospira sp000436475 | fixed | NA | groupFBVLED:time_pointFinal | -0.01 | 0.16 | -0.05 | 41.69 | -0.32 | 0.31 | 0.96 | 0.982 |
| Blautia_A sp900120195 | fixed | NA | groupFBVLED:time_pointFinal | 0.01 | 0.23 | 0.05 | 42.91 | -0.46 | 0.49 | 0.961 | 0.982 |
| TF01 11 sp003524945 | fixed | NA | groupFBVLED:time_pointFinal | -0.01 | 0.17 | -0.05 | 43.01 | -0.35 | 0.33 | 0.961 | 0.982 |
| Porphyromonas MIC7959 | fixed | NA | groupFBVLED:time_pointFinal | -0.01 | 0.25 | -0.05 | 84.00 | -0.51 | 0.49 | 0.964 | 0.982 |
| Eubacterium_E hallii_A | fixed | NA | groupFBVLED:time_pointFinal | 0.01 | 0.26 | 0.04 | 40.69 | -0.51 | 0.53 | 0.965 | 0.982 |

| OUTCOMES - SPECIES | EFFECT | GROUP | TERM | ESTIMATE | STD ERROR | STATISTIC | DF | CONF LOW | CONF HIGH | P VALUE | Q VALUE |
|---|---|---|---|---|---|---|---|---|---|---|---|
| CAG 127 sp900319515 | fixed | NA | groupFBVLED:time_pointFinal | -0.02 | 0.36 | -0.04 | 43.52 | -0.75 | 0.72 | 0.965 | 0.982 |
| Faecalibacterium prausnitzii_E | fixed | NA | groupFBVLED:time_pointFinal | 0.01 | 0.14 | 0.04 | 40.58 | -0.28 | 0.29 | 0.966 | 0.982 |
| Oscillospiraceae MIC7451 | fixed | NA | groupFBVLED:time_pointFinal | 0.00 | 0.04 | -0.04 | 41.50 | -0.09 | 0.08 | 0.969 | 0.983 |
| Alistipes senegalensis | fixed | NA | groupFBVLED:time_pointFinal | 0.00 | 0.12 | 0.03 | 41.38 | -0.25 | 0.26 | 0.973 | 0.986 |
| CAG 170 sp000432135 | fixed | NA | groupFBVLED:time_pointFinal | 0.01 | 0.43 | 0.03 | 44.75 | -0.86 | 0.88 | 0.978 | 0.99 |
| UBA1691 MIC9213 | fixed | NA | groupFBVLED:time_pointFinal | 0.00 | 0.17 | -0.02 | 44.85 | -0.35 | 0.34 | 0.987 | 0.996 |
| Lactobacillus iners | fixed | NA | groupFBVLED:time_pointFinal | 0.00 | 0.15 | 0.02 | 44.79 | -0.30 | 0.30 | 0.988 | 0.996 |
| Blautia_A sp000436615 | fixed | NA | groupFBVLED:time_pointFinal | -0.01 | 0.47 | -0.01 | 42.05 | -0.95 | 0.94 | 0.99 | 0.996 |
| GCA 900066135 sp900066135 | fixed | NA | groupFBVLED:time_pointFinal | 0.00 | 0.17 | 0.01 | 43.38 | -0.33 | 0.34 | 0.993 | 0.996 |
| Faecalicatena sp900120155 | fixed | NA | groupFBVLED:time_pointFinal | 0.00 | 0.07 | 0.01 | 84.00 | -0.13 | 0.13 | 0.993 | 0.996 |
| Clostridium saudiense | fixed | NA | groupFBVLED:time_pointFinal | 0.00 | 0.38 | 0.01 | 41.28 | -0.77 | 0.77 | 0.996 | 0.997 |
| UBA1777 sp003150355 | fixed | NA | groupFBVLED:time_pointFinal | 0.00 | 0.10 | 0.00 | 41.29 | -0.21 | 0.21 | 0.997 | 0.997 |

Unadjusted model: ~ group*timepoint.

Adjusted q-value: The p-value controlling for the false discovery rate using the Benjamini-Hochberg procedure.

Abbreviations: β, Beta-coefficient; CI, Confidence interval.
