## Supplementary material for "The effects of food-based versus supplement-based very low-energy diets on gut microbiome composition and health outcomes in women with high body mass index (The MicroFit Study): a randomised controlled trial": Table S6

**Table S6.** Unadjusted complete case analysis (n=39) of the differential changes in bacterial species between those that consumed a food-based versus supplement-based very low-energy diet for three weeks.

| OUTCOMES - SPECIES | EFFECT | GROUP | TERM | ESTIMATE | STD ERROR | STATISTIC | DF | CONF LOW | CONF HIGH | P VALUE | Q VALUE |
|---|---|---|---|---|---|---|---|---|---|---|---|
| Oscillibacter sp001916835 | fixed | NA | groupFBVLED:time_pointFinal | 0.71 | 0.19 | 3.65 | 39.00 | 0.32 | 1.10 | **0.001** | 0.279 |
| Agathobacter faecis | fixed | NA | groupFBVLED:time_pointFinal | 1.82 | 0.51 | 3.59 | 39.00 | 0.79 | 2.84 | **0.001** | 0.279 |
| Faecalibacterium prausnitzii_G | fixed | NA | groupFBVLED:time_pointFinal | 1.27 | 0.38 | 3.38 | 39.00 | 0.51 | 2.03 | **0.002** | 0.279 |
| Gemmiger sp003476825 | fixed | NA | groupFBVLED:time_pointFinal | 1.23 | 0.38 | 3.24 | 39.00 | 0.46 | 1.99 | **0.002** | 0.279 |
| Anaerostipes hadrus | fixed | NA | groupFBVLED:time_pointFinal | -1.20 | 0.38 | -3.13 | 39.00 | -1.97 | -0.42 | **0.003** | 0.279 |
| Faecalibacterium prausnitzii_D | fixed | NA | groupFBVLED:time_pointFinal | 1.17 | 0.38 | 3.09 | 39.00 | 0.40 | 1.94 | **0.004** | 0.279 |
| Lawsonibacter sp900066645 | fixed | NA | groupFBVLED:time_pointFinal | -0.37 | 0.12 | -3.04 | 39.00 | -0.61 | -0.12 | **0.004** | 0.279 |
| GCA 900066135 MIC6659 | fixed | NA | groupFBVLED:time_pointFinal | 0.72 | 0.24 | 3.02 | 39.00 | 0.24 | 1.21 | **0.004** | 0.279 |
| Alistipes MIC8513 | fixed | NA | groupFBVLED:time_pointFinal | 0.37 | 0.13 | 2.95 | 39.00 | 0.12 | 0.63 | **0.005** | 0.279 |
| Dorea sp000433215 | fixed | NA | groupFBVLED:time_pointFinal | 0.57 | 0.19 | 2.92 | 39.00 | 0.17 | 0.96 | **0.006** | 0.279 |
| GCA 900066905 sp900066905 | fixed | NA | groupFBVLED:time_pointFinal | -0.37 | 0.13 | -2.83 | 78.00 | -0.62 | -0.11 | **0.006** | 0.279 |
| UBA11524 sp000437595 | fixed | NA | groupFBVLED:time_pointFinal | 1.13 | 0.39 | 2.92 | 39.00 | 0.34 | 1.91 | **0.006** | 0.279 |
| Ruminococcus_A sp003011855 | fixed | NA | groupFBVLED:time_pointFinal | -0.89 | 0.31 | -2.91 | 39.00 | -1.51 | -0.27 | **0.006** | 0.279 |
| Lachnospiraceae MIC8879 | fixed | NA | groupFBVLED:time_pointFinal | 0.54 | 0.19 | 2.87 | 39.00 | 0.16 | 0.92 | **0.007** | 0.289 |
| UBA11774 sp003507655 | fixed | NA | groupFBVLED:time_pointFinal | 1.25 | 0.44 | 2.84 | 39.00 | 0.36 | 2.15 | **0.007** | 0.296 |
| Streptococcus thermophilus | fixed | NA | groupFBVLED:time_pointFinal | 1.60 | 0.57 | 2.80 | 39.00 | 0.44 | 2.76 | **0.008** | 0.302 |
| ER4 sp000765235 | fixed | NA | groupFBVLED:time_pointFinal | 0.82 | 0.30 | 2.74 | 39.00 | 0.22 | 1.43 | **0.009** | 0.302 |
| Clostridium_A leptum | fixed | NA | groupFBVLED:time_pointFinal | -0.82 | 0.30 | -2.73 | 39.00 | -1.44 | -0.21 | **0.009** | 0.302 |
| Lachnospiraceae MIC9331 | fixed | NA | groupFBVLED:time_pointFinal | -0.64 | 0.24 | -2.72 | 39.00 | -1.12 | -0.16 | **0.010** | 0.302 |
| Bacteroides finegoldii | fixed | NA | groupFBVLED:time_pointFinal | 0.60 | 0.22 | 2.70 | 39.00 | 0.15 | 1.05 | **0.010** | 0.302 |
| Lachnospira rogosae | fixed | NA | groupFBVLED:time_pointFinal | 1.13 | 0.42 | 2.70 | 39.00 | 0.28 | 1.99 | **0.010** | 0.302 |
| Eisenbergiella sp900066775 | fixed | NA | groupFBVLED:time_pointFinal | 0.72 | 0.27 | 2.66 | 39.00 | 0.17 | 1.27 | **0.011** | 0.304 |
| Bacteroides faecis | fixed | NA | groupFBVLED:time_pointFinal | -0.55 | 0.20 | -2.66 | 39.00 | -0.96 | -0.13 | **0.011** | 0.304 |
| Eisenbergiella massiliensis | fixed | NA | groupFBVLED:time_pointFinal | -0.55 | 0.22 | -2.50 | 78.00 | -0.98 | -0.11 | **0.014** | 0.367 |
| Oscillibacter MIC9597 | fixed | NA | groupFBVLED:time_pointFinal | 0.50 | 0.20 | 2.54 | 39.00 | 0.10 | 0.89 | **0.015** | 0.367 |
| Bifidobacterium longum | fixed | NA | groupFBVLED:time_pointFinal | -1.18 | 0.46 | -2.53 | 39.00 | -2.12 | -0.24 | **0.015** | 0.367 |
| CAG 74 MIC8062 | fixed | NA | groupFBVLED:time_pointFinal | -1.15 | 0.46 | -2.52 | 39.00 | -2.07 | -0.23 | **0.016** | 0.367 |

| OUTCOMES - SPECIES | EFFECT | GROUP | TERM | ESTIMATE | STD ERROR | STATISTIC | DF | CONF LOW | CONF HIGH | P VALUE | Q VALUE |
|---|---|---|---|---|---|---|---|---|---|---|---|
| Eubacterium_F sp000433735 | fixed | NA | groupFBVLED:time_pointFinal | 0.42 | 0.17 | 2.49 | 39.00 | 0.08 | 0.75 | **0.017** | 0.367 |
| Ruthenibacterium lactatiformans | fixed | NA | groupFBVLED:time_pointFinal | -1.19 | 0.48 | -2.49 | 39.00 | -2.16 | -0.22 | **0.017** | 0.367 |
| Faecalicatena lactaris | fixed | NA | groupFBVLED:time_pointFinal | 0.84 | 0.34 | 2.46 | 39.00 | 0.15 | 1.53 | **0.018** | 0.377 |
| Terrisporobacter MIC9205 | fixed | NA | groupFBVLED:time_pointFinal | 0.76 | 0.31 | 2.45 | 39.00 | 0.13 | 1.39 | **0.019** | 0.377 |
| UBA1417 sp003531055 | fixed | NA | groupFBVLED:time_pointFinal | -1.02 | 0.42 | -2.41 | 39.00 | -1.88 | -0.16 | **0.021** | 0.381 |
| Agathobaculum butyriciproducens | fixed | NA | groupFBVLED:time_pointFinal | 0.78 | 0.33 | 2.40 | 39.00 | 0.12 | 1.44 | **0.021** | 0.381 |
| UBA9502 MIC6887 | fixed | NA | groupFBVLED:time_pointFinal | 0.32 | 0.14 | 2.37 | 39.00 | 0.05 | 0.60 | **0.023** | 0.381 |
| Eubacterium_R sp000434995 | fixed | NA | groupFBVLED:time_pointFinal | 0.50 | 0.21 | 2.35 | 39.00 | 0.07 | 0.93 | **0.024** | 0.381 |
| Ruminococcus_D bicirculans | fixed | NA | groupFBVLED:time_pointFinal | 1.24 | 0.53 | 2.35 | 39.00 | 0.17 | 2.31 | **0.024** | 0.381 |
| GCA 900066995 sp900291955 | fixed | NA | groupFBVLED:time_pointFinal | 0.65 | 0.28 | 2.33 | 39.00 | 0.09 | 1.22 | **0.025** | 0.381 |
| Lachnospiraceae MIC6495 | fixed | NA | groupFBVLED:time_pointFinal | 0.42 | 0.18 | 2.32 | 39.00 | 0.05 | 0.78 | **0.026** | 0.381 |
| UBA1820 sp002314265 | fixed | NA | groupFBVLED:time_pointFinal | 0.45 | 0.20 | 2.32 | 39.00 | 0.06 | 0.85 | **0.026** | 0.381 |
| Lachnospira sp900316325 | fixed | NA | groupFBVLED:time_pointFinal | 0.87 | 0.37 | 2.31 | 39.00 | 0.11 | 1.62 | **0.026** | 0.381 |
| Blautia_A MIC7250 | fixed | NA | groupFBVLED:time_pointFinal | 0.22 | 0.10 | 2.27 | 77.94 | 0.03 | 0.42 | **0.026** | 0.381 |
| UBA1191 MIC6579 | fixed | NA | groupFBVLED:time_pointFinal | -0.40 | 0.17 | -2.31 | 39.00 | -0.75 | -0.05 | **0.026** | 0.381 |
| Faecalibacterium MIC8666 | fixed | NA | groupFBVLED:time_pointFinal | 0.94 | 0.41 | 2.30 | 39.00 | 0.11 | 1.77 | **0.027** | 0.381 |
| Eggerthella lenta | fixed | NA | groupFBVLED:time_pointFinal | -0.70 | 0.31 | -2.29 | 39.00 | -1.32 | -0.08 | **0.028** | 0.381 |
| Intestinimonas butyriciproducens | fixed | NA | groupFBVLED:time_pointFinal | -0.55 | 0.24 | -2.29 | 39.00 | -1.04 | -0.06 | **0.028** | 0.381 |
| Hungatella effluvii | fixed | NA | groupFBVLED:time_pointFinal | -0.40 | 0.18 | -2.23 | 78.00 | -0.76 | -0.04 | **0.028** | 0.382 |
| Peptoniphilus_A harei_A | fixed | NA | groupFBVLED:time_pointFinal | 0.34 | 0.15 | 2.24 | 39.00 | 0.03 | 0.65 | **0.031** | 0.410 |
| CAG 269 sp003525075 | fixed | NA | groupFBVLED:time_pointFinal | -0.47 | 0.21 | -2.20 | 39.00 | -0.90 | -0.04 | **0.033** | 0.410 |
| Faecalibacterium MIC7145 | fixed | NA | groupFBVLED:time_pointFinal | 0.80 | 0.36 | 2.20 | 39.00 | 0.07 | 1.54 | **0.034** | 0.410 |
| Dialister invisus | fixed | NA | groupFBVLED:time_pointFinal | 0.43 | 0.20 | 2.20 | 39.00 | 0.04 | 0.83 | **0.034** | 0.410 |
| Parabacteroides goldsteinii | fixed | NA | groupFBVLED:time_pointFinal | -0.75 | 0.34 | -2.20 | 39.00 | -1.44 | -0.06 | **0.034** | 0.410 |
| Coprococcus sp000154245 | fixed | NA | groupFBVLED:time_pointFinal | 0.75 | 0.34 | 2.19 | 39.00 | 0.06 | 1.44 | **0.035** | 0.410 |
| Blautia_A sp900066145 | fixed | NA | groupFBVLED:time_pointFinal | 0.72 | 0.33 | 2.18 | 39.00 | 0.05 | 1.39 | **0.035** | 0.410 |
| Lachnospira eligens_B | fixed | NA | groupFBVLED:time_pointFinal | 1.14 | 0.53 | 2.16 | 39.00 | 0.07 | 2.20 | **0.037** | 0.410 |
| QAND01 sp003150225 | fixed | NA | groupFBVLED:time_pointFinal | -0.24 | 0.11 | -2.13 | 78.00 | -0.47 | -0.02 | **0.037** | 0.410 |
| Blautia_A MIC8343 | fixed | NA | groupFBVLED:time_pointFinal | 0.32 | 0.15 | 2.14 | 39.00 | 0.02 | 0.63 | **0.038** | 0.410 |

| OUTCOMES - SPECIES | EFFECT | GROUP | TERM | ESTIMATE | STD ERROR | STATISTIC | DF | CONF LOW | CONF HIGH | P VALUE | Q VALUE |
|---|---|---|---|---|---|---|---|---|---|---|---|
| Absiella sp000163515 | fixed | NA | groupFBVLED:time_pointFinal | -0.31 | 0.15 | -2.13 | 39.00 | -0.61 | -0.02 | **0.040** | 0.410 |
| Gemmiger MIC9530 | fixed | NA | groupFBVLED:time_pointFinal | 0.58 | 0.28 | 2.12 | 39.00 | 0.03 | 1.14 | **0.040** | 0.410 |
| COE1 sp001916965 | fixed | NA | groupFBVLED:time_pointFinal | 0.45 | 0.21 | 2.11 | 39.00 | 0.02 | 0.88 | **0.042** | 0.410 |
| Clostridium MIC8163 | fixed | NA | groupFBVLED:time_pointFinal | 0.86 | 0.41 | 2.10 | 39.00 | 0.03 | 1.70 | **0.042** | 0.410 |
| 4C28d 15 MIC7065 | fixed | NA | groupFBVLED:time_pointFinal | 0.18 | 0.09 | 2.10 | 39.00 | 0.01 | 0.36 | **0.042** | 0.410 |
| Lachnospiraceae MIC7543 | fixed | NA | groupFBVLED:time_pointFinal | -0.21 | 0.10 | -2.10 | 39.00 | -0.42 | -0.01 | **0.042** | 0.410 |
| Clostridium_M MIC6986 | fixed | NA | groupFBVLED:time_pointFinal | 0.31 | 0.15 | 2.09 | 39.00 | 0.01 | 0.61 | **0.043** | 0.410 |
| Eisenbergiella tayi | fixed | NA | groupFBVLED:time_pointFinal | -0.83 | 0.40 | -2.09 | 39.00 | -1.64 | -0.03 | **0.044** | 0.410 |
| Negativibacillus sp000435195 | fixed | NA | groupFBVLED:time_pointFinal | 0.46 | 0.22 | 2.06 | 39.00 | 0.01 | 0.90 | 0.046 | 0.410 |
| Erysipelatoclostridium ramosum | fixed | NA | groupFBVLED:time_pointFinal | -0.74 | 0.36 | -2.05 | 39.00 | -1.47 | -0.01 | 0.047 | 0.410 |
| Faecalibacterium prausnitzii_I | fixed | NA | groupFBVLED:time_pointFinal | 0.63 | 0.31 | 2.05 | 39.00 | 0.01 | 1.25 | 0.047 | 0.410 |
| An200 sp003268275 | fixed | NA | groupFBVLED:time_pointFinal | -0.22 | 0.11 | -2.05 | 39.00 | -0.43 | 0.00 | 0.047 | 0.410 |
| Lactobacillus_C rhamnosus | fixed | NA | groupFBVLED:time_pointFinal | 0.38 | 0.19 | 2.01 | 78.00 | 0.00 | 0.76 | 0.048 | 0.410 |
| CAG 74 MIC7649 | fixed | NA | groupFBVLED:time_pointFinal | -0.52 | 0.26 | -2.04 | 39.00 | -1.04 | 0.00 | 0.048 | 0.410 |
| Coprococcus_B comes | fixed | NA | groupFBVLED:time_pointFinal | 0.52 | 0.26 | 2.03 | 39.00 | 0.00 | 1.04 | 0.049 | 0.410 |
| Faecalicatena glycyrrhizinilyticum | fixed | NA | groupFBVLED:time_pointFinal | 0.29 | 0.14 | 2.02 | 39.00 | 0.00 | 0.57 | 0.050 | 0.410 |
| UBA9475 MIC7490 | fixed | NA | groupFBVLED:time_pointFinal | 0.19 | 0.09 | 2.02 | 39.00 | 0.00 | 0.38 | 0.051 | 0.410 |
| CAG 103 sp000432375 | fixed | NA | groupFBVLED:time_pointFinal | 0.66 | 0.33 | 2.01 | 39.00 | 0.00 | 1.32 | 0.051 | 0.410 |
| CAG 81 sp900066535 | fixed | NA | groupFBVLED:time_pointFinal | 0.40 | 0.20 | 2.01 | 39.00 | 0.00 | 0.81 | 0.052 | 0.410 |
| Gemmiger MIC8010 | fixed | NA | groupFBVLED:time_pointFinal | -0.54 | 0.27 | -2.01 | 39.00 | -1.09 | 0.00 | 0.052 | 0.410 |
| Lachnospiraceae MIC8643 | fixed | NA | groupFBVLED:time_pointFinal | -0.25 | 0.13 | -2.01 | 39.00 | -0.51 | 0.00 | 0.052 | 0.410 |
| Clostridium sp001916075 | fixed | NA | groupFBVLED:time_pointFinal | 0.79 | 0.40 | 2.01 | 39.00 | -0.01 | 1.59 | 0.052 | 0.410 |
| UBA7102 MIC9705 | fixed | NA | groupFBVLED:time_pointFinal | -0.42 | 0.21 | -2.00 | 39.00 | -0.84 | 0.01 | 0.053 | 0.410 |
| Massilimaliae timonensis | fixed | NA | groupFBVLED:time_pointFinal | -0.64 | 0.32 | -1.96 | 77.99 | -1.28 | 0.01 | 0.054 | 0.410 |
| CAG 313 sp003539625 | fixed | NA | groupFBVLED:time_pointFinal | 0.78 | 0.39 | 1.99 | 39.00 | -0.01 | 1.58 | 0.054 | 0.410 |
| Oxalobacter MIC6654 | fixed | NA | groupFBVLED:time_pointFinal | 0.28 | 0.14 | 1.98 | 39.00 | -0.01 | 0.57 | 0.055 | 0.411 |
| Lachnospira sp000437735 | fixed | NA | groupFBVLED:time_pointFinal | 0.75 | 0.38 | 1.97 | 39.00 | -0.02 | 1.51 | 0.055 | 0.411 |
| Oscillibacter MIC9243 | fixed | NA | groupFBVLED:time_pointFinal | -0.14 | 0.07 | -1.97 | 39.00 | -0.27 | 0.00 | 0.056 | 0.411 |
| Escherichia flexneri | fixed | NA | groupFBVLED:time_pointFinal | -0.82 | 0.42 | -1.94 | 78.00 | -1.66 | 0.02 | 0.057 | 0.411 |

| OUTCOMES - SPECIES | EFFECT | GROUP | TERM | ESTIMATE | STD ERROR | STATISTIC | DF | CONF LOW | CONF HIGH | P VALUE | Q VALUE |
|---|---|---|---|---|---|---|---|---|---|---|---|
| CAG 81 sp900066055 | fixed | NA | groupFBVLED:time_pointFinal | 0.26 | 0.13 | 1.96 | 39.00 | -0.01 | 0.52 | 0.057 | 0.411 |
| Clostridium_Q sp003024715 | fixed | NA | groupFBVLED:time_pointFinal | 0.47 | 0.24 | 1.95 | 39.00 | -0.02 | 0.95 | 0.058 | 0.412 |
| Lachnospirales MIC7715 | fixed | NA | groupFBVLED:time_pointFinal | -0.25 | 0.13 | -1.94 | 39.00 | -0.51 | 0.01 | 0.059 | 0.412 |
| Cloacibacillus evryensis | fixed | NA | groupFBVLED:time_pointFinal | -0.27 | 0.14 | -1.94 | 39.00 | -0.55 | 0.01 | 0.059 | 0.412 |
| CAG 81 sp000435795 | fixed | NA | groupFBVLED:time_pointFinal | 0.42 | 0.22 | 1.93 | 39.00 | -0.02 | 0.86 | 0.060 | 0.413 |
| Bifidobacterium animalis | fixed | NA | groupFBVLED:time_pointFinal | 0.47 | 0.24 | 1.93 | 39.00 | -0.02 | 0.97 | 0.061 | 0.413 |
| CAG 1427 sp000435475 | fixed | NA | groupFBVLED:time_pointFinal | -0.34 | 0.18 | -1.93 | 39.00 | -0.69 | 0.02 | 0.061 | 0.413 |
| Faecalitalea cylindroides | fixed | NA | groupFBVLED:time_pointFinal | -0.48 | 0.25 | -1.92 | 39.00 | -0.98 | 0.03 | 0.062 | 0.414 |
| Lachnospiraceae MIC9183 | fixed | NA | groupFBVLED:time_pointFinal | -0.31 | 0.16 | -1.91 | 39.00 | -0.64 | 0.02 | 0.063 | 0.414 |
| Roseburia inulinivorans | fixed | NA | groupFBVLED:time_pointFinal | 0.62 | 0.33 | 1.89 | 39.00 | -0.04 | 1.28 | 0.066 | 0.425 |
| Odoribacter splanchnicus | fixed | NA | groupFBVLED:time_pointFinal | 0.50 | 0.27 | 1.88 | 39.00 | -0.04 | 1.05 | 0.067 | 0.425 |
| CAG 95 sp900066375 | fixed | NA | groupFBVLED:time_pointFinal | 0.60 | 0.32 | 1.85 | 77.98 | -0.04 | 1.24 | 0.068 | 0.425 |
| Coprococcus_A catus | fixed | NA | groupFBVLED:time_pointFinal | 0.50 | 0.27 | 1.87 | 39.00 | -0.04 | 1.04 | 0.069 | 0.425 |
| Turicibacter sp001543345 | fixed | NA | groupFBVLED:time_pointFinal | 0.35 | 0.19 | 1.87 | 39.00 | -0.03 | 0.73 | 0.069 | 0.425 |
| Streptococcus parasanguinis | fixed | NA | groupFBVLED:time_pointFinal | 0.45 | 0.24 | 1.87 | 39.00 | -0.04 | 0.94 | 0.069 | 0.425 |
| Oscillibacter MIC7603 | fixed | NA | groupFBVLED:time_pointFinal | -0.18 | 0.10 | -1.84 | 78.00 | -0.38 | 0.01 | 0.069 | 0.425 |
| Flavonifractor plautii | fixed | NA | groupFBVLED:time_pointFinal | -0.55 | 0.29 | -1.86 | 39.00 | -1.14 | 0.05 | 0.071 | 0.430 |
| CAG 475 sp000434435 | fixed | NA | groupFBVLED:time_pointFinal | 0.48 | 0.26 | 1.85 | 39.00 | -0.05 | 1.01 | 0.072 | 0.435 |
| Ruminococcus MIC6917 | fixed | NA | groupFBVLED:time_pointFinal | 0.41 | 0.22 | 1.83 | 39.00 | -0.04 | 0.85 | 0.074 | 0.435 |
| UBA1820 sp003150615 | fixed | NA | groupFBVLED:time_pointFinal | -0.28 | 0.15 | -1.83 | 39.00 | -0.58 | 0.03 | 0.075 | 0.435 |
| Eubacterium_F sp003491505 | fixed | NA | groupFBVLED:time_pointFinal | 0.48 | 0.26 | 1.83 | 39.00 | -0.05 | 1.00 | 0.075 | 0.435 |
| Ruthenibacterium MIC9855 | fixed | NA | groupFBVLED:time_pointFinal | -0.28 | 0.15 | -1.82 | 39.00 | -0.60 | 0.03 | 0.076 | 0.435 |
| CAG 74 MIC9837 | fixed | NA | groupFBVLED:time_pointFinal | -0.51 | 0.28 | -1.82 | 39.00 | -1.08 | 0.06 | 0.076 | 0.435 |
| Anaerococcus prevotii_A | fixed | NA | groupFBVLED:time_pointFinal | 0.31 | 0.17 | 1.79 | 78.00 | -0.03 | 0.66 | 0.077 | 0.436 |
| CAG 452 sp000434035 | fixed | NA | groupFBVLED:time_pointFinal | -0.21 | 0.12 | -1.81 | 39.00 | -0.44 | 0.02 | 0.078 | 0.439 |
| Desulfovibrio fairfieldensis | fixed | NA | groupFBVLED:time_pointFinal | -0.32 | 0.18 | -1.80 | 39.00 | -0.67 | 0.04 | 0.079 | 0.442 |
| Victivallis sp002998355 | fixed | NA | groupFBVLED:time_pointFinal | 0.34 | 0.19 | 1.79 | 39.00 | -0.04 | 0.72 | 0.081 | 0.449 |
| Clostridium_Q sp000435655 | fixed | NA | groupFBVLED:time_pointFinal | 0.20 | 0.11 | 1.78 | 39.00 | -0.03 | 0.43 | 0.084 | 0.457 |
| Butyricimonas sp002161485 | fixed | NA | groupFBVLED:time_pointFinal | 0.54 | 0.30 | 1.76 | 39.00 | -0.08 | 1.15 | 0.086 | 0.463 |

| OUTCOMES - SPECIES | EFFECT | GROUP | TERM | ESTIMATE | STD ERROR | STATISTIC | DF | CONF LOW | CONF HIGH | P VALUE | Q VALUE |
|---|---|---|---|---|---|---|---|---|---|---|---|
| QANA01 sp003149735 | fixed | NA | groupFBVLED:time_pointFinal | -0.16 | 0.09 | -1.75 | 39.00 | -0.34 | 0.02 | 0.087 | 0.463 |
| QANA01 MIC6812 | fixed | NA | groupFBVLED:time_pointFinal | 0.18 | 0.10 | 1.75 | 39.00 | -0.03 | 0.38 | 0.087 | 0.463 |
| Faecalibacterium MIC8693 | fixed | NA | groupFBVLED:time_pointFinal | 0.21 | 0.12 | 1.72 | 78.00 | -0.03 | 0.44 | 0.089 | 0.463 |
| CAG 217 sp000436335 | fixed | NA | groupFBVLED:time_pointFinal | 0.32 | 0.19 | 1.74 | 39.00 | -0.05 | 0.70 | 0.089 | 0.463 |
| Alistipes putredinis | fixed | NA | groupFBVLED:time_pointFinal | 0.41 | 0.23 | 1.74 | 39.00 | -0.07 | 0.88 | 0.090 | 0.463 |
| Bacteroides salyersiae | fixed | NA | groupFBVLED:time_pointFinal | 0.32 | 0.19 | 1.72 | 39.00 | -0.06 | 0.70 | 0.093 | 0.463 |
| Parasutterella excrementihominis | fixed | NA | groupFBVLED:time_pointFinal | 0.38 | 0.22 | 1.72 | 39.00 | -0.07 | 0.84 | 0.093 | 0.463 |
| CAG 45 sp900066395 | fixed | NA | groupFBVLED:time_pointFinal | 0.60 | 0.35 | 1.72 | 39.00 | -0.11 | 1.32 | 0.094 | 0.463 |
| Acutalibacter timonensis | fixed | NA | groupFBVLED:time_pointFinal | -0.62 | 0.36 | -1.72 | 39.00 | -1.35 | 0.11 | 0.094 | 0.463 |
| Lachnospira sp000436535 | fixed | NA | groupFBVLED:time_pointFinal | 0.30 | 0.17 | 1.71 | 39.00 | -0.05 | 0.65 | 0.095 | 0.463 |
| Alistipes finegoldii | fixed | NA | groupFBVLED:time_pointFinal | 0.44 | 0.26 | 1.71 | 39.00 | -0.08 | 0.97 | 0.095 | 0.463 |
| Peptoniphilus_C sp900088125 | fixed | NA | groupFBVLED:time_pointFinal | 0.30 | 0.18 | 1.68 | 78.00 | -0.06 | 0.66 | 0.097 | 0.463 |
| Ruminiclostridium_C MIC9168 | fixed | NA | groupFBVLED:time_pointFinal | -0.07 | 0.04 | -1.69 | 39.00 | -0.15 | 0.01 | 0.099 | 0.463 |
| CAG 492 sp000434335 | fixed | NA | groupFBVLED:time_pointFinal | 0.39 | 0.23 | 1.69 | 39.00 | -0.08 | 0.85 | 0.099 | 0.463 |
| Alistipes shahii | fixed | NA | groupFBVLED:time_pointFinal | -0.60 | 0.36 | -1.69 | 39.00 | -1.32 | 0.12 | 0.099 | 0.463 |
| CAG 110 sp000435995 | fixed | NA | groupFBVLED:time_pointFinal | -0.53 | 0.32 | -1.69 | 39.00 | -1.17 | 0.11 | 0.100 | 0.463 |
| CAG 1427 sp000435675 | fixed | NA | groupFBVLED:time_pointFinal | 0.47 | 0.28 | 1.69 | 39.00 | -0.09 | 1.04 | 0.100 | 0.463 |
| Oscillospiraceae MIC8045 | fixed | NA | groupFBVLED:time_pointFinal | 0.17 | 0.10 | 1.68 | 39.00 | -0.03 | 0.37 | 0.100 | 0.463 |
| Oscillibacter sp900066435 | fixed | NA | groupFBVLED:time_pointFinal | -0.67 | 0.40 | -1.68 | 39.00 | -1.48 | 0.14 | 0.100 | 0.463 |
| Oscillibacter MIC6445 | fixed | NA | groupFBVLED:time_pointFinal | 0.20 | 0.12 | 1.68 | 39.00 | -0.04 | 0.43 | 0.100 | 0.463 |
| Anaerococcus MIC7502 | fixed | NA | groupFBVLED:time_pointFinal | 0.31 | 0.19 | 1.68 | 39.00 | -0.06 | 0.69 | 0.101 | 0.463 |
| Eubacterium_G ventriosum | fixed | NA | groupFBVLED:time_pointFinal | 0.38 | 0.23 | 1.68 | 39.00 | -0.08 | 0.85 | 0.102 | 0.463 |
| Alistipes_A sp900240235 | fixed | NA | groupFBVLED:time_pointFinal | 0.33 | 0.20 | 1.67 | 39.00 | -0.07 | 0.74 | 0.102 | 0.463 |
| F23 B02 sp001916715 | fixed | NA | groupFBVLED:time_pointFinal | 0.57 | 0.34 | 1.67 | 39.00 | -0.12 | 1.26 | 0.104 | 0.463 |
| CAG 272 MIC6999 | fixed | NA | groupFBVLED:time_pointFinal | -0.44 | 0.27 | -1.66 | 39.00 | -0.99 | 0.10 | 0.104 | 0.463 |
| Lachnospiraceae MIC7886 | fixed | NA | groupFBVLED:time_pointFinal | 0.17 | 0.10 | 1.66 | 39.00 | -0.04 | 0.37 | 0.105 | 0.463 |
| Blautia sp000436935 | fixed | NA | groupFBVLED:time_pointFinal | 0.50 | 0.30 | 1.65 | 39.00 | -0.11 | 1.12 | 0.106 | 0.463 |
| Blautia_A MIC9663 | fixed | NA | groupFBVLED:time_pointFinal | -0.17 | 0.10 | -1.65 | 39.00 | -0.38 | 0.04 | 0.108 | 0.463 |
| Lawsonibacter sp000177015 | fixed | NA | groupFBVLED:time_pointFinal | 0.10 | 0.06 | 1.65 | 39.00 | -0.02 | 0.22 | 0.108 | 0.463 |

| OUTCOMES - SPECIES | EFFECT | GROUP | TERM | ESTIMATE | STD ERROR | STATISTIC | DF | CONF LOW | CONF HIGH | P VALUE | Q VALUE |
|---|---|---|---|---|---|---|---|---|---|---|---|
| Peptoniphilus_A grossensis_B | fixed | NA | groupFBVLED:time_pointFinal | 0.71 | 0.43 | 1.65 | 39.00 | -0.16 | 1.58 | 0.108 | 0.463 |
| NK3B98 MIC8354 | fixed | NA | groupFBVLED:time_pointFinal | 0.23 | 0.14 | 1.63 | 39.00 | -0.06 | 0.53 | 0.111 | 0.466 |
| Dorea longicatena | fixed | NA | groupFBVLED:time_pointFinal | 0.50 | 0.31 | 1.63 | 39.00 | -0.12 | 1.13 | 0.112 | 0.466 |
| Dorea sp900066555 | fixed | NA | groupFBVLED:time_pointFinal | 0.36 | 0.22 | 1.62 | 39.00 | -0.09 | 0.81 | 0.112 | 0.466 |
| Akkermansia muciniphila | fixed | NA | groupFBVLED:time_pointFinal | -0.80 | 0.49 | -1.62 | 39.00 | -1.79 | 0.20 | 0.113 | 0.466 |
| CAG 314 sp000437915 | fixed | NA | groupFBVLED:time_pointFinal | 0.53 | 0.33 | 1.62 | 39.00 | -0.13 | 1.19 | 0.113 | 0.466 |
| Slackia_A piriformis | fixed | NA | groupFBVLED:time_pointFinal | 0.07 | 0.05 | 1.61 | 39.00 | -0.02 | 0.17 | 0.115 | 0.466 |
| CAG 313 sp000433035 | fixed | NA | groupFBVLED:time_pointFinal | -0.34 | 0.21 | -1.61 | 39.00 | -0.76 | 0.09 | 0.116 | 0.466 |
| Clostridium_M asparagiforme | fixed | NA | groupFBVLED:time_pointFinal | -0.32 | 0.20 | -1.59 | 78.00 | -0.72 | 0.08 | 0.116 | 0.466 |
| Bifidobacterium adolescentis | fixed | NA | groupFBVLED:time_pointFinal | -0.68 | 0.43 | -1.61 | 39.00 | -1.54 | 0.18 | 0.116 | 0.466 |
| ER4 MIC9395 | fixed | NA | groupFBVLED:time_pointFinal | 0.21 | 0.13 | 1.60 | 39.00 | -0.06 | 0.48 | 0.117 | 0.466 |
| Ruminococcus_E bromii_B | fixed | NA | groupFBVLED:time_pointFinal | 0.99 | 0.62 | 1.60 | 39.00 | -0.26 | 2.25 | 0.118 | 0.466 |
| Ruminococcus_E sp003526955 | fixed | NA | groupFBVLED:time_pointFinal | 0.66 | 0.41 | 1.59 | 39.00 | -0.18 | 1.49 | 0.120 | 0.466 |
| Ruthenibacterium sp003149955 | fixed | NA | groupFBVLED:time_pointFinal | -0.28 | 0.18 | -1.57 | 78.00 | -0.63 | 0.07 | 0.120 | 0.466 |
| UBA1191 MIC7533 | fixed | NA | groupFBVLED:time_pointFinal | 0.09 | 0.05 | 1.59 | 39.00 | -0.02 | 0.20 | 0.121 | 0.466 |
| Blautia_A sp900066165 | fixed | NA | groupFBVLED:time_pointFinal | 0.46 | 0.29 | 1.58 | 39.00 | -0.13 | 1.06 | 0.123 | 0.466 |
| CAG 145 MIC8493 | fixed | NA | groupFBVLED:time_pointFinal | 0.12 | 0.08 | 1.58 | 39.00 | -0.03 | 0.28 | 0.123 | 0.466 |
| CAG 74 MIC6989 | fixed | NA | groupFBVLED:time_pointFinal | -0.49 | 0.31 | -1.58 | 39.00 | -1.12 | 0.14 | 0.123 | 0.466 |
| CAG 41 sp900066215 | fixed | NA | groupFBVLED:time_pointFinal | 0.78 | 0.49 | 1.58 | 39.00 | -0.22 | 1.77 | 0.123 | 0.466 |
| Marvinbryantia sp900066075 | fixed | NA | groupFBVLED:time_pointFinal | 0.17 | 0.11 | 1.57 | 39.00 | -0.05 | 0.40 | 0.124 | 0.466 |
| Alistipes obesi | fixed | NA | groupFBVLED:time_pointFinal | 0.55 | 0.35 | 1.57 | 39.00 | -0.16 | 1.26 | 0.124 | 0.466 |
| Anaerostipes caccae | fixed | NA | groupFBVLED:time_pointFinal | -0.33 | 0.21 | -1.57 | 39.00 | -0.75 | 0.09 | 0.125 | 0.467 |
| Erysipelatoclostridium sp000752095 | fixed | NA | groupFBVLED:time_pointFinal | -0.47 | 0.30 | -1.56 | 39.00 | -1.08 | 0.14 | 0.127 | 0.470 |
| TF01 11 sp003529475 | fixed | NA | groupFBVLED:time_pointFinal | 0.54 | 0.35 | 1.56 | 39.00 | -0.16 | 1.24 | 0.127 | 0.470 |
| Pseudoflavonifractor MIC6616 | fixed | NA | groupFBVLED:time_pointFinal | -0.37 | 0.24 | -1.54 | 78.00 | -0.84 | 0.11 | 0.128 | 0.470 |
| CAG 269 sp000437215 | fixed | NA | groupFBVLED:time_pointFinal | 0.24 | 0.15 | 1.55 | 39.00 | -0.07 | 0.55 | 0.130 | 0.473 |
| Bacteroides fragilis | fixed | NA | groupFBVLED:time_pointFinal | 0.39 | 0.25 | 1.55 | 39.00 | -0.12 | 0.90 | 0.130 | 0.473 |
| Ruthenibacterium MIC9423 | fixed | NA | groupFBVLED:time_pointFinal | -0.22 | 0.14 | -1.54 | 39.00 | -0.51 | 0.07 | 0.133 | 0.476 |
| Acutalibacteraceae MIC7526 | fixed | NA | groupFBVLED:time_pointFinal | -0.27 | 0.17 | -1.53 | 39.00 | -0.62 | 0.09 | 0.133 | 0.476 |

| OUTCOMES - SPECIES | EFFECT | GROUP | TERM | ESTIMATE | STD ERROR | STATISTIC | DF | CONF LOW | CONF HIGH | P VALUE | Q VALUE |
|---|---|---|---|---|---|---|---|---|---|---|---|
| QAND01 MIC9470 | fixed | NA | groupFBVLED:time_pointFinal | -0.59 | 0.38 | -1.53 | 39.00 | -1.36 | 0.19 | 0.134 | 0.476 |
| Massilioclostridium coli | fixed | NA | groupFBVLED:time_pointFinal | -0.16 | 0.10 | -1.53 | 39.00 | -0.36 | 0.05 | 0.134 | 0.476 |
| Bifidobacterium bifidum | fixed | NA | groupFBVLED:time_pointFinal | -0.26 | 0.17 | -1.53 | 39.00 | -0.60 | 0.08 | 0.135 | 0.476 |
| CAG 313 MIC9072 | fixed | NA | groupFBVLED:time_pointFinal | 0.33 | 0.21 | 1.52 | 39.00 | -0.11 | 0.76 | 0.136 | 0.476 |
| Gordonibacter pamelaeae | fixed | NA | groupFBVLED:time_pointFinal | -0.30 | 0.20 | -1.51 | 39.00 | -0.70 | 0.10 | 0.138 | 0.476 |
| CAG 103 MIC7540 | fixed | NA | groupFBVLED:time_pointFinal | 0.27 | 0.18 | 1.51 | 39.00 | -0.09 | 0.62 | 0.138 | 0.476 |
| CAG 170 MIC9129 | fixed | NA | groupFBVLED:time_pointFinal | 0.19 | 0.13 | 1.51 | 39.00 | -0.07 | 0.45 | 0.139 | 0.476 |
| UBA5394 sp003150565 | fixed | NA | groupFBVLED:time_pointFinal | -0.40 | 0.27 | -1.49 | 78.00 | -0.93 | 0.13 | 0.139 | 0.476 |
| Blautia_A MIC7329 | fixed | NA | groupFBVLED:time_pointFinal | 0.15 | 0.10 | 1.51 | 39.00 | -0.05 | 0.34 | 0.140 | 0.476 |
| UBA6398 MIC8483 | fixed | NA | groupFBVLED:time_pointFinal | 0.06 | 0.04 | 1.51 | 39.00 | -0.02 | 0.13 | 0.140 | 0.476 |
| CAG 460 sp000437355 | fixed | NA | groupFBVLED:time_pointFinal | 0.12 | 0.08 | 1.50 | 39.00 | -0.04 | 0.28 | 0.142 | 0.481 |
| Bifidobacterium MIC6680 | fixed | NA | groupFBVLED:time_pointFinal | -0.26 | 0.17 | -1.49 | 39.00 | -0.60 | 0.09 | 0.143 | 0.482 |
| Barnesiella intestinihominis | fixed | NA | groupFBVLED:time_pointFinal | 0.50 | 0.34 | 1.49 | 39.00 | -0.18 | 1.19 | 0.145 | 0.484 |
| Acutalibacter MIC8008 | fixed | NA | groupFBVLED:time_pointFinal | -0.45 | 0.31 | -1.47 | 39.00 | -1.07 | 0.17 | 0.149 | 0.492 |
| Peptoniphilus_B duerdenii | fixed | NA | groupFBVLED:time_pointFinal | 0.27 | 0.18 | 1.46 | 78.00 | -0.10 | 0.63 | 0.149 | 0.492 |
| CAG 74 MIC7044 | fixed | NA | groupFBVLED:time_pointFinal | 0.40 | 0.27 | 1.46 | 39.00 | -0.15 | 0.94 | 0.152 | 0.492 |
| CAG 83 MIC7389 | fixed | NA | groupFBVLED:time_pointFinal | -0.31 | 0.21 | -1.46 | 39.00 | -0.74 | 0.12 | 0.153 | 0.492 |
| Intestinibacter bartlettii | fixed | NA | groupFBVLED:time_pointFinal | 0.50 | 0.34 | 1.46 | 39.00 | -0.19 | 1.19 | 0.153 | 0.492 |
| CAG 354 sp001915925 | fixed | NA | groupFBVLED:time_pointFinal | 0.11 | 0.08 | 1.45 | 39.00 | -0.04 | 0.27 | 0.154 | 0.492 |
| Romboutsia timonensis | fixed | NA | groupFBVLED:time_pointFinal | 0.51 | 0.35 | 1.45 | 39.00 | -0.20 | 1.21 | 0.154 | 0.492 |
| Streptococcus anginosus_C | fixed | NA | groupFBVLED:time_pointFinal | 0.29 | 0.20 | 1.44 | 78.00 | -0.11 | 0.69 | 0.155 | 0.492 |
| Bilophila MIC7011 | fixed | NA | groupFBVLED:time_pointFinal | 0.07 | 0.05 | 1.45 | 39.00 | -0.03 | 0.17 | 0.156 | 0.492 |
| Parabacteroides johnsonii | fixed | NA | groupFBVLED:time_pointFinal | -0.21 | 0.15 | -1.45 | 39.00 | -0.51 | 0.08 | 0.156 | 0.492 |
| CAG 272 MIC8971 | fixed | NA | groupFBVLED:time_pointFinal | 0.17 | 0.12 | 1.44 | 39.00 | -0.07 | 0.41 | 0.157 | 0.492 |
| UBA1777 MIC6732 | fixed | NA | groupFBVLED:time_pointFinal | 0.24 | 0.17 | 1.44 | 39.00 | -0.10 | 0.59 | 0.157 | 0.492 |
| UBA1829 sp002338895 | fixed | NA | groupFBVLED:time_pointFinal | 0.17 | 0.12 | 1.44 | 39.00 | -0.07 | 0.40 | 0.158 | 0.492 |
| Desulfovibrio piger | fixed | NA | groupFBVLED:time_pointFinal | -0.12 | 0.08 | -1.44 | 39.00 | -0.29 | 0.05 | 0.159 | 0.492 |
| Bacteroides nordii | fixed | NA | groupFBVLED:time_pointFinal | -0.39 | 0.27 | -1.43 | 39.00 | -0.95 | 0.16 | 0.160 | 0.492 |
| Prevotella sp001275135 | fixed | NA | groupFBVLED:time_pointFinal | 0.09 | 0.06 | 1.43 | 39.00 | -0.04 | 0.21 | 0.160 | 0.492 |

| OUTCOMES - SPECIES | EFFECT | GROUP | TERM | ESTIMATE | STD ERROR | STATISTIC | DF | CONF LOW | CONF HIGH | P VALUE | Q VALUE |
|---|---|---|---|---|---|---|---|---|---|---|---|
| Blautia_A MIC7810 | fixed | NA | groupFBVLED:time_pointFinal | 0.15 | 0.11 | 1.43 | 39.00 | -0.06 | 0.37 | 0.160 | 0.492 |
| Parasutterella sp000980495 | fixed | NA | groupFBVLED:time_pointFinal | 0.19 | 0.14 | 1.43 | 39.00 | -0.08 | 0.47 | 0.162 | 0.492 |
| Oscillospiraceae MIC9607 | fixed | NA | groupFBVLED:time_pointFinal | -0.18 | 0.13 | -1.42 | 39.00 | -0.43 | 0.08 | 0.163 | 0.492 |
| Akkermansia muciniphila_B | fixed | NA | groupFBVLED:time_pointFinal | 0.61 | 0.43 | 1.42 | 39.00 | -0.26 | 1.48 | 0.163 | 0.492 |
| Lachnospiraceae MIC9747 | fixed | NA | groupFBVLED:time_pointFinal | -0.21 | 0.15 | -1.41 | 39.00 | -0.51 | 0.09 | 0.165 | 0.494 |
| Adlercreutzia MIC8014 | fixed | NA | groupFBVLED:time_pointFinal | 0.41 | 0.29 | 1.41 | 39.00 | -0.18 | 1.00 | 0.166 | 0.494 |
| UBA5416 MIC7893 | fixed | NA | groupFBVLED:time_pointFinal | -0.29 | 0.21 | -1.41 | 39.00 | -0.72 | 0.13 | 0.166 | 0.494 |
| Murdochiella MIC8251 | fixed | NA | groupFBVLED:time_pointFinal | 0.30 | 0.21 | 1.41 | 39.00 | -0.13 | 0.73 | 0.167 | 0.494 |
| Faecalibacterium prausnitzii_J | fixed | NA | groupFBVLED:time_pointFinal | 0.25 | 0.18 | 1.41 | 39.00 | -0.11 | 0.62 | 0.168 | 0.494 |
| Flavonifractor sp000508885 | fixed | NA | groupFBVLED:time_pointFinal | -0.45 | 0.32 | -1.39 | 39.00 | -1.10 | 0.20 | 0.171 | 0.498 |
| Mogibacterium massiliense | fixed | NA | groupFBVLED:time_pointFinal | 0.40 | 0.29 | 1.38 | 78.00 | -0.18 | 0.97 | 0.172 | 0.498 |
| UBA738 sp003522945 | fixed | NA | groupFBVLED:time_pointFinal | -0.32 | 0.23 | -1.39 | 39.00 | -0.78 | 0.14 | 0.172 | 0.498 |
| Acidaminococcus intestini | fixed | NA | groupFBVLED:time_pointFinal | 0.06 | 0.04 | 1.39 | 39.00 | -0.03 | 0.15 | 0.172 | 0.498 |
| Blautia_A sp900066505 | fixed | NA | groupFBVLED:time_pointFinal | 0.34 | 0.24 | 1.39 | 39.00 | -0.15 | 0.83 | 0.173 | 0.498 |
| Lachnospiraceae MIC6885 | fixed | NA | groupFBVLED:time_pointFinal | 0.01 | 0.01 | 1.38 | 39.00 | 0.00 | 0.03 | 0.175 | 0.501 |
| CAG 74 MIC8660 | fixed | NA | groupFBVLED:time_pointFinal | -0.14 | 0.10 | -1.38 | 39.00 | -0.34 | 0.07 | 0.177 | 0.504 |
| Coprobacter fastidiosus | fixed | NA | groupFBVLED:time_pointFinal | 0.37 | 0.27 | 1.37 | 39.00 | -0.18 | 0.91 | 0.178 | 0.506 |
| Facklamia hominis | fixed | NA | groupFBVLED:time_pointFinal | 0.30 | 0.22 | 1.35 | 78.00 | -0.14 | 0.73 | 0.181 | 0.509 |
| UBA11452 sp003526375 | fixed | NA | groupFBVLED:time_pointFinal | -0.19 | 0.14 | -1.36 | 39.00 | -0.46 | 0.09 | 0.181 | 0.509 |
| CAG 145 MIC7639 | fixed | NA | groupFBVLED:time_pointFinal | -0.13 | 0.10 | -1.36 | 39.00 | -0.33 | 0.07 | 0.182 | 0.509 |
| CAG 727 MIC8506 | fixed | NA | groupFBVLED:time_pointFinal | 0.28 | 0.21 | 1.36 | 39.00 | -0.14 | 0.70 | 0.183 | 0.511 |
| Ezakiella MIC8494 | fixed | NA | groupFBVLED:time_pointFinal | 0.35 | 0.26 | 1.33 | 78.00 | -0.17 | 0.87 | 0.186 | 0.513 |
| Bacteroides_B MIC8119 | fixed | NA | groupFBVLED:time_pointFinal | -0.18 | 0.13 | -1.35 | 39.00 | -0.45 | 0.09 | 0.186 | 0.513 |
| Lawsonella MIC8032 | fixed | NA | groupFBVLED:time_pointFinal | 0.41 | 0.30 | 1.33 | 78.00 | -0.20 | 1.01 | 0.187 | 0.513 |
| Christensenellales MIC6424 | fixed | NA | groupFBVLED:time_pointFinal | -0.06 | 0.04 | -1.34 | 39.00 | -0.14 | 0.03 | 0.188 | 0.514 |
| CAG 267 sp001917135 | fixed | NA | groupFBVLED:time_pointFinal | -0.29 | 0.21 | -1.33 | 39.00 | -0.72 | 0.15 | 0.191 | 0.518 |
| Gabonibacter massiliensis | fixed | NA | groupFBVLED:time_pointFinal | -0.10 | 0.07 | -1.33 | 39.00 | -0.24 | 0.05 | 0.191 | 0.518 |
| QAND01 MIC9113 | fixed | NA | groupFBVLED:time_pointFinal | -0.36 | 0.27 | -1.32 | 39.00 | -0.90 | 0.19 | 0.193 | 0.520 |
| CAG 312 MIC7338 | fixed | NA | groupFBVLED:time_pointFinal | 0.24 | 0.18 | 1.32 | 39.00 | -0.13 | 0.60 | 0.194 | 0.520 |

| OUTCOMES - SPECIES | EFFECT | GROUP | TERM | ESTIMATE | STD ERROR | STATISTIC | DF | CONF LOW | CONF HIGH | P VALUE | Q VALUE |
|---|---|---|---|---|---|---|---|---|---|---|---|
| Alistipes_A indistinctus | fixed | NA | groupFBVLED:time_pointFinal | -0.40 | 0.30 | -1.32 | 39.00 | -1.00 | 0.21 | 0.194 | 0.520 |
| Faecalibacterium prausnitzii_C | fixed | NA | groupFBVLED:time_pointFinal | 0.42 | 0.32 | 1.31 | 39.00 | -0.23 | 1.08 | 0.198 | 0.528 |
| Clostridium_M sp000431375 | fixed | NA | groupFBVLED:time_pointFinal | 0.51 | 0.39 | 1.30 | 39.00 | -0.28 | 1.31 | 0.200 | 0.530 |
| Bacteroides_B vulgatus | fixed | NA | groupFBVLED:time_pointFinal | 0.51 | 0.39 | 1.30 | 39.00 | -0.28 | 1.29 | 0.200 | 0.530 |
| Peptoniphilus_C MIC7895 | fixed | NA | groupFBVLED:time_pointFinal | 0.28 | 0.22 | 1.28 | 78.00 | -0.16 | 0.73 | 0.204 | 0.533 |
| Tyzzerella MIC9817 | fixed | NA | groupFBVLED:time_pointFinal | -0.22 | 0.17 | -1.29 | 39.00 | -0.56 | 0.12 | 0.204 | 0.533 |
| UBA5394 MIC7155 | fixed | NA | groupFBVLED:time_pointFinal | -0.22 | 0.17 | -1.29 | 39.00 | -0.57 | 0.13 | 0.204 | 0.533 |
| Bacteroides xylanisolvens | fixed | NA | groupFBVLED:time_pointFinal | 0.49 | 0.38 | 1.29 | 39.00 | -0.28 | 1.26 | 0.206 | 0.537 |
| Parvimonas sp001553085 | fixed | NA | groupFBVLED:time_pointFinal | -0.07 | 0.05 | -1.27 | 39.00 | -0.17 | 0.04 | 0.212 | 0.546 |
| CAG 74 MIC9156 | fixed | NA | groupFBVLED:time_pointFinal | -0.45 | 0.35 | -1.27 | 39.00 | -1.16 | 0.27 | 0.213 | 0.546 |
| UBA866 MIC8205 | fixed | NA | groupFBVLED:time_pointFinal | -0.21 | 0.17 | -1.26 | 39.00 | -0.54 | 0.13 | 0.214 | 0.546 |
| Lachnospirales MIC6978 | fixed | NA | groupFBVLED:time_pointFinal | -0.11 | 0.09 | -1.26 | 39.00 | -0.28 | 0.07 | 0.214 | 0.546 |
| CAG 495 sp001917125 | fixed | NA | groupFBVLED:time_pointFinal | 0.15 | 0.12 | 1.26 | 39.00 | -0.09 | 0.39 | 0.215 | 0.546 |
| Lachnospiraceae MIC9280 | fixed | NA | groupFBVLED:time_pointFinal | -0.20 | 0.16 | -1.26 | 39.00 | -0.53 | 0.12 | 0.215 | 0.546 |
| Mailhella sp003150275 | fixed | NA | groupFBVLED:time_pointFinal | -0.09 | 0.07 | -1.25 | 39.00 | -0.22 | 0.05 | 0.217 | 0.549 |
| Anaerotruncus sp900199635 | fixed | NA | groupFBVLED:time_pointFinal | 0.13 | 0.11 | 1.24 | 39.00 | -0.08 | 0.35 | 0.222 | 0.557 |
| Bacteroides intestinalis_A | fixed | NA | groupFBVLED:time_pointFinal | -0.10 | 0.08 | -1.24 | 39.00 | -0.27 | 0.06 | 0.222 | 0.557 |
| Fusicatenibacter MIC7088 | fixed | NA | groupFBVLED:time_pointFinal | 0.10 | 0.08 | 1.22 | 78.00 | -0.06 | 0.26 | 0.225 | 0.561 |
| Dorea sp000509125 | fixed | NA | groupFBVLED:time_pointFinal | -0.21 | 0.17 | -1.23 | 39.00 | -0.54 | 0.13 | 0.226 | 0.561 |
| GCA 900066575 MIC7948 | fixed | NA | groupFBVLED:time_pointFinal | 0.24 | 0.19 | 1.23 | 39.00 | -0.15 | 0.63 | 0.227 | 0.561 |
| Peptoniphilus_A lacrimalis | fixed | NA | groupFBVLED:time_pointFinal | 0.52 | 0.42 | 1.22 | 39.00 | -0.34 | 1.37 | 0.228 | 0.563 |
| Lactobacillus_C paracasei | fixed | NA | groupFBVLED:time_pointFinal | 0.15 | 0.13 | 1.20 | 78.00 | -0.10 | 0.41 | 0.233 | 0.573 |
| Acutalibacteraceae MIC6990 | fixed | NA | groupFBVLED:time_pointFinal | 0.32 | 0.26 | 1.21 | 39.00 | -0.21 | 0.85 | 0.234 | 0.573 |
| CAG 145 sp000435715 | fixed | NA | groupFBVLED:time_pointFinal | -0.37 | 0.31 | -1.21 | 39.00 | -0.99 | 0.25 | 0.235 | 0.573 |
| Anaerostipes hadrus_A | fixed | NA | groupFBVLED:time_pointFinal | -0.38 | 0.31 | -1.20 | 39.00 | -1.01 | 0.26 | 0.238 | 0.577 |
| Clostridium_M citroniae | fixed | NA | groupFBVLED:time_pointFinal | -0.23 | 0.19 | -1.18 | 78.00 | -0.61 | 0.16 | 0.240 | 0.578 |
| CAG 83 MIC7830 | fixed | NA | groupFBVLED:time_pointFinal | 0.14 | 0.12 | 1.19 | 39.00 | -0.10 | 0.38 | 0.243 | 0.578 |
| Haemophilus_D sp001815355 | fixed | NA | groupFBVLED:time_pointFinal | -0.18 | 0.15 | -1.18 | 78.00 | -0.48 | 0.12 | 0.243 | 0.578 |
| UBA1394 MIC7236 | fixed | NA | groupFBVLED:time_pointFinal | -0.08 | 0.06 | -1.18 | 39.00 | -0.21 | 0.05 | 0.243 | 0.578 |

| OUTCOMES - SPECIES | EFFECT | GROUP | TERM | ESTIMATE | STD ERROR | STATISTIC | DF | CONF LOW | CONF HIGH | P VALUE | Q VALUE |
|---|---|---|---|---|---|---|---|---|---|---|---|
| Peptoniphilus_A MIC9267 | fixed | NA | groupFBVLED:time_pointFinal | 0.37 | 0.31 | 1.18 | 39.00 | -0.26 | 0.99 | 0.244 | 0.578 |
| CAG 269 sp001915995 | fixed | NA | groupFBVLED:time_pointFinal | -0.05 | 0.04 | -1.18 | 39.00 | -0.14 | 0.04 | 0.245 | 0.578 |
| CAG 110 MIC9052 | fixed | NA | groupFBVLED:time_pointFinal | -0.20 | 0.17 | -1.18 | 39.00 | -0.55 | 0.14 | 0.245 | 0.578 |
| CAG 417 sp000432835 | fixed | NA | groupFBVLED:time_pointFinal | 0.23 | 0.19 | 1.18 | 39.00 | -0.16 | 0.61 | 0.247 | 0.578 |
| S5 A14a MIC8631 | fixed | NA | groupFBVLED:time_pointFinal | 0.19 | 0.17 | 1.17 | 78.00 | -0.14 | 0.53 | 0.247 | 0.578 |
| Senegalimassilia anaerobia | fixed | NA | groupFBVLED:time_pointFinal | -0.16 | 0.14 | -1.17 | 39.00 | -0.44 | 0.12 | 0.249 | 0.578 |
| CAG 74 MIC8780 | fixed | NA | groupFBVLED:time_pointFinal | -0.08 | 0.07 | -1.17 | 39.00 | -0.23 | 0.06 | 0.249 | 0.578 |
| Lachnospiraceae MIC6593 | fixed | NA | groupFBVLED:time_pointFinal | 0.16 | 0.14 | 1.16 | 39.00 | -0.12 | 0.43 | 0.255 | 0.586 |
| CAG 74 MIC8853 | fixed | NA | groupFBVLED:time_pointFinal | -0.07 | 0.06 | -1.16 | 39.00 | -0.20 | 0.06 | 0.255 | 0.586 |
| Alistipes sp000434235 | fixed | NA | groupFBVLED:time_pointFinal | 0.20 | 0.17 | 1.15 | 39.00 | -0.15 | 0.54 | 0.257 | 0.586 |
| Marseille P4683 sp900232885 | fixed | NA | groupFBVLED:time_pointFinal | -0.20 | 0.17 | -1.15 | 39.00 | -0.54 | 0.15 | 0.257 | 0.586 |
| CAG 312 sp000438015 | fixed | NA | groupFBVLED:time_pointFinal | 0.12 | 0.11 | 1.15 | 39.00 | -0.09 | 0.34 | 0.258 | 0.586 |
| CAG 83 sp001916855 | fixed | NA | groupFBVLED:time_pointFinal | 0.35 | 0.30 | 1.15 | 39.00 | -0.27 | 0.96 | 0.259 | 0.586 |
| Ezakiella coagulans | fixed | NA | groupFBVLED:time_pointFinal | 0.33 | 0.29 | 1.14 | 78.00 | -0.24 | 0.90 | 0.259 | 0.586 |
| CAG 170 MIC6856 | fixed | NA | groupFBVLED:time_pointFinal | 0.29 | 0.25 | 1.14 | 39.00 | -0.22 | 0.79 | 0.260 | 0.586 |
| UBA9502 MIC7149 | fixed | NA | groupFBVLED:time_pointFinal | 0.19 | 0.17 | 1.14 | 39.00 | -0.15 | 0.53 | 0.262 | 0.586 |
| CAG 95 sp000438155 | fixed | NA | groupFBVLED:time_pointFinal | 0.23 | 0.21 | 1.14 | 39.00 | -0.18 | 0.65 | 0.262 | 0.586 |
| CAG 83 sp000435555 | fixed | NA | groupFBVLED:time_pointFinal | -0.46 | 0.41 | -1.14 | 39.00 | -1.29 | 0.36 | 0.262 | 0.586 |
| UBA644 MIC9596 | fixed | NA | groupFBVLED:time_pointFinal | -0.12 | 0.11 | -1.14 | 39.00 | -0.33 | 0.09 | 0.263 | 0.586 |
| Desulfovibrio piger_A | fixed | NA | groupFBVLED:time_pointFinal | -0.06 | 0.05 | -1.13 | 39.00 | -0.17 | 0.05 | 0.265 | 0.587 |
| Anaerovoracaceae MIC7161 | fixed | NA | groupFBVLED:time_pointFinal | 0.10 | 0.09 | 1.12 | 39.00 | -0.08 | 0.27 | 0.271 | 0.599 |
| Acutalibacteraceae MIC7795 | fixed | NA | groupFBVLED:time_pointFinal | -0.24 | 0.22 | -1.11 | 39.00 | -0.69 | 0.20 | 0.273 | 0.600 |
| PeH17 sp000435055 | fixed | NA | groupFBVLED:time_pointFinal | 0.55 | 0.49 | 1.11 | 39.00 | -0.45 | 1.55 | 0.273 | 0.600 |
| Collinsella MIC8209 | fixed | NA | groupFBVLED:time_pointFinal | -0.14 | 0.13 | -1.10 | 39.00 | -0.41 | 0.12 | 0.276 | 0.603 |
| UCG 010 sp003150215 | fixed | NA | groupFBVLED:time_pointFinal | -0.20 | 0.18 | -1.09 | 77.99 | -0.57 | 0.17 | 0.277 | 0.603 |
| Bacteroides_A plebeius_A | fixed | NA | groupFBVLED:time_pointFinal | 0.26 | 0.24 | 1.10 | 39.00 | -0.22 | 0.74 | 0.278 | 0.603 |
| Lactococcus lactis | fixed | NA | groupFBVLED:time_pointFinal | -0.25 | 0.23 | -1.09 | 39.00 | -0.71 | 0.21 | 0.282 | 0.611 |
| Blautia producta | fixed | NA | groupFBVLED:time_pointFinal | -0.21 | 0.19 | -1.08 | 39.00 | -0.60 | 0.18 | 0.288 | 0.619 |
| CAG 312 MIC7045 | fixed | NA | groupFBVLED:time_pointFinal | -0.11 | 0.10 | -1.08 | 39.00 | -0.30 | 0.09 | 0.288 | 0.619 |

| OUTCOMES - SPECIES | EFFECT | GROUP | TERM | ESTIMATE | STD ERROR | STATISTIC | DF | CONF LOW | CONF HIGH | P VALUE | Q VALUE |
|---|---|---|---|---|---|---|---|---|---|---|---|
| Negativibacillus MIC7916 | fixed | NA | groupFBVLED:time_pointFinal | 0.14 | 0.13 | 1.07 | 39.00 | -0.13 | 0.41 | 0.291 | 0.619 |
| Ruminococcus_C sp000980705 | fixed | NA | groupFBVLED:time_pointFinal | 0.39 | 0.36 | 1.07 | 39.00 | -0.34 | 1.11 | 0.291 | 0.619 |
| UBA5446 MIC7746 | fixed | NA | groupFBVLED:time_pointFinal | -0.32 | 0.29 | -1.07 | 39.00 | -0.91 | 0.28 | 0.292 | 0.619 |
| Holdemania sp900120005 | fixed | NA | groupFBVLED:time_pointFinal | 0.11 | 0.10 | 1.06 | 78.00 | -0.09 | 0.31 | 0.292 | 0.619 |
| Holdemania massiliensis | fixed | NA | groupFBVLED:time_pointFinal | 0.11 | 0.11 | 1.06 | 78.00 | -0.10 | 0.33 | 0.294 | 0.622 |
| Haemophilus_D parainfluenzae | fixed | NA | groupFBVLED:time_pointFinal | -0.20 | 0.19 | -1.05 | 39.00 | -0.59 | 0.19 | 0.300 | 0.632 |
| Acutalibacter MIC8974 | fixed | NA | groupFBVLED:time_pointFinal | -0.18 | 0.18 | -1.04 | 78.00 | -0.53 | 0.17 | 0.302 | 0.632 |
| CAG 83 sp000435975 | fixed | NA | groupFBVLED:time_pointFinal | -0.37 | 0.35 | -1.05 | 39.00 | -1.07 | 0.34 | 0.302 | 0.632 |
| Sutterella wadsworthensis_B | fixed | NA | groupFBVLED:time_pointFinal | -0.20 | 0.20 | -1.04 | 39.00 | -0.60 | 0.19 | 0.303 | 0.632 |
| CAG 74 MIC8717 | fixed | NA | groupFBVLED:time_pointFinal | 0.14 | 0.14 | 1.03 | 78.00 | -0.13 | 0.42 | 0.304 | 0.632 |
| Bacteroides sp900066265 | fixed | NA | groupFBVLED:time_pointFinal | 0.08 | 0.08 | 1.04 | 39.00 | -0.08 | 0.25 | 0.306 | 0.632 |
| UBA1394 sp900066845 | fixed | NA | groupFBVLED:time_pointFinal | 0.26 | 0.25 | 1.04 | 39.00 | -0.24 | 0.75 | 0.307 | 0.632 |
| TF01 11 sp001916135 | fixed | NA | groupFBVLED:time_pointFinal | 0.12 | 0.12 | 1.03 | 39.00 | -0.12 | 0.36 | 0.309 | 0.632 |
| Agathobacter rectale | fixed | NA | groupFBVLED:time_pointFinal | 0.52 | 0.50 | 1.03 | 39.00 | -0.50 | 1.53 | 0.309 | 0.632 |
| CAG 194 sp000432915 | fixed | NA | groupFBVLED:time_pointFinal | -0.20 | 0.19 | -1.03 | 39.00 | -0.58 | 0.19 | 0.309 | 0.632 |
| Agathobacter MIC9834 | fixed | NA | groupFBVLED:time_pointFinal | 0.17 | 0.17 | 1.01 | 77.84 | -0.17 | 0.51 | 0.317 | 0.643 |
| CAG 274 sp000432155 | fixed | NA | groupFBVLED:time_pointFinal | 0.30 | 0.29 | 1.01 | 39.00 | -0.30 | 0.89 | 0.317 | 0.643 |
| Urmitella timonensis | fixed | NA | groupFBVLED:time_pointFinal | -0.32 | 0.32 | -1.01 | 39.00 | -0.97 | 0.33 | 0.320 | 0.648 |
| CAG 272 MIC9176 | fixed | NA | groupFBVLED:time_pointFinal | -0.02 | 0.02 | -1.00 | 39.00 | -0.06 | 0.02 | 0.323 | 0.652 |
| UBA11471 sp000434215 | fixed | NA | groupFBVLED:time_pointFinal | 0.24 | 0.24 | 0.99 | 39.00 | -0.25 | 0.72 | 0.328 | 0.659 |
| Prevotella sp003447235 | fixed | NA | groupFBVLED:time_pointFinal | 0.29 | 0.29 | 0.99 | 39.00 | -0.30 | 0.88 | 0.329 | 0.659 |
| 51 20 sp001917175 | fixed | NA | groupFBVLED:time_pointFinal | 0.31 | 0.31 | 0.98 | 39.00 | -0.32 | 0.93 | 0.331 | 0.659 |
| Dorea formicigenerans | fixed | NA | groupFBVLED:time_pointFinal | 0.21 | 0.21 | 0.98 | 39.00 | -0.22 | 0.63 | 0.333 | 0.659 |
| Porphyromonas somerae | fixed | NA | groupFBVLED:time_pointFinal | 0.17 | 0.18 | 0.97 | 78.00 | -0.18 | 0.53 | 0.333 | 0.659 |
| UBA1390 sp002305315 | fixed | NA | groupFBVLED:time_pointFinal | -0.16 | 0.16 | -0.98 | 39.00 | -0.48 | 0.17 | 0.333 | 0.659 |
| UBA7182 MIC8422 | fixed | NA | groupFBVLED:time_pointFinal | 0.17 | 0.17 | 0.97 | 39.00 | -0.18 | 0.52 | 0.336 | 0.663 |
| Blautia_A wexlerae | fixed | NA | groupFBVLED:time_pointFinal | 0.35 | 0.36 | 0.97 | 39.00 | -0.38 | 1.08 | 0.337 | 0.663 |
| CAG 110 MIC9276 | fixed | NA | groupFBVLED:time_pointFinal | -0.27 | 0.28 | -0.96 | 39.00 | -0.82 | 0.29 | 0.342 | 0.669 |
| CAG 170 sp003516765 | fixed | NA | groupFBVLED:time_pointFinal | 0.17 | 0.18 | 0.96 | 39.00 | -0.19 | 0.53 | 0.343 | 0.669 |

| OUTCOMES - SPECIES | EFFECT | GROUP | TERM | ESTIMATE | STD ERROR | STATISTIC | DF | CONF LOW | CONF HIGH | P VALUE | Q VALUE |
|---|---|---|---|---|---|---|---|---|---|---|---|
| UBA7182 MIC8257 | fixed | NA | groupFBVLED:time_pointFinal | 0.11 | 0.11 | 0.95 | 39.00 | -0.12 | 0.34 | 0.349 | 0.678 |
| Campylobacter_B hominis | fixed | NA | groupFBVLED:time_pointFinal | 0.24 | 0.26 | 0.95 | 39.00 | -0.27 | 0.76 | 0.350 | 0.678 |
| CAG 81 sp900066785 | fixed | NA | groupFBVLED:time_pointFinal | 0.16 | 0.17 | 0.95 | 39.00 | -0.19 | 0.52 | 0.350 | 0.678 |
| Streptococcus parasanguinis_B | fixed | NA | groupFBVLED:time_pointFinal | 0.19 | 0.21 | 0.92 | 78.00 | -0.22 | 0.61 | 0.360 | 0.694 |
| UBA5446 MIC7134 | fixed | NA | groupFBVLED:time_pointFinal | -0.32 | 0.35 | -0.92 | 39.00 | -1.03 | 0.38 | 0.362 | 0.695 |
| CAG 138 MIC9630 | fixed | NA | groupFBVLED:time_pointFinal | -0.47 | 0.51 | -0.92 | 39.00 | -1.51 | 0.57 | 0.362 | 0.695 |
| Oscillibacter MIC7169 | fixed | NA | groupFBVLED:time_pointFinal | 0.13 | 0.14 | 0.92 | 39.00 | -0.16 | 0.42 | 0.365 | 0.697 |
| Acetatifactor sp900066565 | fixed | NA | groupFBVLED:time_pointFinal | 0.46 | 0.50 | 0.91 | 39.00 | -0.56 | 1.47 | 0.368 | 0.699 |
| Oscillospiraceae MIC9482 | fixed | NA | groupFBVLED:time_pointFinal | 0.13 | 0.14 | 0.91 | 39.00 | -0.16 | 0.41 | 0.369 | 0.699 |
| Blautia_A sp900066355 | fixed | NA | groupFBVLED:time_pointFinal | 0.26 | 0.28 | 0.91 | 39.00 | -0.32 | 0.83 | 0.371 | 0.699 |
| CAG 110 sp003525905 | fixed | NA | groupFBVLED:time_pointFinal | -0.34 | 0.38 | -0.90 | 39.00 | -1.11 | 0.43 | 0.373 | 0.699 |
| Massiliomicrobiota sp002160815 | fixed | NA | groupFBVLED:time_pointFinal | -0.08 | 0.09 | -0.90 | 39.00 | -0.26 | 0.10 | 0.374 | 0.699 |
| Intestinimonas massiliensis | fixed | NA | groupFBVLED:time_pointFinal | -0.35 | 0.39 | -0.90 | 39.00 | -1.13 | 0.44 | 0.374 | 0.699 |
| Oscillospiraceae MIC9346 | fixed | NA | groupFBVLED:time_pointFinal | 0.11 | 0.12 | 0.90 | 39.00 | -0.13 | 0.35 | 0.376 | 0.699 |
| Clostridium MIC8573 | fixed | NA | groupFBVLED:time_pointFinal | 0.30 | 0.34 | 0.89 | 39.00 | -0.38 | 0.98 | 0.377 | 0.699 |
| Phil1 sp001940855 | fixed | NA | groupFBVLED:time_pointFinal | 0.37 | 0.42 | 0.89 | 39.00 | -0.47 | 1.21 | 0.378 | 0.699 |
| Faecalicatena torques | fixed | NA | groupFBVLED:time_pointFinal | -0.54 | 0.61 | -0.89 | 39.00 | -1.78 | 0.69 | 0.379 | 0.699 |
| Clostridium_P perfringens | fixed | NA | groupFBVLED:time_pointFinal | 0.09 | 0.10 | 0.88 | 77.84 | -0.11 | 0.29 | 0.380 | 0.699 |
| CAG 382 MIC9861 | fixed | NA | groupFBVLED:time_pointFinal | 0.06 | 0.07 | 0.89 | 39.00 | -0.08 | 0.21 | 0.380 | 0.699 |
| Negativibacillus massiliensis | fixed | NA | groupFBVLED:time_pointFinal | 0.03 | 0.04 | 0.88 | 39.00 | -0.04 | 0.10 | 0.384 | 0.702 |
| Butyricimonas virosa | fixed | NA | groupFBVLED:time_pointFinal | -0.18 | 0.21 | -0.88 | 39.00 | -0.61 | 0.24 | 0.385 | 0.702 |
| UC5 1 2E3 sp001304875 | fixed | NA | groupFBVLED:time_pointFinal | -0.06 | 0.07 | -0.88 | 39.00 | -0.21 | 0.08 | 0.386 | 0.702 |
| Blautia_A hydrogenotrophica | fixed | NA | groupFBVLED:time_pointFinal | -0.16 | 0.18 | -0.88 | 39.00 | -0.53 | 0.21 | 0.386 | 0.702 |
| CAG 45 sp000438375 | fixed | NA | groupFBVLED:time_pointFinal | 0.27 | 0.31 | 0.87 | 39.00 | -0.36 | 0.90 | 0.389 | 0.702 |
| Butyricimonas synergistica_A | fixed | NA | groupFBVLED:time_pointFinal | -0.18 | 0.21 | -0.86 | 39.00 | -0.59 | 0.24 | 0.392 | 0.702 |
| Bacteroides clarus | fixed | NA | groupFBVLED:time_pointFinal | 0.17 | 0.19 | 0.86 | 39.00 | -0.22 | 0.56 | 0.393 | 0.702 |
| Christensenellaceae MIC9015 | fixed | NA | groupFBVLED:time_pointFinal | -0.13 | 0.15 | -0.86 | 77.99 | -0.43 | 0.17 | 0.393 | 0.702 |
| Ruminococcus_C sp000437175 | fixed | NA | groupFBVLED:time_pointFinal | 0.11 | 0.13 | 0.86 | 39.00 | -0.15 | 0.37 | 0.393 | 0.702 |
| Bifidobacterium pseudocatenulatum | fixed | NA | groupFBVLED:time_pointFinal | -0.22 | 0.25 | -0.86 | 39.00 | -0.72 | 0.29 | 0.393 | 0.702 |

| OUTCOMES - SPECIES | EFFECT | GROUP | TERM | ESTIMATE | STD ERROR | STATISTIC | DF | CONF LOW | CONF HIGH | P VALUE | Q VALUE |
|---|---|---|---|---|---|---|---|---|---|---|---|
| Acutalibacter sp000435395 | fixed | NA | groupFBVLED:time_pointFinal | -0.08 | 0.10 | -0.86 | 39.00 | -0.28 | 0.11 | 0.393 | 0.702 |
| Lawsonibacter asaccharolyticus | fixed | NA | groupFBVLED:time_pointFinal | -0.19 | 0.22 | -0.86 | 39.00 | -0.64 | 0.26 | 0.395 | 0.702 |
| CAG 272 MIC9439 | fixed | NA | groupFBVLED:time_pointFinal | -0.13 | 0.16 | -0.86 | 39.00 | -0.45 | 0.18 | 0.397 | 0.704 |
| Bacteroides caccae | fixed | NA | groupFBVLED:time_pointFinal | 0.26 | 0.30 | 0.85 | 39.00 | -0.35 | 0.87 | 0.398 | 0.704 |
| Adlercreutzia equolifaciens | fixed | NA | groupFBVLED:time_pointFinal | 0.13 | 0.16 | 0.85 | 39.00 | -0.18 | 0.45 | 0.399 | 0.704 |
| UBA1255 MIC8514 | fixed | NA | groupFBVLED:time_pointFinal | -0.23 | 0.27 | -0.84 | 39.00 | -0.78 | 0.32 | 0.404 | 0.710 |
| Sutterella wadsworthensis_A | fixed | NA | groupFBVLED:time_pointFinal | -0.22 | 0.27 | -0.84 | 39.00 | -0.76 | 0.31 | 0.406 | 0.710 |
| Merdibacter MIC8457 | fixed | NA | groupFBVLED:time_pointFinal | -0.04 | 0.04 | -0.84 | 39.00 | -0.13 | 0.05 | 0.406 | 0.710 |
| CAG 56 sp900066615 | fixed | NA | groupFBVLED:time_pointFinal | 0.36 | 0.44 | 0.83 | 39.00 | -0.52 | 1.24 | 0.410 | 0.714 |
| Bacteroides uniformis | fixed | NA | groupFBVLED:time_pointFinal | -0.34 | 0.41 | -0.83 | 39.00 | -1.16 | 0.48 | 0.411 | 0.714 |
| Ruminiclostridium_C sp000435295 | fixed | NA | groupFBVLED:time_pointFinal | -0.13 | 0.16 | -0.82 | 39.00 | -0.47 | 0.20 | 0.419 | 0.726 |
| Levyella massiliensis | fixed | NA | groupFBVLED:time_pointFinal | -0.26 | 0.31 | -0.81 | 39.00 | -0.89 | 0.38 | 0.421 | 0.728 |
| Erysipelatoclostridium MIC9185 | fixed | NA | groupFBVLED:time_pointFinal | -0.09 | 0.12 | -0.80 | 39.00 | -0.33 | 0.14 | 0.428 | 0.738 |
| CAG 272 MIC7215 | fixed | NA | groupFBVLED:time_pointFinal | 0.19 | 0.24 | 0.80 | 39.00 | -0.29 | 0.67 | 0.431 | 0.738 |
| Anaerotruncus colihominis | fixed | NA | groupFBVLED:time_pointFinal | -0.13 | 0.16 | -0.80 | 39.00 | -0.45 | 0.20 | 0.431 | 0.738 |
| Bifidobacterium MIC7686 | fixed | NA | groupFBVLED:time_pointFinal | -0.10 | 0.12 | -0.79 | 39.00 | -0.35 | 0.15 | 0.432 | 0.738 |
| CAG 145 MIC9666 | fixed | NA | groupFBVLED:time_pointFinal | -0.09 | 0.11 | -0.79 | 39.00 | -0.31 | 0.14 | 0.435 | 0.739 |
| Acutalibacter MIC8741 | fixed | NA | groupFBVLED:time_pointFinal | 0.09 | 0.12 | 0.79 | 39.00 | -0.15 | 0.33 | 0.437 | 0.739 |
| Acetatifactor sp003447295 | fixed | NA | groupFBVLED:time_pointFinal | 0.18 | 0.23 | 0.78 | 39.00 | -0.29 | 0.65 | 0.437 | 0.739 |
| Dorea sp900312975 | fixed | NA | groupFBVLED:time_pointFinal | -0.20 | 0.26 | -0.78 | 39.00 | -0.73 | 0.32 | 0.439 | 0.739 |
| UBA7160 MIC6745 | fixed | NA | groupFBVLED:time_pointFinal | 0.20 | 0.25 | 0.78 | 39.00 | -0.31 | 0.71 | 0.439 | 0.739 |
| CAG 273 sp003507395 | fixed | NA | groupFBVLED:time_pointFinal | -0.19 | 0.25 | -0.78 | 39.00 | -0.69 | 0.31 | 0.439 | 0.739 |
| CAG 83 sp000431575 | fixed | NA | groupFBVLED:time_pointFinal | 0.19 | 0.24 | 0.77 | 39.00 | -0.31 | 0.68 | 0.444 | 0.742 |
| Clostridium_Q symbiosum | fixed | NA | groupFBVLED:time_pointFinal | -0.17 | 0.21 | -0.77 | 39.00 | -0.60 | 0.27 | 0.446 | 0.742 |
| Blautia_A MIC9206 | fixed | NA | groupFBVLED:time_pointFinal | -0.06 | 0.07 | -0.77 | 39.00 | -0.21 | 0.09 | 0.447 | 0.742 |
| UCG 010 MIC8371 | fixed | NA | groupFBVLED:time_pointFinal | 0.10 | 0.13 | 0.77 | 39.00 | -0.17 | 0.37 | 0.448 | 0.742 |
| Anaerococcus senegalensis | fixed | NA | groupFBVLED:time_pointFinal | 0.12 | 0.16 | 0.76 | 78.00 | -0.20 | 0.45 | 0.450 | 0.742 |
| Prevotella bivia | fixed | NA | groupFBVLED:time_pointFinal | -0.13 | 0.17 | -0.76 | 78.00 | -0.48 | 0.21 | 0.451 | 0.742 |
| Coprobacter secundus | fixed | NA | groupFBVLED:time_pointFinal | -0.14 | 0.18 | -0.76 | 39.00 | -0.50 | 0.23 | 0.452 | 0.742 |

| OUTCOMES - SPECIES | EFFECT | GROUP | TERM | ESTIMATE | STD ERROR | STATISTIC | DF | CONF LOW | CONF HIGH | P VALUE | Q VALUE |
|---|---|---|---|---|---|---|---|---|---|---|---|
| CAG 288 sp000437395 | fixed | NA | groupFBVLED:time_pointFinal | 0.19 | 0.24 | 0.76 | 39.00 | -0.31 | 0.68 | 0.452 | 0.742 |
| Fusicatenibacter saccharivorans | fixed | NA | groupFBVLED:time_pointFinal | 0.36 | 0.47 | 0.76 | 39.00 | -0.60 | 1.32 | 0.452 | 0.742 |
| Coprococcus sp900066115 | fixed | NA | groupFBVLED:time_pointFinal | 0.15 | 0.20 | 0.76 | 39.00 | -0.26 | 0.56 | 0.454 | 0.743 |
| Ruminococcaceae MIC8156 | fixed | NA | groupFBVLED:time_pointFinal | 0.07 | 0.10 | 0.75 | 39.00 | -0.12 | 0.27 | 0.456 | 0.745 |
| CAG 302 sp000431795 | fixed | NA | groupFBVLED:time_pointFinal | 0.10 | 0.13 | 0.75 | 39.00 | -0.17 | 0.37 | 0.460 | 0.748 |
| Anaeromassilibacillus sp002159845 | fixed | NA | groupFBVLED:time_pointFinal | -0.09 | 0.12 | -0.74 | 78.00 | -0.34 | 0.16 | 0.461 | 0.748 |
| Hungatella hathewayi | fixed | NA | groupFBVLED:time_pointFinal | -0.31 | 0.41 | -0.74 | 39.00 | -1.15 | 0.53 | 0.462 | 0.749 |
| Bacteroides thetaiotaomicron | fixed | NA | groupFBVLED:time_pointFinal | -0.29 | 0.40 | -0.73 | 39.00 | -1.10 | 0.51 | 0.467 | 0.749 |
| Eubacterium_I ramulus_A | fixed | NA | groupFBVLED:time_pointFinal | 0.13 | 0.18 | 0.73 | 39.00 | -0.23 | 0.50 | 0.469 | 0.749 |
| Eubacterium_R sp000436835 | fixed | NA | groupFBVLED:time_pointFinal | 0.12 | 0.16 | 0.73 | 39.00 | -0.21 | 0.44 | 0.471 | 0.749 |
| Blautia_A MIC7077 | fixed | NA | groupFBVLED:time_pointFinal | 0.23 | 0.31 | 0.73 | 39.00 | -0.40 | 0.85 | 0.473 | 0.749 |
| Coprococcus_B MIC8649 | fixed | NA | groupFBVLED:time_pointFinal | -0.02 | 0.03 | -0.72 | 39.00 | -0.08 | 0.04 | 0.474 | 0.749 |
| Hungatella_A MIC8772 | fixed | NA | groupFBVLED:time_pointFinal | 0.14 | 0.19 | 0.72 | 39.00 | -0.25 | 0.52 | 0.476 | 0.749 |
| UBA737 MIC7964 | fixed | NA | groupFBVLED:time_pointFinal | -0.02 | 0.03 | -0.72 | 39.00 | -0.09 | 0.04 | 0.476 | 0.749 |
| CAG 83 sp003539495 | fixed | NA | groupFBVLED:time_pointFinal | 0.05 | 0.07 | 0.72 | 39.00 | -0.10 | 0.20 | 0.478 | 0.749 |
| Oscillospiraceae MIC8511 | fixed | NA | groupFBVLED:time_pointFinal | 0.08 | 0.11 | 0.71 | 78.00 | -0.14 | 0.29 | 0.480 | 0.749 |
| Peptococcaceae MIC7269 | fixed | NA | groupFBVLED:time_pointFinal | 0.13 | 0.18 | 0.71 | 39.00 | -0.24 | 0.50 | 0.480 | 0.749 |
| Lachnospirales MIC9617 | fixed | NA | groupFBVLED:time_pointFinal | 0.11 | 0.15 | 0.71 | 39.00 | -0.20 | 0.42 | 0.482 | 0.749 |
| Faecalibacterium prausnitzii_K | fixed | NA | groupFBVLED:time_pointFinal | 0.25 | 0.36 | 0.71 | 39.00 | -0.47 | 0.97 | 0.483 | 0.749 |
| QALW01 sp003150515 | fixed | NA | groupFBVLED:time_pointFinal | -0.09 | 0.13 | -0.71 | 39.00 | -0.35 | 0.17 | 0.483 | 0.749 |
| Flavonifractor MIC8104 | fixed | NA | groupFBVLED:time_pointFinal | -0.18 | 0.25 | -0.70 | 78.00 | -0.68 | 0.32 | 0.483 | 0.749 |
| Fenollaria timonensis | fixed | NA | groupFBVLED:time_pointFinal | 0.18 | 0.25 | 0.71 | 39.01 | -0.33 | 0.70 | 0.483 | 0.749 |
| Anaerovoracaceae MIC7478 | fixed | NA | groupFBVLED:time_pointFinal | 0.11 | 0.16 | 0.71 | 39.00 | -0.21 | 0.43 | 0.484 | 0.749 |
| Slackia_A MIC8451 | fixed | NA | groupFBVLED:time_pointFinal | 0.02 | 0.02 | 0.71 | 39.00 | -0.03 | 0.06 | 0.485 | 0.749 |
| ER4 sp003522105 | fixed | NA | groupFBVLED:time_pointFinal | -0.21 | 0.30 | -0.70 | 39.00 | -0.81 | 0.39 | 0.486 | 0.749 |
| Bacteroides cellulosilyticus | fixed | NA | groupFBVLED:time_pointFinal | 0.27 | 0.39 | 0.70 | 39.00 | -0.51 | 1.05 | 0.486 | 0.749 |
| Gemmiger MIC8198 | fixed | NA | groupFBVLED:time_pointFinal | -0.07 | 0.10 | -0.70 | 39.00 | -0.29 | 0.14 | 0.487 | 0.749 |
| Bifidobacterium dentium | fixed | NA | groupFBVLED:time_pointFinal | -0.11 | 0.16 | -0.69 | 39.00 | -0.44 | 0.22 | 0.492 | 0.755 |
| Lawsonella clevelandensis | fixed | NA | groupFBVLED:time_pointFinal | 0.13 | 0.18 | 0.69 | 78.00 | -0.24 | 0.49 | 0.494 | 0.757 |

| OUTCOMES - SPECIES | EFFECT | GROUP | TERM | ESTIMATE | STD ERROR | STATISTIC | DF | CONF LOW | CONF HIGH | P VALUE | Q VALUE |
|---|---|---|---|---|---|---|---|---|---|---|---|
| Faecalicatena gnavus | fixed | NA | groupFBVLED:time_pointFinal | 0.18 | 0.26 | 0.68 | 39.00 | -0.35 | 0.70 | 0.498 | 0.758 |
| QAND01 MIC7514 | fixed | NA | groupFBVLED:time_pointFinal | -0.13 | 0.19 | -0.68 | 39.00 | -0.52 | 0.26 | 0.499 | 0.758 |
| Streptococcus sp001556435 | fixed | NA | groupFBVLED:time_pointFinal | 0.19 | 0.28 | 0.68 | 39.00 | -0.37 | 0.76 | 0.500 | 0.758 |
| Anaerococcus obesiensis | fixed | NA | groupFBVLED:time_pointFinal | 0.20 | 0.30 | 0.68 | 78.00 | -0.39 | 0.79 | 0.500 | 0.758 |
| Blautia_A MIC8910 | fixed | NA | groupFBVLED:time_pointFinal | -0.08 | 0.11 | -0.68 | 39.00 | -0.30 | 0.15 | 0.501 | 0.759 |
| CAG 727 MIC7825 | fixed | NA | groupFBVLED:time_pointFinal | -0.16 | 0.24 | -0.67 | 39.00 | -0.64 | 0.32 | 0.504 | 0.761 |
| QALS01 MIC9566 | fixed | NA | groupFBVLED:time_pointFinal | 0.12 | 0.19 | 0.67 | 39.00 | -0.25 | 0.50 | 0.507 | 0.762 |
| Blautia_A obeum | fixed | NA | groupFBVLED:time_pointFinal | 0.26 | 0.39 | 0.67 | 39.00 | -0.52 | 1.04 | 0.507 | 0.762 |
| CAG 433 sp000433675 | fixed | NA | groupFBVLED:time_pointFinal | -0.17 | 0.26 | -0.66 | 39.00 | -0.69 | 0.35 | 0.511 | 0.764 |
| Peptoniphilus_A harei | fixed | NA | groupFBVLED:time_pointFinal | 0.15 | 0.22 | 0.66 | 78.00 | -0.29 | 0.59 | 0.511 | 0.764 |
| UBA7160 MIC9207 | fixed | NA | groupFBVLED:time_pointFinal | 0.17 | 0.25 | 0.66 | 39.00 | -0.34 | 0.68 | 0.513 | 0.765 |
| Roseburia intestinalis | fixed | NA | groupFBVLED:time_pointFinal | 0.35 | 0.53 | 0.66 | 39.00 | -0.72 | 1.42 | 0.514 | 0.765 |
| Tidjanibacter inops | fixed | NA | groupFBVLED:time_pointFinal | -0.06 | 0.10 | -0.66 | 39.00 | -0.26 | 0.13 | 0.515 | 0.765 |
| Angelakisella MIC6791 | fixed | NA | groupFBVLED:time_pointFinal | 0.15 | 0.24 | 0.65 | 39.00 | -0.32 | 0.63 | 0.516 | 0.765 |
| QANA01 MIC9070 | fixed | NA | groupFBVLED:time_pointFinal | 0.05 | 0.08 | 0.65 | 39.00 | -0.11 | 0.21 | 0.518 | 0.765 |
| UBA9502 MIC8595 | fixed | NA | groupFBVLED:time_pointFinal | -0.09 | 0.14 | -0.65 | 78.00 | -0.38 | 0.20 | 0.521 | 0.765 |
| Eubacterium_E hallii | fixed | NA | groupFBVLED:time_pointFinal | -0.19 | 0.30 | -0.65 | 39.00 | -0.80 | 0.41 | 0.522 | 0.765 |
| Bacteroides_A MIC8078 | fixed | NA | groupFBVLED:time_pointFinal | 0.04 | 0.06 | 0.65 | 39.00 | -0.09 | 0.17 | 0.522 | 0.765 |
| CAG 103 MIC6381 | fixed | NA | groupFBVLED:time_pointFinal | 0.26 | 0.41 | 0.64 | 39.00 | -0.57 | 1.10 | 0.525 | 0.769 |
| QAMH01 MIC6543 | fixed | NA | groupFBVLED:time_pointFinal | 0.10 | 0.15 | 0.64 | 39.00 | -0.21 | 0.40 | 0.527 | 0.769 |
| CAG 196 sp002102975 | fixed | NA | groupFBVLED:time_pointFinal | 0.19 | 0.30 | 0.63 | 39.00 | -0.42 | 0.80 | 0.529 | 0.769 |
| UBA1417 MIC7387 | fixed | NA | groupFBVLED:time_pointFinal | -0.11 | 0.17 | -0.63 | 39.00 | -0.44 | 0.23 | 0.529 | 0.769 |
| TF01 11 sp000436755 | fixed | NA | groupFBVLED:time_pointFinal | 0.07 | 0.12 | 0.63 | 39.00 | -0.16 | 0.31 | 0.531 | 0.769 |
| Dialister MIC7247 | fixed | NA | groupFBVLED:time_pointFinal | 0.11 | 0.18 | 0.62 | 78.00 | -0.24 | 0.46 | 0.535 | 0.774 |
| Oscillospirales MIC7398 | fixed | NA | groupFBVLED:time_pointFinal | -0.09 | 0.14 | -0.62 | 39.00 | -0.37 | 0.19 | 0.537 | 0.775 |
| Clostridium sp000435835 | fixed | NA | groupFBVLED:time_pointFinal | 0.24 | 0.39 | 0.62 | 39.00 | -0.54 | 1.02 | 0.539 | 0.776 |
| Coprococcus eutactus | fixed | NA | groupFBVLED:time_pointFinal | -0.13 | 0.21 | -0.61 | 39.00 | -0.55 | 0.30 | 0.546 | 0.784 |
| Blautia_A MIC8288 | fixed | NA | groupFBVLED:time_pointFinal | -0.07 | 0.12 | -0.60 | 78.00 | -0.30 | 0.16 | 0.547 | 0.784 |
| Ruminococcaceae MIC7581 | fixed | NA | groupFBVLED:time_pointFinal | -0.10 | 0.17 | -0.59 | 39.00 | -0.45 | 0.24 | 0.557 | 0.796 |

| OUTCOMES - SPECIES | EFFECT | GROUP | TERM | ESTIMATE | STD ERROR | STATISTIC | DF | CONF LOW | CONF HIGH | P VALUE | Q VALUE |
|---|---|---|---|---|---|---|---|---|---|---|---|
| Collinsella MIC8447 | fixed | NA | groupFBVLED:time_pointFinal | -0.02 | 0.03 | -0.59 | 39.00 | -0.07 | 0.04 | 0.558 | 0.796 |
| CAG 74 MIC8932 | fixed | NA | groupFBVLED:time_pointFinal | -0.09 | 0.15 | -0.58 | 39.00 | -0.40 | 0.22 | 0.565 | 0.802 |
| Faecalicatena sp000509105 | fixed | NA | groupFBVLED:time_pointFinal | -0.12 | 0.21 | -0.58 | 39.00 | -0.56 | 0.31 | 0.566 | 0.802 |
| Holdemania filiformis | fixed | NA | groupFBVLED:time_pointFinal | -0.06 | 0.10 | -0.58 | 39.00 | -0.26 | 0.14 | 0.566 | 0.802 |
| Finegoldia magna_H | fixed | NA | groupFBVLED:time_pointFinal | 0.19 | 0.34 | 0.58 | 39.00 | -0.49 | 0.87 | 0.568 | 0.803 |
| Prevotella corporis | fixed | NA | groupFBVLED:time_pointFinal | 0.20 | 0.36 | 0.57 | 39.00 | -0.52 | 0.92 | 0.572 | 0.807 |
| CAG 273 sp000438355 | fixed | NA | groupFBVLED:time_pointFinal | -0.15 | 0.26 | -0.57 | 39.00 | -0.67 | 0.38 | 0.574 | 0.807 |
| Streptococcus salivarius | fixed | NA | groupFBVLED:time_pointFinal | 0.19 | 0.34 | 0.56 | 39.00 | -0.50 | 0.88 | 0.577 | 0.809 |
| Marvinbryantia MIC9792 | fixed | NA | groupFBVLED:time_pointFinal | -0.05 | 0.09 | -0.56 | 39.00 | -0.24 | 0.13 | 0.580 | 0.812 |
| Streptococcus vestibularis | fixed | NA | groupFBVLED:time_pointFinal | 0.11 | 0.20 | 0.55 | 78.00 | -0.29 | 0.52 | 0.582 | 0.813 |
| Erysipelatoclostridium spiroforme | fixed | NA | groupFBVLED:time_pointFinal | -0.13 | 0.23 | -0.55 | 39.00 | -0.60 | 0.34 | 0.584 | 0.815 |
| Paraprevotella clara | fixed | NA | groupFBVLED:time_pointFinal | 0.09 | 0.16 | 0.54 | 39.00 | -0.23 | 0.40 | 0.590 | 0.816 |
| CAG 1427 sp000436075 | fixed | NA | groupFBVLED:time_pointFinal | 0.06 | 0.11 | 0.54 | 39.00 | -0.16 | 0.28 | 0.591 | 0.816 |
| Escherichia coli | fixed | NA | groupFBVLED:time_pointFinal | 0.23 | 0.42 | 0.54 | 39.00 | -0.62 | 1.08 | 0.591 | 0.816 |
| CAG 115 sp003531585 | fixed | NA | groupFBVLED:time_pointFinal | 0.22 | 0.42 | 0.54 | 39.00 | -0.62 | 1.06 | 0.594 | 0.816 |
| Faecalibacterium prausnitzii_A | fixed | NA | groupFBVLED:time_pointFinal | -0.10 | 0.19 | -0.54 | 39.00 | -0.49 | 0.29 | 0.594 | 0.816 |
| Anaerotignum sp000436415 | fixed | NA | groupFBVLED:time_pointFinal | -0.11 | 0.20 | -0.53 | 39.00 | -0.51 | 0.30 | 0.598 | 0.816 |
| Streptococcus anginosus | fixed | NA | groupFBVLED:time_pointFinal | 0.16 | 0.30 | 0.53 | 39.00 | -0.45 | 0.78 | 0.599 | 0.816 |
| Blautia_A sp000285855 | fixed | NA | groupFBVLED:time_pointFinal | 0.19 | 0.36 | 0.53 | 39.00 | -0.54 | 0.92 | 0.599 | 0.816 |
| Prevotella MIC8201 | fixed | NA | groupFBVLED:time_pointFinal | 0.11 | 0.20 | 0.53 | 39.00 | -0.30 | 0.52 | 0.600 | 0.816 |
| Ruminococcus_C sp000433635 | fixed | NA | groupFBVLED:time_pointFinal | 0.21 | 0.40 | 0.53 | 39.00 | -0.60 | 1.02 | 0.600 | 0.816 |
| Erysipelatoclostridium sp003024675 | fixed | NA | groupFBVLED:time_pointFinal | 0.04 | 0.07 | 0.53 | 39.00 | -0.11 | 0.19 | 0.600 | 0.816 |
| CAG 353 sp900066885 | fixed | NA | groupFBVLED:time_pointFinal | -0.14 | 0.27 | -0.53 | 39.00 | -0.70 | 0.41 | 0.601 | 0.816 |
| UBA3818 MIC8425 | fixed | NA | groupFBVLED:time_pointFinal | -0.13 | 0.26 | -0.53 | 39.00 | -0.65 | 0.38 | 0.602 | 0.816 |
| Eubacterium_G sp000432355 | fixed | NA | groupFBVLED:time_pointFinal | -0.10 | 0.19 | -0.52 | 39.00 | -0.48 | 0.28 | 0.604 | 0.816 |
| CAG 83 MIC8701 | fixed | NA | groupFBVLED:time_pointFinal | -0.09 | 0.17 | -0.52 | 39.00 | -0.43 | 0.25 | 0.607 | 0.818 |
| Blautia_A MIC8050 | fixed | NA | groupFBVLED:time_pointFinal | -0.19 | 0.37 | -0.51 | 39.00 | -0.93 | 0.55 | 0.611 | 0.820 |
| Streptococcus gordonii | fixed | NA | groupFBVLED:time_pointFinal | 0.07 | 0.13 | 0.51 | 78.00 | -0.19 | 0.32 | 0.612 | 0.820 |
| GCA 900066495 MIC8689 | fixed | NA | groupFBVLED:time_pointFinal | -0.08 | 0.16 | -0.51 | 39.00 | -0.41 | 0.24 | 0.613 | 0.820 |

| OUTCOMES - SPECIES | EFFECT | GROUP | TERM | ESTIMATE | STD ERROR | STATISTIC | DF | CONF LOW | CONF HIGH | P VALUE | Q VALUE |
|---|---|---|---|---|---|---|---|---|---|---|---|
| S5 A14a MIC8656 | fixed | NA | groupFBVLED:time_pointFinal | 0.14 | 0.29 | 0.51 | 78.00 | -0.42 | 0.71 | 0.614 | 0.820 |
| Agathobacter MIC6414 | fixed | NA | groupFBVLED:time_pointFinal | 0.07 | 0.13 | 0.50 | 39.00 | -0.20 | 0.34 | 0.621 | 0.828 |
| UBA4285 MIC9245 | fixed | NA | groupFBVLED:time_pointFinal | -0.07 | 0.14 | -0.50 | 39.00 | -0.36 | 0.22 | 0.623 | 0.829 |
| Peptostreptococcus anaerobius | fixed | NA | groupFBVLED:time_pointFinal | 0.06 | 0.12 | 0.49 | 39.00 | -0.18 | 0.30 | 0.624 | 0.829 |
| CAG 312 MIC7072 | fixed | NA | groupFBVLED:time_pointFinal | 0.08 | 0.17 | 0.49 | 39.00 | -0.27 | 0.43 | 0.630 | 0.833 |
| QALW01 MIC7388 | fixed | NA | groupFBVLED:time_pointFinal | 0.04 | 0.08 | 0.48 | 78.00 | -0.12 | 0.20 | 0.630 | 0.833 |
| Fenollaria massiliensis | fixed | NA | groupFBVLED:time_pointFinal | 0.12 | 0.26 | 0.48 | 78.00 | -0.40 | 0.65 | 0.635 | 0.838 |
| Bacteroides_B dorei | fixed | NA | groupFBVLED:time_pointFinal | -0.16 | 0.33 | -0.47 | 39.00 | -0.83 | 0.51 | 0.638 | 0.840 |
| Escherichia coli_D | fixed | NA | groupFBVLED:time_pointFinal | 0.11 | 0.23 | 0.47 | 39.00 | -0.35 | 0.56 | 0.640 | 0.841 |
| CAG 269 MIC8518 | fixed | NA | groupFBVLED:time_pointFinal | 0.07 | 0.15 | 0.47 | 39.00 | -0.23 | 0.37 | 0.642 | 0.841 |
| Peptoniphilus_C coxii | fixed | NA | groupFBVLED:time_pointFinal | -0.07 | 0.14 | -0.47 | 39.00 | -0.35 | 0.22 | 0.642 | 0.841 |
| Coprococcus eutactus_A | fixed | NA | groupFBVLED:time_pointFinal | 0.21 | 0.44 | 0.46 | 39.00 | -0.69 | 1.10 | 0.645 | 0.842 |
| Acetatifactor sp900066365 | fixed | NA | groupFBVLED:time_pointFinal | -0.21 | 0.47 | -0.46 | 39.00 | -1.16 | 0.73 | 0.648 | 0.844 |
| CAG 83 MIC9166 | fixed | NA | groupFBVLED:time_pointFinal | -0.02 | 0.04 | -0.45 | 39.00 | -0.10 | 0.07 | 0.652 | 0.848 |
| Eubacterium_I ramulus | fixed | NA | groupFBVLED:time_pointFinal | -0.12 | 0.27 | -0.44 | 39.00 | -0.67 | 0.43 | 0.660 | 0.854 |
| CAG 273 sp003534295 | fixed | NA | groupFBVLED:time_pointFinal | 0.16 | 0.37 | 0.44 | 39.00 | -0.59 | 0.92 | 0.661 | 0.854 |
| CAG 170 MIC8868 | fixed | NA | groupFBVLED:time_pointFinal | -0.09 | 0.21 | -0.44 | 78.00 | -0.51 | 0.33 | 0.663 | 0.854 |
| Lawsonibacter sp900066825 | fixed | NA | groupFBVLED:time_pointFinal | -0.05 | 0.10 | -0.44 | 39.00 | -0.25 | 0.16 | 0.664 | 0.854 |
| CAG 354 MIC8844 | fixed | NA | groupFBVLED:time_pointFinal | 0.08 | 0.17 | 0.44 | 39.00 | -0.27 | 0.43 | 0.664 | 0.854 |
| Ruminiclostridium_C MIC7261 | fixed | NA | groupFBVLED:time_pointFinal | 0.05 | 0.10 | 0.43 | 39.00 | -0.17 | 0.26 | 0.667 | 0.857 |
| Clostridium_M sp001517625 | fixed | NA | groupFBVLED:time_pointFinal | 0.09 | 0.21 | 0.43 | 39.00 | -0.34 | 0.52 | 0.670 | 0.858 |
| QALS01 sp003150575 | fixed | NA | groupFBVLED:time_pointFinal | -0.12 | 0.29 | -0.42 | 39.00 | -0.71 | 0.46 | 0.675 | 0.861 |
| Agathobaculum MIC7900 | fixed | NA | groupFBVLED:time_pointFinal | 0.07 | 0.17 | 0.42 | 39.00 | -0.28 | 0.43 | 0.675 | 0.861 |
| An200 MIC7965 | fixed | NA | groupFBVLED:time_pointFinal | -0.07 | 0.16 | -0.42 | 39.00 | -0.39 | 0.25 | 0.678 | 0.864 |
| Bacteroides ovatus | fixed | NA | groupFBVLED:time_pointFinal | -0.21 | 0.50 | -0.41 | 39.00 | -1.22 | 0.81 | 0.681 | 0.866 |
| Blautia_A obeum_B | fixed | NA | groupFBVLED:time_pointFinal | 0.03 | 0.09 | 0.41 | 39.00 | -0.14 | 0.21 | 0.686 | 0.871 |
| Monoglobus pectinilyticus | fixed | NA | groupFBVLED:time_pointFinal | 0.11 | 0.27 | 0.40 | 39.00 | -0.43 | 0.65 | 0.689 | 0.871 |
| Lactobacillus_B ruminis | fixed | NA | groupFBVLED:time_pointFinal | 0.03 | 0.08 | 0.40 | 39.00 | -0.12 | 0.19 | 0.690 | 0.871 |
| Prevotella timonensis | fixed | NA | groupFBVLED:time_pointFinal | -0.08 | 0.20 | -0.40 | 78.00 | -0.47 | 0.31 | 0.692 | 0.871 |

| OUTCOMES - SPECIES | EFFECT | GROUP | TERM | ESTIMATE | STD ERROR | STATISTIC | DF | CONF LOW | CONF HIGH | P VALUE | Q VALUE |
|---|---|---|---|---|---|---|---|---|---|---|---|
| Blautia_A sp900066335 | fixed | NA | groupFBVLED:time_pointFinal | -0.08 | 0.19 | -0.40 | 39.00 | -0.47 | 0.31 | 0.693 | 0.871 |
| Mailhella MIC8103 | fixed | NA | groupFBVLED:time_pointFinal | -0.06 | 0.15 | -0.40 | 39.00 | -0.36 | 0.24 | 0.694 | 0.871 |
| CAG 83 MIC8843 | fixed | NA | groupFBVLED:time_pointFinal | -0.03 | 0.09 | -0.39 | 39.00 | -0.21 | 0.14 | 0.697 | 0.873 |
| Alistipes onderdonkii | fixed | NA | groupFBVLED:time_pointFinal | -0.18 | 0.47 | -0.39 | 39.00 | -1.14 | 0.78 | 0.699 | 0.874 |
| Butyricicoccus_A sp002395695 | fixed | NA | groupFBVLED:time_pointFinal | 0.07 | 0.18 | 0.39 | 39.00 | -0.30 | 0.44 | 0.700 | 0.874 |
| Enorma MIC9272 | fixed | NA | groupFBVLED:time_pointFinal | -0.02 | 0.04 | -0.39 | 39.00 | -0.11 | 0.07 | 0.701 | 0.874 |
| Bacteroides intestinalis | fixed | NA | groupFBVLED:time_pointFinal | 0.07 | 0.18 | 0.38 | 39.00 | -0.30 | 0.43 | 0.707 | 0.878 |
| Bifidobacterium vaginale | fixed | NA | groupFBVLED:time_pointFinal | 0.09 | 0.23 | 0.37 | 78.00 | -0.38 | 0.55 | 0.714 | 0.883 |
| CAG 170 MIC6396 | fixed | NA | groupFBVLED:time_pointFinal | 0.09 | 0.23 | 0.37 | 39.00 | -0.38 | 0.56 | 0.715 | 0.883 |
| KLE1615 sp900066985 | fixed | NA | groupFBVLED:time_pointFinal | 0.12 | 0.33 | 0.37 | 39.00 | -0.54 | 0.78 | 0.715 | 0.883 |
| UBA644 MIC9235 | fixed | NA | groupFBVLED:time_pointFinal | -0.08 | 0.23 | -0.37 | 39.00 | -0.54 | 0.38 | 0.717 | 0.883 |
| Methanobrevibacter_A smithii | fixed | NA | groupFBVLED:time_pointFinal | 0.18 | 0.50 | 0.36 | 39.00 | -0.83 | 1.20 | 0.718 | 0.883 |
| Bacteroides eggerthii | fixed | NA | groupFBVLED:time_pointFinal | 0.10 | 0.27 | 0.36 | 39.00 | -0.45 | 0.65 | 0.719 | 0.883 |
| Phascolarctobacterium_A succinatutens | fixed | NA | groupFBVLED:time_pointFinal | 0.02 | 0.05 | 0.36 | 39.00 | -0.09 | 0.12 | 0.723 | 0.886 |
| Ruminococcus_C callidus | fixed | NA | groupFBVLED:time_pointFinal | 0.13 | 0.38 | 0.35 | 39.00 | -0.63 | 0.89 | 0.728 | 0.891 |
| UBA7102 MIC7325 | fixed | NA | groupFBVLED:time_pointFinal | 0.03 | 0.09 | 0.34 | 39.00 | -0.15 | 0.22 | 0.733 | 0.895 |
| Prevotella copri | fixed | NA | groupFBVLED:time_pointFinal | 0.09 | 0.27 | 0.34 | 39.00 | -0.45 | 0.63 | 0.736 | 0.896 |
| Collinsella sp003487125 | fixed | NA | groupFBVLED:time_pointFinal | -0.05 | 0.16 | -0.33 | 39.00 | -0.37 | 0.27 | 0.739 | 0.899 |
| Faecalibacterium MIC9210 | fixed | NA | groupFBVLED:time_pointFinal | -0.12 | 0.37 | -0.33 | 39.00 | -0.86 | 0.62 | 0.745 | 0.904 |
| Lachnospiraceae MIC7157 | fixed | NA | groupFBVLED:time_pointFinal | -0.04 | 0.14 | -0.31 | 39.00 | -0.32 | 0.23 | 0.755 | 0.914 |
| UBA644_A MIC9239 | fixed | NA | groupFBVLED:time_pointFinal | 0.04 | 0.12 | 0.31 | 39.00 | -0.21 | 0.28 | 0.759 | 0.916 |
| Dorea longicatena_B | fixed | NA | groupFBVLED:time_pointFinal | 0.07 | 0.23 | 0.31 | 39.00 | -0.40 | 0.54 | 0.759 | 0.916 |
| Faecalicatena faecis | fixed | NA | groupFBVLED:time_pointFinal | -0.09 | 0.30 | -0.31 | 39.00 | -0.71 | 0.52 | 0.761 | 0.917 |
| Intestinibacter MIC7152 | fixed | NA | groupFBVLED:time_pointFinal | -0.03 | 0.11 | -0.30 | 78.00 | -0.25 | 0.18 | 0.767 | 0.921 |
| Eubacterium_G sp000435815 | fixed | NA | groupFBVLED:time_pointFinal | 0.04 | 0.14 | 0.29 | 39.00 | -0.24 | 0.33 | 0.770 | 0.921 |
| Turicibacter sanguinis | fixed | NA | groupFBVLED:time_pointFinal | 0.07 | 0.25 | 0.29 | 39.00 | -0.43 | 0.57 | 0.771 | 0.921 |
| Faecalicatena sp900066545 | fixed | NA | groupFBVLED:time_pointFinal | -0.05 | 0.16 | -0.29 | 39.00 | -0.36 | 0.27 | 0.772 | 0.921 |
| Dorea sp000433535 | fixed | NA | groupFBVLED:time_pointFinal | 0.04 | 0.14 | 0.29 | 39.00 | -0.24 | 0.32 | 0.772 | 0.921 |
| Bacteroides_B massiliensis | fixed | NA | groupFBVLED:time_pointFinal | -0.07 | 0.25 | -0.29 | 39.00 | -0.58 | 0.44 | 0.774 | 0.922 |

| OUTCOMES - SPECIES | EFFECT | GROUP | TERM | ESTIMATE | STD ERROR | STATISTIC | DF | CONF LOW | CONF HIGH | P VALUE | Q VALUE |
|---|---|---|---|---|---|---|---|---|---|---|---|
| Oscillibacter MIC6596 | fixed | NA | groupFBVLED:time_pointFinal | 0.02 | 0.06 | 0.29 | 39.00 | -0.10 | 0.13 | 0.777 | 0.922 |
| Pauljensenia sp000466265 | fixed | NA | groupFBVLED:time_pointFinal | -0.05 | 0.16 | -0.28 | 39.00 | -0.37 | 0.28 | 0.777 | 0.922 |
| Lawsonibacter MIC8046 | fixed | NA | groupFBVLED:time_pointFinal | -0.05 | 0.17 | -0.28 | 39.00 | -0.39 | 0.29 | 0.784 | 0.927 |
| Phascolarctobacterium sp000436095 | fixed | NA | groupFBVLED:time_pointFinal | 0.03 | 0.12 | 0.27 | 39.00 | -0.21 | 0.27 | 0.785 | 0.927 |
| Blautia_A massiliensis | fixed | NA | groupFBVLED:time_pointFinal | 0.11 | 0.41 | 0.27 | 39.00 | -0.72 | 0.95 | 0.787 | 0.927 |
| CAG 74 MIC7629 | fixed | NA | groupFBVLED:time_pointFinal | -0.07 | 0.28 | -0.27 | 39.00 | -0.63 | 0.48 | 0.790 | 0.928 |
| CAG 180 sp000432435 | fixed | NA | groupFBVLED:time_pointFinal | 0.09 | 0.35 | 0.27 | 39.00 | -0.62 | 0.81 | 0.790 | 0.928 |
| Alistipes_A ihumii | fixed | NA | groupFBVLED:time_pointFinal | -0.08 | 0.31 | -0.25 | 39.00 | -0.71 | 0.55 | 0.802 | 0.937 |
| Absiella innocuum | fixed | NA | groupFBVLED:time_pointFinal | 0.07 | 0.30 | 0.25 | 78.00 | -0.52 | 0.66 | 0.804 | 0.937 |
| Catenibacterium sp000437715 | fixed | NA | groupFBVLED:time_pointFinal | -0.05 | 0.19 | -0.25 | 39.00 | -0.44 | 0.34 | 0.805 | 0.937 |
| Ruminiclostridium_E siraeum | fixed | NA | groupFBVLED:time_pointFinal | 0.12 | 0.49 | 0.25 | 39.00 | -0.87 | 1.10 | 0.807 | 0.937 |
| UBA5026 MIC9780 | fixed | NA | groupFBVLED:time_pointFinal | 0.02 | 0.09 | 0.24 | 39.00 | -0.16 | 0.21 | 0.809 | 0.937 |
| Bilophila wadsworthia | fixed | NA | groupFBVLED:time_pointFinal | 0.08 | 0.32 | 0.24 | 39.00 | -0.58 | 0.74 | 0.810 | 0.937 |
| Clostridium_M MIC9612 | fixed | NA | groupFBVLED:time_pointFinal | 0.05 | 0.22 | 0.24 | 39.00 | -0.39 | 0.50 | 0.810 | 0.937 |
| S5 A14a sp000758905 | fixed | NA | groupFBVLED:time_pointFinal | -0.04 | 0.18 | -0.24 | 78.00 | -0.40 | 0.32 | 0.813 | 0.937 |
| Lactobacillus crispatus | fixed | NA | groupFBVLED:time_pointFinal | 0.05 | 0.20 | 0.24 | 39.00 | -0.36 | 0.45 | 0.814 | 0.937 |
| Holdemanella MIC8202 | fixed | NA | groupFBVLED:time_pointFinal | 0.02 | 0.08 | 0.24 | 39.00 | -0.14 | 0.17 | 0.814 | 0.937 |
| Acutalibacteraceae MIC6974 | fixed | NA | groupFBVLED:time_pointFinal | 0.06 | 0.27 | 0.24 | 39.00 | -0.47 | 0.60 | 0.815 | 0.937 |
| CAG 245 sp000435175 | fixed | NA | groupFBVLED:time_pointFinal | 0.06 | 0.27 | 0.23 | 39.00 | -0.48 | 0.61 | 0.816 | 0.937 |
| Roseburia hominis | fixed | NA | groupFBVLED:time_pointFinal | 0.09 | 0.38 | 0.23 | 39.00 | -0.67 | 0.85 | 0.820 | 0.939 |
| Eubacterium_G sp000434315 | fixed | NA | groupFBVLED:time_pointFinal | 0.06 | 0.26 | 0.23 | 39.00 | -0.46 | 0.58 | 0.821 | 0.939 |
| CAG 273 sp000435755 | fixed | NA | groupFBVLED:time_pointFinal | 0.03 | 0.15 | 0.23 | 39.00 | -0.26 | 0.33 | 0.822 | 0.939 |
| CAG 302 sp001916775 | fixed | NA | groupFBVLED:time_pointFinal | 0.06 | 0.27 | 0.22 | 39.00 | -0.48 | 0.60 | 0.824 | 0.940 |
| CAG 74 MIC9091 | fixed | NA | groupFBVLED:time_pointFinal | -0.03 | 0.14 | -0.22 | 39.00 | -0.32 | 0.25 | 0.830 | 0.942 |
| Intestinibacter MIC8174 | fixed | NA | groupFBVLED:time_pointFinal | -0.06 | 0.29 | -0.22 | 39.00 | -0.66 | 0.53 | 0.831 | 0.942 |
| Phascolarctobacterium faecium | fixed | NA | groupFBVLED:time_pointFinal | 0.04 | 0.20 | 0.22 | 39.00 | -0.36 | 0.45 | 0.831 | 0.942 |
| Blautia_A sp000433815 | fixed | NA | groupFBVLED:time_pointFinal | 0.06 | 0.30 | 0.21 | 39.00 | -0.54 | 0.66 | 0.833 | 0.942 |
| Parabacteroides merdae | fixed | NA | groupFBVLED:time_pointFinal | 0.04 | 0.21 | 0.21 | 39.00 | -0.39 | 0.48 | 0.834 | 0.943 |
| Bacteroides MIC8726 | fixed | NA | groupFBVLED:time_pointFinal | 0.04 | 0.19 | 0.20 | 39.00 | -0.34 | 0.41 | 0.841 | 0.948 |

| OUTCOMES - SPECIES | EFFECT | GROUP | TERM | ESTIMATE | STD ERROR | STATISTIC | DF | CONF LOW | CONF HIGH | P VALUE | Q VALUE |
|---|---|---|---|---|---|---|---|---|---|---|---|
| Lachnospirales MIC8074 | fixed | NA | groupFBVLED:time_pointFinal | -0.02 | 0.12 | -0.20 | 39.00 | -0.27 | 0.22 | 0.843 | 0.948 |
| Alistipes senegalensis | fixed | NA | groupFBVLED:time_pointFinal | -0.02 | 0.13 | -0.19 | 39.00 | -0.28 | 0.23 | 0.849 | 0.953 |
| CAG 110 sp000434635 | fixed | NA | groupFBVLED:time_pointFinal | -0.05 | 0.27 | -0.19 | 39.00 | -0.61 | 0.50 | 0.851 | 0.953 |
| Porphyromonas MIC9772 | fixed | NA | groupFBVLED:time_pointFinal | -0.04 | 0.23 | -0.19 | 78.00 | -0.50 | 0.41 | 0.852 | 0.953 |
| TF01 11 sp001414325 | fixed | NA | groupFBVLED:time_pointFinal | -0.04 | 0.24 | -0.19 | 39.00 | -0.54 | 0.45 | 0.854 | 0.954 |
| Gemmiger formicilis | fixed | NA | groupFBVLED:time_pointFinal | -0.05 | 0.26 | -0.18 | 39.00 | -0.57 | 0.48 | 0.857 | 0.956 |
| Bacteroides congonensis | fixed | NA | groupFBVLED:time_pointFinal | -0.03 | 0.18 | -0.17 | 39.00 | -0.39 | 0.33 | 0.867 | 0.965 |
| Collinsella aerofaciens_F | fixed | NA | groupFBVLED:time_pointFinal | -0.03 | 0.20 | -0.17 | 39.00 | -0.44 | 0.38 | 0.869 | 0.965 |
| Prevotella disiens | fixed | NA | groupFBVLED:time_pointFinal | 0.03 | 0.17 | 0.16 | 39.00 | -0.32 | 0.37 | 0.870 | 0.965 |
| Coprobacillus cateniformis | fixed | NA | groupFBVLED:time_pointFinal | -0.05 | 0.33 | -0.16 | 39.00 | -0.72 | 0.61 | 0.872 | 0.966 |
| Clostridium_M bolteae | fixed | NA | groupFBVLED:time_pointFinal | 0.04 | 0.27 | 0.16 | 39.00 | -0.51 | 0.59 | 0.876 | 0.966 |
| Dorea sp900066765 | fixed | NA | groupFBVLED:time_pointFinal | -0.02 | 0.12 | -0.15 | 39.00 | -0.27 | 0.23 | 0.878 | 0.966 |
| Akkermansia sp001580195 | fixed | NA | groupFBVLED:time_pointFinal | -0.04 | 0.25 | -0.15 | 39.00 | -0.54 | 0.46 | 0.879 | 0.966 |
| Blastocystis sp subtype 3 | fixed | NA | groupFBVLED:time_pointFinal | 0.01 | 0.07 | 0.15 | 39.00 | -0.14 | 0.16 | 0.881 | 0.966 |
| CAG 83 MIC8731 | fixed | NA | groupFBVLED:time_pointFinal | 0.03 | 0.21 | 0.15 | 39.00 | -0.39 | 0.45 | 0.884 | 0.966 |
| Coprococcus_A MIC9199 | fixed | NA | groupFBVLED:time_pointFinal | 0.03 | 0.21 | 0.15 | 39.00 | -0.40 | 0.47 | 0.885 | 0.966 |
| Holdemanella sp002299315 | fixed | NA | groupFBVLED:time_pointFinal | -0.03 | 0.17 | -0.14 | 39.00 | -0.38 | 0.33 | 0.886 | 0.966 |
| UBA1685 sp002320595 | fixed | NA | groupFBVLED:time_pointFinal | 0.03 | 0.24 | 0.14 | 39.00 | -0.46 | 0.53 | 0.889 | 0.966 |
| ER4 sp900317525 | fixed | NA | groupFBVLED:time_pointFinal | 0.05 | 0.33 | 0.14 | 39.00 | -0.62 | 0.71 | 0.891 | 0.966 |
| Sellimonas intestinalis | fixed | NA | groupFBVLED:time_pointFinal | 0.03 | 0.19 | 0.14 | 39.00 | -0.36 | 0.41 | 0.892 | 0.966 |
| Lachnospira sp000436475 | fixed | NA | groupFBVLED:time_pointFinal | -0.02 | 0.16 | -0.14 | 39.00 | -0.34 | 0.30 | 0.893 | 0.966 |
| Collinsella sp002232035 | fixed | NA | groupFBVLED:time_pointFinal | 0.02 | 0.15 | 0.13 | 39.00 | -0.28 | 0.31 | 0.894 | 0.966 |
| CAG 74 MIC7845 | fixed | NA | groupFBVLED:time_pointFinal | 0.01 | 0.11 | 0.13 | 78.00 | -0.20 | 0.23 | 0.895 | 0.966 |
| Bacteroides_A coprocola | fixed | NA | groupFBVLED:time_pointFinal | 0.03 | 0.20 | 0.13 | 39.00 | -0.37 | 0.42 | 0.898 | 0.966 |
| Lachnospira sp003451515 | fixed | NA | groupFBVLED:time_pointFinal | -0.03 | 0.22 | -0.13 | 39.00 | -0.48 | 0.42 | 0.899 | 0.966 |
| QALS01 MIC6548 | fixed | NA | groupFBVLED:time_pointFinal | -0.03 | 0.20 | -0.13 | 39.00 | -0.43 | 0.38 | 0.899 | 0.966 |
| Porphyromonas MIC7597 | fixed | NA | groupFBVLED:time_pointFinal | -0.01 | 0.06 | -0.12 | 78.00 | -0.13 | 0.12 | 0.901 | 0.966 |
| Blautia_A MIC6897 | fixed | NA | groupFBVLED:time_pointFinal | 0.02 | 0.17 | 0.12 | 39.00 | -0.32 | 0.36 | 0.903 | 0.966 |
| Blautia_A sp900066205 | fixed | NA | groupFBVLED:time_pointFinal | -0.04 | 0.30 | -0.12 | 39.00 | -0.65 | 0.58 | 0.904 | 0.966 |

| OUTCOMES - SPECIES | EFFECT | GROUP | TERM | ESTIMATE | STD ERROR | STATISTIC | DF | CONF LOW | CONF HIGH | P VALUE | Q VALUE |
|---|---|---|---|---|---|---|---|---|---|---|---|
| Eggerthellaceae MIC6427 | fixed | NA | groupFBVLED:time_pointFinal | 0.00 | 0.04 | -0.12 | 39.00 | -0.08 | 0.07 | 0.905 | 0.966 |
| Merdibacter MIC6626 | fixed | NA | groupFBVLED:time_pointFinal | 0.01 | 0.10 | 0.12 | 39.00 | -0.20 | 0.22 | 0.906 | 0.966 |
| Bacteroides stercoris | fixed | NA | groupFBVLED:time_pointFinal | 0.03 | 0.25 | 0.12 | 39.00 | -0.47 | 0.53 | 0.907 | 0.966 |
| Butyricicoccaceae MIC8222 | fixed | NA | groupFBVLED:time_pointFinal | -0.01 | 0.09 | -0.11 | 39.00 | -0.19 | 0.17 | 0.915 | 0.971 |
| Anaerococcus MIC7735 | fixed | NA | groupFBVLED:time_pointFinal | 0.02 | 0.20 | 0.11 | 78.00 | -0.37 | 0.41 | 0.916 | 0.971 |
| Pauljensenia turicensis | fixed | NA | groupFBVLED:time_pointFinal | 0.03 | 0.26 | 0.11 | 39.00 | -0.49 | 0.55 | 0.917 | 0.971 |
| Porphyromonas MIC7959 | fixed | NA | groupFBVLED:time_pointFinal | 0.03 | 0.27 | 0.10 | 78.00 | -0.51 | 0.56 | 0.923 | 0.971 |
| Clostridium saudiense | fixed | NA | groupFBVLED:time_pointFinal | 0.04 | 0.39 | 0.10 | 39.00 | -0.75 | 0.82 | 0.923 | 0.971 |
| Alistipes MIC9770 | fixed | NA | groupFBVLED:time_pointFinal | 0.02 | 0.17 | 0.10 | 39.00 | -0.32 | 0.35 | 0.923 | 0.971 |
| UBA1691 MIC9213 | fixed | NA | groupFBVLED:time_pointFinal | -0.02 | 0.18 | -0.10 | 39.00 | -0.38 | 0.35 | 0.924 | 0.971 |
| Tyzzerella nexilis | fixed | NA | groupFBVLED:time_pointFinal | -0.04 | 0.41 | -0.10 | 39.00 | -0.86 | 0.78 | 0.924 | 0.971 |
| UBA1191 MIC6632 | fixed | NA | groupFBVLED:time_pointFinal | 0.01 | 0.15 | 0.09 | 39.00 | -0.29 | 0.32 | 0.928 | 0.974 |
| CAG 127 sp900319515 | fixed | NA | groupFBVLED:time_pointFinal | -0.03 | 0.38 | -0.08 | 39.00 | -0.79 | 0.73 | 0.934 | 0.978 |
| Faecalibacterium prausnitzii_H | fixed | NA | groupFBVLED:time_pointFinal | 0.01 | 0.11 | 0.07 | 39.00 | -0.21 | 0.23 | 0.942 | 0.983 |
| Ruminococcaceae MIC8509 | fixed | NA | groupFBVLED:time_pointFinal | 0.01 | 0.09 | 0.07 | 39.00 | -0.17 | 0.18 | 0.942 | 0.983 |
| Corynebacterium sp001767255 | fixed | NA | groupFBVLED:time_pointFinal | 0.01 | 0.08 | 0.07 | 78.00 | -0.16 | 0.17 | 0.945 | 0.983 |
| TF01 11 sp003524945 | fixed | NA | groupFBVLED:time_pointFinal | -0.01 | 0.17 | -0.07 | 39.00 | -0.37 | 0.34 | 0.946 | 0.983 |
| UBA1191 sp900066305 | fixed | NA | groupFBVLED:time_pointFinal | -0.01 | 0.13 | -0.07 | 39.00 | -0.28 | 0.26 | 0.946 | 0.983 |
| Blautia_A sp900120195 | fixed | NA | groupFBVLED:time_pointFinal | 0.02 | 0.24 | 0.07 | 39.00 | -0.47 | 0.50 | 0.948 | 0.983 |
| Absiella sp000165065 | fixed | NA | groupFBVLED:time_pointFinal | 0.01 | 0.21 | 0.06 | 39.00 | -0.42 | 0.44 | 0.954 | 0.986 |
| CAG 170 sp000432135 | fixed | NA | groupFBVLED:time_pointFinal | 0.03 | 0.45 | 0.06 | 39.00 | -0.89 | 0.94 | 0.956 | 0.986 |
| Faecalibacterium prausnitzii_E | fixed | NA | groupFBVLED:time_pointFinal | 0.01 | 0.14 | 0.05 | 39.00 | -0.28 | 0.29 | 0.957 | 0.986 |
| UBA10677 MIC8686 | fixed | NA | groupFBVLED:time_pointFinal | 0.00 | 0.08 | 0.05 | 39.00 | -0.15 | 0.16 | 0.959 | 0.986 |
| UBA1191 MIC6696 | fixed | NA | groupFBVLED:time_pointFinal | -0.01 | 0.22 | -0.05 | 39.00 | -0.46 | 0.44 | 0.960 | 0.986 |
| Anaerovoracaceae MIC8502 | fixed | NA | groupFBVLED:time_pointFinal | 0.01 | 0.15 | 0.05 | 39.00 | -0.30 | 0.32 | 0.961 | 0.986 |
| Oscillibacter MIC7430 | fixed | NA | groupFBVLED:time_pointFinal | 0.00 | 0.08 | 0.04 | 78.00 | -0.16 | 0.17 | 0.964 | 0.987 |
| Lactobacillus iners | fixed | NA | groupFBVLED:time_pointFinal | -0.01 | 0.16 | -0.04 | 39.00 | -0.32 | 0.31 | 0.965 | 0.987 |
| Finegoldia magna | fixed | NA | groupFBVLED:time_pointFinal | 0.02 | 0.44 | 0.04 | 39.00 | -0.87 | 0.91 | 0.968 | 0.988 |
| UBA5446 MIC9241 | fixed | NA | groupFBVLED:time_pointFinal | 0.01 | 0.21 | 0.04 | 39.00 | -0.42 | 0.44 | 0.969 | 0.988 |

| OUTCOMES - SPECIES | EFFECT | GROUP | TERM | ESTIMATE | STD ERROR | STATISTIC | DF | CONF LOW | CONF HIGH | P VALUE | Q VALUE |
|---|---|---|---|---|---|---|---|---|---|---|---|
| CAG 495 sp000436375 | fixed | NA | groupFBVLED:time_pointFinal | -0.02 | 0.42 | -0.04 | 39.00 | -0.86 | 0.83 | 0.971 | 0.988 |
| CAG 177 sp003514385 | fixed | NA | groupFBVLED:time_pointFinal | -0.01 | 0.35 | -0.03 | 39.00 | -0.72 | 0.70 | 0.976 | 0.992 |
| Parabacteroides distasonis | fixed | NA | groupFBVLED:time_pointFinal | -0.01 | 0.34 | -0.03 | 39.00 | -0.70 | 0.69 | 0.980 | 0.993 |
| Oscillibacter MIC7608 | fixed | NA | groupFBVLED:time_pointFinal | 0.00 | 0.16 | 0.02 | 39.00 | -0.32 | 0.33 | 0.980 | 0.993 |
| UBA1777 sp003150355 | fixed | NA | groupFBVLED:time_pointFinal | 0.00 | 0.10 | -0.02 | 39.00 | -0.21 | 0.21 | 0.983 | 0.995 |
| Oscillospiraceae MIC7451 | fixed | NA | groupFBVLED:time_pointFinal | 0.00 | 0.04 | -0.01 | 39.00 | -0.09 | 0.08 | 0.990 | 0.997 |
| Blautia_A sp000436615 | fixed | NA | groupFBVLED:time_pointFinal | -0.01 | 0.48 | -0.01 | 39.00 | -0.98 | 0.97 | 0.990 | 0.997 |
| Eubacterium_E hallii_A | fixed | NA | groupFBVLED:time_pointFinal | 0.00 | 0.26 | 0.01 | 39.00 | -0.52 | 0.53 | 0.991 | 0.997 |
| Faecalicatena sp900120155 | fixed | NA | groupFBVLED:time_pointFinal | 0.00 | 0.07 | 0.01 | 78.00 | -0.14 | 0.14 | 0.993 | 0.998 |
| GCA 900066135 sp900066135 | fixed | NA | groupFBVLED:time_pointFinal | 0.00 | 0.17 | 0.01 | 39.00 | -0.35 | 0.35 | 0.995 | 0.998 |
| CAG 170 sp002404795 | fixed | NA | groupFBVLED:time_pointFinal | 0.00 | 0.45 | 0.00 | 39.00 | -0.92 | 0.92 | 0.999 | 1.000 |
| Eubacterium_E MIC8705 | fixed | NA | groupFBVLED:time_pointFinal | 0.00 | 0.18 | 0.00 | 78.00 | -0.36 | 0.36 | 1.000 | 1.000 |

Unadjusted model: ~ group*timepoint.

Adjusted q-value: The p-value controlling for the false discovery rate using the Benjamini-Hochberg procedure.

Abbreviations: β, Beta-coefficient; CI, Confidence interval.
