## Supplementary material for "The effects of food-based versus supplement-based very low-energy diets on gut microbiome composition and health outcomes in women with high body mass index (The MicroFit Study): a randomised controlled trial": Table S7

**Table S7.** Unadjusted modified intention-to-treat analysis (n=45) of the differential changes in bacterial genera between those that consumed a food-based versus supplement-based very low-energy diet for three weeks.

| OUTCOMES - GENERA | EFFECT | GROUP | TERM | ESTIMATE | STD ERROR | STATISTIC | DF | CONF LOW | CONF HIGH | P VALUE | Q VALUE |
|---|---|---|---|---|---|---|---|---|---|---|---|
| GCA_900066905 | fixed | NA | groupFBVLED:time_pointFinal | -1.21 | 0.30 | -3.98 | 46.01 | -1.82 | -0.60 | **0.000** | **0.056** |
| Lachnospira | fixed | NA | groupFBVLED:time_pointFinal | 2.17 | 0.58 | 3.73 | 43.50 | 0.99 | 3.34 | **0.001** | **0.056** |
| Anaerostipes | fixed | NA | groupFBVLED:time_pointFinal | -1.73 | 0.47 | -3.73 | 41.37 | -2.67 | -0.79 | **0.001** | **0.056** |
| Lachnospiraceae_MIC9331 | fixed | NA | groupFBVLED:time_pointFinal | -1.35 | 0.39 | -3.47 | 45.75 | -2.13 | -0.57 | **0.001** | **0.079** |
| Ruminococcus_A | fixed | NA | groupFBVLED:time_pointFinal | -1.05 | 0.31 | -3.40 | 40.07 | -1.68 | -0.43 | **0.002** | **0.079** |
| UBA11774 | fixed | NA | groupFBVLED:time_pointFinal | 2.12 | 0.65 | 3.27 | 42.94 | 0.81 | 3.43 | **0.002** | **0.079** |
| CAG_103 | fixed | NA | groupFBVLED:time_pointFinal | 1.91 | 0.58 | 3.27 | 41.41 | 0.73 | 3.10 | **0.002** | **0.079** |
| Clostridium_A | fixed | NA | groupFBVLED:time_pointFinal | -1.78 | 0.55 | -3.25 | 44.74 | -2.89 | -0.68 | **0.002** | **0.079** |
| Eggerthella | fixed | NA | groupFBVLED:time_pointFinal | -1.69 | 0.55 | -3.05 | 41.49 | -2.81 | -0.57 | **0.004** | **0.122** |
| Lachnospiraceae_MIC7543 | fixed | NA | groupFBVLED:time_pointFinal | -0.60 | 0.20 | -3.01 | 42.01 | -1.01 | -0.20 | **0.004** | **0.122** |
| Agathobaculum | fixed | NA | groupFBVLED:time_pointFinal | 1.51 | 0.51 | 2.98 | 43.47 | 0.49 | 2.53 | **0.005** | **0.122** |
| QAND01 | fixed | NA | groupFBVLED:time_pointFinal | -0.56 | 0.19 | -2.88 | 84.00 | -0.95 | -0.17 | **0.005** | **0.122** |
| Lachnospiraceae_MIC8643 | fixed | NA | groupFBVLED:time_pointFinal | -0.83 | 0.29 | -2.89 | 44.07 | -1.41 | -0.25 | **0.006** | **0.134** |
| CAG_74_MIC8062 | fixed | NA | groupFBVLED:time_pointFinal | -1.81 | 0.64 | -2.84 | 43.98 | -3.10 | -0.52 | **0.007** | **0.139** |
| Cloacibacillus | fixed | NA | groupFBVLED:time_pointFinal | -0.67 | 0.24 | -2.82 | 43.08 | -1.14 | -0.19 | **0.007** | **0.139** |
| Massilimaliae | fixed | NA | groupFBVLED:time_pointFinal | -1.13 | 0.41 | -2.75 | 46.23 | -1.96 | -0.30 | **0.008** | 0.152 |
| Streptococcus | fixed | NA | groupFBVLED:time_pointFinal | 1.76 | 0.64 | 2.73 | 40.76 | 0.46 | 3.05 | **0.009** | 0.154 |
| Desulfovibrio | fixed | NA | groupFBVLED:time_pointFinal | -0.66 | 0.25 | -2.70 | 39.77 | -1.16 | -0.17 | **0.010** | 0.154 |
| CAG_74_MIC7649 | fixed | NA | groupFBVLED:time_pointFinal | -1.13 | 0.42 | -2.69 | 41.56 | -1.98 | -0.28 | **0.010** | 0.154 |
| UBA1417 | fixed | NA | groupFBVLED:time_pointFinal | -1.46 | 0.54 | -2.68 | 40.03 | -2.55 | -0.36 | **0.011** | 0.154 |
| Erysipelatoclostridium | fixed | NA | groupFBVLED:time_pointFinal | -1.26 | 0.47 | -2.65 | 41.57 | -2.22 | -0.30 | **0.011** | 0.157 |
| Ruthenibacterium | fixed | NA | groupFBVLED:time_pointFinal | -1.98 | 0.76 | -2.61 | 43.70 | -3.51 | -0.45 | **0.012** | 0.158 |
| CAG_272_MIC9176 | fixed | NA | groupFBVLED:time_pointFinal | -0.15 | 0.06 | -2.62 | 38.16 | -0.27 | -0.03 | **0.013** | 0.158 |
| Ruminococcus_D | fixed | NA | groupFBVLED:time_pointFinal | 1.83 | 0.72 | 2.54 | 41.25 | 0.38 | 3.28 | **0.015** | 0.176 |
| Intestinimonas | fixed | NA | groupFBVLED:time_pointFinal | -1.46 | 0.58 | -2.52 | 44.93 | -2.63 | -0.29 | **0.015** | 0.176 |
| Ruminococcus_E | fixed | NA | groupFBVLED:time_pointFinal | 2.29 | 0.91 | 2.51 | 40.34 | 0.45 | 4.13 | **0.016** | 0.178 |
| CAG_81 | fixed | NA | groupFBVLED:time_pointFinal | 1.31 | 0.53 | 2.47 | 41.69 | 0.24 | 2.37 | **0.018** | 0.182 |
| Negativibacillus | fixed | NA | groupFBVLED:time_pointFinal | 1.20 | 0.49 | 2.46 | 40.87 | 0.22 | 2.19 | **0.018** | 0.182 |

| OUTCOMES - GENERA | EFFECT | GROUP | TERM | ESTIMATE | STD ERROR | STATISTIC | DF | CONF LOW | CONF HIGH | P VALUE | Q VALUE |
|---|---|---|---|---|---|---|---|---|---|---|---|
| Faecalibacterium | fixed | NA | groupFBVLED:time_pointFinal | 1.11 | 0.45 | 2.46 | 41.62 | 0.20 | 2.02 | **0.018** | 0.182 |
| Massilioclostridium | fixed | NA | groupFBVLED:time_pointFinal | -0.45 | 0.19 | -2.43 | 43.37 | -0.82 | -0.08 | **0.019** | 0.185 |
| Christensenellales_MIC6424 | fixed | NA | groupFBVLED:time_pointFinal | -0.34 | 0.14 | -2.41 | 41.36 | -0.62 | -0.06 | **0.020** | 0.189 |
| Bifidobacterium | fixed | NA | groupFBVLED:time_pointFinal | -1.43 | 0.59 | -2.40 | 40.49 | -2.63 | -0.23 | **0.021** | 0.190 |
| Eubacterium_F | fixed | NA | groupFBVLED:time_pointFinal | 1.43 | 0.60 | 2.37 | 41.07 | 0.21 | 2.64 | **0.022** | 0.196 |
| Lachnospiraceae_MIC8879 | fixed | NA | groupFBVLED:time_pointFinal | 0.71 | 0.30 | 2.35 | 41.95 | 0.10 | 1.31 | **0.023** | 0.198 |
| GCA_900066995 | fixed | NA | groupFBVLED:time_pointFinal | 1.23 | 0.53 | 2.32 | 44.75 | 0.16 | 2.30 | **0.025** | 0.199 |
| Oscillospiraceae_MIC9607 | fixed | NA | groupFBVLED:time_pointFinal | -0.56 | 0.24 | -2.32 | 45.11 | -1.05 | -0.07 | **0.025** | 0.199 |
| Lachnospiraceae_MIC6885 | fixed | NA | groupFBVLED:time_pointFinal | -0.13 | 0.06 | -2.32 | 38.96 | -0.25 | -0.02 | **0.026** | 0.199 |
| CAG_452 | fixed | NA | groupFBVLED:time_pointFinal | -0.42 | 0.18 | -2.30 | 40.50 | -0.79 | -0.05 | **0.027** | 0.202 |
| CAG_74_MIC9837 | fixed | NA | groupFBVLED:time_pointFinal | -0.93 | 0.41 | -2.26 | 42.89 | -1.75 | -0.10 | **0.029** | 0.212 |
| Acutalibacteraceae_MIC7526 | fixed | NA | groupFBVLED:time_pointFinal | -0.71 | 0.32 | -2.24 | 43.59 | -1.34 | -0.07 | **0.030** | 0.213 |
| UBA11524 | fixed | NA | groupFBVLED:time_pointFinal | 1.71 | 0.76 | 2.24 | 41.71 | 0.17 | 3.24 | **0.030** | 0.213 |
| Senegalimassilia | fixed | NA | groupFBVLED:time_pointFinal | -0.30 | 0.14 | -2.20 | 39.22 | -0.58 | -0.02 | **0.034** | 0.221 |
| Parasutterella | fixed | NA | groupFBVLED:time_pointFinal | 0.79 | 0.36 | 2.18 | 39.43 | 0.06 | 1.51 | **0.035** | 0.221 |
| Parvimonas | fixed | NA | groupFBVLED:time_pointFinal | -0.27 | 0.13 | -2.17 | 40.47 | -0.53 | -0.02 | **0.036** | 0.221 |
| Clostridium | fixed | NA | groupFBVLED:time_pointFinal | 1.76 | 0.82 | 2.16 | 42.13 | 0.12 | 3.41 | **0.036** | 0.221 |
| Parabacteroides | fixed | NA | groupFBVLED:time_pointFinal | -0.68 | 0.32 | -2.15 | 41.73 | -1.33 | -0.04 | **0.038** | 0.221 |
| Terrisporobacter | fixed | NA | groupFBVLED:time_pointFinal | 1.14 | 0.53 | 2.15 | 40.48 | 0.07 | 2.21 | **0.038** | 0.221 |
| UBA737 | fixed | NA | groupFBVLED:time_pointFinal | -0.27 | 0.13 | -2.14 | 40.81 | -0.52 | -0.02 | **0.038** | 0.221 |
| Lachnospirales_MIC6978 | fixed | NA | groupFBVLED:time_pointFinal | -0.33 | 0.16 | -2.14 | 41.68 | -0.65 | -0.02 | **0.038** | 0.221 |
| CAG_45 | fixed | NA | groupFBVLED:time_pointFinal | 1.77 | 0.83 | 2.13 | 44.19 | 0.09 | 3.44 | **0.039** | 0.221 |
| Oscillospiraceae_MIC7451 | fixed | NA | groupFBVLED:time_pointFinal | -0.22 | 0.11 | -2.13 | 41.24 | -0.44 | -0.01 | **0.039** | 0.221 |
| CAG_95 | fixed | NA | groupFBVLED:time_pointFinal | 1.39 | 0.66 | 2.11 | 45.69 | 0.06 | 2.71 | **0.040** | 0.221 |
| Enorma | fixed | NA | groupFBVLED:time_pointFinal | -0.16 | 0.08 | -2.12 | 39.65 | -0.31 | -0.01 | **0.041** | 0.221 |
| Agathobacter | fixed | NA | groupFBVLED:time_pointFinal | 1.50 | 0.72 | 2.09 | 42.53 | 0.06 | 2.95 | **0.042** | 0.223 |
| CAG_272_MIC6999 | fixed | NA | groupFBVLED:time_pointFinal | -0.74 | 0.36 | -2.09 | 44.08 | -1.46 | -0.03 | **0.043** | 0.223 |
| QAND01_MIC9113 | fixed | NA | groupFBVLED:time_pointFinal | -0.78 | 0.38 | -2.07 | 42.48 | -1.54 | -0.02 | **0.044** | 0.227 |
| Flavonifractor | fixed | NA | groupFBVLED:time_pointFinal | -1.41 | 0.69 | -2.04 | 43.58 | -2.79 | -0.02 | 0.047 | 0.235 |
| COE1 | fixed | NA | groupFBVLED:time_pointFinal | 0.74 | 0.36 | 2.05 | 39.92 | 0.01 | 1.48 | 0.047 | 0.235 |

| OUTCOMES - GENERA | EFFECT | GROUP | TERM | ESTIMATE | STD ERROR | STATISTIC | DF | CONF LOW | CONF HIGH | P VALUE | Q VALUE |
|---|---|---|---|---|---|---|---|---|---|---|---|
| Oxalobacter | fixed | NA | groupFBVLED:time_pointFinal | 0.46 | 0.23 | 2.02 | 43.14 | 0.00 | 0.92 | 0.049 | 0.241 |
| Massiliomicrobiota | fixed | NA | groupFBVLED:time_pointFinal | -0.42 | 0.21 | -2.01 | 41.44 | -0.85 | 0.00 | 0.051 | 0.243 |
| Lachnospiraceae_MIC9183 | fixed | NA | groupFBVLED:time_pointFinal | -0.60 | 0.30 | -2.00 | 44.58 | -1.21 | 0.00 | 0.051 | 0.243 |
| Hungatella | fixed | NA | groupFBVLED:time_pointFinal | -1.33 | 0.67 | -1.99 | 44.54 | -2.69 | 0.02 | 0.053 | 0.247 |
| CAG_74_MIC8780 | fixed | NA | groupFBVLED:time_pointFinal | -0.47 | 0.24 | -1.97 | 44.12 | -0.95 | 0.01 | 0.055 | 0.250 |
| Acutalibacter | fixed | NA | groupFBVLED:time_pointFinal | -1.25 | 0.64 | -1.97 | 43.57 | -2.53 | 0.03 | 0.056 | 0.250 |
| Lachnospiraceae_MIC9280 | fixed | NA | groupFBVLED:time_pointFinal | -0.39 | 0.20 | -1.96 | 39.79 | -0.79 | 0.01 | 0.057 | 0.254 |
| UBA5394 | fixed | NA | groupFBVLED:time_pointFinal | -0.96 | 0.49 | -1.94 | 45.88 | -1.95 | 0.04 | 0.058 | 0.254 |
| QAND01_MIC9470 | fixed | NA | groupFBVLED:time_pointFinal | -0.82 | 0.43 | -1.93 | 45.95 | -1.68 | 0.04 | 0.060 | 0.254 |
| CAG_74_MIC6989 | fixed | NA | groupFBVLED:time_pointFinal | -0.96 | 0.50 | -1.93 | 45.56 | -1.96 | 0.04 | 0.060 | 0.254 |
| Levyella | fixed | NA | groupFBVLED:time_pointFinal | -0.89 | 0.46 | -1.92 | 41.66 | -1.82 | 0.04 | 0.061 | 0.257 |
| UBA9502 | fixed | NA | groupFBVLED:time_pointFinal | 1.01 | 0.54 | 1.88 | 41.31 | -0.07 | 2.09 | 0.067 | 0.274 |
| UBA5416 | fixed | NA | groupFBVLED:time_pointFinal | -0.55 | 0.30 | -1.82 | 42.61 | -1.16 | 0.06 | 0.076 | 0.302 |
| Dialister | fixed | NA | groupFBVLED:time_pointFinal | 0.94 | 0.52 | 1.82 | 39.03 | -0.10 | 1.99 | 0.076 | 0.302 |
| Gordonibacter | fixed | NA | groupFBVLED:time_pointFinal | -0.68 | 0.37 | -1.81 | 43.10 | -1.43 | 0.08 | 0.077 | 0.302 |
| Turicibacter | fixed | NA | groupFBVLED:time_pointFinal | 0.65 | 0.36 | 1.80 | 42.27 | -0.08 | 1.38 | 0.080 | 0.308 |
| CAG_267 | fixed | NA | groupFBVLED:time_pointFinal | -0.39 | 0.22 | -1.79 | 39.02 | -0.84 | 0.05 | 0.081 | 0.308 |
| ER4 | fixed | NA | groupFBVLED:time_pointFinal | 0.65 | 0.37 | 1.79 | 39.88 | -0.08 | 1.39 | 0.081 | 0.308 |
| Pseudoflavonifractor | fixed | NA | groupFBVLED:time_pointFinal | -0.65 | 0.37 | -1.74 | 84.00 | -1.39 | 0.09 | 0.086 | 0.314 |
| Eubacterium_R | fixed | NA | groupFBVLED:time_pointFinal | 0.82 | 0.47 | 1.76 | 40.31 | -0.12 | 1.77 | 0.086 | 0.314 |
| GCA_900066135 | fixed | NA | groupFBVLED:time_pointFinal | 0.72 | 0.41 | 1.76 | 42.30 | -0.11 | 1.54 | 0.086 | 0.314 |
| Lachnospirales_MIC7715 | fixed | NA | groupFBVLED:time_pointFinal | -0.59 | 0.34 | -1.75 | 43.32 | -1.26 | 0.09 | 0.087 | 0.314 |
| UBA644_MIC9596 | fixed | NA | groupFBVLED:time_pointFinal | -0.29 | 0.17 | -1.70 | 43.33 | -0.64 | 0.05 | 0.096 | 0.336 |
| Lachnospiraceae_MIC6495 | fixed | NA | groupFBVLED:time_pointFinal | 1.00 | 0.59 | 1.70 | 42.53 | -0.19 | 2.18 | 0.096 | 0.336 |
| Eubacterium_G | fixed | NA | groupFBVLED:time_pointFinal | 1.14 | 0.67 | 1.70 | 42.61 | -0.21 | 2.50 | 0.097 | 0.336 |
| Lachnospiraceae_MIC7886 | fixed | NA | groupFBVLED:time_pointFinal | 0.40 | 0.23 | 1.68 | 42.28 | -0.08 | 0.87 | 0.099 | 0.341 |
| TF01_11 | fixed | NA | groupFBVLED:time_pointFinal | 1.15 | 0.69 | 1.66 | 40.84 | -0.25 | 2.55 | 0.105 | 0.350 |
| UBA866 | fixed | NA | groupFBVLED:time_pointFinal | -0.46 | 0.28 | -1.66 | 43.56 | -1.03 | 0.10 | 0.105 | 0.350 |
| CAG_110 | fixed | NA | groupFBVLED:time_pointFinal | -0.87 | 0.53 | -1.64 | 39.70 | -1.94 | 0.20 | 0.108 | 0.350 |
| 4C28d_15_MIC7065 | fixed | NA | groupFBVLED:time_pointFinal | 0.33 | 0.20 | 1.64 | 43.66 | -0.08 | 0.74 | 0.108 | 0.350 |

| OUTCOMES - GENERA | EFFECT | GROUP | TERM | ESTIMATE | STD ERROR | STATISTIC | DF | CONF LOW | CONF HIGH | P VALUE | Q VALUE |
|---|---|---|---|---|---|---|---|---|---|---|---|
| Lachnospiraceae_MIC9747 | fixed | NA | groupFBVLED:time_pointFinal | -0.54 | 0.33 | -1.64 | 44.03 | -1.21 | 0.13 | 0.109 | 0.350 |
| Oscillospiraceae_MIC8045 | fixed | NA | groupFBVLED:time_pointFinal | 0.44 | 0.27 | 1.64 | 36.87 | -0.10 | 0.98 | 0.109 | 0.350 |
| UBA7102 | fixed | NA | groupFBVLED:time_pointFinal | -0.69 | 0.42 | -1.62 | 43.72 | -1.54 | 0.17 | 0.112 | 0.350 |
| UBA5446 | fixed | NA | groupFBVLED:time_pointFinal | -1.01 | 0.62 | -1.62 | 44.67 | -2.27 | 0.25 | 0.113 | 0.350 |
| Faecalitalea | fixed | NA | groupFBVLED:time_pointFinal | -0.57 | 0.35 | -1.62 | 39.37 | -1.28 | 0.14 | 0.113 | 0.350 |
| CAG_74_MIC9156 | fixed | NA | groupFBVLED:time_pointFinal | -0.81 | 0.51 | -1.58 | 45.96 | -1.85 | 0.22 | 0.122 | 0.373 |
| Sutterella | fixed | NA | groupFBVLED:time_pointFinal | -0.62 | 0.40 | -1.55 | 39.54 | -1.44 | 0.19 | 0.129 | 0.391 |
| UBA10677 | fixed | NA | groupFBVLED:time_pointFinal | -0.21 | 0.14 | -1.53 | 41.72 | -0.49 | 0.07 | 0.133 | 0.397 |
| Christensenellaceae_MIC9015 | fixed | NA | groupFBVLED:time_pointFinal | -0.44 | 0.29 | -1.51 | 84.00 | -1.02 | 0.14 | 0.134 | 0.397 |
| UBA738 | fixed | NA | groupFBVLED:time_pointFinal | -0.71 | 0.47 | -1.51 | 42.77 | -1.67 | 0.24 | 0.140 | 0.410 |
| Ruminococcus | fixed | NA | groupFBVLED:time_pointFinal | 0.40 | 0.27 | 1.48 | 42.04 | -0.14 | 0.94 | 0.145 | 0.417 |
| CAG_492 | fixed | NA | groupFBVLED:time_pointFinal | 0.55 | 0.37 | 1.48 | 44.30 | -0.20 | 1.30 | 0.145 | 0.417 |
| Lawsonibacter | fixed | NA | groupFBVLED:time_pointFinal | -0.80 | 0.54 | -1.48 | 41.66 | -1.88 | 0.29 | 0.146 | 0.417 |
| Marseille_P4683 | fixed | NA | groupFBVLED:time_pointFinal | -0.43 | 0.30 | -1.46 | 39.39 | -1.03 | 0.17 | 0.152 | 0.428 |
| CAG_312 | fixed | NA | groupFBVLED:time_pointFinal | 0.55 | 0.38 | 1.45 | 41.20 | -0.22 | 1.32 | 0.154 | 0.431 |
| CAG_314 | fixed | NA | groupFBVLED:time_pointFinal | 0.64 | 0.44 | 1.44 | 43.06 | -0.26 | 1.53 | 0.157 | 0.433 |
| CAG_74_MIC7044 | fixed | NA | groupFBVLED:time_pointFinal | 0.78 | 0.55 | 1.44 | 42.37 | -0.32 | 1.88 | 0.158 | 0.433 |
| Lactococcus | fixed | NA | groupFBVLED:time_pointFinal | -0.72 | 0.51 | -1.42 | 45.97 | -1.74 | 0.30 | 0.163 | 0.436 |
| Gemmiger | fixed | NA | groupFBVLED:time_pointFinal | 0.81 | 0.57 | 1.42 | 44.71 | -0.34 | 1.97 | 0.163 | 0.436 |
| CAG_74_MIC9091 | fixed | NA | groupFBVLED:time_pointFinal | -0.37 | 0.26 | -1.42 | 43.27 | -0.90 | 0.16 | 0.164 | 0.436 |
| Barnesiella | fixed | NA | groupFBVLED:time_pointFinal | 0.62 | 0.44 | 1.40 | 40.36 | -0.28 | 1.52 | 0.170 | 0.447 |
| An200 | fixed | NA | groupFBVLED:time_pointFinal | -0.52 | 0.38 | -1.39 | 45.54 | -1.28 | 0.23 | 0.171 | 0.447 |
| Blastocystis | fixed | NA | groupFBVLED:time_pointFinal | -0.13 | 0.10 | -1.36 | 39.37 | -0.33 | 0.06 | 0.180 | 0.464 |
| UBA1390 | fixed | NA | groupFBVLED:time_pointFinal | -0.36 | 0.26 | -1.36 | 42.88 | -0.89 | 0.17 | 0.180 | 0.464 |
| CAG_74_MIC8853 | fixed | NA | groupFBVLED:time_pointFinal | -0.14 | 0.10 | -1.35 | 39.64 | -0.34 | 0.07 | 0.184 | 0.470 |
| CAG_353 | fixed | NA | groupFBVLED:time_pointFinal | -0.51 | 0.38 | -1.34 | 42.37 | -1.28 | 0.26 | 0.188 | 0.475 |
| Adlercreutzia | fixed | NA | groupFBVLED:time_pointFinal | 0.61 | 0.46 | 1.33 | 40.15 | -0.32 | 1.53 | 0.192 | 0.481 |
| F23_B02 | fixed | NA | groupFBVLED:time_pointFinal | 0.76 | 0.58 | 1.31 | 43.61 | -0.41 | 1.94 | 0.198 | 0.490 |
| Coprococcus_A | fixed | NA | groupFBVLED:time_pointFinal | 0.37 | 0.28 | 1.31 | 39.81 | -0.20 | 0.94 | 0.199 | 0.490 |
| Ruminococcaceae_MIC8156 | fixed | NA | groupFBVLED:time_pointFinal | -0.20 | 0.16 | -1.30 | 39.54 | -0.52 | 0.11 | 0.202 | 0.493 |

| OUTCOMES - GENERA | EFFECT | GROUP | TERM | ESTIMATE | STD ERROR | STATISTIC | DF | CONF LOW | CONF HIGH | P VALUE | Q VALUE |
|---|---|---|---|---|---|---|---|---|---|---|---|
| UBA11452 | fixed | NA | groupFBVLED:time_pointFinal | -0.41 | 0.32 | -1.27 | 42.48 | -1.06 | 0.24 | 0.212 | 0.504 |
| CAG_145 | fixed | NA | groupFBVLED:time_pointFinal | -0.71 | 0.56 | -1.26 | 43.83 | -1.84 | 0.42 | 0.213 | 0.504 |
| Ruminiclostridium_C | fixed | NA | groupFBVLED:time_pointFinal | -0.37 | 0.29 | -1.26 | 39.92 | -0.97 | 0.22 | 0.213 | 0.504 |
| UBA1255_MIC8514 | fixed | NA | groupFBVLED:time_pointFinal | -0.45 | 0.36 | -1.26 | 44.81 | -1.17 | 0.27 | 0.214 | 0.504 |
| UBA9475 | fixed | NA | groupFBVLED:time_pointFinal | 0.29 | 0.23 | 1.25 | 41.13 | -0.18 | 0.76 | 0.218 | 0.506 |
| CAG_217 | fixed | NA | groupFBVLED:time_pointFinal | 0.37 | 0.29 | 1.25 | 39.52 | -0.23 | 0.96 | 0.219 | 0.506 |
| Ruminococcaceae_MIC7581 | fixed | NA | groupFBVLED:time_pointFinal | -0.42 | 0.34 | -1.25 | 45.33 | -1.09 | 0.26 | 0.219 | 0.506 |
| UBA6398 | fixed | NA | groupFBVLED:time_pointFinal | -0.09 | 0.07 | -1.23 | 39.09 | -0.23 | 0.06 | 0.228 | 0.519 |
| Odoribacter | fixed | NA | groupFBVLED:time_pointFinal | 0.49 | 0.40 | 1.22 | 39.30 | -0.32 | 1.30 | 0.229 | 0.519 |
| PeH17 | fixed | NA | groupFBVLED:time_pointFinal | 0.67 | 0.55 | 1.22 | 41.26 | -0.44 | 1.77 | 0.231 | 0.519 |
| Gabonibacter | fixed | NA | groupFBVLED:time_pointFinal | -0.23 | 0.19 | -1.21 | 42.29 | -0.60 | 0.15 | 0.233 | 0.521 |
| Coprococcus | fixed | NA | groupFBVLED:time_pointFinal | 0.87 | 0.72 | 1.20 | 40.66 | -0.59 | 2.32 | 0.236 | 0.524 |
| UBA1777 | fixed | NA | groupFBVLED:time_pointFinal | 0.38 | 0.32 | 1.17 | 41.32 | -0.28 | 1.03 | 0.250 | 0.535 |
| UBA11471 | fixed | NA | groupFBVLED:time_pointFinal | 0.49 | 0.42 | 1.17 | 40.56 | -0.36 | 1.33 | 0.250 | 0.535 |
| Romboutsia | fixed | NA | groupFBVLED:time_pointFinal | 0.59 | 0.51 | 1.16 | 42.36 | -0.44 | 1.62 | 0.252 | 0.535 |
| Dorea | fixed | NA | groupFBVLED:time_pointFinal | 0.30 | 0.26 | 1.16 | 42.00 | -0.22 | 0.81 | 0.254 | 0.535 |
| Bacteroides | fixed | NA | groupFBVLED:time_pointFinal | -0.29 | 0.25 | -1.16 | 42.02 | -0.79 | 0.21 | 0.254 | 0.535 |
| CAG_313 | fixed | NA | groupFBVLED:time_pointFinal | 0.74 | 0.64 | 1.16 | 42.88 | -0.55 | 2.03 | 0.254 | 0.535 |
| Intestinibacter | fixed | NA | groupFBVLED:time_pointFinal | 0.71 | 0.62 | 1.16 | 40.39 | -0.54 | 1.96 | 0.255 | 0.535 |
| Holdemanella | fixed | NA | groupFBVLED:time_pointFinal | -0.33 | 0.29 | -1.14 | 39.24 | -0.91 | 0.25 | 0.262 | 0.543 |
| Haemophilus_D | fixed | NA | groupFBVLED:time_pointFinal | -0.41 | 0.36 | -1.13 | 43.43 | -1.14 | 0.32 | 0.264 | 0.543 |
| Eubacterium_E | fixed | NA | groupFBVLED:time_pointFinal | -0.48 | 0.42 | -1.13 | 41.94 | -1.34 | 0.38 | 0.264 | 0.543 |
| Lachnospiraceae_MIC7157 | fixed | NA | groupFBVLED:time_pointFinal | -0.34 | 0.30 | -1.12 | 45.58 | -0.95 | 0.27 | 0.268 | 0.548 |
| CAG_475 | fixed | NA | groupFBVLED:time_pointFinal | 0.45 | 0.40 | 1.11 | 41.88 | -0.36 | 1.25 | 0.272 | 0.550 |
| Anaerotignum | fixed | NA | groupFBVLED:time_pointFinal | -0.52 | 0.47 | -1.11 | 40.44 | -1.47 | 0.43 | 0.273 | 0.550 |
| Bacteroides_A | fixed | NA | groupFBVLED:time_pointFinal | 0.62 | 0.57 | 1.10 | 40.45 | -0.52 | 1.77 | 0.276 | 0.552 |
| Lactobacillus_C | fixed | NA | groupFBVLED:time_pointFinal | 0.45 | 0.41 | 1.09 | 84.00 | -0.37 | 1.26 | 0.278 | 0.552 |
| CAG_170 | fixed | NA | groupFBVLED:time_pointFinal | 0.83 | 0.76 | 1.09 | 42.13 | -0.70 | 2.37 | 0.281 | 0.554 |
| CAG_138_MIC9630 | fixed | NA | groupFBVLED:time_pointFinal | -0.70 | 0.64 | -1.09 | 43.36 | -1.99 | 0.60 | 0.284 | 0.556 |
| UC5_1_2E3 | fixed | NA | groupFBVLED:time_pointFinal | -0.29 | 0.27 | -1.08 | 43.95 | -0.82 | 0.25 | 0.287 | 0.558 |

| OUTCOMES - GENERA | EFFECT | GROUP | TERM | ESTIMATE | STD ERROR | STATISTIC | DF | CONF LOW | CONF HIGH | P VALUE | Q VALUE |
|---|---|---|---|---|---|---|---|---|---|---|---|
| CAG_74_MIC8660 | fixed | NA | groupFBVLED:time_pointFinal | -0.29 | 0.27 | -1.07 | 44.56 | -0.83 | 0.26 | 0.292 | 0.563 |
| Slackia_A | fixed | NA | groupFBVLED:time_pointFinal | -0.07 | 0.06 | -1.07 | 39.05 | -0.19 | 0.06 | 0.293 | 0.563 |
| Butyricicoccaceae_MIC8222 | fixed | NA | groupFBVLED:time_pointFinal | -0.17 | 0.16 | -1.05 | 41.88 | -0.49 | 0.15 | 0.298 | 0.568 |
| Collinsella | fixed | NA | groupFBVLED:time_pointFinal | -0.61 | 0.58 | -1.04 | 40.30 | -1.79 | 0.57 | 0.303 | 0.570 |
| CAG_41 | fixed | NA | groupFBVLED:time_pointFinal | 0.72 | 0.69 | 1.04 | 44.32 | -0.67 | 2.10 | 0.304 | 0.570 |
| Tidjanibacter | fixed | NA | groupFBVLED:time_pointFinal | -0.13 | 0.13 | -1.04 | 39.59 | -0.39 | 0.12 | 0.306 | 0.570 |
| Acetatifactor | fixed | NA | groupFBVLED:time_pointFinal | 1.00 | 0.98 | 1.03 | 43.25 | -0.96 | 2.97 | 0.309 | 0.570 |
| UBA7182 | fixed | NA | groupFBVLED:time_pointFinal | 0.41 | 0.40 | 1.03 | 41.63 | -0.39 | 1.20 | 0.311 | 0.570 |
| Coprococcus_B | fixed | NA | groupFBVLED:time_pointFinal | 0.52 | 0.51 | 1.03 | 42.07 | -0.50 | 1.55 | 0.311 | 0.570 |
| Oscillospirales_MIC7398 | fixed | NA | groupFBVLED:time_pointFinal | -0.20 | 0.20 | -1.02 | 44.86 | -0.61 | 0.20 | 0.315 | 0.574 |
| UBA7160 | fixed | NA | groupFBVLED:time_pointFinal | 0.30 | 0.30 | 1.01 | 40.00 | -0.30 | 0.90 | 0.319 | 0.576 |
| Acutalibacteraceae_MIC6974 | fixed | NA | groupFBVLED:time_pointFinal | -0.50 | 0.50 | -1.01 | 42.86 | -1.50 | 0.50 | 0.320 | 0.576 |
| QALW01 | fixed | NA | groupFBVLED:time_pointFinal | -0.31 | 0.32 | -0.98 | 43.44 | -0.96 | 0.33 | 0.332 | 0.594 |
| UBA644_A_MIC9239 | fixed | NA | groupFBVLED:time_pointFinal | -0.17 | 0.17 | -0.97 | 44.01 | -0.52 | 0.18 | 0.337 | 0.599 |
| QAND01_MIC7514 | fixed | NA | groupFBVLED:time_pointFinal | -0.27 | 0.28 | -0.96 | 42.82 | -0.84 | 0.30 | 0.342 | 0.603 |
| Roseburia | fixed | NA | groupFBVLED:time_pointFinal | 0.84 | 0.87 | 0.96 | 43.33 | -0.93 | 2.60 | 0.343 | 0.603 |
| Escherichia | fixed | NA | groupFBVLED:time_pointFinal | -0.88 | 0.93 | -0.95 | 43.09 | -2.75 | 0.99 | 0.346 | 0.603 |
| UBA4285 | fixed | NA | groupFBVLED:time_pointFinal | -0.14 | 0.14 | -0.95 | 39.32 | -0.43 | 0.16 | 0.349 | 0.603 |
| CAG_288 | fixed | NA | groupFBVLED:time_pointFinal | 0.38 | 0.40 | 0.95 | 42.56 | -0.43 | 1.19 | 0.350 | 0.603 |
| UCG_010 | fixed | NA | groupFBVLED:time_pointFinal | -0.31 | 0.33 | -0.94 | 44.73 | -0.98 | 0.36 | 0.354 | 0.603 |
| Lachnospirales_MIC8074 | fixed | NA | groupFBVLED:time_pointFinal | -0.23 | 0.25 | -0.94 | 44.53 | -0.73 | 0.27 | 0.355 | 0.603 |
| Acutalibacteraceae_MIC7795 | fixed | NA | groupFBVLED:time_pointFinal | -0.30 | 0.32 | -0.93 | 40.86 | -0.95 | 0.35 | 0.356 | 0.603 |
| Clostridium_M | fixed | NA | groupFBVLED:time_pointFinal | 0.55 | 0.59 | 0.92 | 42.16 | -0.65 | 1.74 | 0.361 | 0.607 |
| Finegoldia | fixed | NA | groupFBVLED:time_pointFinal | -0.64 | 0.69 | -0.92 | 40.26 | -2.03 | 0.76 | 0.364 | 0.610 |
| CAG_274 | fixed | NA | groupFBVLED:time_pointFinal | 0.49 | 0.53 | 0.91 | 39.67 | -0.59 | 1.57 | 0.367 | 0.610 |
| UBA1829 | fixed | NA | groupFBVLED:time_pointFinal | 0.19 | 0.21 | 0.90 | 41.79 | -0.24 | 0.62 | 0.371 | 0.610 |
| Mailhella | fixed | NA | groupFBVLED:time_pointFinal | -0.27 | 0.30 | -0.90 | 41.85 | -0.89 | 0.34 | 0.372 | 0.610 |
| GCA_900066575 | fixed | NA | groupFBVLED:time_pointFinal | 0.36 | 0.41 | 0.90 | 43.62 | -0.46 | 1.18 | 0.375 | 0.610 |
| Marvinbryantia | fixed | NA | groupFBVLED:time_pointFinal | 0.25 | 0.28 | 0.89 | 42.58 | -0.31 | 0.81 | 0.376 | 0.610 |
| Faecalicatena | fixed | NA | groupFBVLED:time_pointFinal | -0.42 | 0.48 | -0.89 | 44.11 | -1.38 | 0.53 | 0.377 | 0.610 |

| OUTCOMES - GENERA | EFFECT | GROUP | TERM | ESTIMATE | STD ERROR | STATISTIC | DF | CONF LOW | CONF HIGH | P VALUE | Q VALUE |
|---|---|---|---|---|---|---|---|---|---|---|---|
| Catenibacterium | fixed | NA | groupFBVLED:time_pointFinal | -0.18 | 0.22 | -0.84 | 39.65 | -0.62 | 0.25 | 0.406 | 0.653 |
| Paraprevotella | fixed | NA | groupFBVLED:time_pointFinal | 0.30 | 0.36 | 0.83 | 41.94 | -0.43 | 1.04 | 0.412 | 0.659 |
| CAG_460 | fixed | NA | groupFBVLED:time_pointFinal | 0.16 | 0.20 | 0.82 | 41.29 | -0.24 | 0.57 | 0.419 | 0.666 |
| Clostridium_P | fixed | NA | groupFBVLED:time_pointFinal | -0.32 | 0.39 | -0.83 | 12.46 | -1.16 | 0.52 | 0.421 | 0.666 |
| Lachnospiraceae_MIC6593 | fixed | NA | groupFBVLED:time_pointFinal | 0.25 | 0.31 | 0.81 | 37.34 | -0.38 | 0.88 | 0.424 | 0.666 |
| Absiella | fixed | NA | groupFBVLED:time_pointFinal | -0.49 | 0.61 | -0.80 | 43.69 | -1.73 | 0.74 | 0.426 | 0.666 |
| Prevotella | fixed | NA | groupFBVLED:time_pointFinal | 0.53 | 0.66 | 0.80 | 40.51 | -0.80 | 1.86 | 0.428 | 0.667 |
| Coprobacillus | fixed | NA | groupFBVLED:time_pointFinal | -0.36 | 0.46 | -0.78 | 43.82 | -1.27 | 0.56 | 0.439 | 0.680 |
| Phascolarctobacterium_A | fixed | NA | groupFBVLED:time_pointFinal | -0.06 | 0.07 | -0.76 | 39.18 | -0.21 | 0.09 | 0.450 | 0.691 |
| GCA_900066495 | fixed | NA | groupFBVLED:time_pointFinal | -0.19 | 0.25 | -0.76 | 40.12 | -0.70 | 0.32 | 0.452 | 0.691 |
| CAG_433 | fixed | NA | groupFBVLED:time_pointFinal | -0.33 | 0.44 | -0.75 | 37.14 | -1.21 | 0.55 | 0.456 | 0.691 |
| CAG_196 | fixed | NA | groupFBVLED:time_pointFinal | 0.42 | 0.57 | 0.74 | 40.07 | -0.72 | 1.57 | 0.461 | 0.691 |
| CAG_273 | fixed | NA | groupFBVLED:time_pointFinal | -0.35 | 0.48 | -0.74 | 40.70 | -1.31 | 0.61 | 0.461 | 0.691 |
| CAG_272_MIC8971 | fixed | NA | groupFBVLED:time_pointFinal | 0.26 | 0.35 | 0.74 | 44.59 | -0.44 | 0.95 | 0.462 | 0.691 |
| Acutalibacteraceae_MIC6990 | fixed | NA | groupFBVLED:time_pointFinal | 0.24 | 0.32 | 0.74 | 41.27 | -0.41 | 0.89 | 0.463 | 0.691 |
| CAG_727_MIC8506 | fixed | NA | groupFBVLED:time_pointFinal | 0.22 | 0.31 | 0.73 | 42.26 | -0.39 | 0.84 | 0.469 | 0.695 |
| Porphyromonas | fixed | NA | groupFBVLED:time_pointFinal | -0.38 | 0.52 | -0.72 | 84.00 | -1.41 | 0.66 | 0.470 | 0.695 |
| Alistipes | fixed | NA | groupFBVLED:time_pointFinal | -0.16 | 0.23 | -0.72 | 40.69 | -0.63 | 0.30 | 0.476 | 0.699 |
| UBA1691 | fixed | NA | groupFBVLED:time_pointFinal | -0.31 | 0.45 | -0.70 | 44.39 | -1.22 | 0.59 | 0.491 | 0.712 |
| Eubacterium_I | fixed | NA | groupFBVLED:time_pointFinal | 0.39 | 0.57 | 0.69 | 40.97 | -0.75 | 1.54 | 0.492 | 0.712 |
| Holdemania | fixed | NA | groupFBVLED:time_pointFinal | 0.27 | 0.40 | 0.69 | 44.94 | -0.52 | 1.07 | 0.492 | 0.712 |
| Anaerovoracaceae_MIC7161 | fixed | NA | groupFBVLED:time_pointFinal | 0.12 | 0.18 | 0.68 | 45.12 | -0.24 | 0.49 | 0.497 | 0.715 |
| Pauljensenia | fixed | NA | groupFBVLED:time_pointFinal | -0.24 | 0.36 | -0.67 | 42.88 | -0.98 | 0.49 | 0.507 | 0.715 |
| CAG_302 | fixed | NA | groupFBVLED:time_pointFinal | 0.22 | 0.33 | 0.67 | 40.18 | -0.44 | 0.88 | 0.508 | 0.715 |
| Phil1 | fixed | NA | groupFBVLED:time_pointFinal | 0.41 | 0.61 | 0.66 | 43.62 | -0.83 | 1.64 | 0.510 | 0.715 |
| Peptococcaceae_MIC7269 | fixed | NA | groupFBVLED:time_pointFinal | 0.18 | 0.27 | 0.66 | 40.91 | -0.36 | 0.72 | 0.511 | 0.715 |
| Merdibacter | fixed | NA | groupFBVLED:time_pointFinal | -0.15 | 0.23 | -0.66 | 40.51 | -0.62 | 0.31 | 0.511 | 0.715 |
| Peptoniphilus_B | fixed | NA | groupFBVLED:time_pointFinal | 0.23 | 0.34 | 0.66 | 45.81 | -0.46 | 0.92 | 0.511 | 0.715 |
| Ezakiella | fixed | NA | groupFBVLED:time_pointFinal | 0.39 | 0.60 | 0.65 | 84.00 | -0.80 | 1.57 | 0.518 | 0.721 |
| Anaeromassilibacillus | fixed | NA | groupFBVLED:time_pointFinal | -0.23 | 0.36 | -0.64 | 42.16 | -0.95 | 0.49 | 0.525 | 0.727 |

| OUTCOMES - GENERA | EFFECT | GROUP | TERM | ESTIMATE | STD ERROR | STATISTIC | DF | CONF LOW | CONF HIGH | P VALUE | Q VALUE |
|---|---|---|---|---|---|---|---|---|---|---|---|
| Blautia | fixed | NA | groupFBVLED:time_pointFinal | 0.38 | 0.59 | 0.64 | 39.66 | -0.82 | 1.58 | 0.528 | 0.728 |
| CAG_417 | fixed | NA | groupFBVLED:time_pointFinal | 0.25 | 0.40 | 0.63 | 43.98 | -0.55 | 1.05 | 0.534 | 0.730 |
| Angelakisella | fixed | NA | groupFBVLED:time_pointFinal | 0.29 | 0.46 | 0.63 | 41.51 | -0.65 | 1.22 | 0.535 | 0.730 |
| CAG_727_MIC7825 | fixed | NA | groupFBVLED:time_pointFinal | -0.19 | 0.31 | -0.61 | 42.93 | -0.80 | 0.43 | 0.543 | 0.737 |
| Ruminococcaceae_MIC8509 | fixed | NA | groupFBVLED:time_pointFinal | -0.13 | 0.22 | -0.61 | 43.14 | -0.57 | 0.31 | 0.548 | 0.740 |
| CAG_272_MIC9439 | fixed | NA | groupFBVLED:time_pointFinal | -0.16 | 0.26 | -0.60 | 42.50 | -0.68 | 0.37 | 0.550 | 0.740 |
| Ruminococcus_C | fixed | NA | groupFBVLED:time_pointFinal | 0.41 | 0.71 | 0.57 | 40.97 | -1.03 | 1.84 | 0.569 | 0.762 |
| Corynebacterium | fixed | NA | groupFBVLED:time_pointFinal | -0.16 | 0.28 | -0.57 | 45.77 | -0.73 | 0.41 | 0.574 | 0.766 |
| UBA3818 | fixed | NA | groupFBVLED:time_pointFinal | -0.20 | 0.37 | -0.54 | 43.02 | -0.95 | 0.55 | 0.591 | 0.784 |
| Oscillospiraceae_MIC9346 | fixed | NA | groupFBVLED:time_pointFinal | 0.14 | 0.27 | 0.54 | 41.78 | -0.40 | 0.69 | 0.595 | 0.786 |
| CAG_56 | fixed | NA | groupFBVLED:time_pointFinal | 0.32 | 0.61 | 0.52 | 40.37 | -0.92 | 1.55 | 0.607 | 0.795 |
| Butyricimonas | fixed | NA | groupFBVLED:time_pointFinal | 0.26 | 0.51 | 0.52 | 41.65 | -0.77 | 1.30 | 0.608 | 0.795 |
| CAG_272_MIC7215 | fixed | NA | groupFBVLED:time_pointFinal | 0.20 | 0.38 | 0.51 | 44.53 | -0.57 | 0.96 | 0.610 | 0.795 |
| Fenollaria | fixed | NA | groupFBVLED:time_pointFinal | -0.27 | 0.54 | -0.49 | 43.08 | -1.37 | 0.83 | 0.624 | 0.808 |
| CAG_354 | fixed | NA | groupFBVLED:time_pointFinal | 0.13 | 0.27 | 0.49 | 38.48 | -0.41 | 0.67 | 0.626 | 0.808 |
| Mogibacterium | fixed | NA | groupFBVLED:time_pointFinal | 0.18 | 0.37 | 0.49 | 84.00 | -0.56 | 0.91 | 0.629 | 0.808 |
| Murdochiella | fixed | NA | groupFBVLED:time_pointFinal | 0.13 | 0.27 | 0.47 | 45.42 | -0.41 | 0.66 | 0.637 | 0.816 |
| Bilophila | fixed | NA | groupFBVLED:time_pointFinal | -0.22 | 0.48 | -0.46 | 41.42 | -1.19 | 0.75 | 0.648 | 0.826 |
| CAG_269 | fixed | NA | groupFBVLED:time_pointFinal | -0.16 | 0.35 | -0.46 | 40.88 | -0.86 | 0.54 | 0.651 | 0.826 |
| Eggerthellaceae_MIC6427 | fixed | NA | groupFBVLED:time_pointFinal | -0.05 | 0.13 | -0.43 | 40.81 | -0.31 | 0.20 | 0.671 | 0.848 |
| CAG_74_MIC8932 | fixed | NA | groupFBVLED:time_pointFinal | -0.20 | 0.48 | -0.41 | 44.40 | -1.16 | 0.76 | 0.684 | 0.859 |
| Hungatella_A | fixed | NA | groupFBVLED:time_pointFinal | 0.18 | 0.45 | 0.41 | 42.47 | -0.73 | 1.10 | 0.687 | 0.859 |
| CAG_74_MIC7629 | fixed | NA | groupFBVLED:time_pointFinal | -0.18 | 0.45 | -0.40 | 45.86 | -1.08 | 0.72 | 0.689 | 0.859 |
| UBA1191 | fixed | NA | groupFBVLED:time_pointFinal | 0.21 | 0.52 | 0.40 | 43.20 | -0.84 | 1.25 | 0.692 | 0.859 |
| Bacteroides_B | fixed | NA | groupFBVLED:time_pointFinal | 0.12 | 0.29 | 0.40 | 42.20 | -0.47 | 0.70 | 0.695 | 0.859 |
| Anaerotruncus | fixed | NA | groupFBVLED:time_pointFinal | -0.12 | 0.32 | -0.39 | 42.77 | -0.76 | 0.52 | 0.700 | 0.859 |
| UBA1820 | fixed | NA | groupFBVLED:time_pointFinal | 0.16 | 0.41 | 0.39 | 43.65 | -0.67 | 0.98 | 0.702 | 0.859 |
| Fusicatenibacter | fixed | NA | groupFBVLED:time_pointFinal | 0.25 | 0.65 | 0.38 | 43.77 | -1.06 | 1.56 | 0.704 | 0.859 |
| CAG_180 | fixed | NA | groupFBVLED:time_pointFinal | 0.21 | 0.57 | 0.37 | 41.33 | -0.94 | 1.36 | 0.714 | 0.868 |
| Lachnospirales_MIC9617 | fixed | NA | groupFBVLED:time_pointFinal | 0.07 | 0.21 | 0.35 | 41.22 | -0.35 | 0.49 | 0.731 | 0.885 |

| OUTCOMES - GENERA | EFFECT | GROUP | TERM | ESTIMATE | STD ERROR | STATISTIC | DF | CONF LOW | CONF HIGH | P VALUE | Q VALUE |
|---|---|---|---|---|---|---|---|---|---|---|---|
| Oscillospiraceae_MIC9482 | fixed | NA | groupFBVLED:time_pointFinal | 0.12 | 0.35 | 0.33 | 44.47 | -0.60 | 0.83 | 0.743 | 0.895 |
| Eisenbergiella | fixed | NA | groupFBVLED:time_pointFinal | -0.30 | 0.92 | -0.32 | 43.38 | -2.15 | 1.56 | 0.750 | 0.900 |
| UBA5026 | fixed | NA | groupFBVLED:time_pointFinal | 0.06 | 0.20 | 0.31 | 41.63 | -0.35 | 0.47 | 0.754 | 0.902 |
| CAG_83 | fixed | NA | groupFBVLED:time_pointFinal | -0.13 | 0.41 | -0.30 | 39.12 | -0.96 | 0.71 | 0.764 | 0.906 |
| QANA01 | fixed | NA | groupFBVLED:time_pointFinal | -0.08 | 0.26 | -0.30 | 44.51 | -0.60 | 0.44 | 0.766 | 0.906 |
| KLE1615 | fixed | NA | groupFBVLED:time_pointFinal | 0.15 | 0.51 | 0.30 | 42.02 | -0.88 | 1.18 | 0.769 | 0.906 |
| UBA644 | fixed | NA | groupFBVLED:time_pointFinal | -0.10 | 0.33 | -0.29 | 44.07 | -0.75 | 0.56 | 0.771 | 0.906 |
| Campylobacter_B | fixed | NA | groupFBVLED:time_pointFinal | -0.12 | 0.41 | -0.29 | 43.54 | -0.94 | 0.70 | 0.776 | 0.908 |
| Coprobacter | fixed | NA | groupFBVLED:time_pointFinal | 0.13 | 0.48 | 0.28 | 41.05 | -0.83 | 1.09 | 0.782 | 0.912 |
| NK3B98 | fixed | NA | groupFBVLED:time_pointFinal | 0.08 | 0.30 | 0.27 | 41.07 | -0.52 | 0.68 | 0.787 | 0.914 |
| Sellimonas | fixed | NA | groupFBVLED:time_pointFinal | 0.08 | 0.28 | 0.27 | 42.75 | -0.50 | 0.65 | 0.791 | 0.915 |
| CAG_245 | fixed | NA | groupFBVLED:time_pointFinal | -0.10 | 0.40 | -0.25 | 42.40 | -0.92 | 0.71 | 0.800 | 0.922 |
| Peptoniphilus_C | fixed | NA | groupFBVLED:time_pointFinal | 0.13 | 0.57 | 0.23 | 84.00 | -1.00 | 1.26 | 0.822 | 0.944 |
| Acidaminococcus | fixed | NA | groupFBVLED:time_pointFinal | 0.04 | 0.21 | 0.21 | 38.01 | -0.37 | 0.46 | 0.832 | 0.944 |
| QAMH01 | fixed | NA | groupFBVLED:time_pointFinal | -0.05 | 0.24 | -0.21 | 40.87 | -0.53 | 0.43 | 0.836 | 0.944 |
| Oscillospiraceae_MIC8511 | fixed | NA | groupFBVLED:time_pointFinal | -0.04 | 0.21 | -0.21 | 84.00 | -0.47 | 0.38 | 0.837 | 0.944 |
| CAG_74_MIC7845 | fixed | NA | groupFBVLED:time_pointFinal | -0.08 | 0.37 | -0.20 | 84.00 | -0.81 | 0.66 | 0.839 | 0.944 |
| Methanobrevibacter_A | fixed | NA | groupFBVLED:time_pointFinal | 0.13 | 0.63 | 0.20 | 41.18 | -1.14 | 1.40 | 0.839 | 0.944 |
| CAG_1427 | fixed | NA | groupFBVLED:time_pointFinal | 0.09 | 0.49 | 0.19 | 41.09 | -0.90 | 1.08 | 0.850 | 0.951 |
| Peptostreptococcus | fixed | NA | groupFBVLED:time_pointFinal | -0.05 | 0.24 | -0.19 | 43.27 | -0.54 | 0.44 | 0.852 | 0.951 |
| CAG_127 | fixed | NA | groupFBVLED:time_pointFinal | -0.09 | 0.52 | -0.18 | 43.05 | -1.15 | 0.96 | 0.857 | 0.953 |
| Ruminiclostridium_E | fixed | NA | groupFBVLED:time_pointFinal | 0.12 | 0.70 | 0.18 | 40.84 | -1.30 | 1.55 | 0.861 | 0.954 |
| Phascolarctobacterium | fixed | NA | groupFBVLED:time_pointFinal | 0.04 | 0.25 | 0.17 | 39.27 | -0.47 | 0.55 | 0.868 | 0.957 |
| 51_20 | fixed | NA | groupFBVLED:time_pointFinal | 0.06 | 0.35 | 0.16 | 40.38 | -0.64 | 0.76 | 0.871 | 0.957 |
| Anaerovoracaceae_MIC7478 | fixed | NA | groupFBVLED:time_pointFinal | 0.04 | 0.27 | 0.15 | 39.81 | -0.51 | 0.59 | 0.885 | 0.966 |
| S5_A14a | fixed | NA | groupFBVLED:time_pointFinal | 0.07 | 0.50 | 0.14 | 84.00 | -0.92 | 1.06 | 0.886 | 0.966 |
| Peptoniphilus_A | fixed | NA | groupFBVLED:time_pointFinal | 0.10 | 0.76 | 0.13 | 40.43 | -1.43 | 1.63 | 0.896 | 0.973 |
| Monoglobus | fixed | NA | groupFBVLED:time_pointFinal | -0.05 | 0.37 | -0.12 | 43.42 | -0.80 | 0.70 | 0.901 | 0.973 |
| Alistipes_A | fixed | NA | groupFBVLED:time_pointFinal | 0.06 | 0.50 | 0.12 | 41.23 | -0.94 | 1.06 | 0.904 | 0.973 |
| Akkermansia | fixed | NA | groupFBVLED:time_pointFinal | -0.09 | 0.77 | -0.12 | 41.52 | -1.65 | 1.46 | 0.906 | 0.973 |

| OUTCOMES - GENERA | EFFECT | GROUP | TERM | ESTIMATE | STD ERROR | STATISTIC | DF | CONF LOW | CONF HIGH | P VALUE | Q VALUE |
|---|---|---|---|---|---|---|---|---|---|---|---|
| Lawsonella | fixed | NA | groupFBVLED:time_pointFinal | -0.05 | 0.45 | -0.11 | 84.00 | -0.95 | 0.85 | 0.914 | 0.978 |
| Blautia_A | fixed | NA | groupFBVLED:time_pointFinal | -0.02 | 0.25 | -0.09 | 40.67 | -0.53 | 0.48 | 0.927 | 0.987 |
| Anaerovoracaceae_MIC8502 | fixed | NA | groupFBVLED:time_pointFinal | 0.03 | 0.36 | 0.09 | 45.44 | -0.69 | 0.75 | 0.931 | 0.987 |
| Lactobacillus | fixed | NA | groupFBVLED:time_pointFinal | -0.03 | 0.41 | -0.09 | 44.48 | -0.85 | 0.78 | 0.932 | 0.987 |
| Butyricicoccus_A | fixed | NA | groupFBVLED:time_pointFinal | -0.03 | 0.34 | -0.08 | 44.25 | -0.71 | 0.66 | 0.940 | 0.991 |
| CAG_115 | fixed | NA | groupFBVLED:time_pointFinal | 0.04 | 0.60 | 0.07 | 40.84 | -1.17 | 1.26 | 0.943 | 0.991 |
| Facklamia | fixed | NA | groupFBVLED:time_pointFinal | 0.03 | 0.40 | 0.06 | 41.41 | -0.79 | 0.84 | 0.949 | 0.991 |
| Oscillibacter | fixed | NA | groupFBVLED:time_pointFinal | -0.03 | 0.55 | -0.06 | 39.51 | -1.15 | 1.09 | 0.952 | 0.991 |
| Anaerococcus | fixed | NA | groupFBVLED:time_pointFinal | -0.04 | 0.66 | -0.06 | 40.17 | -1.37 | 1.30 | 0.954 | 0.991 |
| UBA1685 | fixed | NA | groupFBVLED:time_pointFinal | -0.02 | 0.33 | -0.05 | 42.29 | -0.68 | 0.65 | 0.957 | 0.991 |
| Lactobacillus_B | fixed | NA | groupFBVLED:time_pointFinal | 0.01 | 0.14 | 0.05 | 39.63 | -0.28 | 0.29 | 0.962 | 0.993 |
| UBA1394 | fixed | NA | groupFBVLED:time_pointFinal | -0.02 | 0.38 | -0.04 | 40.07 | -0.79 | 0.76 | 0.966 | 0.993 |
| CAG_194 | fixed | NA | groupFBVLED:time_pointFinal | -0.01 | 0.41 | -0.03 | 43.45 | -0.84 | 0.81 | 0.974 | 0.994 |
| CAG_74_MIC8717 | fixed | NA | groupFBVLED:time_pointFinal | 0.01 | 0.25 | 0.03 | 45.75 | -0.50 | 0.52 | 0.976 | 0.994 |
| CAG_382_MIC9861 | fixed | NA | groupFBVLED:time_pointFinal | 0.00 | 0.16 | -0.03 | 44.18 | -0.33 | 0.32 | 0.978 | 0.994 |
| CAG_177 | fixed | NA | groupFBVLED:time_pointFinal | -0.01 | 0.52 | -0.02 | 40.53 | -1.06 | 1.03 | 0.980 | 0.994 |
| CAG_495 | fixed | NA | groupFBVLED:time_pointFinal | 0.01 | 0.45 | 0.02 | 39.88 | -0.90 | 0.92 | 0.984 | 0.994 |
| QALS01 | fixed | NA | groupFBVLED:time_pointFinal | 0.01 | 0.55 | 0.01 | 42.23 | -1.10 | 1.12 | 0.990 | 0.994 |
| Clostridium_Q | fixed | NA | groupFBVLED:time_pointFinal | -0.01 | 0.60 | -0.01 | 42.66 | -1.22 | 1.21 | 0.993 | 0.994 |
| Tyzzerella | fixed | NA | groupFBVLED:time_pointFinal | 0.00 | 0.45 | -0.01 | 41.24 | -0.92 | 0.91 | 0.994 | 0.994 |

Unadjusted model: ~ group*timepoint

Adjusted q-value: The p-value controlling for the false discovery rate using the Benjamini-Hochberg procedure.

Abbreviations: β, Beta-coefficient; CI, Confidence interval.
