## Supplementary material for "The effects of food-based versus supplement-based very low-energy diets on gut microbiome composition and health outcomes in women with high body mass index (The MicroFit Study): a randomised controlled trial": Table S8

**Table S8.** Unadjusted modified intention-to-treat analysis (n=45) of the differential changes in bacterial families between those that consumed a food-based versus supplement-based very low-energy diet for three weeks.

| OUTCOMES - FAMILY | EFFECT | GROUP | TERM | ESTIMATE | STD ERROR | STATISTIC | DF | CONF LOW | CONF HIGH | P VALUE | Q VALUE |
|---|---|---|---|---|---|---|---|---|---|---|---|
| GCA_900066905 | fixed | NA | groupFBVLED:time_pointFinal | -1.19 | 0.31 | -3.89 | 45.58 | -1.81 | -0.57 | **0.000** | **0.028** |
| QAND01 | fixed | NA | groupFBVLED:time_pointFinal | -1.66 | 0.52 | -3.18 | 44.65 | -2.71 | -0.61 | **0.003** | **0.115** |
| Butyricicoccaceae | fixed | NA | groupFBVLED:time_pointFinal | 1.34 | 0.50 | 2.69 | 43.37 | 0.33 | 2.35 | **0.010** | 0.220 |
| Synergistaceae | fixed | NA | groupFBVLED:time_pointFinal | -0.65 | 0.24 | -2.68 | 43.29 | -1.14 | -0.16 | **0.010** | 0.220 |
| Streptococcaceae | fixed | NA | groupFBVLED:time_pointFinal | 1.46 | 0.60 | 2.43 | 40.45 | 0.25 | 2.68 | **0.020** | 0.296 |
| Bifidobacteriaceae | fixed | NA | groupFBVLED:time_pointFinal | -1.41 | 0.59 | -2.40 | 40.39 | -2.60 | -0.23 | **0.021** | 0.296 |
| Acutalibacteraceae | fixed | NA | groupFBVLED:time_pointFinal | 0.74 | 0.33 | 2.22 | 31.85 | 0.06 | 1.41 | **0.033** | 0.360 |
| Christensenellales_MIC6424 | fixed | NA | groupFBVLED:time_pointFinal | -0.32 | 0.15 | -2.19 | 41.17 | -0.62 | -0.03 | **0.034** | 0.360 |
| Eggerthellaceae | fixed | NA | groupFBVLED:time_pointFinal | -0.93 | 0.45 | -2.05 | 43.57 | -1.84 | -0.02 | 0.046 | 0.405 |
| Tannerellaceae | fixed | NA | groupFBVLED:time_pointFinal | -0.67 | 0.33 | -2.03 | 41.92 | -1.33 | 0.00 | 0.049 | 0.405 |
| Lachnospirales_MIC6978 | fixed | NA | groupFBVLED:time_pointFinal | -0.32 | 0.16 | -1.96 | 41.69 | -0.64 | 0.01 | 0.057 | 0.405 |
| Clostridiaceae | fixed | NA | groupFBVLED:time_pointFinal | 1.62 | 0.83 | 1.94 | 41.52 | -0.06 | 3.29 | 0.059 | 0.405 |
| Dialisteraceae | fixed | NA | groupFBVLED:time_pointFinal | 0.96 | 0.50 | 1.92 | 38.85 | -0.05 | 1.98 | 0.062 | 0.405 |
| Turicibacteraceae | fixed | NA | groupFBVLED:time_pointFinal | 0.67 | 0.36 | 1.83 | 42.46 | -0.07 | 1.40 | 0.075 | 0.454 |
| 4C28d_15_MIC7065 | fixed | NA | groupFBVLED:time_pointFinal | 0.35 | 0.20 | 1.75 | 43.09 | -0.05 | 0.75 | 0.087 | 0.495 |
| Lachnospirales_MIC7715 | fixed | NA | groupFBVLED:time_pointFinal | -0.57 | 0.34 | -1.67 | 43.04 | -1.26 | 0.12 | 0.103 | 0.525 |
| Victivallaceae | fixed | NA | groupFBVLED:time_pointFinal | 0.65 | 0.39 | 1.65 | 44.09 | -0.14 | 1.44 | 0.106 | 0.525 |
| Erysipelatoclostridiaceae | fixed | NA | groupFBVLED:time_pointFinal | -0.56 | 0.34 | -1.63 | 42.32 | -1.25 | 0.13 | 0.111 | 0.525 |
| UBA1750 | fixed | NA | groupFBVLED:time_pointFinal | -0.67 | 0.42 | -1.59 | 43.76 | -1.52 | 0.18 | 0.119 | 0.532 |
| CAG_312 | fixed | NA | groupFBVLED:time_pointFinal | 0.57 | 0.37 | 1.53 | 41.14 | -0.18 | 1.32 | 0.134 | 0.564 |
| CAG_314 | fixed | NA | groupFBVLED:time_pointFinal | 0.65 | 0.43 | 1.51 | 43.20 | -0.22 | 1.53 | 0.139 | 0.564 |
| Barnesiellaceae | fixed | NA | groupFBVLED:time_pointFinal | 0.64 | 0.44 | 1.43 | 40.42 | -0.26 | 1.53 | 0.159 | 0.615 |
| UBA1390 | fixed | NA | groupFBVLED:time_pointFinal | -0.34 | 0.26 | -1.30 | 42.96 | -0.87 | 0.19 | 0.199 | 0.675 |
| CAG_552 | fixed | NA | groupFBVLED:time_pointFinal | -0.20 | 0.15 | -1.29 | 41.90 | -0.50 | 0.11 | 0.204 | 0.675 |
| Peptostreptococcaceae | fixed | NA | groupFBVLED:time_pointFinal | 0.67 | 0.52 | 1.29 | 42.08 | -0.38 | 1.71 | 0.205 | 0.675 |
| Erysipelotrichaceae | fixed | NA | groupFBVLED:time_pointFinal | -0.77 | 0.60 | -1.28 | 40.36 | -1.99 | 0.44 | 0.206 | 0.675 |
| UBA1255 | fixed | NA | groupFBVLED:time_pointFinal | -0.44 | 0.36 | -1.22 | 44.77 | -1.16 | 0.28 | 0.227 | 0.691 |
| CAG_313 | fixed | NA | groupFBVLED:time_pointFinal | 0.76 | 0.62 | 1.22 | 42.83 | -0.50 | 2.01 | 0.230 | 0.691 |

| OUTCOMES - FAMILY | EFFECT | GROUP | TERM | ESTIMATE | STD ERROR | STATISTIC | DF | CONF LOW | CONF HIGH | P VALUE | Q VALUE |
|---|---|---|---|---|---|---|---|---|---|---|---|
| UBA11471 | fixed | NA | groupFBVLED:time_pointFinal | 0.50 | 0.42 | 1.20 | 40.53 | -0.34 | 1.34 | 0.236 | 0.691 |
| CAG_917 | fixed | NA | groupFBVLED:time_pointFinal | 0.46 | 0.40 | 1.15 | 42.03 | -0.34 | 1.26 | 0.255 | 0.691 |
| Lactobacillaceae | fixed | NA | groupFBVLED:time_pointFinal | 0.68 | 0.60 | 1.13 | 43.18 | -0.54 | 1.90 | 0.265 | 0.691 |
| Pasteurellaceae | fixed | NA | groupFBVLED:time_pointFinal | -0.40 | 0.36 | -1.12 | 43.17 | -1.12 | 0.32 | 0.269 | 0.691 |
| Anaerotignaceae | fixed | NA | groupFBVLED:time_pointFinal | -0.51 | 0.47 | -1.08 | 40.43 | -1.46 | 0.44 | 0.285 | 0.691 |
| Coriobacteriaceae | fixed | NA | groupFBVLED:time_pointFinal | -0.65 | 0.61 | -1.06 | 40.43 | -1.87 | 0.58 | 0.294 | 0.691 |
| Blastocystidae | fixed | NA | groupFBVLED:time_pointFinal | -0.12 | 0.11 | -1.04 | 39.48 | -0.35 | 0.11 | 0.306 | 0.691 |
| UBA1381 | fixed | NA | groupFBVLED:time_pointFinal | 0.73 | 0.71 | 1.03 | 44.37 | -0.70 | 2.17 | 0.309 | 0.691 |
| CAG_274 | fixed | NA | groupFBVLED:time_pointFinal | 0.51 | 0.53 | 0.96 | 39.49 | -0.56 | 1.58 | 0.341 | 0.691 |
| Helcococcaceae | fixed | NA | groupFBVLED:time_pointFinal | -0.72 | 0.75 | -0.96 | 40.57 | -2.24 | 0.80 | 0.345 | 0.691 |
| Enterobacteriaceae | fixed | NA | groupFBVLED:time_pointFinal | -0.87 | 0.92 | -0.95 | 43.16 | -2.72 | 0.97 | 0.346 | 0.691 |
| CAG_288 | fixed | NA | groupFBVLED:time_pointFinal | 0.40 | 0.42 | 0.94 | 42.68 | -0.45 | 1.25 | 0.350 | 0.691 |
| Actinomycetaceae | fixed | NA | groupFBVLED:time_pointFinal | -0.46 | 0.49 | -0.94 | 43.07 | -1.46 | 0.53 | 0.353 | 0.691 |
| Oscillospirales_MIC7398 | fixed | NA | groupFBVLED:time_pointFinal | -0.19 | 0.20 | -0.94 | 44.66 | -0.59 | 0.22 | 0.354 | 0.691 |
| QALW01 | fixed | NA | groupFBVLED:time_pointFinal | -0.30 | 0.32 | -0.93 | 43.55 | -0.95 | 0.35 | 0.357 | 0.691 |
| CAG_1000 | fixed | NA | groupFBVLED:time_pointFinal | 0.18 | 0.20 | 0.88 | 41.42 | -0.23 | 0.58 | 0.381 | 0.691 |
| Marinifilaceae | fixed | NA | groupFBVLED:time_pointFinal | 0.36 | 0.41 | 0.87 | 39.16 | -0.48 | 1.20 | 0.391 | 0.691 |
| Christensenellaceae | fixed | NA | groupFBVLED:time_pointFinal | -0.26 | 0.30 | -0.86 | 44.29 | -0.86 | 0.35 | 0.395 | 0.691 |
| UBA644_A | fixed | NA | groupFBVLED:time_pointFinal | -0.15 | 0.18 | -0.86 | 43.70 | -0.52 | 0.21 | 0.395 | 0.691 |
| Rikenellaceae | fixed | NA | groupFBVLED:time_pointFinal | -0.19 | 0.22 | -0.86 | 40.62 | -0.65 | 0.26 | 0.395 | 0.691 |
| Lachnospirales_MIC8074 | fixed | NA | groupFBVLED:time_pointFinal | -0.22 | 0.25 | -0.85 | 44.26 | -0.72 | 0.29 | 0.399 | 0.691 |
| Gastranaerophilaceae | fixed | NA | groupFBVLED:time_pointFinal | 0.44 | 0.57 | 0.77 | 40.10 | -0.71 | 1.58 | 0.448 | 0.761 |
| Acidaminococcaceae | fixed | NA | groupFBVLED:time_pointFinal | 0.18 | 0.26 | 0.71 | 39.54 | -0.34 | 0.71 | 0.482 | 0.766 |
| Porphyromonadaceae | fixed | NA | groupFBVLED:time_pointFinal | -0.36 | 0.51 | -0.70 | 84.00 | -1.38 | 0.66 | 0.484 | 0.766 |
| Peptococcaceae | fixed | NA | groupFBVLED:time_pointFinal | 0.19 | 0.28 | 0.70 | 41.07 | -0.37 | 0.75 | 0.487 | 0.766 |
| CAG_302 | fixed | NA | groupFBVLED:time_pointFinal | 0.24 | 0.34 | 0.70 | 40.26 | -0.45 | 0.92 | 0.489 | 0.766 |
| CAG_433 | fixed | NA | groupFBVLED:time_pointFinal | -0.30 | 0.45 | -0.68 | 36.72 | -1.21 | 0.60 | 0.501 | 0.766 |
| CAG_382 | fixed | NA | groupFBVLED:time_pointFinal | -0.22 | 0.33 | -0.67 | 44.39 | -0.89 | 0.44 | 0.505 | 0.766 |
| Bacteroidaceae | fixed | NA | groupFBVLED:time_pointFinal | -0.15 | 0.23 | -0.65 | 42.68 | -0.62 | 0.32 | 0.520 | 0.766 |
| CAG_611 | fixed | NA | groupFBVLED:time_pointFinal | 0.26 | 0.41 | 0.64 | 44.07 | -0.56 | 1.09 | 0.524 | 0.766 |

| OUTCOMES - FAMILY | EFFECT | GROUP | TERM | ESTIMATE | STD ERROR | STATISTIC | DF | CONF LOW | CONF HIGH | P VALUE | Q VALUE |
|---|---|---|---|---|---|---|---|---|---|---|---|
| UBA644 | fixed | NA | groupFBVLED:time_pointFinal | -0.22 | 0.34 | -0.63 | 44.16 | -0.91 | 0.48 | 0.533 | 0.766 |
| Oscillospiraceae | fixed | NA | groupFBVLED:time_pointFinal | -0.16 | 0.26 | -0.62 | 34.63 | -0.69 | 0.37 | 0.541 | 0.766 |
| Mycobacteriaceae | fixed | NA | groupFBVLED:time_pointFinal | -0.25 | 0.49 | -0.52 | 84.00 | -1.22 | 0.71 | 0.606 | 0.844 |
| UBA1820 | fixed | NA | groupFBVLED:time_pointFinal | 0.17 | 0.40 | 0.42 | 43.54 | -0.64 | 0.99 | 0.674 | 0.895 |
| Ruminococcaceae | fixed | NA | groupFBVLED:time_pointFinal | 0.09 | 0.22 | 0.41 | 44.08 | -0.35 | 0.53 | 0.685 | 0.895 |
| Anaerovoracaceae | fixed | NA | groupFBVLED:time_pointFinal | -0.19 | 0.46 | -0.41 | 41.84 | -1.13 | 0.75 | 0.687 | 0.895 |
| Lachnospirales_MIC9617 | fixed | NA | groupFBVLED:time_pointFinal | 0.09 | 0.22 | 0.40 | 41.25 | -0.35 | 0.53 | 0.692 | 0.895 |
| CAG_272 | fixed | NA | groupFBVLED:time_pointFinal | -0.16 | 0.40 | -0.40 | 41.41 | -0.97 | 0.65 | 0.695 | 0.895 |
| CAG_822 | fixed | NA | groupFBVLED:time_pointFinal | 0.08 | 0.22 | 0.35 | 42.14 | -0.37 | 0.53 | 0.726 | 0.911 |
| CAG_239 | fixed | NA | groupFBVLED:time_pointFinal | -0.20 | 0.59 | -0.35 | 40.46 | -1.39 | 0.98 | 0.729 | 0.911 |
| Coprobacteraceae | fixed | NA | groupFBVLED:time_pointFinal | 0.15 | 0.47 | 0.31 | 41.12 | -0.80 | 1.10 | 0.757 | 0.933 |
| Desulfovibrionaceae | fixed | NA | groupFBVLED:time_pointFinal | -0.11 | 0.39 | -0.29 | 39.58 | -0.91 | 0.68 | 0.772 | 0.937 |
| Campylobacteraceae | fixed | NA | groupFBVLED:time_pointFinal | -0.10 | 0.40 | -0.26 | 43.51 | -0.90 | 0.70 | 0.799 | 0.957 |
| Methanobacteriaceae | fixed | NA | groupFBVLED:time_pointFinal | 0.14 | 0.61 | 0.24 | 41.17 | -1.10 | 1.39 | 0.815 | 0.962 |
| CAG_727 | fixed | NA | groupFBVLED:time_pointFinal | 0.11 | 0.50 | 0.22 | 41.88 | -0.91 | 1.13 | 0.826 | 0.962 |
| CAG_138 | fixed | NA | groupFBVLED:time_pointFinal | -0.12 | 0.65 | -0.19 | 41.25 | -1.44 | 1.19 | 0.850 | 0.969 |
| Burkholderiaceae | fixed | NA | groupFBVLED:time_pointFinal | 0.09 | 0.50 | 0.17 | 42.12 | -0.92 | 1.09 | 0.863 | 0.969 |
| UBA1829 | fixed | NA | groupFBVLED:time_pointFinal | -0.06 | 0.38 | -0.15 | 42.34 | -0.83 | 0.71 | 0.880 | 0.969 |
| Aerococcaceae | fixed | NA | groupFBVLED:time_pointFinal | 0.05 | 0.39 | 0.14 | 41.32 | -0.73 | 0.84 | 0.889 | 0.969 |
| QAMH01 | fixed | NA | groupFBVLED:time_pointFinal | -0.03 | 0.25 | -0.13 | 41.24 | -0.55 | 0.48 | 0.899 | 0.969 |
| Peptoniphilaceae | fixed | NA | groupFBVLED:time_pointFinal | 0.10 | 0.78 | 0.13 | 40.10 | -1.47 | 1.67 | 0.901 | 0.969 |
| Akkermansiaceae | fixed | NA | groupFBVLED:time_pointFinal | -0.08 | 0.75 | -0.11 | 41.49 | -1.60 | 1.44 | 0.914 | 0.971 |
| Monoglobaceae | fixed | NA | groupFBVLED:time_pointFinal | -0.03 | 0.39 | -0.08 | 43.51 | -0.81 | 0.75 | 0.935 | 0.973 |
| CAG_508 | fixed | NA | groupFBVLED:time_pointFinal | -0.03 | 0.49 | -0.07 | 39.77 | -1.02 | 0.95 | 0.947 | 0.973 |
| Lachnospiraceae | fixed | NA | groupFBVLED:time_pointFinal | -0.01 | 0.17 | -0.06 | 40.67 | -0.36 | 0.34 | 0.950 | 0.973 |
| Ezakiellaceae | fixed | NA | groupFBVLED:time_pointFinal | 0.02 | 0.67 | 0.04 | 41.49 | -1.33 | 1.38 | 0.971 | 0.975 |
| CAG_74 | fixed | NA | groupFBVLED:time_pointFinal | -0.02 | 0.54 | -0.03 | 40.40 | -1.10 | 1.07 | 0.975 | 0.975 |

Unadjusted model: ~ group*timepoint

Adjusted q-value: The p-value controlling for the false discovery rate using the Benjamini-Hochberg procedure.

Abbreviations: β, Beta-coefficient; CI, Confidence interval.
