## Supplementary material for "The effects of food-based versus supplement-based very low-energy diets on gut microbiome composition and health outcomes in women with high body mass index (The MicroFit Study): a randomised controlled trial": Table S9

**Table S9.** Unadjusted modified intention-to-treat analysis (n=45) of the differential changes in bacterial phyla between those that consumed a food-based versus supplement-based very low-energy diet for three weeks.

| OUTCOMES - PHYLUM | EFFECT | GROUP | TERM | ESTIMATE | STD ERROR | STATISTIC | DF | CONF LOW | CONF HIGH | P VALUE | Q VALUE |
|---|---|---|---|---|---|---|---|---|---|---|---|
| Synergistota | fixed | NA | groupFBVLED:time_pointFinal | -0.39 | 0.17 | -2.26 | 43.29 | -0.74 | -0.04 | **0.029** | 0.404 |
| Actinobacteriota | fixed | NA | groupFBVLED:time_pointFinal | -0.66 | 0.34 | -1.95 | 40.30 | -1.35 | 0.03 | 0.059 | 0.411 |
| Firmicutes_C | fixed | NA | groupFBVLED:time_pointFinal | 0.40 | 0.23 | 1.74 | 40.13 | -0.06 | 0.87 | 0.089 | 0.417 |
| Firmicutes_B | fixed | NA | groupFBVLED:time_pointFinal | 0.19 | 0.23 | 0.84 | 41.88 | -0.27 | 0.66 | 0.408 | 0.923 |
| Bacteroidota | fixed | NA | groupFBVLED:time_pointFinal | -0.14 | 0.19 | -0.72 | 42.76 | -0.52 | 0.24 | 0.474 | 0.923 |
| Cyanobacteria | fixed | NA | groupFBVLED:time_pointFinal | 0.29 | 0.41 | 0.71 | 40.13 | -0.54 | 1.13 | 0.480 | 0.923 |
| Firmicutes_A | fixed | NA | groupFBVLED:time_pointFinal | 0.11 | 0.15 | 0.70 | 43.61 | -0.20 | 0.41 | 0.488 | 0.923 |
| Blastocystidae_phylum | fixed | NA | groupFBVLED:time_pointFinal | -0.05 | 0.12 | -0.46 | 40.13 | -0.29 | 0.18 | 0.647 | 0.923 |
| Euryarchaeota | fixed | NA | groupFBVLED:time_pointFinal | 0.15 | 0.48 | 0.32 | 41.22 | -0.82 | 1.12 | 0.751 | 0.923 |
| Campylobacterota | fixed | NA | groupFBVLED:time_pointFinal | 0.08 | 0.27 | 0.30 | 43.17 | -0.46 | 0.62 | 0.764 | 0.923 |
| Firmicutes | fixed | NA | groupFBVLED:time_pointFinal | -0.05 | 0.24 | -0.19 | 42.80 | -0.54 | 0.44 | 0.853 | 0.923 |
| Verrucomicrobiota | fixed | NA | groupFBVLED:time_pointFinal | 0.09 | 0.56 | 0.16 | 40.45 | -1.04 | 1.21 | 0.877 | 0.923 |
| Proteobacteria | fixed | NA | groupFBVLED:time_pointFinal | 0.06 | 0.37 | 0.15 | 43.13 | -0.69 | 0.81 | 0.878 | 0.923 |
| Desulfobacterota_A | fixed | NA | groupFBVLED:time_pointFinal | -0.03 | 0.27 | -0.10 | 41.04 | -0.58 | 0.53 | 0.923 | 0.923 |

Unadjusted model: ~ group*timepoint.

Adjusted q-value: The p-value controlling for the false discovery rate using the Benjamini-Hochberg procedure.

Abbreviations: β, Beta-coefficient; CI, Confidence interval.
