## Supplementary material for "The effects of food-based versus supplement-based very low-energy diets on gut microbiome composition and health outcomes in women with high body mass index (The MicroFit Study): a randomised controlled trial": Table S10

**Table S10.** Unadjusted modified intention-to-treat analysis (n=45) of the differential changes in functional MetaCyc pathway alpha diversity between those that consumed a food-based versus supplement-based very low-energy diet for three weeks.

| OUTCOMES – METACYC PATHWAY | EFFECT | GROUP | TERM | ESTIMATE | STD ERROR | STATISTIC | DF | CONF LOW | CONF HIGH | P VALUE | Q VALUE |
|---|---|---|---|---|---|---|---|---|---|---|---|
| Alcohol Degradation | fixed | NA | groupFBVLED:time_pointFinal | -0.50 | 0.20 | -2.47 | 43.09 | -0.90 | -0.09 | **0.017** | 0.350 |
| Fatty Acid and Lipid Biosynthesis | fixed | NA | groupFBVLED:time_pointFinal | -0.22 | 0.10 | -2.28 | 41.23 | -0.42 | -0.03 | **0.028** | 0.350 |
| Amine and Polyamine Degradation | fixed | NA | groupFBVLED:time_pointFinal | -0.43 | 0.19 | -2.22 | 43.44 | -0.82 | -0.04 | **0.032** | 0.350 |
| Secondary Metabolite Degradation | fixed | NA | groupFBVLED:time_pointFinal | -0.19 | 0.09 | -2.21 | 41.44 | -0.36 | -0.02 | **0.033** | 0.350 |
| Entner Duodoroff Pathways | fixed | NA | groupFBVLED:time_pointFinal | -1.12 | 0.52 | -2.14 | 43.39 | -2.18 | -0.07 | **0.038** | 0.350 |
| Storage Compound Biosynthesis | fixed | NA | groupFBVLED:time_pointFinal | 0.79 | 0.38 | 2.09 | 38.96 | 0.03 | 1.56 | **0.043** | 0.350 |
| Antibiotic Resistance | fixed | NA | groupFBVLED:time_pointFinal | -1.23 | 0.66 | -1.88 | 43.05 | -2.56 | 0.09 | 0.067 | 0.439 |
| TCA cycle | fixed | NA | groupFBVLED:time_pointFinal | -1.46 | 0.81 | -1.81 | 43.10 | -3.08 | 0.16 | 0.077 | 0.439 |
| O Antigen Biosynthesis | fixed | NA | groupFBVLED:time_pointFinal | -1.03 | 0.58 | -1.78 | 44.32 | -2.20 | 0.13 | 0.082 | 0.439 |
| Other | fixed | NA | groupFBVLED:time_pointFinal | -1.10 | 0.65 | -1.70 | 43.50 | -2.41 | 0.21 | 0.096 | 0.439 |
| Cofactor Prosthetic Group Electron Carrier Degradation | fixed | NA | groupFBVLED:time_pointFinal | -0.88 | 0.52 | -1.68 | 43.53 | -1.93 | 0.17 | 0.099 | 0.439 |
| Chemoautotrophic Energy Metabolism | fixed | NA | groupFBVLED:time_pointFinal | -0.36 | 0.22 | -1.63 | 42.57 | -0.81 | 0.09 | 0.110 | 0.439 |
| C1 Compound Utilization and Assimilation | fixed | NA | groupFBVLED:time_pointFinal | -0.08 | 0.05 | -1.53 | 41.09 | -0.19 | 0.03 | 0.135 | 0.439 |
| Aromatic Compound Degradation | fixed | NA | groupFBVLED:time_pointFinal | -0.42 | 0.28 | -1.50 | 44.51 | -0.99 | 0.14 | 0.140 | 0.439 |
| Metabolic Regulator Biosynthesis | fixed | NA | groupFBVLED:time_pointFinal | -0.20 | 0.14 | -1.46 | 41.21 | -0.48 | 0.08 | 0.152 | 0.439 |
| Respiration | fixed | NA | groupFBVLED:time_pointFinal | -0.41 | 0.29 | -1.42 | 43.86 | -1.00 | 0.17 | 0.162 | 0.439 |
| Secondary Metabolite Biosynthesis | fixed | NA | groupFBVLED:time_pointFinal | -0.12 | 0.09 | -1.42 | 40.92 | -0.30 | 0.05 | 0.164 | 0.439 |
| Carbohydrate Degradation | fixed | NA | groupFBVLED:time_pointFinal | -0.11 | 0.08 | -1.37 | 40.97 | -0.26 | 0.05 | 0.177 | 0.439 |
| Methylglyoxal Detoxification | fixed | NA | groupFBVLED:time_pointFinal | -0.76 | 0.56 | -1.37 | 44.27 | -1.88 | 0.36 | 0.179 | 0.439 |
| Unclassified Pathways | fixed | NA | groupFBVLED:time_pointFinal | -0.10 | 0.07 | -1.37 | 41.07 | -0.24 | 0.05 | 0.179 | 0.439 |
| Amine and Polyamine Biosynthesis | fixed | NA | groupFBVLED:time_pointFinal | 0.18 | 0.14 | 1.30 | 43.94 | -0.10 | 0.46 | 0.199 | 0.464 |
| Carbohydrate Biosynthesis | fixed | NA | groupFBVLED:time_pointFinal | -0.08 | 0.06 | -1.26 | 41.64 | -0.20 | 0.05 | 0.215 | 0.478 |
| Nucleic Acid Processing | fixed | NA | groupFBVLED:time_pointFinal | 0.07 | 0.06 | 1.16 | 43.68 | -0.05 | 0.20 | 0.251 | 0.487 |
| Glycolysis | fixed | NA | groupFBVLED:time_pointFinal | -0.12 | 0.11 | -1.16 | 44.30 | -0.34 | 0.09 | 0.254 | 0.487 |
| Chlorinated Compound Degradation | fixed | NA | groupFBVLED:time_pointFinal | -0.26 | 0.23 | -1.15 | 40.27 | -0.72 | 0.20 | 0.258 | 0.487 |
| Polymeric Compound Degradation | fixed | NA | groupFBVLED:time_pointFinal | 0.39 | 0.35 | 1.13 | 41.62 | -0.31 | 1.10 | 0.263 | 0.487 |
| Acetyl CoA Biosynthesis | fixed | NA | groupFBVLED:time_pointFinal | 0.25 | 0.22 | 1.12 | 42.99 | -0.20 | 0.69 | 0.269 | 0.487 |

| OUTCOMES – METACYC PATHWAY | EFFECT | GROUP | TERM | ESTIMATE | STD ERROR | STATISTIC | DF | CONF LOW | CONF HIGH | P VALUE | Q VALUE |
|---|---|---|---|---|---|---|---|---|---|---|---|
| Cofactor Prosthetic Group Electron Carrier and Vitamin Biosynthesis | fixed | NA | groupFBVLED:time_pointFinal | -0.08 | 0.07 | -1.03 | 42.52 | -0.22 | 0.07 | 0.309 | 0.519 |
| Degradation Utilization Assimilation Other | fixed | NA | groupFBVLED:time_pointFinal | -0.31 | 0.30 | -1.02 | 41.45 | -0.92 | 0.30 | 0.315 | 0.519 |
| Amino Acid Degradation | fixed | NA | groupFBVLED:time_pointFinal | -0.07 | 0.07 | -1.01 | 41.73 | -0.21 | 0.07 | 0.318 | 0.519 |
| Fermentation | fixed | NA | groupFBVLED:time_pointFinal | -0.09 | 0.09 | -0.95 | 40.94 | -0.28 | 0.10 | 0.348 | 0.549 |
| Nucleoside and Nucleotide Degradation | fixed | NA | groupFBVLED:time_pointFinal | -0.09 | 0.10 | -0.91 | 42.30 | -0.29 | 0.11 | 0.370 | 0.558 |
| Other Biosynthesis | fixed | NA | groupFBVLED:time_pointFinal | 0.22 | 0.25 | 0.90 | 43.82 | -0.28 | 0.72 | 0.376 | 0.558 |
| Fatty Acid and Lipid Degradation | fixed | NA | groupFBVLED:time_pointFinal | -0.18 | 0.23 | -0.82 | 44.00 | -0.64 | 0.27 | 0.419 | 0.603 |
| Amino Acid Biosynthesis | fixed | NA | groupFBVLED:time_pointFinal | -0.06 | 0.08 | -0.78 | 42.46 | -0.22 | 0.10 | 0.441 | 0.617 |
| Carboxylate Degradation | fixed | NA | groupFBVLED:time_pointFinal | -0.08 | 0.12 | -0.66 | 43.18 | -0.31 | 0.16 | 0.513 | 0.673 |
| Glycan Degradation | fixed | NA | groupFBVLED:time_pointFinal | -0.08 | 0.12 | -0.65 | 40.88 | -0.31 | 0.16 | 0.518 | 0.673 |
| Inorganic Nutrient Metabolism | fixed | NA | groupFBVLED:time_pointFinal | -0.06 | 0.09 | -0.65 | 42.18 | -0.24 | 0.12 | 0.522 | 0.673 |
| Hydrogen Production | fixed | NA | groupFBVLED:time_pointFinal | -0.04 | 0.08 | -0.52 | 41.32 | -0.20 | 0.12 | 0.604 | 0.756 |
| Hormone Biosynthesis | fixed | NA | groupFBVLED:time_pointFinal | 0.03 | 0.05 | 0.50 | 41.26 | -0.08 | 0.14 | 0.617 | 0.756 |
| Protein Modification | fixed | NA | groupFBVLED:time_pointFinal | -0.09 | 0.20 | -0.45 | 46.17 | -0.49 | 0.31 | 0.656 | 0.784 |
| Reactive Oxygen Species Degradation | fixed | NA | groupFBVLED:time_pointFinal | 0.07 | 0.22 | 0.32 | 42.33 | -0.37 | 0.51 | 0.750 | 0.875 |
| Acid Resistance | fixed | NA | groupFBVLED:time_pointFinal | 0.02 | 0.08 | 0.28 | 41.98 | -0.14 | 0.18 | 0.778 | 0.887 |
| Glycan Biosynthesis | fixed | NA | groupFBVLED:time_pointFinal | -0.04 | 0.16 | -0.24 | 41.60 | -0.36 | 0.29 | 0.815 | 0.908 |
| Aromatic Compound Biosynthesis | fixed | NA | groupFBVLED:time_pointFinal | -0.01 | 0.07 | -0.20 | 42.58 | -0.16 | 0.13 | 0.843 | 0.918 |
| Nucleoside and Nucleotide Biosynthesis | fixed | NA | groupFBVLED:time_pointFinal | -0.01 | 0.06 | -0.15 | 43.63 | -0.14 | 0.12 | 0.879 | 0.936 |
| Aldehyde Degradation | fixed | NA | groupFBVLED:time_pointFinal | 0.01 | 0.08 | 0.10 | 40.74 | -0.15 | 0.17 | 0.924 | 0.944 |
| Pentose Phosphate Pathways | fixed | NA | groupFBVLED:time_pointFinal | 0.01 | 0.06 | 0.09 | 40.04 | -0.12 | 0.14 | 0.925 | 0.944 |
| Cell Structure Biosynthesis | fixed | NA | groupFBVLED:time_pointFinal | 0.00 | 0.05 | 0.04 | 41.63 | -0.10 | 0.11 | 0.967 | 0.967 |
