## Supplementary material for "The effects of food-based versus supplement-based very low-energy diets on gut microbiome composition and health outcomes in women with high body mass index (The MicroFit Study): a randomised controlled trial": Table S11

**Table S11.** Modified intention-to-treat sensitivity analyses (n=45) of the differential changes in Shannon index between those that consumed a food-based versus supplement-based very low-energy diet for three weeks.

| | β (95%CI) | p-value | q-value |
|---|---|---|---|
| **SENSITIVITY – BMI ADJUSTED** | | | |
| Shannon ITT (n=45) | 0.42 (0.19 to 0.66) | 0.001 | 0.001 |
| **SENSITIVITY – LOW READ SAMPLE REMOVED** | | | |
| Shannon ITT (n=45) | 0.35 (0.13 to 0.57) | 0.003 | 0.003 |
