## Supplementary Methods for "The effects of food-based versus supplement-based very low-energy diets on gut microbiome composition and health outcomes in women with high body mass index (The MicroFit Study): a randomised controlled trial"

### *Table S1.* CONSORT 2010 checklist of information to include when reporting a randomised trial*

| Section/Topic | Item No | Checklist item | Reported on page No |
|---|---|---|---|
| **Title and abstract** | | | |
| | 1a | Identification as a randomised trial in the title | Page 1 |
| | 1b | Structured summary of trial design, methods, results, and conclusions (for specific guidance see CONSORT for abstracts) | Page 5 |
| **Introduction** | | | |
| Background and objectives | 2a | Scientific background and explanation of rationale | Page 6 |
| | 2b | Specific objectives or hypotheses | Page 6 |
| **Methods** | | | |
| Trial design | 3a | Description of trial design (such as parallel, factorial) including allocation ratio | Page 7 |
| | 3b | Important changes to methods after trial commencement (such as eligibility criteria), with reasons | Page 7 |
| Participants | 4a | Eligibility criteria for participants | Page 8 |
| | 4b | Settings and locations where the data were collected | Page 7 |
| Interventions | 5 | The interventions for each group with sufficient details to allow replication, including how and when they were actually administered | Page 9 |
| Outcomes | 6a | Completely defined pre-specified primary and secondary outcome measures, including how and when they were assessed | Pages 10-12 |
| | 6b | Any changes to trial outcomes after the trial commenced, with reasons | N/A |
| Sample size | 7a | How sample size was determined | Page 10 |
| | 7b | When applicable, explanation of any interim analyses and stopping guidelines | N/A |
| Randomisation: | | | |
| Sequence generation | 8a | Method used to generate the random allocation sequence | Page 8 |
| | 8b | Type of randomisation; details of any restriction (such as blocking and block size) | Page 8 |
| Allocation concealment mechanism | 9 | Mechanism used to implement the random allocation sequence (such as sequentially numbered containers), describing any steps taken to conceal the sequence until interventions were assigned | Page 8 |
| Implementation | 10 | Who generated the random allocation sequence, who enrolled participants, and who assigned participants to interventions | Page 8 |
| Blinding | 11a | If done, who was blinded after assignment to interventions (for example, participants, care providers, those assessing outcomes) and how | Page 8 |
| | 11b | If relevant, description of the similarity of interventions | N/A |
| Statistical methods | 12a | Statistical methods used to compare groups for primary and secondary outcomes | Page 13 |
| | 12b | Methods for additional analyses, such as subgroup analyses and adjusted analyses | Page 13 |
| **Results** | | | |

| | | | |
|---|---|---|---|
| Participant flow (a diagram is strongly recommended) | 13a | For each group, the numbers of participants who were randomly assigned, received intended treatment, and were analysed for the primary outcome | Page 14 |
| | 13b | For each group, losses and exclusions after randomisation, together with reasons | Page 14 |
| Recruitment | 14a | Dates defining the periods of recruitment and follow-up | Page 7 |
| | 14b | Why the trial ended or was stopped | N/A |
| Baseline data | 15 | A table showing baseline demographic and clinical characteristics for each group | Page 15 |
| Numbers analysed | 16 | For each group, number of participants (denominator) included in each analysis and whether the analysis was by original assigned groups | Page 14 |
| Outcomes and estimation | 17a | For each primary and secondary outcome, results for each group, and the estimated effect size and its precision (such as 95% confidence interval) | Pages 16-17 |
| | 17b | For binary outcomes, presentation of both absolute and relative effect sizes is recommended | N/A |
| Ancillary analyses | 18 | Results of any other analyses performed, including subgroup analyses and adjusted analyses, distinguishing pre-specified from exploratory | Pages 16-17 |
| Harms | 19 | All important harms or unintended effects in each group (for specific guidance see CONSORT for harms) | Page 16 |
| **Discussion** | | | |
| Limitations | 20 | Trial limitations, addressing sources of potential bias, imprecision, and, if relevant, multiplicity of analyses | Page 19 |
| Generalisability | 21 | Generalisability (external validity, applicability) of the trial findings | Page 19 |
| Interpretation | 22 | Interpretation consistent with results, balancing benefits and harms, and considering other relevant evidence | Pages 19-20 |
| **Other information** | | | |
| Registration | 23 | Registration number and name of trial registry | Page 7 |
| Protocol | 24 | Where the full trial protocol can be accessed, if available | Page 7 |
| Funding | 25 | Sources of funding and other support (such as supply of drugs), role of funders | Page 21 |

*We strongly recommend reading this statement in conjunction with the CONSORT 2010 Explanation and Elaboration for important clarifications on all the items. If relevant, we also recommend reading CONSORT extensions for cluster randomised trials, non-inferiority and equivalence trials, non-pharmacological treatments, herbal interventions, and pragmatic trials. Additional extensions are forthcoming: for those and for up to date references relevant to this checklist, see www.consort-statement.org.

***Table S2.*** Strengthening the Organising and Reporting of Microbiome Studies (STORMS) checklist

| Number | Item | Recommendation | Item Source | Additional Guidance | Yes/No/NA | Comments or location in manuscript |
|---|---|---|---|---|---|---|
| **Abstract** | | | | | | |
| 1.0 | Structured or Unstructured Abstract | Abstract should include information on background, methods, results, and conclusions in structured or unstructured format. | STORMS | | Yes | Page 5 |
| 1.1 | Study Design | State study design in abstract. | STORMS | See 3.0 for additional information on study design. | Yes | Page 5 |
| 1.2 | Sequencing methods | State the strategy used for metagenomic classification. | STORMS | For example, targeted 16S by qPCR or sequencing, shotgun metagenomics, metatranscriptomics, etc. | Yes | Page 5 |
| 1.3 | Specimens | Describe body site(s) studied. | STORMS | | Yes | Page 5 |
| **Introduction** | | | | | | |
| 2.0 | Background and Rationale | Summarize the underlying background, scientific evidence, or theory driving the current hypothesis as well as the study objectives. | STORMS | | Yes | Page 6 |
| 2.1 | Hypotheses | State the pre-specified hypothesis. If the study is exploratory, state any pre-specified study objectives. | STORMS | | Yes | Page 6 |
| **Methods** | | | | | | |
| 3.0 | Study Design | Describe the study design. | STORMS | Observational (Case-Control, Cohort, Cross-sectional survey, etc.) or Experimental (Randomized controlled trial, Non-randomized controlled trial, etc.). For a brief description of common study designs see: DOI: 10.11613/BM.2014.022 | Yes | Page 7 |

| | | | | | | |
|---|---|---|---|---|---|---|
| | | | | If applicable, describe any blinding (e.g. single or double-blinding) used in the course of the study. | | |
| 3.1 | Participants | State what the population of interest is, and the method by which participants are sampled from that population. Include relevant information on physiological state of the subjects or stage in the life history of disease under study when participants were sampled. | STORMS | Examples of the population of interest could be: adults with no chronic health conditions, adults with type II diabetes, newborns, etc. This is the total population to whom the study is hoped to be generalizable to. The sampling method describes how potential participants were selected from that population.<br><br>If the participants are from a substudy of a larger study, provide a brief description of that study and cite that study.<br><br>Clearly state how cases and controls are defined.<br><br>An example of relevant physiological state might be pre/post menopausal for a vaginal microbiome study; examples of stage in the life history of disease could be whether specimens were collected during active or dormant disease, or before or after treatment. | Yes | Page 7 |
| 3.2 | Geographic location | State the geographic region(s) where participants were sampled from. | MIxS: geographic location (country and/or sea,region) | Geographic coordinates can be reported to prevent potential ambiguities if necessary. | Yes | Page 7 |

| | | | | | | |
|---|---|---|---|---|---|---|
| 3.3 | Relevant Dates | State the start and end dates for recruitment, follow-up, and data collection. | STORMS | Recruitment is the period in which participants are recruited for the study. In longitudinal studies, follow-up is the date range in which participants are asked to complete a specific assessment. Finally, data collection is the total period in which data is being collected from participants including during initial recruitment through all follow-ups. | Yes | Page 7 |
| 3.4 | Eligibility criteria | List any criteria for inclusion and exclusion of recruited participants. | Modified STROBE | Among potential recruited participants, how were some chosen and others not? This could include criteria such as sex, diet, age, health status, or BMI.<br><br>If there is a primary and validation sample, describe inclusion/exclusion criteria for each. | Yes | Page 8 |
| 3.5 | Antibiotics Usage | List what is known about antibiotics usage before or during sample collection. | STORMS | If participants were excluded due to current or recent antibiotics usage, state this here.<br><br>Other factors (e.g. proton pump inhibitors, probiotics, etc.) that may influence the microbiome should also be described as well. | Yes | Page 8 |
| 3.6 | Analytic sample size | Explain how the final analytic sample size was calculated, including the number of cases and controls if relevant, and reasons for dropout at each stage of the study. This should include the number of individuals in whom microbiome sequencing was attempted and the number in whom microbiome sequencing was successful. | STORMS | Consider use of a flow diagram (see template at https://stormsmicrobiome.org/figures). Also state sample size in abstract.<br><br>If power analysis was used to calculate sample size, describe those calculations. | Yes | Page 10 |
| 3.7 | Longitudinal Studies | For longitudinal studies, state how many follow-ups were conducted, describe sample size at follow-up by group or condition, and discuss any loss to follow-up. | STORMS | If there is loss to follow-up, discuss the likelihood that drop-out is associated with exposures, treatments, or outcomes of interest. | N/A | N/A |

| | | | | | | |
|---|---|---|---|---|---|---|
| 3.8 | Matching | For matched studies, give matching criteria. | Modified STROBE | "Matched" refers to matching between comparable study participants as cases and controls or exposed / unexposed.<br><br>Indicate whether participants were individual or frequency matched and in what ratio were they matched (e.g. 1 case to 1 control). | N/A | N/A |
| 3.9 | Ethics | State the name of the institutional review board that approved the study and protocols, protocol number and date of approval, and procedures for obtaining informed consent from participants. | STORMS | | Yes | Page 7 |
| 4.0 | Laboratory methods | State the laboratory/center where laboratory work was done. | STORMS | Provide a reference to complete lab protocols if previously published elsewhere such as on protocols.io. Note any modifications of lab protocols and the reason for protocol modifications. | Yes | Pages 7, 10, 11 |
| 4.1 | Specimen collection | State the body site(s) sampled from and how specimens were collected. | MIxS: sample collection device or method; host body site | Use terms from the Uber-anatomy Ontology (https://www.ebi.ac.uk/ols/ontologies/uberon) to describe body sites in a standardized format. | Yes | Page 11 |
| 4.2 | Shipping | Describe how samples were stored and shipped to the laboratory. | STORMS | Include length of time from collection to receipt by the lab and if temperature control was used during shipping. | Yes | Page 11 |
| 4.3 | Storage | Describe how the laboratory stored samples, including time between collection and storage and any preservation buffers or refrigeration used. | STORMS | State where each procedure or lot of samples was done if not all in the same place.<br><br>Include reagent/lot/catalogue #s for storage buffers. | Yes | Page 11 |
| 4.4 | DNA extraction | Provide DNA extraction method, including kit and version if relevant. | MIxS: nucleic acid extraction | If any DNA quantification methods were used prior to DNA amplification or at the pooling step of library preparation, state so here. | Yes | Page 11 |
| 4.5 | Human DNA sequence depletion or microbial DNA enrichment | Describe whether human DNA sequence depletion or enrichment of microbial or viral DNA was performed. | STORMS | | Yes | Page 11 |

| | | | | | | |
|---|---|---|---|---|---|---|
| 4.6 | Primer selection | Provide primer selection and DNA amplification methods as well as variable region sequenced (if applicable). | MIxS: pcr primers | | N/A | N/A |
| 4.7 | Positive Controls | Describe any positive controls (mock communities) if used. | STORMS | If used, should be deposited under guidance provided in the 8.X items. | Yes | Page 11 |
| 4.8 | Negative Controls | Describe any negative controls if used. | STORMS | If used, should be deposited under guidance provided in the 8.X items. | Yes | Page 11 |
| 4.9 | Contaminant mitigation and identification | Provide any laboratory or computational methods used to control for or identify microbiome contamination from the environment, reagents, or laboratory. | STORMS | Includes filtering of reagents and other steps to minimize contamination. It is relevant to state whether the specimens of interest have low microbial load, which makes contamination especially relevant. | Yes | Page 11 |
| 4.10 | Replication | Describe any biological or technical replicates included in the sequencing, including which steps were replicated between them. | STORMS | Replication may be biological (redundant biological specimens) or technical (aliquots taken at different stages of analysis) and used in extraction, sequencing, preprocessing, and/or data analysis. | N/A | N/A |
| 4.11 | Sequencing strategy | Major divisions of strategy, such as shotgun or amplicon sequencing. | MIxS: sequencing method | For amplicon sequencing (for example, 16S variable region), state the region selected. State the model of sequencer used. | Yes | Page 11 |
| 4.12 | Sequencing methods | State whether experimental quantification was used (QMP/cell count based, spike-in based) or whether relative abundance methods were applied. | STORMS | These include read length, sequencing depth per sample (average and minimum), whether reads are paired, and other parameters. | Yes | Page 11, 12 |
| 4.13 | Batch effects | Detail any blocking or randomization used in study design to avoid confounding of batches with exposures or outcomes. Discuss any likely sources of batch effects, if known. | STORMS | Sources of batch effects include sample collection, storage, library preparation, and sequencing and are commonly unavoidable in all but the smallest of studies. | N/A | N/A |
| 4.14 | Metatranscriptomics | Detail whether any mRNA enrichment was performed and whether/how retrotranscription was performed prior to sequencing. Provide size range of isolated transcripts. Describe whether the sequencing library was stranded or not. Provide details on sequencing methods and platforms. | STORMS | Provide details on any internal standards which may have been used as well as parameters and versions of any software or databases used. | N/A | N/A |

| | | | | | | |
|---|---|---|---|---|---|---|
| 4.15 | Metaproteomics | Detail which protease was used for digestion. Provide details on proteomic methods and platforms (e.g. LC-MS/MS, instrument type, column type, mass range, resolution, scan speed, maximum injection time, isolation window, normalised collision energy, and resolution). | STORMS | Provide details on any internal standards which may have been used as well as parameters and versions of any software or databases used. | N/A | N/A |
| 4.16 | Metabolomics | Specify the analytic method used (such as nuclear magnetic resonance spectroscopy or mass spectrometry). For mass spectrometry, detail which fractions were obtained (polar and/or non-polar) and how these were analyzed. Provide details on metabolomics methods and platforms (e.g. derivatization, instrument type, injection type, column type and instrument settings). | STORMS | Provide details on any internal standards which may have been used as well as parameters and versions of any software or databases used. | N/A | N/A |
| 5.0 | Data sources/ measurement | For each non-microbiome variable, including the health condition, intervention, or other variable of interest, state how it was defined, how it was measured or collected, and any transformations applied to the variable prior to analysis. | MIxS: host disease status | State any sources of potential bias in measurements, for example multiple interviewers or measurement instruments, and whether these potential biases were assessed or accounted for in study design.<br><br>Use terms from a standardized ontology such as the Experimental Factor Ontology (https://www.ebi.ac.uk/efo/) to describe variables of interest in a standardized format. | Yes | Page 10 |
| 6.0 | Research design for causal inference | Discuss any potential for confounding by variables that may influence both the outcome and exposure of interest. State any variables controlled for and the rationale for controlling for them. | STORMS | For causal inference, this item refers to describing the assumptions that would be required to draw causal inferences from observational data. See Vujkovic-Cvijin, I., Sklar, J., Jiang, L. et al. Host variables confound gut microbiota studies of human disease. Nature 587, 448–454 (2020). https://doi.org/10.1038/s41586-020-2881-9 for more details on confounding in observational microbiome studies.<br><br>For example, hypothesized confounders may be controlled for by multivariable adjustment. Consider using a directed acyclic graph (DAG) to describe your causal model and justify any variables controlled for. DAGs can be made using www.dagitty.net. | Yes | Page 13 |

| | | | | | | |
|---|---|---|---|---|---|---|
| 6.1 | Selection bias | Discuss potential for selection or survival bias. | STORMS | Selection bias can occur when some members of the target study population are more likely to be included in the study/final analytic sample than others. Some examples include survival bias (where part of the target study population is more likely to die before they can be studied), convenience sampling (where members of the target study population are not selected at random), and loss to follow-up (when probability of dropping out is related to one of the things being studied). | N/A | N/A |
| 7.0 | Bioinformatic and Statistical Methods | Describe any transformations to quantitative variables used in analyses (e.g. use of percentages instead of counts, normalization, rarefaction, categorization). | STORMS | If a variable is analyzed using different transformations, state rationale for the transformation and for each analyses which version of the variable is used.<br><br>In case of any complex or multistep transformations, give enumerated instructions for reproducing those transformations. | Yes | Page 12 |
| 7.1 | Quality Control | Describe any methods to identify or filter low quality reads or samples. | MIxS: sequence quality check | If samples were excluded based on quality or read depth, list the criteria used, the number of samples excluded, and the final sample size after quality control. | Yes | Page 12 |
| 7.2 | Sequence analysis | Describe any taxonomic, functional profiling, or other sequence analysis performed. | MIxS: feature prediction; similarity search method | | Yes | Pages 12, 13 |

| | | | | | | |
|---|---|---|---|---|---|---|
| 7.3 | Statistical methods | Describe all statistical methods. | Modified STROBE | Describe any statistical tests used, exploratory data analysis performed, dimension reduction methods/unsupervised analysis, alpha/beta metrics, and/or methods for adjusting for measurement bias.<br><br>If multiple statistical methods are possible, discuss why the methods used were selected.<br><br>If a multiple hypothesis testing correction method was used, describe the type of correction used.<br><br>State which taxonomic levels are analyzed. | Yes | Page 13 |
| 7.4 | Longitudinal analysis | If the study is longitudinal, include a section that explicitly states what analysis methods were used (if any) to account for grouping of measurements by individual or patterns over time. | STORMS | | N/A | N/A |
| 7.5 | Subgroup analysis | Describe any methods used to examine subgroups and interactions. | STROBE | | Yes | Page 13 |
| 7.6 | Missing data | Explain how missing data were addressed. | STROBE | "Missing data" refers to participant measurements such as covariates, exposures, outcomes, or time points that should have been collected but were not, not to zeros in taxonomic abundance tables or data points not applicable to that observation. | Yes | Page 13 |
| 7.7 | Sensitivity analyses | Describe any sensitivity analyses. | STROBE | | Yes | Page 13 |
| 7.8 | Findings | State criteria used to select findings for reporting. | STORMS | For example, false discovery rate with total number of tests, effect size threshold, significance threshold, microbes of interest. | Yes | Page 13 |

| | | | | | | |
|---|---|---|---|---|---|---|
| 7.9 | Software | Cite all software (including read mapping software) and databases (including any used for taxonomic reference or annotating amplicons, if applicable) used. Include version numbers. | Modified STREGA | Installed packages, add-ons or libraries should be stated and cited in addition to the software used.<br><br>All parameters employed that differ from the default of that software/version should be provided.<br><br>This is in addition to, not a replacement for, publishing of code as outlined in the section Reproducible Research. | Yes | Pages 11, 12, 13 |
| 8.0 | Reproducible research | Make a statement about whether and how others can reproduce the reported analysis. | STORMS | Any protected information that has been excluded or provided under controlled access should be listed along with any relevant data access procedures. "On request from authors" is not sufficiently detailed; formal data access procedures and conditions should be defined.<br><br>If data are unavailable, state so clearly.<br><br>Consider using a specialized rubric for reproducible research (such as: https://mbio.asm.org/content/9/3/e00525-18.short).<br><br>Consider preregistering the study protocol (such as on osf.io or https://plos.org/open-science/preregistration/). | Yes | Page 23 |
| 8.1 | Raw data access | State where raw data may be accessed including demultiplexing information. | STORMS | Robust, long-term databases such as those hosted by NCBI and EBI are preferred. If using a private repository, provide rationale. | Yes | Page 23 |

| | | | | | | |
|---|---|---|---|---|---|---|
| 8.2 | Processed data access | State where processed data may be accessed. | STORMS | Unfiltered data should be provided.<br><br>Robust, long-term databases such as those hosted by NCBI and EBI-EMBL are preferred. Repositories like zenodo (https://zenodo.org/) or publisso (https://www.publisso.de/en/working-for-you/doi-service/) can be used to provide a DOI and long-term storage for processed datasets, even those which cannot be published openly. | Yes | Page 23 |
| 8.3 | Participant data access | State where individual participant data such as demographics and other covariates may be accessed, and how they can be matched to the microbiome data. | STORMS | If re-categorized, transformed, or otherwise derived variables were used in the analysis, these variables or code for deriving them should be provided.<br><br>Examples of how participant data can be matched to microbiome data are: using the same set of anonymized identifiers, or using different anonymized identifiers but providing a map.<br><br>Provided data should be sufficient to independently replicate the current analysis. | Yes | Page 23 |
| 8.4 | Source code access | State where code may be accessed. | STORMS | If a standard or formalized workflow was employed, reference it here. | Yes | Page 23 |
| 8.5 | Full results | Provide full results of all analyses, in computer-readable format, in supplementary materials. | STORMS | For example, any fold-changes, p-values, or FDR values calculated, provided as a spreadsheet.<br><br>Use a machine-readable, plain-text format such as csv or tsv. | Yes | Supplementary |
| **Results** | | | | | | |

| | | | | | | |
|---|---|---|---|---|---|---|
| 9.0 | Descriptive data | Give characteristics of study participants (e.g. dietary, demographic, clinical, social) and information on exposures and potential confounders. | STROBE | Typically reported in a table included in the paper or as a supplementary table. Indicate number of participants with missing data for each variable of interest.<br><br>This includes environmental and lifestyle factors that may affect the relationship between the microbiome and the condition of interest. Participant diet and medication use should be summarized, if known.<br><br>At minimum, age and sex of all participants should be summarized. | Yes | Page 15 |
| 10.0 | Microbiome data | Report descriptive findings for microbiome analyses with all applicable outcomes and covariates. | STORMS | This includes measures of diversity as well as relative abundances. These descriptive findings should be reported both for the sample overall and for individual groups. | Yes | Pages 16, 17 |
| 10.1 | Taxonomy | Identify taxonomy using standardized taxon classifications that are sufficient to uniquely identify taxa. | STORMS | If not using full taxonomic hierarchy, make sure it is clear whether names stated are species, genera, family, etc.<br><br>Italicize genus/species pairs. Consult journal guidelines or standardized references on taxonomic nomenclature. For instance, https://wwwnc.cdc.gov/eid/page/scientific-nomenclature | Yes | Page 17 |
| 10.2 | Differential abundance | Report results of differential abundance analysis by the variable of interest and (if applicable) by time, clearly indicating the direction of change and total number of taxa tested. | STORMS | If there are more than two groups, include omnibus (multigroup) test results if applicable to the research question.<br><br>If applicable, reported effect sizes should include a measure of uncertainty such as the confidence interval. | Yes | Page 17 |
| 10.3 | Other data types | Report other data analyzed--e.g. metabolic function, functional potential, MAG assembly, and RNAseq. | STORMS | | Yes | Pages 16, 17 |

| | | | | | | |
|---|---|---|---|---|---|---|
| 10.4 | Other statistical analysis | Report any statistical data analysis not covered above. | STORMS | This could include subgroup analysis, sensitivity analyses, and cluster analysis.<br><br>Visualizations should be easily interpretable and colorblind-friendly. The caption and/or main text should provide a detailed description of visualizations for visually-impaired readers. | Yes | Page 16 |
| **Discussion** | | | | | | |
| 11.0 | Key results | Summarise key results with reference to study objectives | STROBE | | Yes | Page 19 |
| 12.0 | Interpretation | Give a cautious overall interpretation of results considering objectives, limitations, multiplicity of analyses, results from similar studies, and other relevant evidence. | STROBE | Define or clarify any subjective terms such as "dominant," "dysbiosis," and similar words used in interpretation of results.<br><br>When interpreting the findings, consider how the interpretation of the findings may be summarized or quoted for the general public such as in press releases or news articles.<br><br>If causal language is used in the interpretation (such as "alters," "affects," "results in," "causes," or "impacts"), assumptions made for causal inference should be explicitly stated as part of 6.0 and 13.0.<br><br>Distinguish between function potential (ie inferred from metagenomics) and observed activity (ie metatranscriptomic, metabolomic, proteomic) if discussing microbial function. | Yes | Pages 19, 20 |
| 13.0 | Limitations | Discuss limitations of the study, taking into account sources of potential bias or imprecision. | STROBE | Also consider limitations resulting from the methods (especially novel methods), the study design, and the sample size. | Yes | Pages 19, 20 |
| 13.1 | Bias | Discuss any potential for bias to influence study findings. | STORMS | May include sampling method, representativeness of study participants, or potential confounding. | Yes | Pages 19, 20 |

| | | | | | | |
|---|---|---|---|---|---|---|
| 13.2 | Generalizability | Discuss the generalisability (external validity) of the study results | STROBE | To what populations or other settings do you expect the conclusions to generalize? | Yes | Pages 19, 20 |
| 14.0 | Ongoing/future work | Describe potential future research or ongoing research based on the study's findings. | STORMS | | Yes | Pages 19, 20 |
| **Other information** | | | | | | |
| 15.0 | Funding | Give the source of funding and the role of the funders for the present study and, if applicable, for the original study on which the present article is based | STROBE | | Yes | Pages 21, 22 |
| 15.1 | Acknowledgements | Include acknowledgements of those who contributed to the research but did not meet criteria for authorship. | STORMS | For general guidelines on authorship, see http://www.icmje.org and https://www.elsevier.com/authors/journal-authors/policies-and-ethics/credit-author-statement | Yes | Page 19 |
| 15.2 | Conflicts of Interest | Include a conflicts of interest statement. | STORMS | | Yes | Pages 21, 22 |
| 16.0 | Supplements | Indicate where supplements may be accessed and what materials they contain. | STORMS | | Yes | Page 23 |
| 17.0 | Supplementary data | Provide supplementary data files of results with for all taxa and all outcome variables analyzed. Indicate the taxonomic level of all taxa. | STORMS | Depending on the analysis performed, examples of the supplemental results included could be mean relative abundance, differential abundance, raw p-value, multiple hypothesis testing-adjusted p-values, and standard error.<br><br>All discussed taxa should include the taxonomic level (e.g. class, order, genus). | Yes | Supplementary |

***Table S3.*** Complete list of food- and supplement-based options with ingredient compositions

| **Food-based meal replacements and snacks** | |
|---|---|
| Item | Ingredient composition |
| 5 Veg Eggs | Egg (36%), Egg White (18%), Leek (11%), Mushroom (11%), Pumpkin (11%), Spinach (3.5%), Spring Onion (3.5%), Fetta Cheese (2%), Light Tasty Cheese (2%), Olive Oil (1.5%), Pepper (0.5%), Pink Salt (0%) |
| Almond and Flaxseed Porridge | Oats (45%), Protein (32%) (Whey Protein Isolate, Whey Protein Concentrate, Vanilla Flavour, Guar Gum, Stevia, Salt, Soy Lecithin), Flaxseed Flakes (13%), Almond Meal (8%), Cinnamon (0.5%) |
| Apricot & Coconut Low Carbohydrate Cookie (snack) | Lupin Flour (25%), Whole Egg, Almond Meal, Gluten Free Flour (Maize Starch, Rice Flour, Tapioca Starch, Rice Bran, Guar Gum), Erythritol, Dried Apricots (8%), Vegetable Glycerin, Desiccated Coconut (5%), Canola Oil, Soluble Fibre (Polydextrose), Natural Flavours, Monk Fruit Extract, Baking Powder. |
| Baked Bean and Feta bowl | Diced Tomato (25%) (Tomato, Citric Acid), Cannellini Beans (15%), Fetta (9%) (Pasteurised Milk, Vegetable Oil, Salt, Lactic Cultures, Non-Animal Rennet), Red Capsicum (9%), Tomato Paste (8%) (Tomato Paste, Citric Acid), Carrot (7%), Onion (7%), Celery (7%), Spinach (5%), Light Tasty Cheese (3%), Faba Bean Protein (1.5%), Spring Onion (1%), Olive Oil (0.5%), Parsley (0.5%), Garlic (0.5%), Cumin, Paprika, Corn Starch, Chilli. |
| Banana Coconut Protein Muffin | High Protein Muffin Mix (21%) (Almond Meal, Lupin, Golden Flax meal, Whey Protein, Kibbled Sunflower, Coconut Flour, Vanilla Bean Powder, Gluten Free Baking Powder, Natural Sweetener, Natural Flavour, Salt), Banana (21%), Zucchini (21%), Milk (15%), Egg White (11%), Dates (7%), Protein (1.5%) (Whey Protein Isolate, Whey Protein Concentrate, Oligofructose, Vanilla Flavour, Guar Gum, Stevia, Soy Lecithin), Sultana (1%), Stevia (1%), Coconut (1%), Nutmeg (0%), Banana Essence (0%) |
| Banana Maple Smoothie | Cashew Nuts, Banana, Maple Essence, Water, Erythritol, Pea Protein |
| Banana Spice Protein Bircher | Yoghurt (52%) (Skim Milk, Live Cultures), Ricotta (15%) (Whey, Milk, Food Acid), Banana (7%), Stevia (3%), Shredded Coconut (3%), Black Chia Seeds (2.5%), LSA (2.5%) (Linseed, Sunflower Kernel, Almond), Pepitas (2%), Slivered Almond (2%), Puffed Amaranth (2%), Protein (1%), Puffed Quinoa (1%), Gelatine (1%), Dates (1%), Salted Peanuts (1%), Almonds (1%), Orange Juice (1%), Sunflower Seed (0.5%), Pumpkin Seed (0.5%), Cinnamon (0.5%), Nutmeg (0%), Vanilla Essence (0%), Desiccated Coconut (0%), Flaxseed (0%). |
| Beef Chow Mein | Beef Mince (32%), Green Cabbage (17%), Carrot (9%), Peas (9%), Zucchini (9%), Onion (6%), Brown Rice (5%), Gluten Free Soy Sauce (2%), Sesame Seeds (1%), Olive Oil (0.5%), Garlic (0.5%), Ginger (0.5%), Sesame Oil (0.5%), Curry Powder (0.5%), Chinese Five Spice (0%), Pink Salt (0%). |

| | |
|---|---|
| Beef Madras Curry | Beef (30%), Diced Tomato (24%) (Tomato, Citric Acid), Mushroom (16%), Bok Choy (6%), Brown Rice (6%), Green Beans (3%), Onion (3%), Coconut Milk (2.5%), Green Lentils (2%), Beef Stock (2%), Tomato Paste (2%), Gluten Free Soy Sauce (0.5%), Garlic (0.5%), Ginger (0.5%), Curry Powder (0.5%), Ground Coriander (0.5%), Fresh Coriander (0%), Cumin (0%), Olive Oil (0%), Corn Starch (0%), Pink Salt (0%), Mixed Herbs (0%), Turmeric (0%), Cardamom (0%). |
| Protein Balls (snack) | Dates (43%), Almond Meal (22%), Protein Powder (22%) (Whey Protein Isolate, Whey Protein Concentrate, Oligofructose, Chocolate Flavour, Cocoa, Stevia, Lactobacillus Plantarum, Guar Gum, Soy Lecithin), Cacao Powder (6%), Cacao Nibs (4.5%), Coconut (1.5%), Coffee (0.5%). |
| Caramelised Onion & Parmesan Egg Bites (snack) | Pasteurised Egg (62%), Water, Caramelised Onion (8%) [Onion, Vinegar, Canola Oil, Sugar, Food Acid (330)], Parmesan Cheese (6%) [Pasteurised Milk, Salt, Starter Culture, Non-Animal Rennet, Lipase, Anticaking Agent (460), Tapioca Starch, Preservative (200)], Skim Milk Powder (4%), Sunflower Oil (4%), Cheese (4%), Thickener (0.5%) (1442) [From Maize], Stabiliser (0.5%) (415, 412), Salt (0.5%), Stock Powder (0.5%), Pepper (0%), Chives (0%). |
| Carrot Cake Muffin | High Protein Muffin Mix (21%) (Almond Meal, Lupin, Golden Flax meal, Whey Protein, Kibbled Sunflower, Coconut Flour, Vanilla Bean Powder, Gluten Free Baking Powder, Natural Sweetener, Natural Flavour, Salt), Apple (17%), Carrot (15%), Milk (13%), Egg White (12%), Zucchini (11%), Dates (7%), Protein (1.5%), Sultana (1%), Stevia (1%), Cinnamon (0.5%), Nutmeg (0%). |
| Cauliflower Fried Rice and Chicken | Cauliflower Rice (31%) (Cauliflower, Turmeric Powder), Chicken (17%), Peas (10%), Carrot (10%), Egg (6%)(Pasteurised Egg Pulp), Red Capsicum (6%), Quinoa (5%), Celery (5%), Onion (2.5%), Spring Onion (2.5%), Garlic (1%), Peanuts (0.5%) (Peanuts, Peanut Oil), Gluten Free Soy Sauce (0.5%), Moroccan Spice (0.5%), Olive Oil (0.5%), Chilli (0.5%), Pink Salt (0%), Ginger (0%). |
| Cauliflower, Leek and Bacon Soup | Cauliflower (29%) (Cauliflower, Turmeric), Light Milk (17%), Ricotta Cheese (14%) (Whey, Milk, Salt, Food Acid), Cannellini Beans (9%), Bacon (5%), Leek (4.5%), Onion (3%), Potato (3%), Faba Bean Protein (1.5%), Parmesan Cheese (1%), Olive Oil (0.5%), Garlic (0.5%), Chicken Stock (0.5%), Pepper (0%), Cumin (0%). |
| Beef and Vegetable Ragout | Diced Tomato (32%) (Tomato, Citric Acid), Beef Mince (18%), Onion (16%), Zucchini (16%), Red Capsicum (11%), Light Milk (2%), Egg (2%), Gluten Free Breadcrumbs (1%), Garlic (1%), Olive Oil (0.5%), Corn Starch (0.5%), Mozzarella Cheese (0%), Pink Salt (0%), Pepper (0%), Dried Basil (0%), Oregano (0%). |
| Chilli and Ginger Baked Fish | Hoki Fish (34%), Broccoli (2%), Carrot (2%), Bok Choy (2%), Red Capsicum (2%), Celery (1.5%), Brown Rice (1%), Zucchini (0.5%), Cashews (0.5%), Onion (0.5%), Gluten Free Soy Sauce (0.5%), Olive Oil (0%), Fresh Coriander (0%), Garlic (0%), Rice Vinegar (0%), Sesame Oil (0%), Ginger (0%), Natvia (0%), Corn Starch (0%), Chilli (0%), Chinese Five Spice (0%). |

| | |
|---|---|
| Chilli Con Carne | Beef Mince (29%), Diced Tomato (17%) (Tomato, Citric Acid), Red Kidney Beans (12%), Red Capsicum (8%), Mushroom (6%), Zucchini (6%), Carrot (6%), Onion (4.5%), Tomato Paste (3.5%), Corn (3%), Gluten Free Soy Sauce (1%), Fresh Coriander (0.5%), Beef Stock (0.5%), Paprika (0.5%), Cumin (0%), Garlic (0%), Cinnamon (0%), Olive Oil (0%), Chilli Powder (0%), Corn Starch (0%). |
| Chocolate Caramel Nut Milk Smoothie | Pea Protein (6%), Cashew Nuts (5%), Peanuts (5%), Dates (4%), Cocoa (3%), Erythritol (0%), . |
| Chocolate Coconut Protein Bircher | Greek Yoghurt (49%) (Skim Milk, Live Cultures), Ricotta Cheese (18%) (Whey, Milk, Salt, Food Acid), Natvia (2%), Dates (2%), Faba Bean Protein (1.5%), Almonds (1.5%), Black Chia Seeds (1.5%), LSA (1.5%) (Linseed, Sunflower Kernel, Almond), Shredded Coconut (1.5%), Brown Rice Flakes (1.5%), 99% Sugar Free Choc Chips (1.5%), Pepitas (1.5%), Cocoa (1.5%), Vanilla Extract (0%), Xanthan Gum (0%), Cinnamon (0%), Pink Salt (0%). |
| Chocolate Truffle Low Carbohydrate Cookie (snack) | Lupin, Egg, Wholemeal Flour, Erythritol, Almond Meal, Vegetable Glycerin, Canola Oil, Natural Cocoa (5%), Desiccated Coconut (5%), Dates, Vanilla, Stevia, Coconut Essence, Baking Powder |
| Chunky Chicken, Ham and Sweetcorn Soup | Chicken (26%), Celery (11%), Corn Kernels (9%), Light Milk (9%), Leek (7%), Ham (5%), Onion (5%), Egg White (2.5%), Spring Onion (2%), Olive Oil (1.5%), Corn Starch (0.5%), Chicken Stock (0.5%), Gluten Free Soy Sauce (0.5%), Ginger (0.5%), Pepper (0%). |
| Cottage Pie with Cauliflower Mash | Beef Mince (22%), Cauliflower (19%) (Cauliflower, Turmeric Powder), Diced Tomato (8%) (Tomato, Citric Acid), Cannellini Beans (8%), Potato (8%), Mushroom (7%), Green Peas (4%), Carrot (4%), Onion (4%), Zucchini (4%), Egg White (4%), Tasty Cheese (2%), Beef Stock (1%), Tomato Paste (1%), Ricotta Cheese (1%), Gluten Free Soy Sauce (0.5%), Parmesan Cheese (0.5%), Pepper (0%), Garlic (0%), Rice Vinegar (0%), Olive Oil (0%), Corn Starch (0%), Thyme (0%), Pink Salt (0%). |
| Chicken, Pea and Ham Soup | Chicken (20%), Green Split Peas (8%), Carrot (5%), Onion (5%), Celery (5%), Zucchini (5%), Ham (5%), Parsnip (3.5%), Leek (3.5%), Cannellini Beans (2%), Chicken Stock (0.5%), Olive Oil (0.5%), Garlic (0.5%), Thyme (0%), Oregano (0%), Pepper (0%), Gluten Free Soy Sauce (0%). |
| Curried Pumpkin and Chicken Soup | Pumpkin (30%), Chicken (24%), Water (13%), Leek (9%), Carrot (7%), Sweet Potato (7%), Onion (6%), Olive Oil (1.5%), Chicken Stock (0.5%), Fresh Coriander (0.5%), Curry Powder (0.5%), Garlic (0.5%), Pink Salt (0.5%), Cumin (0%), Pepper (0%). |
| Dark Chocolate & Hazelnut Protein Balls (snack) | Dates (37%), Hazelnut Meal (30%), Protein Powder (19%) (Whey Protein Isolate, Whey Protein Concentrate, Oligofructose, Chocolate Flavour, Cocoa, Stevia, Lactobacillus Plantarum, Guar Gum, Soy Lecithin), Almond Meal (7%), Cacao Powder (6%), Coconut (1.5%), Coffee (0.5%). |
| Dim Sim (snack) | Green Cabbage (25%), Beef Mince (20%), Pork Mince (20%), Dim Sim Wrapper (11%) (Wheat Flour, Water, Salt), Mushroom (6%), Carrot (6%), Zucchini (4%), Tapioca Starch (2%), Textured Vegetable Protein (2%), Gluten Free Soy Sauce (1.5%), Beef Stock (0.5%), Natvia (0.5%), Pepper (0%), Garlic Powder (0%), Ginger Powder (0%). |

| | |
|---|---|
| Double Chocolate Muffin | High Protein Muffin Mix (20%) (Almond Meal, Lupin, Golden Flax meal, Whey Protein, Kibbled Sunflower, Coconut Flour, Vanilla Bean Powder, Gluten Free Baking Powder, Natural Sweetener, Natural Flavour, Salt), Egg White (18%), Light Milk (18%), Zucchini (12%), Sugar Free Chocolate (12%) (Cocoa Solids (45%), Soy Lecithin, Natural Vanilla Flavour), Greek Style Yoghurt (9%) (Pasteurised Whole Milk, Milk Solids, Live Cultures), Cocoa Powder (6%), Pumpkin (5%), Natural Sweetener (1%), Vanilla (0.5%) (Natural Flavour). |
| French Eggs | Egg (49%), Egg White (24%), Bacon (9%) (Pork (95%), Water, Salt, Mineral Salts (451, 452), Dextrose (Maize), Antioxidant (316), Nitrite (250), Hydrolysed Vegetable Protein (Maize)), Onion (7%), Spinach (4%), Parmesan Cheese (2.5%), Spring Onion (1.5%), Olive Oil (1.5%), Chives (0.5%), Garlic (0%), Pepper (0%). |
| Ham, Spinach and Feta Muffin | High Protein Muffin Mix (24%) (Lupin, Almond Meal, Whey Protein, Kibbled Sunflower, Coconut Flour, Gluten Free Baking Powder, Herbs, Natural Flavour, Salt), Zucchini (18%), Ham (15%) (Meat 68%, Water, Potato Starch, Acidity Regulator (325), Salt, Soy Protein, Sugar, Mineral Salt (451), Vegetable Powder, Flavouring, Antioxidant (316), Preservative (250), Fermented Rice, Spice Extract, Flavour Enhancer), Egg White (11%), Fetta Cheese (9%) (Pasteurised Milk, Salt, Starter Culture, Non-Animal Coagulant, Colour (171), Rennet), Spinach (7%) |
| Indian Chicken Curry | Chicken (35%), Diced Tomato (19%) (Tomato, Citric Acid), Potato (14%), Green Beans (7%), Coconut Milk (7%) (Coconut Cream, Xanthan Gum), Onion (5%), Peas (4.5%), Chicken Stock (2%), Gluten Free Soy Sauce (1.5%), Ginger (1%), Garlic (1%), Tomato Paste (1%), Corn Starch (0.5%), Fresh Coriander (0.5%), Curry Powder (0%), Coriander Powder (0%), Cumin (0%), Turmeric (0%), Mixed Herbs (0%), Cardamom (0%), Olive Oil (0%). |
| Indian Fish Curry | Hoki Fish (34%), Broccoli (13%), Zucchini (13%), Diced Tomato (10%) (Tomato, Citric Acid), Coconut Milk (4.5%), Carrot (3.5%), Almonds (2.5%), Onion (2.5%), Milk Powder (1.5%), Olive Oil (1%), Ginger (1%), Garlic (1%), Tomato Paste (0.5%), Corn Starch (0.5%), Coriander (0.5%), Pink Salt (0%), Chicken Stock (0%), Cumin (0%), Turmeric (0%), Ground Coriander (0%), Curry Powder (0%), Thyme (0%), Marjoram (0%), Sage (0%), Cardamom (0%). |
| Italian Beef Meatballs | Diced Tomato (24%) (Tomato, Citric Acid), Beef Mince (18%), Mushroom (14%), Zucchini (8%), Green Beans (7%), Onion (6%), Red Capsicum (4.5%), Gluten Free Pasta Penne (4.5%) (Maize Starch, Soy Flour, Potato Starch, Rice Starch), Parmesan Cheese (3.5%), Tomato Paste (2.5%), Light Milk (2%), Egg (2%), Gluten Free Breadcrumbs (1.5%), Garlic (1%), Olive Oil (0.5%), Gluten Free Soy Sauce (0.5%), Beef Stock (0%), Pepper (0%), Pink Salt (0%), Basil (0%), Oregano (0%), Corn Starch (0%), Rosemary (0%). |
| Italian Meatball Soup | Diced Tomato (29%) (Tomato, Citric Acid), Beef Mince (16%), Zucchini (11%), Carrot (7%), Green Beans (6%), Tomato Paste (4%), Light Milk (2%), Egg (1.5%), Garlic (1.5%), Gluten Free Breadcrumbs (1.5%), |

| | |
|---|---|
| | Chicken Stock (0.5%), Olive Oil (0.5%), Oregano (0%), Pink Salt (0%), Dried Basil (0%), Pepper (0%), Rosemary (0%). |
| Lamb Kofta with Middle Eastern Quinoa | Lamb Mince (27%), Zucchini (17%), Green Beans (17%), Broccoli (9%), Onion (8%), Red Capsicum (7%), Quinoa (5%), Olive Oil (2%), Almonds (1.5%), Parsley (1%), Garlic (1%), Lemon Rind (0.5%), Vegetable Stock (0.5%), Cumin (0.5%), Lemon Juice (0.5%), Pink Salt (0%), Chilli (0%), Paprika (0%), Chicken Stock (0%), Cardamom (0%), Oregano (0%), Thyme (0%), Pepper (0%), Ground Coriander (0%), Chilli Powder (0%). |
| Lemon & Coconut Protein Balls (snack) | Almond Meal (29%), Protein Powder (27%) (Whey Protein Isolate, Whey Protein Concentrate, Oligofructose, Vanilla Flavour, Salt, Stevia, Lactobacillus Plantarum, Guar Gum, Soy Lecithin), Dried Apple (Apple, Preservative (220)), Lemon Juice (22%), Coconut (1%), Lemon Essence (0.2%). |
| Low Carbohydrate Protein Loaf (snack) | Water, Protein 6 Seeds Bread Mix (43%) (Lupin Flour, Seeds (24% (Linseeds, Poppy Seeds, Sesame, Sunflower, Pepita, Chia Seeds), Whey Protein, Golden Flax meal, Egg Protein, Almond Meal, Baking Powder, Salt.), Egg White (13%) (Egg White, Water, Thickener (Xanthan Gum), Emulsifiers (Triethyl Citrate, Guam Gum)), Vinegar (1.5%). |
| Malaysian Spiced Pumpkin & Chickpea Soup | Pumpkin (28%), Carrot (10%), Chickpeas (10%), Coconut Milk (8%) (Coconut Cream, Xanthan Gum), Onion (5%), Faba Bean Protein (4%), Leek (2.5%), Sweet Potato (2.5%), Lemon Grass (0.5%), Vegetable Stock (0.5%), Curry Powder (0.5%), Fresh Coriander (0.5%), Garlic (0.5%), Olive Oil (0.5%), Pink Salt (0.5%), Kaffir Lime (0%), Pepper (0%), Chilli Powder (0%). |
| Mexican Stovetop Penne | Diced Tomato (27%) (Tomato, Citric Acid), Beef Mince (22%), Broccoli (7%), Carrot (7%), Zucchini (7%), Onion (7%), Gluten Free Pasta Penne (7%) (Maize Starch, Soy Flour, Potato Starch, Rice Starch).), Tomato Paste (3.5%), Parmesan Cheese (3%), Ricotta (2.5%), Jalapenos (1%), Beef Stock (1%), Parsley (1%), Light Milk (0.5%), Olive Oil (0.5%), Garlic (0.5%), Paprika (0%), Cumin (0%), Oregano (0%), Pink Salt (0%), Mixed Herbs (0%), Corn Starch (0%), Pepper (0%). |
| Mint Chocolate Protein Balls (snack) | Dates (43%), Almond Meal (22%), Protein Powder (22%) (Whey Protein Isolate, Whey Protein Concentrate, Oligofructose, Chocolate Flavour, Cocoa, Stevia, Lactobacillus Plantarum, Guar Gum, Soy Lecithin), Cacao Powder (6%), Cacao Nibs (4.5%), Coconut (1.5%), Coffee (0.5%), Mint Flavour (0%). |
| Mixed Berry Protein Bircher | Greek Yoghurt (46%) (Skim Milk, Live Yoghurt Cultures.), Mixed Berries (20%) (Blueberries, Raspberries, Blackberries, Red Currants), Ricotta Cheese (17%) (Whey, Milk, Salt, Food Acid), Natvia (2%), Dates (2%), Almonds (1.5%), Brown Rice Flakes (1.5%), Chia Seeds (1.5%), Faba Bean Protein (1.5%), LSA (1.5%) (Linseed, Sunflower Kernel, Almond), Shredded Coconut (1.5%), Pepitas (1.5%), Vanilla Extract (0%), Cinnamon (0%), Xanthan Gum (0%), Pink Salt (0%). |
| Naked Burrito Bowl | Chicken (33%), Carrot (13%), Green Capsicum (11%), Red Capsicum (11%), Red Kidney Beans (6%) (Red Kidney Beans, Salt, Firming Agent (509)), Corn Kernels (5%), Black beans (5%), Quinoa (5%), Cheese |

| | |
|---|---|
| | (3%), Onion (3%), Coriander (2.5%), Spring Onion (1%), Chicken Stock (0.5%), Garlic (0.5%), Olive Oil (0.5%), Pink Salt (0.5%), Ground Coriander (0%), Cumin (0%), Paprika (0%), Oregano (0%), Pepper (0%), Chilli Powder (0%). |
| Nutty Protein Balls (snack) | Dates (67%), Peanuts (16%), Vanilla Protein Powder (Whey Protein Isolate, Whey Protein Concentrate, Oligofructose, Vanilla Flavour, Salt, Stevia, Lactobacillus Plantarum, Guar Gum, Soy Lecithin), Coconut (1%), Pink Salt (0.5%). |
| Protein + Bolognese | Beef Mince (21%), Diced Tomato (21%) (Tomato, Citric Acid), Broccoli (17%), Gluten Free Pasta Penne (10%) (Maize Starch, Soy Flour, Potato Starch, Rice Starch).), Carrot (8%), Zucchini (8%), Onion (6%), Tomato Paste (3.5%), Parmesan Cheese (3%), Olive Oil (1%), Beef Stock (1%), Garlic (0.5%), Pink Salt (0%), Corn Starch (0%), Dried Basil (0%), Mixed Herbs (0%), Pepper (0%). |
| Quinoa and Protein Porridge | Quinoa Flakes (40%), Protein (27%) (Whey Protein Isolate, Whey Protein Concentrate, Vanilla Flavour, Guar Gum, Stevia, Salt, Soy Lecithin), Flaxseed Flakes (27%), Almond Meal (7%), Cinnamon (0.5%), |
| Quinoa Protein Granola | Puffed Amaranth (16%), Protein (16%) (Whey Protein Isolate, Whey Protein Concentrate, Oligofructose, Vanilla Flavour, Guar Gum, Stevia, Soy Lecithin), Puffed Quinoa (11%), Almonds (11%), Salted Peanuts (11%), Pumpkin Seed (9%), Orange Juice (7%), Sunflower Seed (6%), Dates (6%), Desiccated Coconut (4.5%), Black Chia Seed (2%), Flaxseed (1%), Cinnamon (0%) |
| Satay Chicken | Chicken (27%), Green Cabbage (13%), Carrot (13%), Red Cabbage (13%), Spring Onion (10%), Water (9%), Onion (6%), Coconut Milk (2.5%), Fresh Coriander (2%), Peanut Butter (1.5%), Olive Oil (1.5%), Turmeric (0.5%), Gluten Free Soy Sauce (0.5%), Cumin (0.5%), Ground Coriander (0.5%), Garlic (0%), Vegetable Stock (0%), Pink Salt (0%), Chilli (0%) (Red Chilli (74%), Vegetable Oil (11%), Vinegar (11%), Salt (3.50%), Acetic Acid (0.50%)), Corn Starch (0%). |
| Slow Cooked Sicilian Beef | Vegetables (57%) (Zucchini, Tomato (Diced Tomato, Tomato Juice, Citric Acid (330)), Red Onion, Carrot, Tomato Paste), Beef (40%), Olive Oil (0.5%), Corn Starch (0.5%), Garlic (0.5%), Smoked Paprika (0%), Natural Beef Stock (0%), Thyme (0%), Rosemary (0%). |
| South American Chilli Bean and Vegetable | Diced Tomato (28%) (Tomato, Citric Acid), Mushroom (7%), Red Kidney Beans (7%), Broccoli (6%), Red Capsicum (6%), Zucchini (6%), Carrot (6%), Tofu (5%), Onion (4%), Tomato Paste (4%), Green Peas (2.5%), Kale (2.5%), Corn Kernels (2.5%), Textured Vegetable Protein (2.5%), Leek (1.5%), Gluten Free Soy Sauce (1.5%), Olive Oil (1%), Faba Bean Protein (1%). Garlic (0.5%), Paprika (0.5%), Vegetable Stock (0.5%), Cumin (0%), Cinnamon (0%), Corn Starch (0%), Pepper (0%), Chilli Powder (0%). |
| Spanish Eggs | Egg (44%), Egg White (22%), Spinach (10%), Red Capsicum (8%), Chorizo (7%) (Pork, Salt, Spices, Maltodextrin (Maize), Garlic, Mineral Salts (451, 450), Antioxidant (316), Preservative (250), Natural Hog Casing, Wood Smoke.), Corn Kernels (4.5%), Spring Onion (3.5%), Olive Oil (1.5%), Garlic (0%), Pepper (0%). |

| | |
|---|---|
| Spiced Lentil Dahl | Tofu (15%), Broccoli (11%), Cauliflower Rice (11%) (Cauliflower, Turmeric), Mushroom (11%), Red Lentils (11%), Diced Tomato (9%) (Tomato, Citric Acid), Onion (5%), Coconut Milk (2%), Faba Bean Protein (2%), Vegetable Stock (1.5%), Olive Oil (1%), Gluten Free Soy Sauce (1%), Garlic (0.5%), Fresh Coriander (0.5%), Cumin (0%), Curry Powder (0%), Ginger (0%), Pink Salt (0%), Turmeric (0%), Garam Masala (0%), Cinnamon (0%), Chilli Powder (0%). |
| Spicy Mexican Pulled Beef | Beef (25%), Diced Tomato (17%) (Tomato, Citric Acid), Red Capsicum (11%), Green Capsicum (10%), Carrot (10%), Corn Kernels (6%), Black Beans (5%), Red Kidney Beans (5%), Tomato Paste (3%), Coriander (2%), Onion (0.5%), Gluten Free Soy Sauce (0.5%), Garlic (0.5%), Olive Oil (0.5%), Chicken Stock (0.5%), Paprika (0%), Cumin (0%), Pepper (0%), Oregano (0%), Corn Starch (0%), Chilli Powder (0%). |
| Sticky Date Protein Balls (snack) | Dates (41%), Almond Meal (21%), Vanilla Protein Powder (Whey Protein Isolate, Whey Protein Concentrate, Oligofructose, Vanilla Flavour, Salt, Stevia, Lactobacillus Plantarum, Guar Gum, Soy Lecithin Traces), Walnuts (16%), Coconut (1.5%). |
| Sunset Crush Smoothie | Mango, Orange, Passionfruit, Pea Protein (5.8%). |
| Super Barley Nut and Flaxseed Granola | Barleymax Tm Wholegrain Flakes (37%), Almonds (11%), Pumpkin Seed (11%), Salted Peanuts (11%), Protein (6%) (Whey Protein Isolate, Whey Protein Concentrate, Oligofructose, Vanilla Flavour, Guar Gum, Stevia, Soy Lecithin), Dates (6%), Orange Juice (6%), Sunflower Seed (4.5%), Black Chia Seed (3.5%), Coconut (2%), Flaxseed (2%), Cinnamon (0%). |
| Super Barley, Almond & Flaxseed Porridge | Barleymax Tm Wholegrain Flakes (29%), Protein (26%) (Whey Protein Isolate, Whey Protein Concentrate, Vanilla Flavour, Guar Gum, Stevia, Salt, Soy Lecithin), Oats (23%), Flaxseed Flakes (13%), Almond Meal (8%), Cinnamon (0.5%). |
| Super Green Protein Smoothie | Apple (23%), Cucumber (23%), Kiwi (14%), Pineapple (14%), Zucchini (12%), Faba Bean Protein (7%), Broccoli (5%), Spinach (1%), Kale (0.5%), Mint (0%). |
| Thai Green Chicken Curry | Chicken (31%), Broccoli (12%), Light Milk (12%), Brown Rice (7%), Coconut Milk (5%), Onion (5%), Eggplant (5%), Zucchini (5%), Coconut Cream (3.5%), Green Peas (3.5%), Spinach (3.5%), Diced Tomato (3%), Green Curry Paste (1%), Gluten Free Soy Sauce (0.5%), Fresh Coriander (0.5%), Garlic (0.5%), Ginger (0.5%), Lemon Grass (0.5%), Chilli (0%), Corn Starch (0%), Kaffir Lime (0%). |
| Trio of Green Soup | Broccoli (33%), Ricotta Cheese (17%) (Whey, Milk, Salt, Food Acid), Green Peas (10%), Edamame (10%), Spinach (8%), Light Milk (7%), Onion (3.5%), Potato (3.5%), Cannellini Beans (2.5%), Leek (2.5%), Faba Bean Protein (1%), Vegetable Stock (0.5%), Olive Oil (0.5%), Garlic (0.5%), Cumin (0%), Pepper (0%), Pink Salt (0%). |
| Vegan Bolognese | Diced Tomato (24%) (Tomato, Citric Acid), Broccoli (8%), Carrot (8%), Zucchini (8%), Gluten Free Pasta Penne (8%) (Maize Starch, Soy Flour, Potato Starch, Rice Starch), Celery (7%), Mushroom (7%), Onion |

| | |
|---|---|
| | (7%), Tomato Paste (4%), Green Lentils (4%), Walnuts (3.5%), Textured Vegetable Protein (2.5%), Faba Bean Protein (1.5%), Olive Oil (0.5%), Garlic (0.5%), Vegetable Stock (0.5%), Pink Salt (0.5%), Corn Starch (0%), Dried Basil (0%), Mixed Herbs (0%), Pepper (0%). |
| Vegetable and Chicken Cauliflower Rice | Vegetables (35%) (Peas, Carrot, Celery, Onion, Capsicum, Spring Onion), Cauliflower (28%), Chicken Breast (24%), Egg (8%), Garlic (2%), Ginger (1%), Olive Oil (0.5%), Peanuts (0.5%), Chilli Crushed (0%), Soy Sauce (0%) (Water, Soybean, Rice, Salt), Natural Chicken Stock (0%), Black Pepper (0%). |
| Vegetable and Chickpea Frittata | Egg (17%), Egg White (17%), Pumpkin (14%), Chickpeas (10%), Broccoli (9%), Red Capsicum (7%), Green Beans (7%), Sweet Potato (6%), Fetta Cheese (5%), Ricotta Cheese (2.5%), Spring Onion (2.5%), Light Tasty Cheese (2%), Olive Oil (0.5%), Garlic (0%), Parsley (0%), Pink Salt (0%), Curry Powder (0%), Canola Oil (0%), Pepper (0%). |
| Vegetarian Bolognese | Tomatoes (33%), Textured Soy Protein (9%) (Textured Soybean Protein (Soy Flour, Caramel Colour 150a)), Tomato Paste (5%), Broccoli (4.5%), Mushroom (4.5%), Celery (3.5%), Carrot (3.5%), Gluten Free Penne Pasta (3.5%), Onion (2.5%), Gluten Free Soy Sauce (1%), Stock (0.5%), Mixed Herbs (0.5%), Olive Oil (0.5%), Garlic (0%), Black Pepper (0%). |
| Vegetarian Masala | Vegetables (54%) (Cauliflower Floret, Pumpkin, Tomato (Diced Tomato, Tomato Juice, Citric Acid (330)), Paneer Cheese (19%) (Whey, Milk, Food Acid (330), Salt, Preservative (202)), Tofu (13%), Yoghurt (6%) (Pasteurised Milk, Milk Solids, Cultures), Milk Powder (2.5%), Olive Oil (1.5%), Coriander (1.5%), Ginger (1%), Garlic (0.5%), Sweet Paprika (0.5%), Cumin (0.5%), Garam Masala (0.5%), Corn Starch (0%), Chilli Powder (0%). |
| Wholemeal Beef Lasagne | Diced Tomato (26%) (Tomato, Citric Acid), Beef Mince (22%), Wholemeal Pasta Sheets (10%), Broccoli (8%), Carrot (8%), Zucchini (8%), Onion (6%), Tomato Paste (3.5%), Parmesan Cheese (3%), Ricotta Cheese (2.5%), Beef Stock (1%), Olive Oil (1%), Light Milk (0.5%), Garlic (0.5%), Pink Salt (0%), Dried Basil (0%), Mixed Herbs (0%), Corn Starch (0%), Pepper (0%). |
| Yellow Chicken Curry | Chicken (24%), Bok Choy (10%), Broccoli (10%), Choy Sum (10%), Zucchini (10%), Carrot (7%), Coconut Milk (6%) (Coconut Cream (51%), Water, Thickener (415)), Onion (5%), Milk Powder (2.5%), Almonds (1.5%), Yellow Curry Paste (1.5%), Coriander (1%), Corn Starch (0.5%), Olive Oil (0.5%), Sesame Oil (0.5%), Garlic (0.5%), Ginger (0.5%), Lemon Grass (0%), Turmeric (0%), Curry Powder (0%), Vegetable Stock (0%), Kaffir Lime (0%), Chilli (0%). |
| Yellow Vegetable Curry | Tofu (16%), Broccoli (11%), Coconut Milk (11%) (Coconut Cream, Xanthan Gum), Diced Tomato (11%) (Tomato, Citric Acid), Eggplant (11%), Edamame (7%), Zucchini (7%), Brown Rice (6%), Onion (6%), Peanuts (3.5%), Green Peas (2%), Faba Bean Protein (2%), Yellow Curry Paste (1.5%), Fresh Coriander (0.5%), Garlic (0.5%), Ginger (0.5%), Olive Oil (0.5%), Lemon Grass (0.5%), Vegetable Stock (0.5%), Gluten Free Soy Sauce (0.5%), Turmeric (0%), Chilli (0%), Kaffir Lime (0%), Corn Starch (0%). |

| Supplement-based meal replacements | |
|---|---|
| Item | Ingredient Composition |
| Bar Almond Butter & Date Flavour | Whey Crisps (Whey Protein (Milk), Starch (Corn)), Whey Protein (Milk), Date Paste, Almond, Honey, Inulin, Minerals (Potassium Hydrogen Phosphate, Calcium Phosphate, Sodium Chloride, Magnesium Carbonate, Potassium Chloride, Ferrous Sulphate, Zinc Sulphate, Manganese Sulphate, Copper Sulphate, Sodium Molybdate, Potassium Iodide, Sodium Selenite, Chromium Chloride), Flaxseed, Oat, Vitamins (C, E, Niacin, Pantothenic Acid, B6, Folic Acid, B12, Biotin, K, Thiamine, A, D, Riboflavin), Flavour. |
| Bar Berry Crunch Flavour | Soy Crisp (Soy Protein Isolate, Tapioca Starch, Salt), MILK Chocolate (17%) [Sugar, Cocoa Solids (6.6%), Whole MILK Powder, Emulsifier (Soy Lecithin), Flavour], Soy Protein Isolate, Polydextrose, Sorbitol, Soy Cores, Minerals (Potassium Citrate, Calcium Carbonate, Sodium Phosphate, Magnesium Carbonate, Sodium Citrate, Magnesium Phosphate, Calcium Phosphate, Copper Sulphate, Iron Pyrophosphate, Potassium Iodate, Sodium Selenate, Zinc Oxide, Manganese Sulphate), Glycerol, Fruit Preparation (1.5%) [Sugar, Raspberry Puree (0.3%), Fructose Syrup, Raspberry Juice Concentrate (0.2%), Apple Puree (0.2%), Lactose, Cherry Juice Concentrate (0.1%)], Maltodextrin (Corn), Inulin, Rapeseed Oil, Fructose-Glucose Syrup, Water, Vitamins (Ascorbic Acid, Vitamin E Acetate, Niacinamide, Vitamin A Acetate, Calcium Pantothenate, Biotin, Cyanocobalamin, Folic Acid, Cholecalciferol, Pyridoxine Hydrochloride, Riboflavin, Phylloquinone, Thiamine Mononitrate), Acidity Regulator (330), Flavour, Sweeteners (Sucralose, Acesulfame Potassium), Emulsifier (Soy Lecithin). |
| Bar Cappuccino Flavour | Soy Crisp (Soy Protein Isolate, Tapioca Starch, Salt), MILK Chocolate (17%) [Sugar, Cocoa Solids (6.6%), Whole MILK Powder, Emulsifier (Soy Lecithin), Flavour], Sorbitol, Soy Protein Isolate, Soybean, Minerals (Potassium Citrate, Calcium Carbonate, Sodium Phosphate, Sodium Citrate, Magnesium Carbonate, Magnesium Phosphate, Calcium Phosphate, Copper Sulphate, Iron Pyrophosphate, Potassium Iodate, Sodium Selenate, Zinc Oxide, Manganese Sulphate), Polydextrose, Maltodextrin (Corn), Glycerol, Water, Fructose-Glucose Syrup, Inulin, Rapeseed Oil, Flavour, Vitamins (Ascorbic Acid, Vitamin E Acetate, Niacinamide, Vitamin A Acetate, Calcium Pantothenate, Biotin, Cyanocobalamin, Folic Acid, Cholecalciferol, Pyridoxine Hydrochloride, Riboflavin, Phylloquinone, Thiamine Mononitrate), Sweeteners (Sucralose, Acesulfame Potassium), Emulsifier (Soy Lecithin). |
| Bar Chocolate Flavour | MILK Proteins, MILK Chocolate (14.5%) [Sugar, Cocoa Solids (6%), Whole MILK Powder, Emulsifier (Soy Lecithin), Flavour], Polydextrose, Sorbitol, Glycerol, Minerals (Potassium Citrate, Calcium Carbonate, Sodium Citrate, Sodium Phosphate, Magnesium Carbonate, Magnesium Phosphate, Calcium Phosphate, Copper Sulphate, Iron Pyrophosphate, Potassium Iodate, Sodium Selenate, Zinc Oxide, Manganese Sulphate), Cocoa Powder (5%), Vegetable Oils (Rapeseed, Safflower), Fructose Syrup, Soy |

| | |
|---|---|
| | Protein Isolate, Glucose Syrup, Flavour, Vitamins (Ascorbic Acid, Vitamin E Acetate, Niacinamide, Vitamin A Acetate, Calcium Pantothenate, Biotin, Cyanocobalamin, Folic Acid, Cholecalciferol, Pyridoxine Hydrochloride, Riboflavin, Phylloquinone, Thiamine Mononitrate), Emulsifier (Soy Lecithin). |
| Bar Cereal | Soy Crisp (Soy Protein Isolate, Tapioca Starch, Salt), Soy Protein Isolate, Sorbitol, Soy Cores, Polydextrose, Oat Flakes (7%), Maltodextrin (Corn), Minerals (Potassium Citrate, Calcium Carbonate, Sodium Phosphate, Sodium Citrate, Magnesium Carbonate, Magnesium Phosphate, Calcium Phosphate, Copper Sulphate, Iron Pyrophosphate, Potassium Iodate, Sodium Selenate, Zinc Oxide, Manganese Sulphate), Glycerol, Fructose-Glucose Syrup, Cranberries in Syrup [Cranberries (2.4%), Sugar, Sunflower Oil], Rapeseed Oil, Water, Vitamins (Ascorbic Acid, Vitamin E Acetate, Niacinamide, Vitamin A Acetate, Calcium Pantothenate, Biotin, Cyanocobalamin, Folic Acid, Cholecalciferol, Pyridoxine Hydrochloride, Riboflavin, Phylloquinone, Thiamine Mononitrate), Sweeteners (Sucralose, Acesulfame Potassium), Flavour, Emulsifier (Soy Lecithin). |
| Dessert Chocolate Flavour | Milk Protein (17%), Calcium Caseinate (9.5%), Sodium Caseinate (5.5%), Skimmed Milk Powder (24%), Vegetable Oil (Canola, Sunflower), Minerals (Potassium Citrate, Sodium Phosphate, Calcium Carbonate, Magnesium Carbonate, Potassium Chloride, Ferric Pyrophosphate, Zinc Sulphate, Copper Gluconate, Manganese Sulphate, Sodium Fluoride, Potassium Iodide, Sodium Selenite, Sodium Molybdate, Chromium Chloride), Maltodextrin (Corn), Cocoa Powder (5%), Vegetable Gum (414), Starch (Potato), Fructo-Oligosaccharide, Inulin, Glucose Syrup (Corn), Medium Chain Triglycerides, Sugar, Fish Oil, Flavour, Sweeteners (Aspartame, Acesulfame Potassium), Emulsifiers (472c, Soy Lecithin, 471), Antioxidants (301, 304, 306), Vitamins (E, Niacin, C, Pantothenic Acid, B6, B1, A, B2, Folic Acid, K, Biotin, D, B12). |
| Dessert Lemon Creme | MILK Protein (32%), Skimmed MILK Powder (25%), Maltodextrin (Corn), Minerals (Potassium Citrate, Sodium Phosphate, Magnesium Carbonate, Calcium Carbonate, Potassium Chloride, Sodium Molybdate, Ferric Pyrophosphate, Chromium Chloride, Zinc Sulphate, Copper Gluconate, Manganese Sulphate, Sodium Fluoride, Potassium Iodide, Sodium Selenite), Vegetable Oil (Canola, Sunflower), Vegetable Gum (414), Fructo-Oligosaccharide, Starch (Potato), Inulin, Glucose Syrup (Corn), Medium Chain Triglycerides, Sugar, Fish Oil, Sweeteners (Aspartame, Acesulfame Potassium), Flavour, Emulsifiers (472c, Soy Lecithin, 471), Antioxidants (301, 304, 306), Vitamins (E, Niacin, C, Pantothenic Acid, B6, B1, A, B2, Folic Acid, K, Biotin, D3, B12) Colour (Curcumin). |
| Shake Banana flavour | Skimmed MILK Powder (31%), MILK Proteins [Calcium Caseinate (20%), Sodium Caseinate (10%)], Maltodextrin (Corn), Vegetable Oil (Canola, Sunflower), Minerals (Potassium Citrate, Magnesium Carbonate, Calcium Phosphate, Sodium Chloride, Potassium Phosphate, Ferric Pyrophosphate, Copper Gluconate, Zinc Sulphate, Manganese Sulphate, Sodium Fluoride, Potassium Iodide, Sodium |

| | |
|---|---|
| | Molybdate, Sodium Selenite, Chromium Chloride), Vegetable Gum (414), Fructo-Oligosaccharide, Inulin, Medium Chain Triglycerides, Glucose Syrup (Corn), Sugar, Fish Oil, Emulsifiers (472c, Soy Lecithin, 471), Sweeteners (Aspartame, Acesulfame Potassium), Antioxidants (301, 304, 306), Vitamins (Vitamin E Acetate, Nicotinamide, Calcium Pantothenate, Sodium Ascorbate, Pyridoxine Hydrochloride, Thiamine Hydrochloride, Vitamin A Acetate, Riboflavin, Folic Acid, Phytomenadione, Cholecalciferol, Cyanocobalamin, Biotin), Colour (Curcumin, Beetroot), Flavour. |
| Shake Caramel flavour | Skimmed Milk Powder (31%), Milk Proteins [Calcium Caseinate (20%), Sodium Caseinate (10%)], Maltodextrin (Corn), Vegetable Oil (Canola, Sunflower), Minerals (Potassium Citrate, Magnesium Carbonate, Calcium Phosphate, Sodium Chloride, Potassium Phosphate, Ferric Pyrophosphate, Copper Gluconate, Zinc Sulphate, Manganese Sulphate, Sodium Fluoride, Potassium Iodide, Sodium Molybdate, Sodium Selenite, Chromium Chloride), Vegetable Gum (414), Fructo-Oligosaccharide, Inulin, Medium Chain Triglycerides, Glucose Syrup (Corn), Sugar, Fish Oil, Colour (150a), Emulsifiers (472c, Soy Lecithin, 471), Flavour, Antioxidants (301, 304, 306), Sweeteners (Aspartame, Acesulfame Potassium), Vitamins (Vitamin E Acetate, Nicotinamide, Calcium Pantothenate, Sodium Ascorbate, Pyridoxine Hydrochloride, Thiamine Hydrochloride, Vitamin A Acetate, Riboflavin, Folic Acid, Phytomenadione, Cholecalciferol, Cyanocobalamin, Biotin). |
| Shake Chocolate flavour | Skimmed Milk Powder (40%), Milk Proteins [Calcium Caseinate (19%), Sodium Caseinate (6%)], Vegetable Oils (Rapeseed, Sunflower), Maltodextrin (Corn), Cocoa (5%), Minerals (Potassium Citrate, Magnesium Carbonate, Sodium Citrate, Sodium Chloride, Potassium Hydrogen Phosphate, Ferric Pyrophosphate, Zinc Sulphate, Copper Gluconate, Manganese Sulphate, Potassium Iodide, Sodium Selenite, Chromium Chloride, Sodium Molybdate), Vegetable Gum (414), Fructo-oligosaccharide, Inulin, Medium Chain Triglycerides, Acidity Regulator (501), Vitamins (Sodium Ascorbate, Vitamin E Acetate, Nicotinamide, Calcium Pantothenate, Pyridoxine Hydrochloride, Riboflavin, Thiamine Hydrochloride, Vitamin A Acetate, Folic Acid, Phytomenadione, Biotin, Cholecalciferol, Cyanocobalamin), Sweeteners (Aspartame, Acesulfame Potassium), Flavour. |
| Shake Coffee Flavour | Skimmed Milk Powder (40%), Milk Proteins [Calcium Caseinate (19%), Sodium Caseinate (7%)], Vegetable Oils (Rapeseed, Sunflower), Maltodextrin (Corn), Minerals (Potassium Citrate, Magnesium Carbonate, Potassium Hydrogen Phosphate, Sodium Chloride, Sodium Citrate, Ferric Pyrophosphate, Zinc Sulphate, Copper Gluconate, Manganese Sulphate, Sodium Selenite, Chromium Chloride, Sodium Molybdate, Potassium Iodide), Coffee Extract (4.7%), Vegetable Gum (414), Fructo-oligosaccharides, Inulin, Medium Chain Triglycerides, Vitamins (Sodium Ascorbate, Vitamin E Acetate, Niacinamide, Calcium Pantothenate, Pyridoxine Hydrochloride, Riboflavin, Thiamine Hydrochloride, Vitamin A |

| | |
|---|---|
| | Acetate, Folic Acid, Phytomenadione, Biotin, Cholecalciferol, Cyanocobalamin) Sweeteners (Aspartame, Acesulfame Potassium), Flavour. |
| Shake Mocha Flavour | Skimmed MILK Powder (31%), MILK Proteins [Calcium Caseinate (20%), Sodium Caseinate (10%)], Maltodextrin (Corn), Vegetable Oil (Canola, Sunflower), Minerals (Potassium Citrate, Magnesium Carbonate, Calcium Phosphate, Sodium Chloride, Potassium Phosphate, Ferric Pyrophosphate, Copper Gluconate, Zinc Sulphate, Manganese Sulphate, Sodium Fluoride, Potassium Iodide, Sodium Molybdate, Sodium Selenite, Chromium Chloride), Coffee Extract (3.5%), Vegetable Gum (414), Fructo-Oligosaccharide, Cocoa Powder (2%), Inulin, Medium Chain Triglycerides, Glucose Syrup (Corn), Fish Oil, Sugar, Colour (Beetroot), Sweeteners (Aspartame, Acesulfame Potassium), Emulsifiers (472c, Soy Lecithin, 471), Antioxidants (301, 304, 306), Flavour, Vitamins (Vitamin E Acetate, Nicotinamide, Calcium Pantothenate, Sodium Ascorbate, Pyridoxine Hydrochloride, Thiamine Hydrochloride, Vitamin A Acetate, Riboflavin, Folic Acid, Phytomenadione, Cholecalciferol, Cyanocobalamin, Biotin). |
| Shake Strawberry flavour | Skimmed Milk Powder (31%), Milk Proteins [Calcium Caseinate (20%), Sodium Caseinate (10%)], Maltodextrin (Corn), Vegetable Oil (Canola, Sunflower), Minerals (Potassium Citrate, Magnesium Carbonate, Calcium Phosphate, Sodium Chloride, Potassium Phosphate, Ferric Pyrophosphate, Copper Gluconate, Zinc Sulphate, Manganese Sulphate, Sodium Fluoride, Potassium Iodide, Sodium Molybdate, Sodium Selenite, Chromium Chloride), Vegetable Gum (414), Fructo-Oligosaccharide, Inulin, Medium Chain Triglycerides, Glucose Syrup (Corn), Sugar, Fish Oil, Emulsifiers (472c, Soy Lecithin, 471), Colour (Beetroot), Sweeteners (Aspartame, Acesulfame Potassium), Antioxidants (301, 304, 306), Flavour, Vitamins (Vitamin E Acetate, Nicotinamide, Calcium Pantothenate, Sodium Ascorbate, Pyridoxine Hydrochloride, Thiamine Hydrochloride, Vitamin A Acetate, Riboflavin, Folic Acid, Phytomenadione, Cholecalciferol, Cyanocobalamin, Biotin). |
| Shake Vanilla flavour | Skimmed MILK Powder (31%), MILK Proteins [Calcium Caseinate (20%), Sodium Caseinate (10%)], Maltodextrin (Corn), Vegetable Oil (Canola, Sunflower), Minerals (Potassium Citrate, Magnesium Carbonate, Calcium Phosphate, Sodium Chloride, Potassium Phosphate, Ferric Pyrophosphate, Copper Gluconate, Zinc Sulphate, Manganese Sulphate, Sodium Fluoride, Potassium Iodide, Sodium Molybdate, Sodium Selenite, Chromium Chloride), Vegetable Gum (414), Fructo-Oligosaccharide, Inulin, Medium Chain Triglycerides, Glucose Syrup (Corn), Sugar, Fish Oil, Flavour, Emulsifiers (472c, Soy Lecithin, 471), Sweeteners (Aspartame, Acesulfame Potassium), Antioxidants (301, 304, 306), Vitamins (Vitamin E Acetate, Nicotinamide, Calcium Pantothenate, Sodium Ascorbate, Pyridoxine Hydrochloride, Thiamine Hydrochloride, Vitamin A Acetate, Riboflavin, Folic Acid, Phytomenadione, Cholecalciferol, Cyanocobalamin, Biotin), Colour (Curcumin). |

| | |
|---|---|
| Soup Vegetable | MILK Protein (Calcium Caseinate, Whey Protein), Hydrolysed Collagen (Bovine), Corn Starch, Minerals (Sodium Chloride, Potassium Phosphate, Magnesium Citrate, Potassium Citrate, Calcium Citrate, Calcium Phosphate, Ferric Pyrophosphate, Zinc Sulphate, Copper Gluconate, Manganese Sulphate, Sodium Selenite, Chromium Chloride, Sodium Molybdate, Potassium lodide), Vegetable Oil (Rapeseed, Sunflower), Potato (7%), Thickener (1442), Acacia Gum, Dried Leek (2.3%), Fructo-Oligosaccharide, Spinach, Inulin, Flavour, Dried Chive, Emulsifier (Soy Lecithin), Vitamins (C, E, Niacin, Pantothenic Acid, B6, B2, B1, A,K, Biotin, Folic Acid, D, B12), Glucose, Sweetener (Acesulfame Potassium), Colour (Curcumin). |
| Soup Chicken | MILK Protein (Calcium Caseinate, Whey Protein), Corn Starch, Hydrolysed Collagen (Bovine), Minerals (Potassium Phosphate, Sodium Chloride, Calcium Citrate, Magnesium Citrate, Potassium Citrate, Ferric Pyrophosphate, Calcium Carbonate, Zinc Sulphate, Copper Gluconate, Manganese Sulphate, Sodium Selenite, Chromium Chloride, Sodium Molybdate, Potassium lodide), Thickener (1442), Vegetable Oil (Rapeseed, Sunflower), Acacia Gum, Fructo-Oligosaccharides, Flavour, Inulin, Dried Parsley, Emulsifier (Soy Lecithin), Vitamins (C, E, Niacin, Pantothenic Acid, B6, B2, B1, A, K, Biotin, Folic Acid, D, B12), Glucose, Sweetener (Acesulfame Potassium), Colour (Curcumin). |

***Table S4.*** Food- and supplement-based recommended extras

<table>
<tr><th colspan="6">Food-based group recommended extras</th></tr>
<tr><td colspan="6">Fruit snacks</td></tr>
<tr><td colspan="3">0-5g Carbs<br>- 2-3 Passionfruit<br>- 220g Cooked rhubarb<br>- 30g Grapes<br>- 3 Small slices (~225g) of watermelon<br>- ¼ Small banana (firmer bananas will contain less sugar)<br>- 1 Guava ~90g</td><td colspan="3">5-10g Carbs<br>- 2 Fresh figs<br>- 2 Medium fresh apricots<br>- 2 Dates<br>- 1 Kiwi fruit<br>- 1 Orange<br>- 60g Cherries<br>- 200g Strawberries<br>- 80g Blueberries (fresh or frozen)<br>- 100g Raspberries (fresh or frozen)<br>- 50g Apple (~1/2 medium)<br>- 50g Pear (fresh or in natural juice, drained)<br>- 70g Plums<br>- 3 Prunes<br>- 4 Wedges of pineapple<br>- 1 Medium peach<br>- 1 Small wedge (125g) honeydew melon<br>- 1 Medium Mandarin<br>- 3 Tsps Goji Berries (~10g)<br>- 3/4 Small grapefruit<br>- 1 Small nectarine (~100g) (white or yellow)<br>- 130g Papaya</td></tr>
<tr><td colspan="6">Protein snacks</td></tr>
<tr><td colspan="2">General Protein snacks<br>1 Protein Ball<br>1 Cheesecake<br>- 4 egg whites (1 large whole egg)<br>- 100g tuna (tinned) in spring water</td><td colspan="2">Milk<br>Low/ no fat cows milk or soy milk<br>- Sanitarium Health and Wellbeing, So Good Unsweetened Almond</td><td colspan="2">Yoghurt<br>- Chobani 0.5% Fat Free plain Greek yoghurt<br>- YoPro Natural plain or flavoured yoghurt<br>- ProCal Icelandic SKYR</td></tr>
</table>

<table>
<tr><td>- 50g skinless chicken, turkey, lean trimmed meats (lamb, beef, pork)<br>- 70g white fish (non-oily; flake, flathead, perch etc. NOT crumbed, battered, with sauce<br>- 50g Oily Fish (salmon, ocean trout) - 50g 5-star pre-cooked lean/diet beefs mince - Tofu 150g</td><td colspan="2">Milk<br>Cheese<br>- Bega Super slim<br>- South Cape Tasmanian Reduced Fat Fetta<br>- Dairylea 97% Fat Free Slices<br>- Laughing Cow Light (3 wedges)</td><td>Natural<br>- Rockeby Farms QUARK<br>Natural Yoghurt<br>- Farmers Union Greek high<br>protein 0.2% fat yoghurt<br>– Aldi protein Greek<br>yoghurt - Woolworths<br>SKYR yoghurt</td></tr>
<tr><td colspan="4">Side Vegetables & Side Salad Options</td></tr>
<tr><td colspan="2">Salad – (1 cup per serve)<br>- Alfalfa sprouts<br>- Bamboo shoots<br>- Bean sprouts<br>- Cabbage (red or green)<br>- Capsicum<br>- Celery<br>- Cucumber<br>- Iceberg lettuce<br>- Kale<br>- Mung beans<br>- Onion - Radish<br>- Rocket<br>- Snow peas<br>- Spinach<br>- Spring onion<br>- Tomatoes<br>- Water chestnuts<br>- Watercress</td><td colspan="2">Vegetables – (1 cup per serve)<br>- Artichoke<br>- Asparagus<br>- Bok Choy<br>- Broccoli<br>- Broccolini<br>- Brussel sprouts<br>- Carrot<br>- Cauliflower<br>- Eggplant</td></tr>
<tr><td colspan="4">Optional Extras</td></tr>
<tr><td colspan="2">Fats<br>< 50 Calories per serve and<br>< 2g Carbs/protein<br>Suitable in side meals, as required</td><td colspan="2">- Olive oil - 1 tsp<br>- Coconut oil - 1 tsp<br>- Avocado – 1 tbsp<br>- Almonds x 6</td></tr>
</table>

| | |
|---|---|
| Females: 0-1 serves/day | - Other nuts x 5 |
| Herbs and Spices (fresh or dried)<br>Suitable to add to meals and sides | - All Spice<br>- Basil<br>- Celery flakes<br>- Chilli<br>- Chives<br>- Cinnamon<br>- Cloves<br>- Coriander<br>- Cumin<br>- Curry powder<br>- Dill<br>- Fennel<br>- Garlic<br>- Ginger<br>- Mint<br>- Mustard<br>- Nutmeg<br>- Oregano<br>- Paprika<br>- Parsley<br>- Pepper<br>- Rosemary<br>- Sage<br>- Tarragon<br>- Thyme<br>- Turmeric |
| Sauces and Condiments<br>Suitable to add to meals and sides<br>(Consider sodium may be high – use sparingly) | - Stock cube (low sodium)<br>- Vegetable soup (made from the above low starch vegetables)<br>- Miso<br>- Bonox<br>- Mustard<br>- Worcestershire sauce |

| | |
|---|---|
| | - Lemon / lime juice<br>- Salt or Lite Salt (reduced sodium salt)<br>- Pepper<br>- Balsamic vinegar<br>- Apple cider vinegar |
| Others<br>Not recommended for daily use (sparingly if needed only) | - Artificial sweeteners<br>- Unsweetened lollies / gum<br>- Diet jelly<br>- Dash of milk – any type (30ml) |
| **Drink Options** | |
| Minimum of 2-3 litres of fluid per day | Allowed<br>- Water (still or Sparkling)<br>- Coffee (black)<br>- Tea (black)<br>- Herbal Teas<br><br>Occasionally<br>- Diet Cordial<br>- Diet Soft Drink<br>- Kombucha |
| Vegetables to limit to under 2 tablespoons per day<br>*Please note that if you have, for example, 2 tablespoons of potato with your dinner, this will be in place of the 1 cup of side salad and/ or veg from page 1. | |
| - Potato<br>- Sweet Pumpkin<br>- Green Peas<br>- Corn<br>- Legumes<br>- Lentils | |
| **Supplement-based group recommended extras** | |
| **Low starch vegetables** | |
| Allowed<br>- Alfalfa sprouts | Avoid<br>- Potato |

| | |
|---|---|
| - Asparagus<br>- Bean Sprouts<br>- Bok Choy<br>- Broccoli<br>- Brussels sprouts<br>- Cabbage<br>- Capsicum<br>- Carrots<br>- Cauliflower<br>- Celery<br>- Cucumber<br>- Eggplant<br>- Green Beans<br>- Konjac Noodles (Slendier/Slim Pasta range)<br>- Lettuce (all types)<br>- Leeks<br>- Mushrooms<br>- Onions<br>- Radish<br>- Shallots<br>- Silver Beet<br>- Snow Peas<br>- Spinach<br>- Squash<br>- Tomatoes<br>- Watercress<br>- Zucchini | - Sweet Potato<br>- Green Peas<br>- Corn<br>- Legumes<br>- Lentils<br>- Parsnip<br>- Pumpkin<br>- Turnip |
| **Soups** | |
| Allowed<br>- Stock cubes<br>- Bonox (in moderation)<br>- Vegetables soups made from allowed vegetables<br>- Miso soup | Avoid<br>- All other soups |

| **Sauces and condiments** | |
|---|---|
| Allowed<br>- Lemon and lime juice<br>- Vinegar<br>- Worcestershire sauce<br>- Tabasco sauce<br>- Soy sauce (in moderation)<br>- Chilli<br>- Diet, oil free or fat free dressings<br>- Mustard<br>- Tomato paste | Avoid<br>- Cream<br>- High calorie simmer sauces and dressings |
| **Herbs and spices** | |
| Allowed<br>- All Spice<br>- Basil<br>- Celery flakes<br>- Chilli<br>- Chives<br>- Cinnamon<br>- Cloves<br>- Coriander<br>- Cumin<br>- Curry powder<br>- Dill<br>- Fennel<br>- Garlic<br>- Ginger<br>- Mint<br>- Mustard seed<br>- Nutmeg<br>- Oregano<br>- Paprika<br>- Parsley | Avoid<br>- None |

| | |
|---|---|
| - Pepper<br>- Rosemary<br>- Sage<br>- Tarragon<br>- Thyme<br>- Turmeric | |
| **Miscellaneous** | |
| Allowed<br>- Artificial sweeteners<br>- Sugar free lollies and gum<br>- Diet jelly<br>- Flavour essence<br>- Diet topping | Avoid<br>- None |
| **Low energy drinks** | |
| Allowed<br>- Water<br>- Soda water<br>- Diet soft drinks and cordial<br>- Plain mineral water<br>- Tea and coffee (no or 30mL skin milk no sugar)<br>- Herbal teas | Avoid<br>- Fruit juice<br>- Alcohol<br>- Soft drinks<br>- Cordial |

**Detailed methods for gut microbiome sample collection and secondary outcome measures**

*Sample collection*

Faecal and blood samples were collected from participants at both the baseline and three-week follow-up visits. Faecal samples (15 grams) were collected using a Copan Italia SPA FLOQSwab in an active drying tube (FLOQSwab-ADT) that includes an internal desiccant for the purpose of preserving and storing the samples at room temperature for up to four weeks. The faecal collections took place within 24–48 hours of participants attending face-to-face appointments at Australian Clinical Labs (ACL, Geelong, Australia). After preparing and cleaning the skin with Briemarpak alcohol wipes (containing 70% isopropyl alcohol), a total of 40 millilitres of blood samples were collected using BD Vacutainer® tubes (yellow top), butterfly needles, and barrels. Blood collection took place in the morning after an overnight fast, before participants had breakfast. The collected faecal and blood samples were then processed and analysed by the accredited and contracted laboratories, Microba Pty Ltd and ACL, respectively. Following the processing and analysis procedures, any remaining faecal and blood material was returned to the investigators and securely stored in a freezer maintained at a temperature of minus 80 degrees Celsius. This freezer was in a protected facility at Barwon Health in Geelong, Victoria, Australia, accessible only via swipe card access.

*Measurements of serum metabolic and inflammatory parameters*

Serum markers of inflammation (including homocysteine, interleukin-beta (IL-β), interleukin-6 (IL-6, pg/mL), and tumour necrosis factor-alpha (TNF-α, pg/mL)) were assayed using the BD^tm Cytomteric Bead Array assay platform by SA Pathology at Adelaide Women's & Children's Hospital, Adelaide, South Australia, Australia. Serum leptin was assayed using Merck Millipore radioimmunoassay (Iodine 125) kit by the Royal Prince Alfred's Central Sydney Pathology Services, Camperdown, New South Wales, Australia. All other serum biomarkers (glucose, insulin, markers of liver function (alanine aminotransferase (ALT), gamma-glutamyl transpeptidase (GGT), alkaline phosphatase (ALP), aspartate aminotransferase (AST), total bilirubin, total albumin, total protein, and total globulin), markers of lipids (total cholesterol, high-density lipoprotein (HDL) cholesterol, low-density lipoprotein (LDL) cholesterol, non-HDL cholesterol, LDL/HDL ratio, cholesterol/HDL ratio, triglycerides) were assayed using Siemens' ADVIA® Chemistry kits by Australian Clinical Labs, Clayton, Victoria, Australia.

*Microbiome analysis by whole-genome metagenomic sequencing*

*Library preparation and sequencing*

DNA Extraction: Faecal samples underwent DNA extraction using the Qiagen DNeasy 96 PowerSoil Pro QIAcube HT Kit (Qiagen 47021) in a 2-millilitre deep well plate format, following the manufacturer's instructions. A modified initial processing step was applied on the QIAcube HT DNA extraction system (Qiagen 9001793). Mechanical lysis was performed using PowerBead Pro beads (Qiagen 19311) and optimised chemistry to enhance bacteria and fungi lysis. The kit included technology to remove inhibitors typically present in faecal samples. The resulting DNA was quantified using a high-sensitivity dsDNA fluorometric assay (QuantIT, ThermoFisher, Q33120), and samples needed to reach a minimum concentration of 0.2 nanograms per microlitre to meet quality control requirements.

Library Preparation: Libraries were constructed with the Illumina DNA Prep (M) Tagmentation Kit (Illumina, 20018705) and IDT for Illumina DNA/RNA UD Index Sets A-D (Illumina 20027213-16), following the manufacturer's guidelines. Adjustments were made to accommodate processing in a

384-plate format. The libraries were evaluated using a high-sensitivity dsDNA fluorometric assay (QuantIT, ThermoFisher, Q33120) and were visualised with capillary gel electrophoresis using the QIAxcel DNA High Resolution Kit (Qiagen, 929002). The libraries needed to meet specific criteria for average size, smallest and largest fragment gating, and concentration.

Sequencing: Individual libraries were combined in equimolar quantities to create sequencing pools. These pools were assessed using a high-sensitivity dsDNA fluorometric assay (QuantIT, ThermoFisher, Q33120) and capillary gel electrophoresis with the QIAxcel DNA High Resolution Kit (Qiagen, 929002). The pools were required to meet specific criteria for average size, smallest and largest fragment gating, and concentration as per Microba standards. The sequencing pools were loaded and sequenced on the NovaSeq6000 (Illumina) using v1.5 300 bp PE sequencing reagents, following the manufacturer's instructions. Sequence data underwent checks for minimum performance requirements regarding yield and sequence quality. Additionally, the data was reviewed for the performance of known control samples, included in each processing run, to ensure proper reagent contribution and reporting accuracy based on statistical measures and Hellinger distance assessment.

*Sequence processing and bioinformatics*

To quantify the abundance of genes and pathways within the metagenomic samples, the Microba Gene and Pathway Profiler (MGPP) v1.0 was employed. MGPP operates in a two-step process. Initially, all Open Reading Frames (ORFs) from genomes within the Microba Genome Database (MGDB) were grouped, utilizing a 90% identity threshold over 80% of read length, with the assistance of MMSeqs2 Release 10-6d92c (1). Gene clusters were then annotated with UniRef90 (2) identifiers and linked to Enzyme Commission annotations (accessed via UniProt 2019/04) and Transporter Classification Database (3) annotations, facilitated through the UniProt ID mapping service (www.uniprot.org/uploadlists/).

Enzyme Commission annotations were instrumental in identifying the presence of MetaCyc (4) pathways within each genome using the enrichM tool (https://github.com/geronimp/enrichM). Pathways that exhibited completeness levels exceeding 80% were classified as encoded. In the subsequent step, the accumulation of DNA sequencing read pairs aligning with gene sequences from proteins within an MGENES protein cluster was computed. This facilitated the estimation of pathway abundances for species identified by MCP, achieved by averaging the read counts of all genes associated with each enzyme in the respective pathway.

*Metabolic and inflammatory parameters*

Anthropometric measurements were recorded for height, weight, and hip and waist circumferences, using weighing scales and a measuring tape. BMI was estimated using the following formula:

$$BMI = (weight) / (height)^2$$

Where:

Weight is the participant's weight in kilograms.
Height is the participant's height in metres.

The following serum biomarkers were collected in fasted state:

- Leptin (measured in nanograms per millilitre, ng/mL)
- Glucose (measured in millimoles per litre, mmol/L) and insulin (measured in milliunits per litre, mU/L)

- Markers of liver function, including ALT (measured in units per litre, (U/L)), GGT (U/L), ALP (U/L), AST (U/L), total bilirubin (measured in micromoles per litre, μmol/L), total albumin (measured in grams per litre, g/L), total protein (g/L), and total globulin (g/L)
- Markers of lipids, including CHO (mmol/L), HDL (mmol/L), LDL (mmol/L), non-HDL (mmol/L), LDL/HDL ratio (mmol/L), CHO/HDL ratio (mmol/L), and triglycerides (mmol/L).
- Markers of inflammation, including homocysteine (measured in micromoles per litre, μmol/L), IL-β (measured in picograms per millilitre, pg/mL), IL-6 (pg/mL), TNF-α (pg/mL).

*Mental health symptoms and perceived well-being*
Depressive, anxiety, and stress symptoms were evaluated using the Depression Anxiety Stress Scale-21 (DASS-21). This validated self-report questionnaire comprises 21 items grouped into three 7-item subscales for depression, anxiety, and stress (5). The items are applicable to both clinical and non-clinical populations, encompassing various affect-related physical and mental symptoms. Participants rated each item on a 4-point Likert scale, ranging from 0 (not applicable to me at all) to 3 (highly applicable to me most of the time). Higher scores indicated more pronounced symptoms of dysphoric mood, while a score of 0 denoted the absence of mood-related disturbances. Participants reported on their experiences over the past week. Previous research has demonstrated good validity and high internal consistency for all three DASS-21 subscales (6).

Perceived well-being was assessed using the World Health Organization Wellbeing Scale (WHO-5), a validated 5-item questionnaire (7). Participants rated the relevance of five statements to their experiences over the preceding two weeks on a Likert scale ranging from 0 to 5. Higher scores indicated better well-being, with 0 representing the poorest imaginable well-being. A raw score between 0 and 25 was multiplied by 4 to derive a final score ranging from 0 (representing the worst imaginable well-being) to 100 (representing the best imaginable well-being) (8).

The Athens Insomnia Scale (AIS) is an 8-item self-reported assessment of sleep-related difficulties, quality, and duration during the past three weeks. Each item is rated on a 4-point Likert scale, ranging from 0 to 3, with higher values indicating more severe sleep-related issues. The maximum total score achievable is 24 (9). The AIS designed in accordance with the criteria outlined in the International Statistical Classification of Diseases and Related Health Problems (ICD-10) by the World Health Organization (9). It explores sleep induction, awakenings during the night, total sleep duration, sleep quality, as well as well-being, functional capacity, and daytime sleepiness. The AIS exhibits strong consistency, reliability, and validity and has been validated in both clinical and non-clinical populations (9).

*Gastrointestinal symptoms*
The Visual Analogue Scale for Irritable Bowel Syndrome (VAS-IBS) is a validated 9-item questionnaire designed to discriminate self-reported symptoms and the degree of symptom change between individuals with and without IBS (10, 11). The questionnaire captured changes within the preceding two weeks. Symptom severity for items 1 to 7 was presented on a 10-centimeter horizontal line, with higher scores indicating better symptom outcomes. The furthest left point represented 0, indicating "very bad," while the furthest right point represented 100, indicating "very good." The 8th and 9th items, related to defecation urgency and level of defecation emptiness, were answered in a yes/no format. Individual items scores were not combined into sub-scales or a total score. Instead, scores for each individual item were assessed between and within groups (10, 11).

*Stool consistency*
Stool consistency was measured using the Bristol Stool Form Scale (BSFS), a 7-point self-reported scale that spans from the firmest stool (Type 1) to the softest stool (Type 7). The BSFS captured participants' stool consistency during the preceding two weeks (12). A mean score was estimated for participants

in each group across one week. The BSFS is used extensively in clinical practice and research for stool form measurement and has acceptable validity and reliability in healthy adults and individuals with diarrhoea-predominant IBS (12, 13).

*Physical activity*
The International Physical Activity Questionnaire-Short Form (IPAQ-SF) measured levels of physical in/activity (14). The IPAQ-SF consists of seven open-ended questions relating to participants' physical activity during the preceding seven days, as well as measures of inactivity and intensity of physical activity (15). It captures Metabolic Equivalent of Task (MET) minutes per week. A categorical score of low, medium, or high was calculated for each participant as a summary of walking, moderate and vigorous MET scores (16) estimated as per guidelines (17). The IPAQ-SF has shown an excellent test-retest reliability (18) and adequate measurement properties for monitoring population levels of physical activity in adults across diverse settings (19).

*Dietary intake*
Dietary intake was measured at baseline by the Dietary Questionnaire for Epidemiological Studies v3.2 (DQES v3.2) (Appendix 8). This tool is modified food frequency tool developed by Cancer Council Victoria. The DQES v3.2 incorporates 80 items across five main food categories. Questions relate to the past 12 months and include overall intake of energy, fibre, macro and micronutrients. The Nutritional Assessment Office, Cancer Council Victoria scores these data. This instrument is commonly used in Australian research (20). As per the methods employed elsewhere (21), the habitual consumption of ultra-processed foods at baseline in weight (absolute and proportion of total grams per day) was estimated by applying the NOVA food classification system to DQES v3.2. Examples of ultra-processed foods include: soft drinks, sweet or savoury packaged snacks, confectionery, packaged breads and buns, margarine, reconstituted meat products and pre-prepared frozen or shelf-stable dishes (22).

Self-reported data pertaining to sociodemographic- (age, country of birth, marital status, employment status, and household income), dietary- (food intolerance, special dietary requirements), and health- (smoking status, medication use) were also collected.
